# Genetic analysis of cognitive preservation in the midwestern Amish reveals a novel locus on chromosome 2

**DOI:** 10.1101/2023.12.13.23299932

**Authors:** Leighanne R Main, Yeunjoo E Song, Audrey Lynn, Renee A Laux, Kristy L Miskimen, Michael D Osterman, Michael L Cuccaro, Paula K Ogrocki, Alan J Lerner, Jeffery M Vance, M Denise Fuzzell, Sarada L Fuzzell, Sherri D Hochstetler, Daniel A Dorfsman, Laura J Caywood, Michael B Prough, Larry D Adams, Jason E Clouse, Sharlene D Herington, William K Scott, Margaret A Pericak-Vance, Jonathan L Haines

**Author notes:** Corresponding Authors: Leighanne R Main, and Jonathan L Haines.

## Abstract

**INTRODUCTION:** Alzheimer disease (AD) remains a debilitating condition with limited treatments and additional therapeutic targets needed. Identifying AD protective genetic loci may identify new targets and accelerate identification of therapeutic treatments. We examined a founder population to identify loci associated with cognitive preservation into advanced age.

**METHODS:** Genome-wide association and linkage analyses were performed on 946 examined and sampled Amish individuals, aged 76-95, who were either cognitively unimpaired (CU) or impaired (CI).

**RESULTS:** 12 SNPs demonstrated suggestive association (P≤5x10^-4^) with cognitive preservation. Genetic linkage analyses identified >100 significant (LOD≥3.3) SNPs, some which overlapped with the association results. Only one locus on chromosome 2 retained significance across multiple analyses.

**DISCUSSION:** A novel significant result for cognitive preservation on chromosome 2 includes the genes *LRRTM4* and *CTNNA2*. Additionally, the lead SNP, rs1402906, impacts the POU3F2 transcription factor binding affinity, which regulates *LRRTM4* and *CTNNA2*.

## 1 BACKGROUND

Dementia, defined broadly as a decline or loss in memory and other cognitive abilities, is an overarching term that incorporates many underlying conditions. Alzheimer disease (AD), the most common form of dementia (∼60-80% of cases), affects more than five million people in the U.S. and is becoming more prevalent as the world population ages.[1] To date there have been over 70 genetic loci associated with *increased* risk for AD-associated dementia,[2–4] some of which have been targets for therapeutic development. However, no existing treatments or therapies addressing AD and other dementias have been sufficient to significantly ameliorate AD pathology or delay onset and decline.[5] An alternative and less often used approach for generating therapeutic targets is the identification of genetic variation that associates with a *decreased* risk for AD-associated dementia (e.g., protective loci). Focusing on protective loci is a strategy demonstrating higher success rates than focusing on loci with increased risk when translated into therapeutics.[6]

Phenotypically, protective AD genetic loci result in maintenance of cognitive abilities (e.g., cognitive preservation). To identify loci promoting cognitive preservation, we analyzed members of the Ohio and Indiana Amish, a founder population of European ancestry. For our analyses, we identified older individuals who were cognitively unimpaired (CU) but at high risk for developing AD, defined as having a first-degree relative with dementia. Analyses included cognitively impaired (CI) individuals as controls. The relatively uniform lifestyle of the midwestern Amish reduces the variability of environmental influences on many phenotypes, while their endogamous cultural norms increase the frequency of some normally rare genomic variants.[7,8] Both genetic association analyses and family-based linkage analyses were used to identify loci associated with cognitive preservation.

## 2 METHODS

### 2.1 Data Collection and Cognitive Screening

The analyzed dataset consisted of Amish individuals from Ohio and Indiana who were 76-95 years old at their last exam. Individuals underwent a panel of neurocognitive assessments to evaluate their cognitive status. Assessments included the Consortium to Establish a Registry for Alzheimer’s Disease (CERAD) neuropsychological assessment[9], the Modified Mini-Mental State (3MS) exam[10], the AD8 Dementia Screening Interview[11], and the Trail Making Test for Dementia Studies.[12] Cognitive data from 2,096 individuals were available for analysis. Individuals were classified as cognitively impaired (CI) based on established testing thresholds, with determinations confirmed by a clinical adjudication board. Categories defined by the adjudication board included cognitively unimpaired for age-normed benchmarks (CU), mild cognitive impairment (MCI), borderline impairment, or cognitively impaired (CI). Additional personal and medical information was assessed to provide context for other factors related to cognition scores, such as self-reported depression. Individuals unable to be categorized as CI or CU were excluded from genetic analyses. All participants provided written informed consent for participation in this study.

### 2.2 Genotyping

Blood was collected from participants for DNA extraction. All samples were genotyped via the Multi-Ethnic Genotyping Array (MEGA^ex^) or Global Screening Array (GSA) genotyping chips. The MEGA^ex^ chip recognized over 2 million loci, and the GSA chip recognized 660,000 loci. Custom markers were also included on the MEGA^ex^ chip (6,000 SNPs), including disease-associated loci previously identified in Amish and European populations.[13,14]

### 2.3 Genotype Quality Control

Initial quality control (QC) was done as previously described.[15] 256,978 SNPs passed QC across both chips. After additional pruning for linkage disequilibrium (LD), 167,196 SNPs were used to inform the number of effective independent tests done in the association analyses. Genotyped individuals were filtered for sex discrepancies and low call rates (<96%). Familial relationships were verified across samples. Age was defined as follows for all analyses: for CU individuals, age at the last study visit; for CI individuals, age at CI diagnosis. Participants whose age or familial relationships were missing were not included in the current analyses. Additional samples were dropped due to being under 75 years of age (n = 748) or a diagnosis other than CI/CU (n = 343). After QC, 946 individuals were available for analysis (66% CU; 59% female, Supplementary Table 1). Mean age for both CU and CI individuals was 82 years (Supplementary Figure 1).

### 2.4 Amish Genealogy

To evaluate familial relationships in this dataset, we consulted the Anabaptist Genealogical Database (AGDB).[16] All individuals were connected into a single, large, 14-generation pedigree (n=8,222) generated using pedigraph[17] (Figure 1A). These relationships were used to correct for relatedness in our genetic analyses. To conduct chromosome-wide genetic linkage analyses smaller pedigrees were required, so PedCut[18] was used to create sub-pedigrees with at least two cognitively preserved individuals in each (see Figure 1B for an example). The 14-generation pedigree was split based on pedigree size, or bit-size. Bit sizes evaluated were 21 (n = 103 sub-pedigrees) and 22 (n = 98) for linkage analysis on the autosomes. An additional 23-bit (n = 94) pedigree size was used for linkage analysis on the X chromosome.

**Figure 1.**
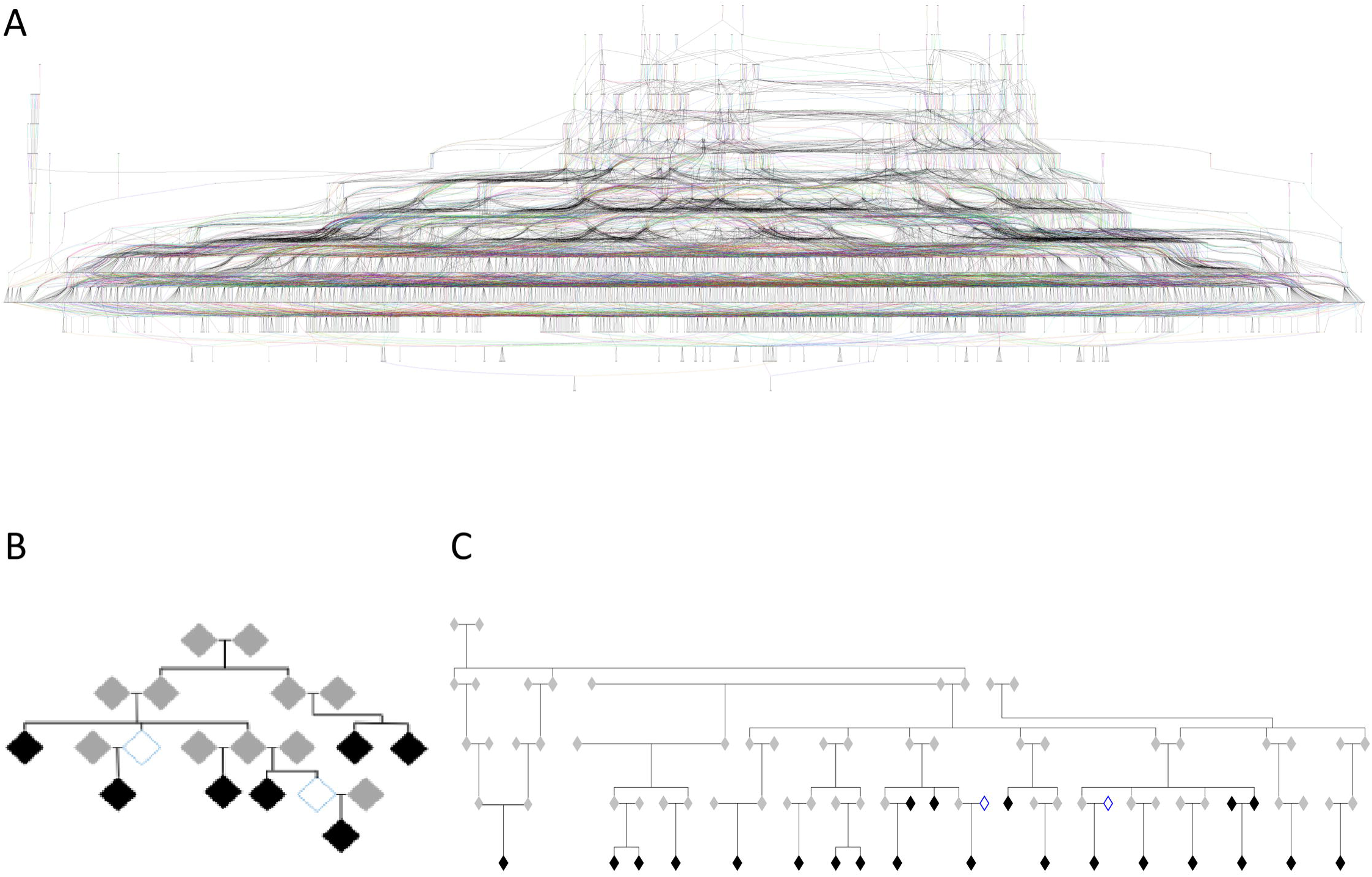
Amish pedigrees. (A) 8,222 individuals spanning 14 generations are present in this large pedigree. Relationship data was drawn from the AGDB to incorporate ascertained individuals into existing family trees. Colored lines are used to connect generations between families. (B) Example of a 21-bit sub-pedigree generated via PedCut from the 14-generation pedigree to create computationally tractable units for analyses. An individual’s sex was included during analyses but was removed here for privacy protection. Black shading indicates individuals who were CU at their last assessment; clear shading indicates individuals with CI. Grey shading indicates individuals for whom no information besides relationship data was available. (C) Larger pedigree connecting chromosome 2-linked smaller pedigrees. To protect privacy, sex information was included for analyses but was removed here. Shading indicates the same phenotypes as in 1B.

### 2.5 Principal Component Analysis and Association Analysis

Principal Component Analysis (PCA) was performed via Principal Component Analysis in Related Samples (PC-AiR), part of the GENetic Estimation and Inference in Structured samples (GENESIS) R package.[19] PC-AiR adjusted for cryptic relatedness to generate principal components (PCs) based on the overall population structure, not just family structure. PC-AiR created kinship estimates for each individual based on the pedigree relationship information obtained from the 14-generation pedigree (Figure 1A). Model-free estimation of recent genetic relatedness (PC-Relate)[20] then used the PCs generated from PC-AiR to adjust for the population substructure and ancestry to provide an accurate estimate of recent genetic relatedness due to family structure, correcting for the extensive and complex relatedness in this dataset by generating a Genetic Relationship Matrix (GRM). GENESIS software was then used for association analysis.[19] Association analysis was completed using covariates of age, sex, and the GRM, adjusting for the first two PCs.

To accommodate the LD structure specific to the Amish, we used the simpleM method[21–23] to define appropriate significance thresholds for the study population. SimpleM uses PCA and linkage disequilibrium (LD) between SNPs to help estimate the effective number of independent tests for the dataset. This method generated a significance threshold of 6.4×10^-7^.

The X chromosome was also investigated for association. 9,852 X-linked SNPs were available after QC. XWAS[24] was used to perform sex-stratified association testing. Two tests were performed, one treating males as heterozygous for SNPs on the X chromosome, and the other treating males as homozygous for SNPs on the X chromosome, both yielded to the same results. Age and the first five PCs were included as covariates.

### 2.6 Linkage Analysis

Due to computational complexity, the 14-generation 8,222-person Amish pedigree (Figure 1A) was divided into smaller sub-pedigrees (e.g., Figure 1B) to perform both nonparametric (NPL) and parametric linkage analysis. A total of 295 sub-pedigrees were used in linkage analyses (Supplementary Table 2).

Model-based (parametric) and model-free (nonparametric or NPL) linkage analyses were performed using MERLIN.[25] To test for linkage, the null hypothesis of no linkage was compared to the likelihood of linkage (0 ≤ L□*₁* < 0.5) between the phenotype and SNP within each sub-pedigree. A logarithm of the odds (LOD) score was calculated within each sub-pedigree and then summed across sub-pedigrees. In addition, the proportion of sub-pedigrees linked to a particular SNP (α) was calculated as part of a heterogeneity LOD score (HLOD).

For parametric two-point and multipoint linkage tests, both dominant and recessive models were tested on sub-pedigrees. These pedigrees each contained at least three but up to 11 CU individuals, with varying numbers of CI individuals ranging from zero to two. For tests modeling a dominant inheritance model, a trait allele frequency (AF) of 0.001 was used with penetrances of 0.0001, 1.0, and 1.0 for zero, one, and two copies of the trait allele, respectively. For the recessive model, the trait AF was 0.05 with penetrance of 0.0001, 0.0001, and 1.0 for zero, one, and two copies of the trait allele, respectively.

In nonparametric linkage (NPL) and parametric analyses, both two-point and multipoint linkage was examined. Two-point NPL and parametric analyses were performed on the full set of post-QC SNPs (n = 256,978), while multipoint NPL and parametric analyses were carried out on two different sets of pruned SNPs to minimize the impact of linkage disequilibrium (LD). An r^2^ threshold of 0.16 was used to define a set of 85,857 pruned SNPs for the multipoint analyses.[26] MERLIN’s NPL_all_ function was utilized for NPL analyses to construct a linear model of linkage. LOD and HLOD scores surpassing the defined thresholds for suggestive (≥ 1.86) and significant linkage (≥ 3.3)[27] were examined in more depth.

MINX software[25], an extension of MERLIN, was used for X chromosome linkage analysis. For two-point NPL and parametric analyses, a set of 9,852 X chromosome SNPs was used, and for multipoint NPL and parametric analyses, a pruned set of 153 SNPs with an r^2^ < 0.02 and MAF ≥ 0.40 was used. Multipoint NPL analysis utilized a sex-averaged genetic map[28], and model penetrances, adjusted for males and females, were the same as defined for the autosomal linkage analyses above.

For the subset of the loci where HLOD scores surpassed the significance threshold, 21- and 22-bit pedigrees driving the positive HLOD scores were examined for more distant connections allowing for the construction of larger pedigrees. 21- or 22-bit sub-pedigrees were merged when either identical AGBD IDs or first-degree relative AGBD IDs appeared (see Figure 1C for a larger pedigree example constructed for chromosome 2-linked sub-pedigrees). Six loci, one on each of chromosomes 1, 2, 3, 7, 11 and 17 met significance criteria, and larger pedigrees were possible for five of these loci, the exception being the chromosome 3 locus. MORGAN[29,30] was used to calculate region-specific LOD scores for these larger, more complex pedigrees using both dominant and recessive models. Any linkage originally appearing under dominant inheritance patterns was verified by MCMC analysis also utilizing dominant parameters, recessive results were treated similarly.

MORGAN allowed for larger pedigrees and provided an alternative linkage method, Markov chain Monte Carlo (MCMC), to investigate the same loci. The trait allele frequency for CU was coded as very rare (CU = 0.0001) for all analyses, similar to, but slightly more restrictive than previous models run with MERLIN, in order to specify a single founder for each allele. Additionally, MCMC penetrance models utilized the same penetrances specified in MERLIN analyses. MCMC was performed with a maximum of 8 SNPs, spaced at 1-5 cM intervals. The genetic maps generated by Halldorsson et al., 2019 were used.[31] D’ and r^2^ values were generated specifically for our Amish population to verify that the resulting LOD scores were not influenced by LD.

## 3 RESULTS

### 3.1 Genome-Wide Association Analysis for Cognitive Preservation

No SNPs reached the genome-wide significance threshold defined by simpleM for this population, but eleven loci (chromosomes 1, 3, 5, 6, 7, 11, 13, 14, 15, 16) surpassed the suggestive threshold (p ≤ 10^-4^) (Supplementary Table 3 and Figure 2). Genomic inflation was 0.98 after the GRM, age, and sex covariates were applied to the model (Supplementary Figure 2A).

**Figure 2.**
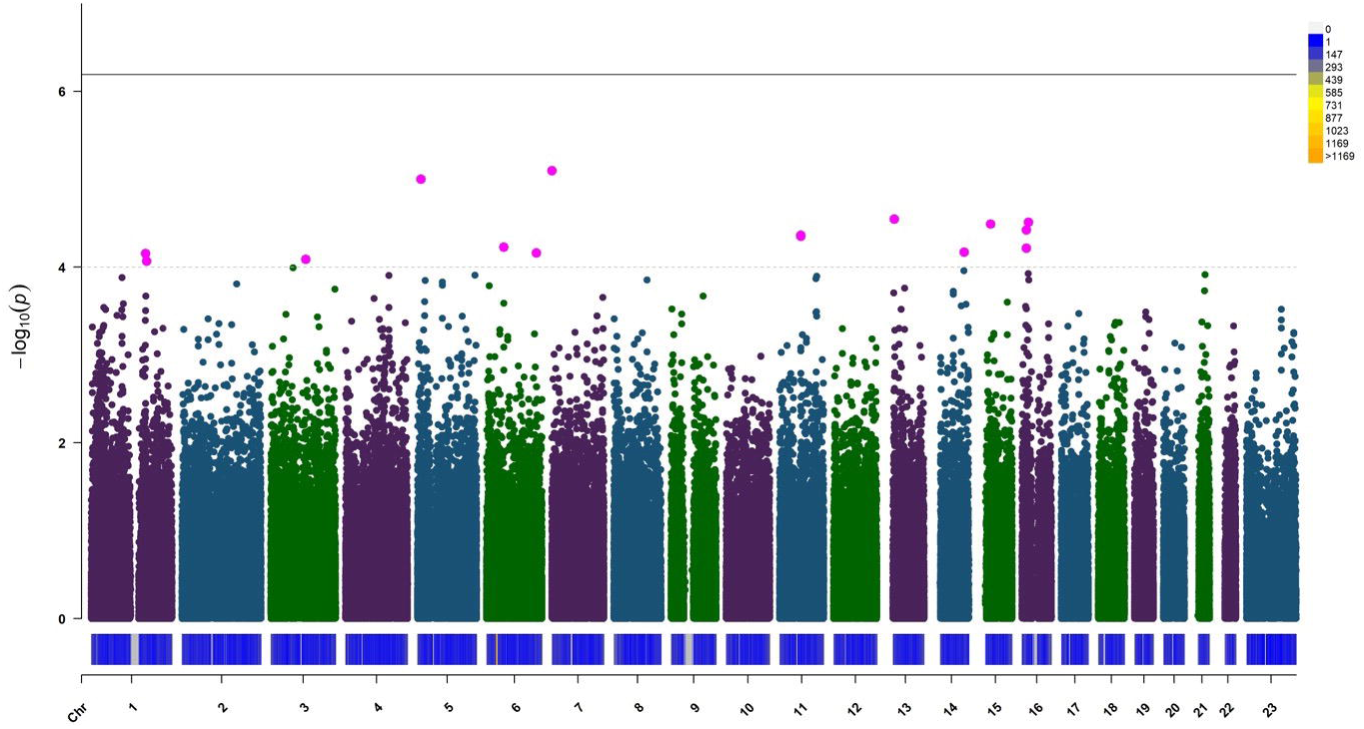
Genome-wide association study (GWAS) for cognitive preservation. The suggestive association threshold (p=1x10^-4^) is depicted with a dashed line; the significant association threshold (p=6.4x10^-7^) was defined using the simple-M method and is depicted with a solid line. SNPs are represented as dots corresponding to genomic coordinates within their chromosome (horizontal axis). SNP density within each chromosome is shown as a heat map above each chromosome number. Chromosome 23 is the X chromosome. Eleven loci demonstrated suggestive association, located on chromosomes 1, 3, 5, 6, 7, 11, 13, 14, 15, and 16 (and depicted as pink dots on this figure). No SNP surpassed the significance threshold for association.

No SNPs on the X chromosome were significant or suggestive under either sex-stratified model (Figure 2). The genomic inflation value was 0.98 for the X chromosome (Supplementary Figure 2B).

### 3.2 Linkage Analyses for Cognitive Preservation

Pedigrees were grouped via bit-size, chosen based on computational tractability for our analyses. For NPL analyses, families of bit size 21 and 22 were examined (n = 201). In the two-point NPL analyses, 17 loci on 13 chromosomes surpassed the threshold for suggestive linkage (≥ 1.86) (Supplementary Figure 3A). Multipoint NPL analyses resulted in 10 suggestive loci on 10 different chromosomes (Supplementary Figure 3B). No SNPs reached statistical significance in any NPL analysis.

Two-point dominant linkage analyses generated 72 significant (≥ 3.3) HLOD results spread across most chromosomes for both the 21- and 22-bit pedigrees (Figure 3 and Supplementary Figure 4A). In multipoint analyses, only chromosomes 12 and 17 had significant results for the dominant model (Supplementary Figure 4B). Two-point recessive linkage analyses generated significant results on 11 different chromosomes (Supplementary Figure 4C). No loci reached significance with a multipoint recessive model (Supplementary Figure 4D). Two-point parametric linkage analysis on the X chromosome led to 14 loci surpassing the threshold for suggestive linkage, but no loci were significant. No SNPs were seen as suggestive or significant in multipoint analyses on the X chromosome. See Supplemental Tables 3 and 4 for a full list of significant and suggestive loci, respectively, from linkage analyses.

**Figure 3.**
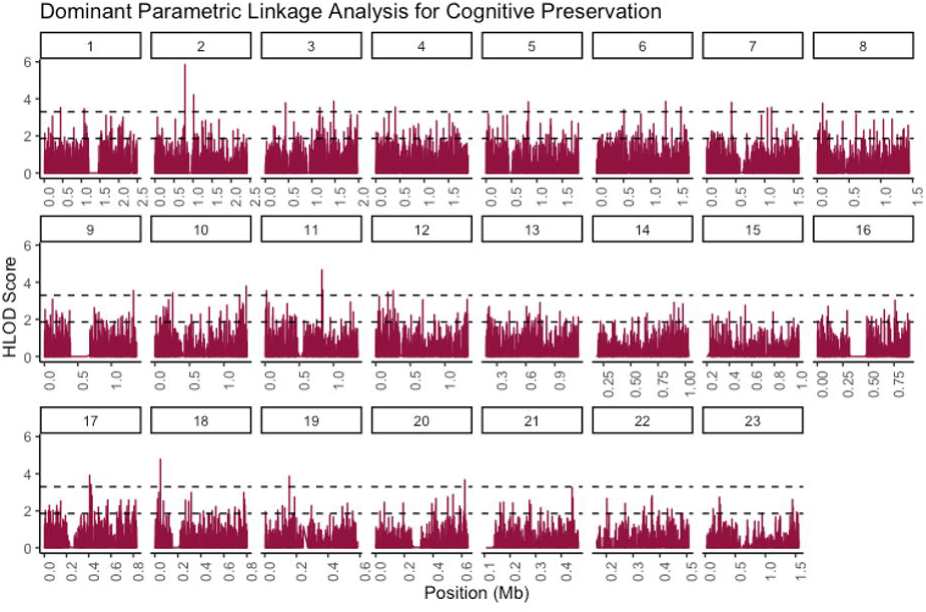
Dominant two-point parametric linkage analysis for cognitive preservation. Dominant linkage analysis on 21-bit pedigrees across all autosomes and the X chromosome, labeled as chromosome 23. Suggestive (LOD ≥ 1.86) and significant (LOD ≥ 3.3) thresholds are denoted as the lower and upper dashed horizontal lines, respectively. A total of 72 significant results were seen across most chromosomes (excluding chromosomes 14, 16, 22, and 23). See Supplemental Figure 3 and Supplemental Tables 4 and 4 for the complete list of all loci surpassing significant and suggestive thresholds.

### 3.3 Assessment of Significant and Suggestive Genomic Regions

The need to use sub-pedigrees for linkage analysis introduced an unknown amount of variability into the linkage analysis calculations.[32] To address this issue, we implemented an approach to filter the large number of loci surpassing thresholds for suggestive or significant results in GWAS and/or linkage. Some differences were seen across the various bit size pedigrees, therefore, significant HLOD scores in multiple linkage analyses in the same region were a requirement for the locus to undergo further investigation. Regions selected for follow-up were required to have 1) two significant HLOD scores (≥3.3) in any two linkage analyses and a significant or suggestive LOD score (≥1.86) in at least one other linkage analysis; or 2) one suggestive result in the GWAS (within 5 Mb), a significant HLOD score in any linkage analysis, <u>and</u> a suggestive HLOD score in any other linkage analysis. After applying these criteria, 6 regions of interest were identified for further investigation, located on chromosomes 1, 2, 3, 7, 11, and 17 (Table 1). When probing within a 10 Mb region around each of these loci, we observed two known AD-related genes (adsp.niagads.org), *PICALM* (chromosome 11) and *EPHA1* (chromosome 7).

**Table 1.** Loci meeting *ad-hoc* filtering conditions for further investigation. Chromosome number and coordinates are listed for each of the top LOD scores. P-value is listed for suggestive GWAS result near the same location. For 21- and 22-bit pedigree analyses, significant (≥ 3.3) and suggestive (≥ 1.86) HLOD scores are listed, with significant results in bold. Empty cells indicate that no loci surpassed the suggestive or significant threshold for that analysis.

| Chr | Location | GWAS | 21-bit pedigrees |  |  |  | 22-bit pedigrees |  |  |  |
| --- | --- | --- | --- | --- | --- | --- | --- | --- | --- | --- |
|  |  | p-value | Two-point NPL | Two-point Dominant | Two-point Recessive | Multipoint Dominant | Two-point NPL | Two-point Dominant | Two-point Recessive | Multipoint Dominant |
| 1 | 165,478,920 | 7.00E-05 | 1.9 |  | 3.4 |  |  |  | 3.2 |  |
| 2 | 78,873,098 |  | 2.4 | 5.9 | 4.6 |  | 1.9 | 4.9 | 3 |  |
| 3 | 108,251,237 | 8.20E-05 |  | 3.3 |  |  | 2 | 3.6 |  |  |
| 7 | 139,996,449 |  | 2.1 | 2.4 | 4.1 |  |  |  | 3.8 |  |
| 11 | 83,081,483 |  | 2 | 4.7 | 2.9 |  |  |  | 3.3 |  |
| 17 | 4,487,906 |  |  | 2.3 |  | 2.3 |  | 4.1 |  | 4.1 |

By examining which sub-pedigrees displayed the highest chromosome-specific LOD scores across multiple analyses, it was noted that some of these sub-pedigrees could be connected either by a specific individual or through a first degree relative. These related sub-pedigrees were combined into larger family pedigrees for five of the loci (chromosomes 1, 2, 7, 11, and 17), but were unable to be merged for the chromosome 3 locus. These five larger pedigrees were then used for the MCMC linkage analyses.

Only the chromosome 2 locus, centered around SNP rs1402906, remained significant (HLOD = 4.87) after connection of the sub-pedigrees. MCMC analyses with identical parameters were performed to further verify these results, this time adding additional SNPs upstream and downstream of rs1402906 to widen the genomic window considering potential linkage. These additional analyses each considered 5 SNPs, and again rs1402906 retained the highest significance with a dominant model (LOD=4.14) (see Table 2 and Figure 4). The rs1402906 T allele frequency is 12% in the Amish, compared to 19% in the ALFA European population (dbSNP).[33] Of the 24 CU individuals who had genotype data available, the genotype counts for rs1402906 were 7 TT, 12 CT, and 3 CC (Supplementary Table 6). These CU individuals in the chromosome 2-linked pedigree were all under 88 years of age. There were two CI individuals in the chromosome 2-linked pedigree, both displayed the CC genotype at rs1402906 and were >89 years. Despite the co-segregation of the T allele with most CU individuals in this pedigree, the genotype frequencies are essentially the same between CU and CI individuals in the entire Amish population in the study (Supplementary Table 6).

**Figure 4.**
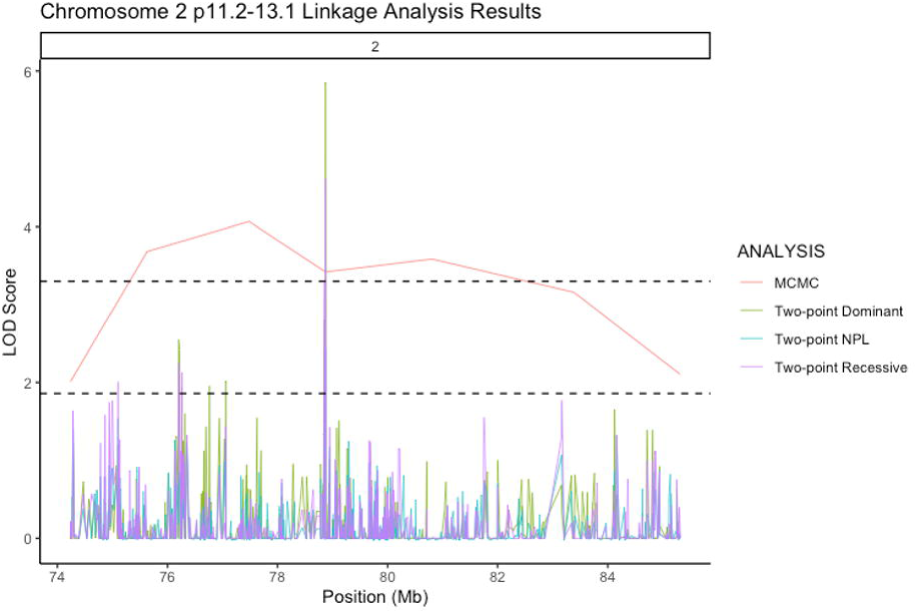
Chromosome 2p11.2-13.1 linkage analysis results. Two-point dominant (green) and recessive (purple) linkage results, two-point NPL (blue) and multipoint MCMC (orange) (significance >3.3, upper dashed line; suggestive >1.86, lower dashed line).

**Table 2.** MCMC linkage analysis for cognitive preservation. Chromosome, genomic coordinates, and LOD score are depicted in the table. LOD scores in bold surpass

| Chromosome | Location (bp) GRCh38 | Top LOD Score |
| --- | --- | --- |
| 1 | 159,476,580 – 167,672,055 | 1.97 |
| 2 | 72,972,490 – 80,875,139 | **4.14** |
| 7 | 138,047,463 – 141,972,804 | 0.6 |
| 11 | 81,074,649 – 85,224,030 | 2.64 |
| 17 | 3,492,832 – 5,478,547 | 0.13 |

### 3.4 Chromosome 2p *in Silico* Annotation

The UCSC genome browser was used to identify protein coding genes on chromosome 2p11.2-13.1 spanning 10 cM around the peak LOD score at rs1402906.[34] The protein coding genes in this region include *REG1A*, *REG1B*, *CTNNA2*, and *LRRTM1* and *LRRTM4* (Figure 5). In addition, the Open Targets Platform eQTL and PheWAS databases specified an eQTL associated with decreased *CTNNA2* expression with the “T” allele at rs1402906, as well as an increased incidence within a study population of individuals with autism.[35,36] The PheWAS database also indicated a miniscule, but still significant, diminishment within a study population of Bipolar individuals.

**Figure 5.**
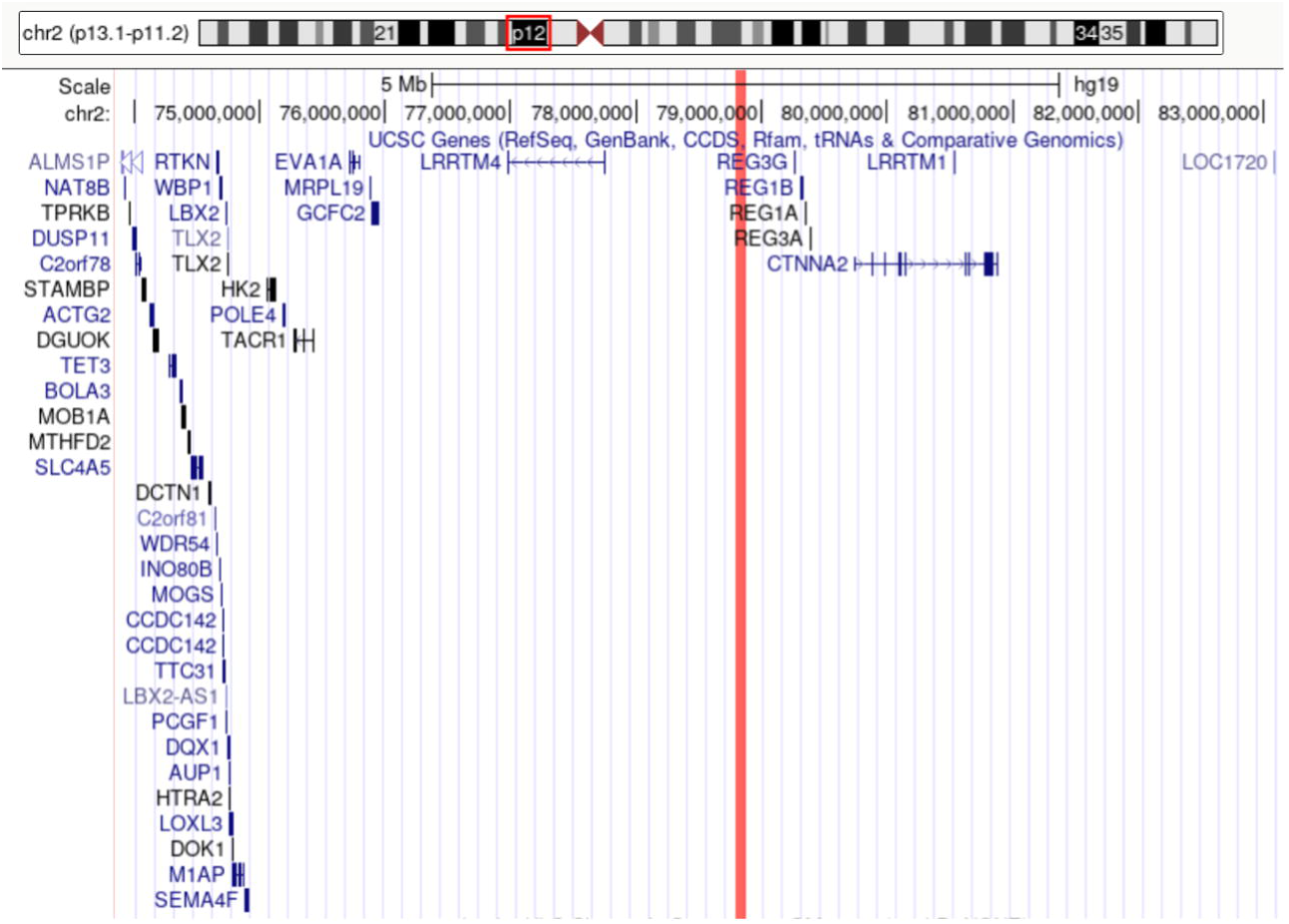
UCSC Genome browser for chromosome 2p11.2-13.1. UCSC Genome Browser tracks of protein coding genes in the significant linkage region on chromosome 2 p11.2-13.1. The observed chromosomal region is indicated by a red box on the genomic coordinate slider. rs1402906 is located at 79Mb, highlighted by a vertical red line across the viewer, with 5Mb on either side shown. This region is located upstream of the genes *CTNNA2* and *LRRTM1*.

Sequences around the SNP demonstrating the peak LOD score, rs1402906, were investigated for potential promoter regions in the Eukaryotic Promoter Database (EPD)[37], as well as analyzed through PROMO, a transcription factor binding site (TFBS) identification tool developed by ALGGEN.[38,39] Cap Analysis of Gene Expression (CAGE) reads from the FANTOM5 project were also examined to investigate transcription start sites (TSS).[40]

Although the 2p11.2-13.1 genomic region is relatively devoid of known genes, examination of transcription factor binding sites in this region via ALGGEN shows the transcription factor POU3F2 as having a perfect binding site consensus beginning 4bp upstream of rs1402906 when the genomic sequence contained the T allele at rs1402906.[41] Substitution of the alternative allele C nearly abolished the binding potential for POU3F2, with a low predicted binding score due to two base-pair mismatches in the recognition sequence. Another transcription factor, POU1F1a, demonstrates a perfect binding consensus 10bp upstream of rs1402906. However, the alleles at rs1402906 do not disrupt the binding. Utilizing the EPD as well as CAGE reads uncovered HK3me1, HK3me3, and H3K27ac sites within 10bp of rs1402906, indicating a potential TSS as well as predicted enhancer activity.[42,43]

## 4 DISCUSSION

Identifying protective genetic loci as a strategy for finding potential therapeutic targets is a methodology that yields higher success rates for clinical trials than other approaches.[6] For dementia, protective loci are associated with an outcome of cognitive preservation, which can result from either cognitive resilience or resistance. Cognitive resilience is observed when patients display typical dementia pathology, specifically a buildup of amyloid plaques and tangles in the brain in AD, without demonstrating any cognitive impairment or other dementia symptoms.[44] Cognitive resistance is defined by a lack of dementia-associated symptoms without presenting dementia pathologies.[44] As the Amish undergo only pre-mortem diagnosis, their classification falls under cognitive preservation, which is either cognitive resilient or resistant. Previous studies for AD have identified multiple genes that possess alleles that appear to confer some protection against developing AD including, *APP*[45]*, APOE* LJ*2*[46]*, PLCG2*[47]*, MS4A*[48]*, ABCA1*[49]*, RAB10*[50]*, SORL1*[51]*, PICALM*[52]*, EPHA1*[4], and *CASS4*[4]. Although our *ad hoc* filtering suggested possible effects near *PICALM* and *EPHA1,* no SNPs in either gene demonstrated a protective effect.

GWAS and linkage analyses identified 106 suggestive or significant loci for cognitive preservation in the Amish (Supplemental Table 3). To limit false positives, we focused only on areas yielding suggestive or significant associations in multiple analyses. This limited our results to six areas of interest across chromosomes 1, 2, 3, 7, 11, and 17 (Table 1 and Table 2). Notably, an effect of *APOE* LJ*2* was not detected in any analysis, however this is likely because of APOE LJ*3’s* high allele frequency within the Amish and its neutral phenotypic outcome, thereby preventing its enrichment in either the case or control groups. *APOE* LJ*2* was infrequent in this Amish population, with an allele frequency of just 4.2%, compared to 19.5% in non-Hispanic white populations,[53] making it likely too rare for analyses to detect. A region on chromosome 2p11.2-13.1, centered around rs1402906, demonstrated statistical significance consistently across multiple linkage analyses. By connecting several of the sub-pedigrees, we confirmed the transmission, providing additional evidence of linkage.

Because significant allelic association or genetic linkage results do not necessarily identify causal variations, a 10 Mb region around the chromosome 2 locus peak was evaluated to develop a broader assessment of possible functional elements. While this region was not accompanied by association in the GWAS, PROMO predicted a POU3F2 TFBS with perfect consensus beginning 4bp upstream of rs1402906 when the genomic sequence contained the T allele. Though there remained a possibility of binding in the presence of the C allele at that site, consensus was markedly diminished as any potential binding would require only partial adherence to the site, with two of the seven base pairs in the binding locus being mismatched.[41] A majority (87%) of the individuals demonstrating cognitive preservation in the chromosome 2-linked pedigree (Figure 1C and Figure 4) have this T allele. Of the three CC-genotyped CU individuals, two married into the pedigree (i.e., were not otherwise closely related to the other pedigree members), with the third CC genotyped individual (age 80) being below the average age of onset for CI in the Amish population (age 82) and a child of one of the married-ins. Additionally, the average age of onset for CI was 89.5 in this chromosome 2-linked pedigree used for MCMC analysis, implying that these individuals may still be too young to display CI onset or may have other protective factors located elsewhere in the genome as the average age of onset for this family is older than the general Amish population.

POU3F2 is a known regulator of the nearby gene *Leucine Rich Repeat Transmembrane neuronal 4* (*LRRTM4*).[54] The leucine-rich repeat transmembrane proteins (LRRTMs) are primarily expressed in the nervous system where they assist in synaptic differentiation and development.[55,56] Murine *in vivo* studies demonstrated that reduced *LRRTM4* expression led to deficits in excitatory synapse density in dentate gyrus granule cells.[57] Increased expression of *LRRTM4* in the hippocampus improves memory in aging rats.[58] Clinically, *LRRTM4* has been associated with Tourette syndrome and autism spectrum disorder, as well as increased risk of suicide attempt in bipolar disorder.[59,60] It is possible that altered transcription in an upstream regulatory region, possibly due to the potential for POU3F2 binding, may play a role in expression differences of *LRRTM4*, which could further contribute to improved cognitive ability in CU patients, as observed in rodent models.

In eQTL and PheWAS databases searched, the “T” allele at rs1402906 was associated with increased incidence of autism spectrum disorder as well as lower *CTNNA2* expression. Notably, the POU1F1a TF also had perfect consensus binding in the region containing rs1402906 and POU1F1a regulates the *CTNNA2* gene[61]. POU1F1a overexpression induces increased expression of *Leucine Rich Repeat Transmembrane neuronal 1* (*LRRTM1*),[61] a gene 312 kb away from rs1402906 and within the region of significant linkage on 2p11.2-13.1. *LRRTM1* is highly expressed in the brain and is associated with handedness and schizophrenia in humans.[62] Deletions of this gene lead to hippocampal shrinking, loss of synaptic density, reduced long-term potentiation in hippocampal neurons, and memory deficiencies in mice.[63,64]

*LRRTM1* resides within the seventh intron of the *Catenin Alpha 2* (*CTNNA2*) gene in an anti-sense orientation, 262kb away from rs1402906. *CTNNA2* shares promoters with *LRRTM1*.[65] These shared promoters induce alternative transcripts of *CTNNA2* containing additional 5’ exons expressed almost exclusively in the brain. Importantly, these promoters are bi-directional and may regulate both *LRRTM1* and the alternative splicing of *CTNNA2* as a single functional unit.[66] Exon VIIb of *CTNNA2*, located on the sense strand corresponding to the *LRRM1* location, is included more frequently in transcription than other alternative exons, especially in the brain. The ratio of expression levels of full-length *CTNNA2* and this isoform may be important for regulating cadherin-dependent adhesion.[66] We note that a previous publication from our group using a subset of our Amish dataset found linkage to dementia in the region surrounding *CTNNA2* and suggested it as a potential risk locus for dementia.[32] Here, the “T” allele at rs1402906, linked to cognitive preservation, was an eQTL noted to decrease *CTNNA2* expression.

*CTNNA2* is highly expressed in the brain in both mice and humans.[32,67] It is well categorized in mice, where it regulates function during murine neuronal development, specifically synapse formation and plasticity.[67] Mechanistically, *CTNNA2* regulates synaptic spine formation, stabilization, and turnover.[68] *CTNNA2* also forms a complex with beta-catenin, which interacts with presenilin, a known contributor to AD pathology. Destabilization of this beta-catenin complex can lead to neuronal apoptosis in mice and humans.[69] The location of potential POU3F2 and POU1F1a binding sites at rs1402906, near HK3me1, HK3me3, and H3K27ac sites indicate predicted enhancer activity nearby a TSS. Thus, POU1F1a binding may initiate transcription of either *LRRTM1*, an alternative splicing variant of *CTNNA2*, or both, as transcription of these genes has been shown to be coregulated, with multiple TSSs for each gene existing within 30 bps of each other.[66] Similarly, POU3F2 binding may initiate altered transcription of *LRRTM4*, another gene involved in neurocognition.[58]

Our analyses highlight the genetic complexity of cognitive preservation in the Amish, with different sub-pedigrees appearing to drive disparate regions of the genome associated with cognitive preservation. Numerous regions yielded significant or suggestive results, but a region on chromosome 2p11.2-13.1 showed the most robust linkage in an area previously associated with dementia in the Amish, suggesting the possibility of both protective and risk regulatory elements for cognition in this region of the genome. These results lay the foundation for additional studies to identify loci conferring protection against cognitive impairment.

## Data Availability

Data produced in the present work are contained in the manuscript or available upon reasonable request to the authors, barring inquiries impinging upon the privacy of individuals in our dataset.

## ABBREVIATIONS

AD: Alzheimer Disease
AGDB: Anabaptist Genealogical Database
CI: Cognitively impaired
CU: Cognitively unimpaired
GWAS: Genome-wide association study
HLOD: Heterogeneity logarithm of the odds
LD: Linkage disequilibrium
LOD: Logarithm of the odds
MCI: Mild cognitive impairment
MCMC: Markov chain Monte Carlo
QC: Quality control

## 6 AUTHOR CONTRIBUTIONS

Jonathan L Haines, William K Scott, and Margaret A Pericak-Vance helped design the study and obtain funding for this research. Ascertainment of data was collected and managed by Renee A Laux, Kristy L Miskimen, M Denise Fuzzell, Sarada L Fuzzell, Sherri D Hochstetler, Laura J Caywood, Jason E Clouse, and Sharlene D Herington. Cognitive status was determined by Michael L Cuccaro, Paula K Ogrocki, Alan J Lerner, and Jeffery M Vance. Yeunjoo E Song, Leighanne R Main, Michael D Osterman, Audrey Lynn, Daniel A Dorfsman, Larry D Adams, and Michael B Prough provided data management, organization, and QC. Analyses were performed by Leighanne R Main, with assistance from Yeunjoo Song and Michael D. Osterman. Manuscript writing and drafting was performed by Jonathan L Haines, Audrey Lynn, and Leighanne R Main. Editing completed by all authors.

## 7 ACKNOWLEDGMENTS

This research was supported under the National institutes of Health and National Institute on Aging, grant AG058066. This research was also supported by an Alzheimer’s Disease Translational Data Science Training Grant, 5T32AG071474-02.

## 9 CONSENT STATEMENT

All individuals ascertained in this study supplied written informed consent under protocols approved by the IRBs at Case Western Reserve University and the University of Miami.

## 10 Diversity, Equity, and Inclusion Statement

Our study considers an isolated founder population, not only genetically, but also culturally, as their lifestyles deviate from modern technocratic habits, making the Amish an under-represented population in studies. Additionally, informed consent and community engagement informed our sampling practices and subsequent analyses. We also made efforts to ensure our study cohort consisted of comparable proportions across each sex.

