## Supplemental Figure 1 for "Genetic analysis of cognitive preservation in the midwestern Amish reveals a novel locus on chromosome 2"

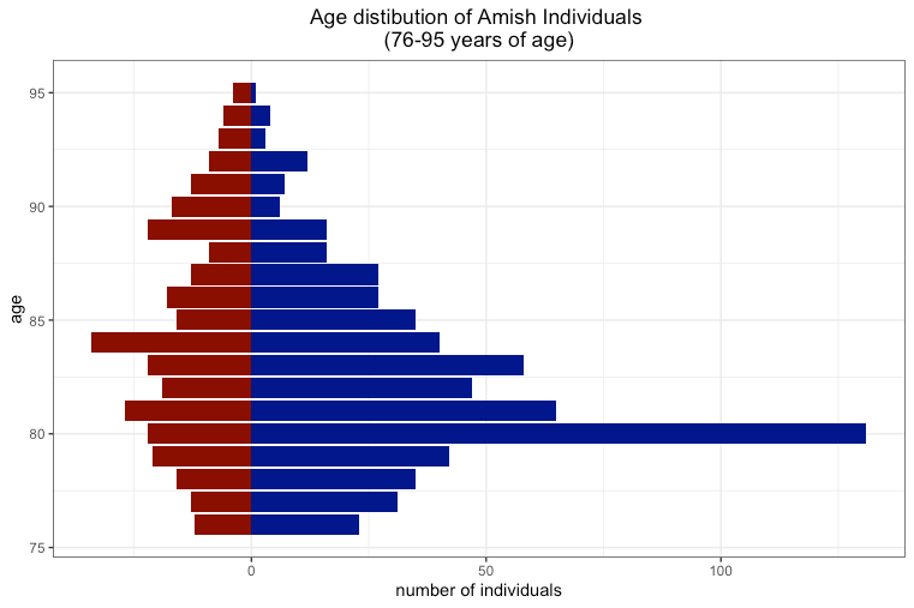

### SUPPLEMENTARY FIGURE 1 (Leighanne Main)

**Supplementary Figure 1 – Age distribution of Amish individuals.** Ages for cognitively impaired individuals (CI) are displayed in red and cognitively unimpaired individuals (CU) in blue. Age is on the y-axis and sample size is on the x-axis. Our study was limited to individuals aged 76-95. The mean age for both CI and CU groups was 82 years.
