## Supplemental Figure 2 for "Genetic analysis of cognitive preservation in the midwestern Amish reveals a novel locus on chromosome 2"

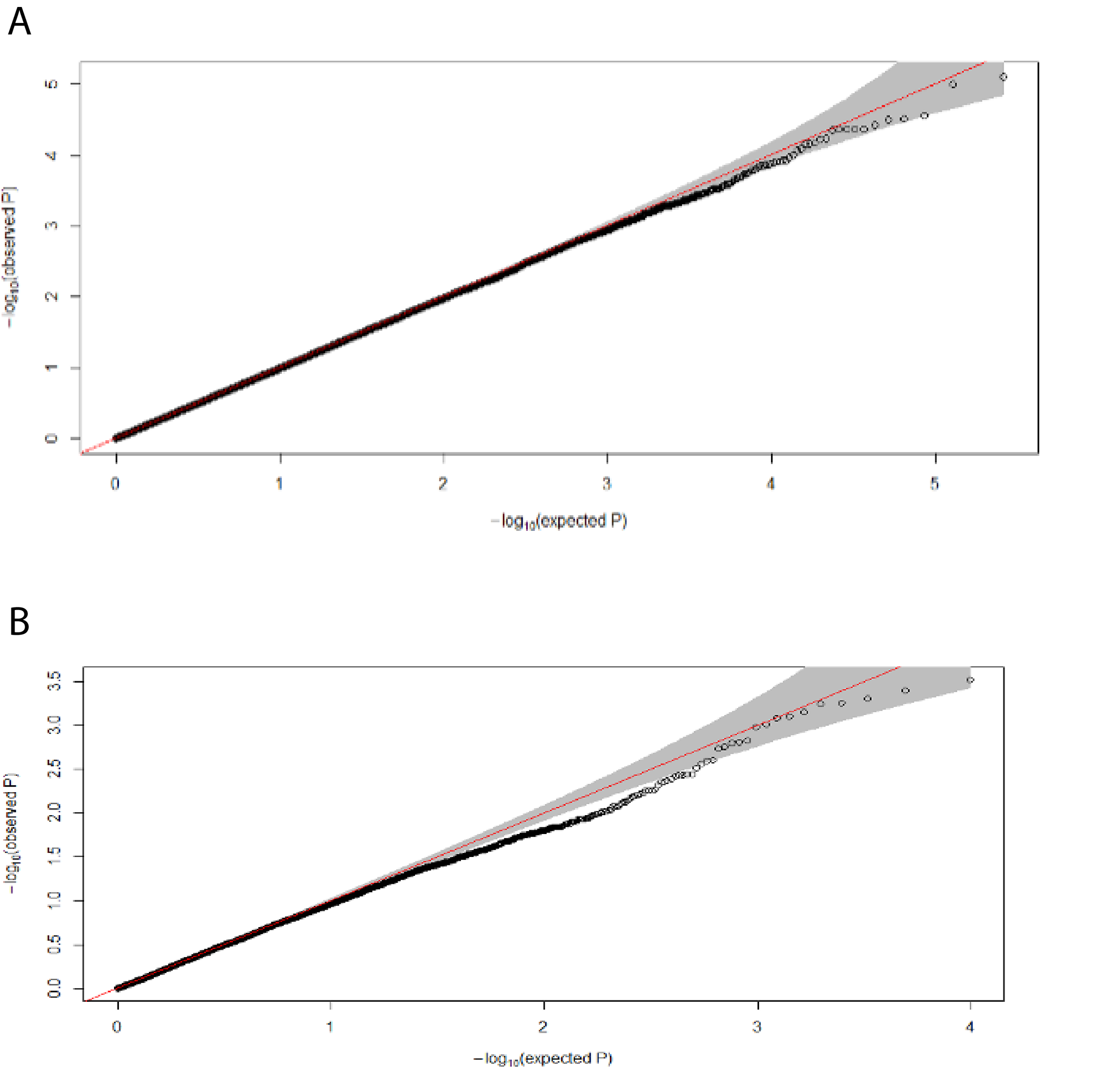

SUPPLEMENTARY FIGURE 2 (Leighanne Main)

**Supplementary Figure 2 – Quantile-quantile (QQ) plot of GWAS results.** (A) QQ plot for autosomal association. The genomic inflation (lambda) was 0.98 after adjusting for the GRM (which considers PC1 and PC2) as well as sex and age. (B) QQ plot for X chromosomal association. The genomic inflation (lambda) was 0.98 after adjusting for age and PCs 1-5. This sex-stratified analysis was run with males coded as both homozygotes and heterozygotes for the X chromosome, with no change in genomic inflation.
