## Supplemental Figure 3 for "Genetic analysis of cognitive preservation in the midwestern Amish reveals a novel locus on chromosome 2"

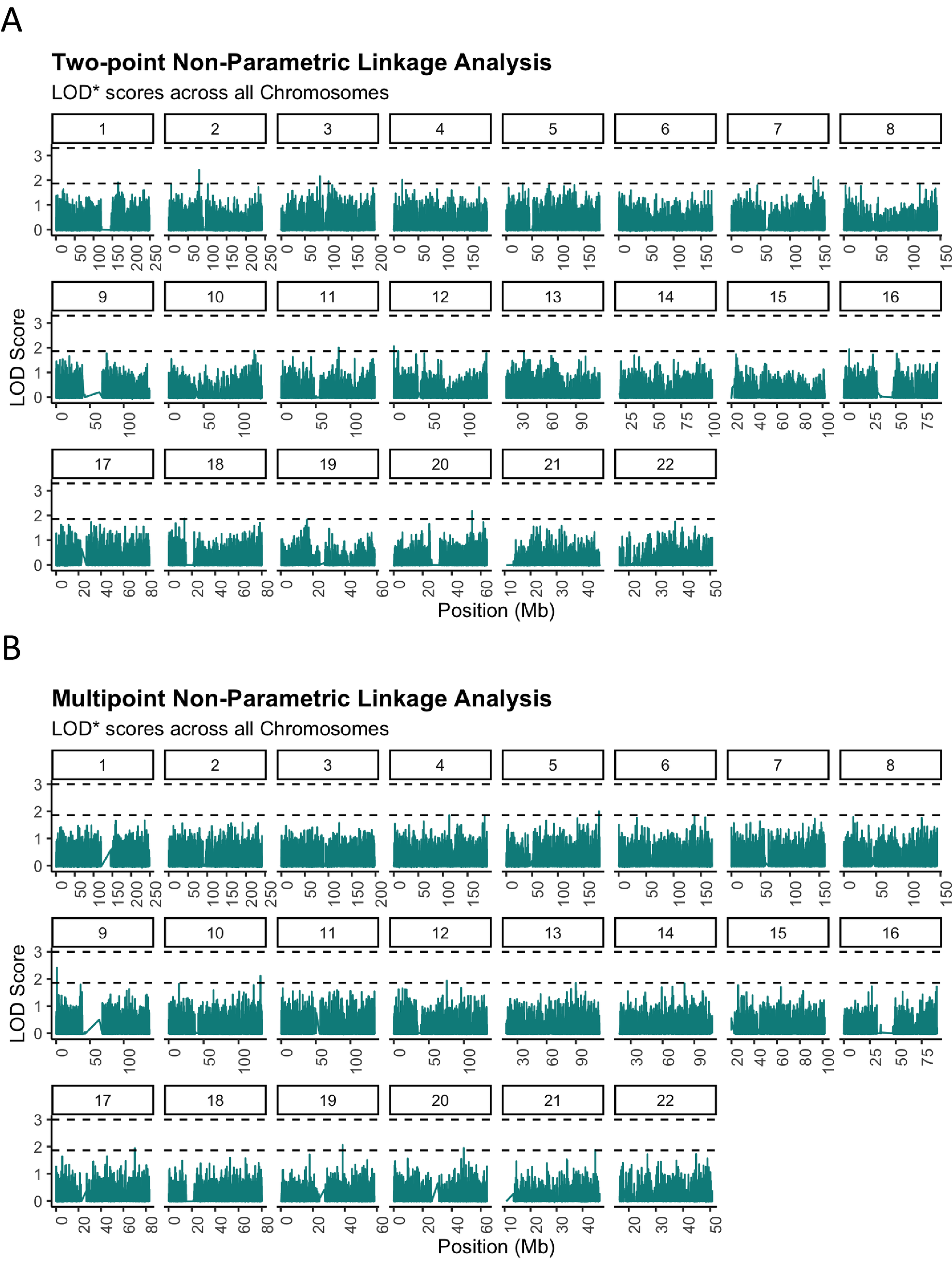

SUPPLEMENTARY FIGURE 3 (Leighanne Main)

**Supplementary Figure 3 –** **Non-parametric linkage analysis for cognitive preservation.** Two-point (A) and multipoint (B) non-parametric linkage analyses (NPL) was performed using MERLIN software. Chromosomes are labeled in boxes above each plot, with thresholds for suggestive (LOD* ≥ 1.86) and significant linkage (LOD* ≥ 3.3) represented as lower and upper dashed lines, respectively. (A) Two-point analyses resulted in suggestive loci on chromosomes 1, 2, 3, 4, 5, 7, 8, 10, 11, 12, 16, 18, and 20. (B) Multipoint analyses resulted in suggestive loci on chromosomes 4, 5, 9, 10, 12, 13, 17, 19, 20, and 21. No SNPs surpassed the threshold for significant linkage in either two-point or multipoint NPL analyses.
