## Supplemental Figure 4 for "Genetic analysis of cognitive preservation in the midwestern Amish reveals a novel locus on chromosome 2"

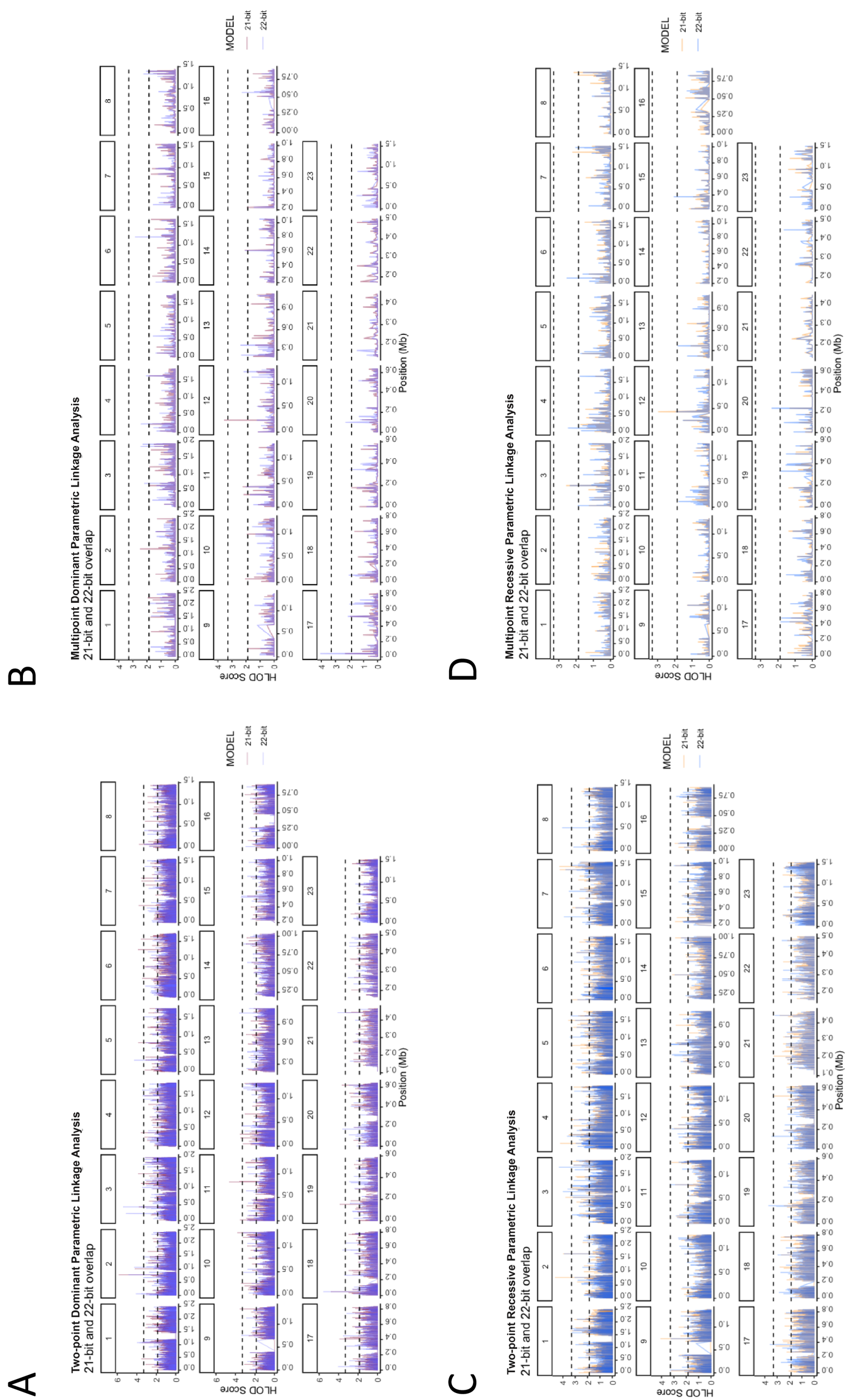

SUPPLEMENTARY FIGURE 4 (Leighanne Main)

**Supplementary Figure 4 –** **Parametric linkage analysis for cognitive preservation.** Dominant (A and B) and Recessive (C and D) linkage analyses; Two-point (A and C) and Multipoint (B and D) linkage analyses. Chromosomes are labeled in boxes above each plot, with the X chromosome labeled as chromosome 23. Suggestive (LOD ≥ 1.86) and significant (LOD ≥ 3.3) thresholds are denoted as the lower and upper dashed horizontal lines, respectively. Pedigree sizes are coded as blue for analyses of 22-bit size pedigrees, and as red or yellow for 21-bit size pedigrees (dominant or recessive, respectively). (A) Two-point dominant analyses resulted in 72 significant results across all chromosomes except chromosomes 14, 16, and 22. (B) Multipoint dominant analyses resulted in significant loci on chromosomes 12 and 17. (C) Two-point recessive analyses led to significant loci on chromosomes 1-5, 7-9, 11, and 17. (D) Multipoint recessive analyses generated no significant loci. See Supplemental Tables 3 and 4 for a complete list of all loci surpassing significant or suggestive thresholds.
