## Supplemental Table 1 for "Genetic analysis of cognitive preservation in the midwestern Amish reveals a novel locus on chromosome 2"

| Cognitive Status by Sex | | | |
| --- | --- | --- | --- |
| Sex | CU | CI | Total |
| Male | 242 (62.9%) | 143 (37.1%) | 385 (40.7%) |
| Female | 384 (68.4%) | 177 (31.6%) | 561 (59.3%) |
| Total | 626 (66.2%) | 320 (33.8%) | 946 (100%) |

SUPPLEMENTARY TABLE 1 (Leighanne Main)

**Supplementary Table 1 – Cognitive status by sex.** Our sample was largely female and unimpaired.
