## Supplemental Table 2 for "Genetic analysis of cognitive preservation in the midwestern Amish reveals a novel locus on chromosome 2"

| Sub-pedigree set | Number of Pedigrees | CU individuals per sub-pedigree | | CI individuals per sub-pedigree | |
| --- | --- | --- | --- | --- | --- |
|  |  | Mean | Range | Mean | Range |
| 21-bit | 103 | 6 | 10-Mar | 0 | 0-2 |
| 22-bit | 98 | 6 | 10-Mar | 0 | 0-2 |
| 23-bit | 94 | 7 | 11-Mar | 0 | 0-2 |
| Merged pedigrees for MCMC | 5 | 27 | 22-35 | 0 | 0-2 |

SUPPLEMENTARY TABLE 2 (Leighanne Main)

**Supplementary Table 2 – Pedigree subsets in linkage analyses.** Descriptive statistics provided for each subset of pedigrees generated by PedCut for linkage analyses.
