## Supplemental Table 3 for "Genetic analysis of cognitive preservation in the midwestern Amish reveals a novel locus on chromosome 2"

| Chrom | Location | SNP | p-value |
| --- | --- | --- | --- |
| 1 | 166,737,906 | GSA-rs487726 | 7.00E-05 |
| 1 | 170,646,692 | rs12129225 | 8.58E-05 |
| 3 | 106,423,768 | JHU_3.106142614 | 8.17E-05 |
| 5 | 10,303,626 | rs13177199 | 9.99E-06 |
| 6 | 53,111,161 | rs80240944 | 5.93E-05 |
| 6 | 153,208,028 | rs685341 | 6.92E-05 |
| 7 | 302,511 | rs140733969 | 7.96E-06 |
| 11 | 64,725,290 | 11:64492762-A-C | 4.35E-05 |
| 11 | 64,728,885 | exm2250129 | 4.35E-05 |
| 11 | 64,757,744 | rs589691 | 4.35E-05 |
| 11 | 64,765,107 | exm2259835 | 4.35E-05 |
| 11 | 64,778,919 | rs606458 | 4.46E-05 |
| 13 | 23,216,503 | rs681938 | 2.83E-05 |
| 14 | 92,320,706 | rs4900111 | 6.77E-05 |
| 15 | 36,056,972 | rs1347455 | 3.24E-05 |
| 16 | 13,370,376 | rs2023768 | 6.12E-05 |
| 16 | 13,370,634 | GSA-rs886218 | 3.80E-05 |
| 16 | 19,926,831 | rs12596728 | 3.08E-05 |

SUPPLEMENTARY TABLE 3 (Leighanne Main)

**Supplementary Table 3 – GWAS results surpassing the suggestive threshold.** 18 SNPs surpassed the suggestive threshold (p<10^-4^). Listed for each SNP are chromosome number, genomic coordinates (hg38), SNP identifier, and p-value.
