## Supplemental Table 4 for "Genetic analysis of cognitive preservation in the midwestern Amish reveals a novel locus on chromosome 2"

| CHR | POS | SNP | LOD | ANALYSIS |
| --- | --- | --- | --- | --- |
| 1 | 44,133,088.00 | rs2254746 | 3.5271 | 21_bit_twopoint_parametric_dominant |
| 1 | 107,721,589.00 | rs12059570 | 3.7129 | 22_bit_twopoint_parametric_dominant |
| 1 | 107,744,349.00 | GSA-rs6583045 | 3.4594 | 21_bit_twopoint_parametric_dominant |
| 1 | 107,744,349.00 | GSA-rs6583045 | 3.9667 | 22_bit_twopoint_parametric_dominant |
| 1 | 110,827,750.00 | rs343769 | 3.3912 | 22_bit_twopoint_parametric_dominant |
| 1 | 110,827,750.00 | rs343769 | 3.3912 | 22_bit_twopoint_parametric_dominant |
| 1 | 165,478,920.00 | GSA-rs10918196 | 3.3757 | 21_bit_twopoint_parametric_recessive |
| 1 | 173,492,069.00 | JHU_1.173461207 | 3.3636 | 22_bit_twopoint_parametric_recessive |
| 2 | 6,541,346.00 | rs17802554 | 4.0966 | 22_bit_twopoint_parametric_dominant |
| 2 | 6,589,575.00 | rs17803843 | 3.4924 | 22_bit_twopoint_parametric_dominant |
| 2 | 68,395,858.00 | rs6725743 | 3.3332 | 22_bit_twopoint_parametric_dominant |
| 2 | 68,395,858.00 | rs6725743 | 3.3332 | 22_bit_twopoint_parametric_dominant |
| 2 | 78,873,098.00 | rs1402906 | 4.6123 | 21_bit_twopoint_parametric_recessive |
| 2 | 78,873,098.00 | rs1402906 | 4.8588 | 22_bit_twopoint_parametric_dominant |
| 2 | 78,873,098.00 | rs1402906 | 5.8538 | 21_bit_twopoint_parametric_dominant |
| 2 | 101,617,269.00 | rs6543081 | 4.2113 | 21_bit_twopoint_parametric_dominant |
| 2 | 107,806,354.00 | rs4972265 | 4.2523 | 22_bit_twopoint_parametric_dominant |
| 2 | 147,380,514.00 | exm2261081 | 3.3321 | 22_bit_twopoint_parametric_dominant |
| 2 | 147,380,514.00 | exm2261081 | 3.3321 | 22_bit_twopoint_parametric_dominant |
| 2 | 170,217,377.00 | rs2161916 | 3.8849 | 22_bit_twopoint_parametric_recessive |
| 2 | 170,217,377.00 | rs2161916 | 3.956 | 21_bit_twopoint_parametric_recessive |
| 3 | 22,990,734.00 | rs680110 | 3.6268 | 22_bit_twopoint_parametric_dominant |
| 3 | 23,013,017.00 | GSA-rs537660 | 5.096 | 22_bit_twopoint_parametric_dominant |
| 3 | 23,086,636.00 | JHU_3.23128126 | 4.3551 | 22_bit_twopoint_parametric_dominant |
| 3 | 23,105,281.00 | GSA-rs6807204 | 4.1879 | 22_bit_twopoint_parametric_dominant |
| 3 | 23,111,582.00 | rs6550726 | 4.8556 | 22_bit_twopoint_parametric_dominant |
| 3 | 43,676,038.00 | rs4682900 | 3.772 | 21_bit_twopoint_parametric_dominant |
| 3 | 43,676,038.00 | rs4682900 | 3.772 | 21_bit_twopoint_parametric_dominant |
| 3 | 43,676,038.00 | rs4682900 | 5.3929 | 22_bit_twopoint_parametric_dominant |
| 3 | 43,728,831.00 | GSA-rs3733155 | 3.5632 | 22_bit_twopoint_parametric_dominant |
| 3 | 82,190,248.00 | rs4561854 | 3.7613 | 21_bit_twopoint_parametric_recessive |
| 3 | 82,190,248.00 | rs4561854 | 3.7613 | 21_bit_twopoint_parametric_recessive |
| 3 | 87,749,292.00 | JHU_3.87798441 | 3.3956 | 22_bit_twopoint_parametric_recessive |
| 3 | 100,185,939.00 | GSA-rs12629505 | 3.97 | 22_bit_twopoint_parametric_recessive |
| 3 | 100,188,149.00 | rs9290003 | 3.4477 | 22_bit_twopoint_parametric_recessive |
| 3 | 104,224,854.00 | rs2895264 | 3.405 | 22_bit_twopoint_parametric_dominant |
| 3 | 108,251,237.00 | GSA-rs1531191 | 3.6033 | 22_bit_twopoint_parametric_recessive |
| 3 | 112,735,768.00 | JHU_3.112454614 | 3.3444 | 21_bit_twopoint_parametric_recessive |
| 3 | 112,735,768.00 | JHU_3.112454614 | 3.3444 | 21_bit_twopoint_parametric_recessive |
| 3 | 117,295,218.00 | rs4855941 | 3.5299 | 21_bit_twopoint_parametric_dominant |
| 3 | 117,295,218.00 | rs4855941 | 3.5299 | 21_bit_twopoint_parametric_dominant |
| 3 | 117,295,218.00 | rs4855941 | 3.5299 | 21_bit_twopoint_parametric_dominant |
| 3 | 117,295,218.00 | rs4855941 | 3.5299 | 21_bit_twopoint_parametric_dominant |
| 3 | 119,237,041.00 | rs3806698 | 3.3455 | 22_bit_twopoint_parametric_recessive |
| 3 | 147,308,854.00 | rs9831738 | 3.3542 | 21_bit_twopoint_parametric_dominant |
| 3 | 147,308,854.00 | rs9831738 | 3.3542 | 21_bit_twopoint_parametric_dominant |
| 3 | 147,317,800.00 | rs6440477 | 3.8575 | 21_bit_twopoint_parametric_dominant |
| 3 | 147,317,800.00 | rs6440477 | 3.8575 | 21_bit_twopoint_parametric_dominant |
| 3 | 176,976,778.00 | rs41333946 | 3.3656 | 22_bit_twopoint_parametric_dominant |
| 3 | 176,976,778.00 | rs41333946 | 4.2974 | 22_bit_twopoint_parametric_recessive |
| 4 | 17,329,137.00 | rs13107521 | 4.2108 | 21_bit_twopoint_parametric_recessive |
| 4 | 17,383,928.00 | rs2122574 | 3.3222 | 21_bit_twopoint_parametric_recessive |
| 4 | 38,300,069.00 | rs2890608 | 3.6381 | 22_bit_twopoint_parametric_recessive |
| 4 | 40,226,051.00 | rs11729556 | 3.5528 | 21_bit_twopoint_parametric_dominant |
| 4 | 42,193,360.00 | rs34809764 | 3.5193 | 21_bit_twopoint_parametric_recessive |
| 4 | 43,518,573.00 | rs4861284 | 3.4716 | 21_bit_twopoint_parametric_recessive |
| 4 | 88,249,578.00 | rs17013965 | 3.3713 | 21_bit_twopoint_parametric_recessive |
| 4 | 141,411,826.00 | rs59709790 | 3.3442 | 22_bit_twopoint_parametric_dominant |
| 4 | 151,550,916.00 | rs11726545 | 3.3432 | 22_bit_twopoint_parametric_dominant |
| 4 | 151,627,960.00 | rs12504273 | 3.3672 | 22_bit_twopoint_parametric_dominant |
| 4 | 151,669,185.00 | GSA-rs17027780 | 3.4045 | 22_bit_twopoint_parametric_dominant |
| 4 | 168,617,275.00 | rs2723687 | 3.4375 | 22_bit_twopoint_parametric_dominant |
| 5 | 33,011,880.00 | rs4867131 | 4.2588 | 22_bit_twopoint_parametric_dominant |
| 5 | 83,294,728.00 | GSA-rs28360285 | 3.319 | 21_bit_twopoint_parametric_recessive |
| 5 | 83,294,728.00 | GSA-rs28360285 | 3.8296 | 21_bit_twopoint_parametric_dominant |
| 5 | 107,030,138.00 | rs4073517 | 3.4294 | 22_bit_twopoint_parametric_recessive |
| 5 | 155,078,345.00 | rs1319694 | 3.5375 | 21_bit_twopoint_parametric_recessive |
| 5 | 167,636,509.00 | rs10516034 | 3.3466 | 22_bit_twopoint_parametric_recessive |
| 6 | 50,853,227.00 | GSA-rs2635727 | 3.4129 | 21_bit_twopoint_parametric_dominant |
| 6 | 127,985,973.00 | rs4341027 | 3.8552 | 21_bit_twopoint_parametric_dominant |
| 6 | 156,175,326.00 | rs7742923 | 3.5556 | 21_bit_twopoint_parametric_dominant |
| 7 | 21,775,270.00 | rs74418216 | 3.8888 | 22_bit_twopoint_parametric_recessive |
| 7 | 42,770,779.00 | rs2656510 | 3.8073 | 21_bit_twopoint_parametric_dominant |
| 7 | 85,902,146.00 | rs17339155 | 3.4859 | 22_bit_twopoint_parametric_recessive |
| 7 | 104,800,178.00 | rs2470957 | 3.4977 | 21_bit_twopoint_parametric_dominant |
| 7 | 111,860,497.00 | 7:111500553-C-T | 3.4072 | 21_bit_twopoint_parametric_dominant |
| 7 | 111,860,766.00 | 7:111500822-G-A | 3.4072 | 21_bit_twopoint_parametric_dominant |
| 7 | 111,860,880.00 | 7:111500936-G-A | 3.4072 | 21_bit_twopoint_parametric_dominant |
| 7 | 111,863,604.00 | 7:111503660-G-A | 3.4072 | 21_bit_twopoint_parametric_dominant |
| 7 | 111,886,263.00 | 7:111526319-T-C | 3.3882 | 21_bit_twopoint_parametric_dominant |
| 7 | 111,887,504.00 | rs10255299 | 3.393 | 21_bit_twopoint_parametric_dominant |
| 7 | 111,893,464.00 | 7:111533520-T-C | 3.5376 | 21_bit_twopoint_parametric_dominant |
| 7 | 111,897,412.00 | 7:111537468-T-C | 3.393 | 21_bit_twopoint_parametric_dominant |
| 7 | 111,899,490.00 | 7:111539546-G-A | 3.3986 | 21_bit_twopoint_parametric_dominant |
| 7 | 111,900,776.00 | 7:111540832-A-G | 3.393 | 21_bit_twopoint_parametric_dominant |
| 7 | 139,996,449.00 | rs2267705 | 3.7599 | 22_bit_twopoint_parametric_recessive |
| 7 | 139,996,449.00 | rs2267705 | 4.1465 | 21_bit_twopoint_parametric_recessive |
| 7 | 140,007,340.00 | rs740204 | 3.3445 | 22_bit_twopoint_parametric_recessive |
| 7 | 140,007,340.00 | rs740204 | 4.2437 | 21_bit_twopoint_parametric_recessive |
| 7 | 148,277,563.00 | JHU_7.147974654 | 4.2642 | 21_bit_twopoint_parametric_recessive |
| 8 | 8,614,994.00 | rs9329273 | 3.7602 | 21_bit_twopoint_parametric_dominant |
| 8 | 53,144,786.00 | rs1373387 | 4.0342 | 22_bit_twopoint_parametric_recessive |
| 8 | 53,151,988.00 | GSA-rs2114108 | 3.7893 | 22_bit_twopoint_parametric_recessive |
| 9 | 5,854,501.00 | GSA-rs1418746 | 3.3082 | 22_bit_twopoint_parametric_recessive |
| 9 | 5,854,501.00 | GSA-rs1418746 | 3.3082 | 22_bit_twopoint_parametric_recessive |
| 9 | 74,685,003.00 | GSA-rs77600772 | 3.8551 | 21_bit_twopoint_parametric_recessive |
| 9 | 74,696,783.00 | GSA-rs79485647 | 3.5879 | 21_bit_twopoint_parametric_recessive |
| 9 | 74,702,207.00 | GSA-rs11791579 | 4.0649 | 21_bit_twopoint_parametric_recessive |
| 9 | 75,529,916.00 | GSA-rs12000133 | 3.3947 | 22_bit_twopoint_parametric_dominant |
| 9 | 133,028,610.00 | GSA-rs623489 | 3.5551 | 21_bit_twopoint_parametric_dominant |
| 10 | 25,690,862.00 | rs7914182 | 3.4201 | 21_bit_twopoint_parametric_dominant |
| 10 | 132,260,382.00 | rs11146274 | 3.7958 | 21_bit_twopoint_parametric_dominant |
| 11 | 2,000,263.00 | JHU_11.2021492 | 3.5437 | 21_bit_twopoint_parametric_dominant |
| 11 | 83,081,483.00 | GSA-rs6592098 | 3.3226 | 22_bit_twopoint_parametric_recessive |
| 11 | 83,081,483.00 | GSA-rs6592098 | 4.6642 | 21_bit_twopoint_parametric_dominant |
| 11 | 84,435,099.00 | exm2259886 | 3.5832 | 21_bit_twopoint_parametric_dominant |
| 12 | 17,608,245.00 | rs10840763 | 3.462 | 21_bit_twopoint_parametric_dominant |
| 12 | 25,296,623.00 | GSA-rs4281557 | 3.5456 | 21_bit_multipoint_parametric_dominant |
| 12 | 25,296,623.00 | GSA-rs4281557 | 3.5456 | 21_bit_twopoint_parametric_dominant |
| 12 | 42,663,926.00 | rs4556609 | 3.4083 | 22_bit_twopoint_parametric_dominant |
| 12 | 124,435,245.00 | rs2660376 | 3.3172 | 22_bit_twopoint_parametric_dominant |
| 13 | 107,366,187.00 | rs4468469 | 3.5914 | 22_bit_twopoint_parametric_dominant |
| 15 | 52,728,736.00 | rs11634814 | 3.4247 | 22_bit_twopoint_parametric_dominant |
| 15 | 54,095,122.00 | rs12594549 | 3.3959 | 22_bit_twopoint_parametric_dominant |
| 15 | 54,095,122.00 | rs12594549 | 3.3959 | 22_bit_twopoint_parametric_dominant |
| 17 | 4,487,906.00 | rs2291742 | 4.0501 | 22_bit_twopoint_parametric_dominant |
| 17 | 4,487,906.00 | rs2291742 | 4.0641 | 22_bit_multipoint_parametric_dominant |
| 17 | 37,132,758.00 | rs7208415 | 3.5627 | 22_bit_twopoint_parametric_recessive |
| 17 | 40,854,610.00 | 17:39010862-A-C | 3.9088 | 21_bit_twopoint_parametric_dominant |
| 17 | 40,854,610.00 | 17:39010862-A-C | 3.9088 | 21_bit_twopoint_parametric_dominant |
| 17 | 42,166,716.00 | rs9912576 | 3.4176 | 21_bit_twopoint_parametric_dominant |
| 17 | 42,166,716.00 | rs9912576 | 3.4176 | 21_bit_twopoint_parametric_dominant |
| 17 | 44,983,400.00 | rs75193889 | 3.5212 | 22_bit_twopoint_parametric_dominant |
| 18 | 4,836,051.00 | rs9807161 | 4.7839 | 21_bit_twopoint_parametric_dominant |
| 18 | 4,836,051.00 | rs9807161 | 5.5792 | 22_bit_twopoint_parametric_dominant |
| 18 | 25,509,746.00 | rs7239453 | 3.4199 | 22_bit_twopoint_parametric_dominant |
| 18 | 57,648,852.00 | JHU_18.55316083 | 3.6165 | 22_bit_twopoint_parametric_dominant |
| 19 | 15,478,890.00 | rs1961562 | 3.872 | 21_bit_twopoint_parametric_dominant |
| 19 | 16,776,634.00 | rs12974841 | 3.6865 | 22_bit_twopoint_parametric_recessive |
| 20 | 12,955,872.00 | JHU_20.12936519 | 3.4116 | 22_bit_twopoint_parametric_recessive |
| 20 | 17,523,456.00 | rs2269051 | 3.5935 | 22_bit_twopoint_parametric_dominant |
| 20 | 62,160,042.00 | rs6121926 | 3.6736 | 21_bit_twopoint_parametric_dominant |
| 21 | 44,239,562.00 | JHU_21.45659444 | 4.1122 | 22_bit_twopoint_parametric_dominant |

SUPPLEMENTARY TABLE 4 (Leighanne Main)

**Supplementary Table 4 – Significant Linkage Analysis results.** Loci that surpassed the significant linkage threshold (≥ 3.3) for all analyses. For each SNP, chromosome number is followed by genomic coordinates (hg38), SNP identifier, LOD score, and to which analysis this LOD score corresponds.
