## Supplemental Table 5 for "Genetic analysis of cognitive preservation in the midwestern Amish reveals a novel locus on chromosome 2"

| CHR | POS | SNP | LOD | ANALYSIS |
| --- | --- | --- | --- | --- |
| 1 | 1,143,818.00 | rs11260603 | 2.6852 | 21_bit_twopoint_parametric_recessive |
| 1 | 1,755,523.00 | JHU_1.1686961 | 2.1377 | 21_bit_twopoint_parametric_dominant |
| 1 | 4,359,937.00 | rs17410050 | 2.1911 | 21_bit_twopoint_parametric_recessive |
| 1 | 4,421,155.00 | rs4654432 | 2.3002 | 21_bit_twopoint_parametric_recessive |
| 1 | 9,319,634.00 | rs1294046 | 1.8752 | 22_bit_twopoint_parametric_dominant |
| 1 | 11,866,999.00 | rs12562952 | 2.4333 | 21_bit_twopoint_parametric_dominant |
| 1 | 11,876,457.00 | GSA-rs11804222 | 1.8947 | 21_bit_twopoint_parametric_dominant |
| 1 | 13,610,786.00 | rs4391657 | 2.0932 | 21_bit_twopoint_parametric_recessive |
| 1 | 13,869,717.00 | rs16853069 | 2.3921 | 21_bit_twopoint_parametric_recessive |
| 1 | 13,869,717.00 | rs16853069 | 3.0341 | 22_bit_twopoint_parametric_recessive |
| 1 | 14,325,870.00 | rs12568762 | 1.8763 | 21_bit_twopoint_parametric_recessive |
| 1 | 14,341,822.00 | rs16853991 | 1.8763 | 21_bit_twopoint_parametric_recessive |
| 1 | 14,351,587.00 | rs77171346 | 1.8763 | 21_bit_twopoint_parametric_recessive |
| 1 | 17,460,819.00 | rs113343434 | 2.1353 | 21_bit_twopoint_parametric_dominant |
| 1 | 18,202,354.00 | rs1033190 | 2.1774 | 22_bit_twopoint_parametric_recessive |
| 1 | 18,567,915.00 | rs76644082 | 2.1519 | 22_bit_twopoint_parametric_recessive |
| 1 | 18,567,915.00 | rs76644082 | 2.7717 | 21_bit_twopoint_parametric_recessive |
| 1 | 19,760,913.00 | GSA-rs1473688 | 2.0127 | 21_bit_twopoint_parametric_dominant |
| 1 | 19,760,913.00 | GSA-rs1473688 | 2.077 | 21_bit_twopoint_parametric_recessive |
| 1 | 19,769,683.00 | GSA-rs1266438 | 1.9879 | 22_bit_twopoint_parametric_dominant |
| 1 | 20,288,602.00 | rs1022470 | 2.0019 | 21_bit_twopoint_parametric_dominant |
| 1 | 21,468,895.00 | exm27957 | 2.2534 | 22_bit_twopoint_parametric_dominant |
| 1 | 21,575,445.00 | 1:21901938-G-A | 1.9731 | 22_bit_twopoint_parametric_recessive |
| 1 | 22,159,904.00 | rs12729516 | 1.9425 | 21_bit_twopoint_parametric_dominant |
| 1 | 22,327,391.00 | rs12137132 | 2.2932 | 21_bit_twopoint_parametric_dominant |
| 1 | 22,330,370.00 | rs11580249 | 1.9789 | 22_bit_twopoint_parametric_recessive |
| 1 | 22,330,370.00 | rs11580249 | 2.1563 | 22_bit_twopoint_parametric_dominant |
| 1 | 22,330,370.00 | rs11580249 | 2.3486 | 21_bit_twopoint_parametric_recessive |
| 1 | 22,330,370.00 | rs11580249 | 3.0732 | 21_bit_twopoint_parametric_dominant |
| 1 | 23,397,442.00 | rs7516962 | 1.9896 | 21_bit_twopoint_parametric_recessive |
| 1 | 23,397,442.00 | rs7516962 | 1.9896 | 21_bit_twopoint_parametric_recessive |
| 1 | 24,180,371.00 | rs3897440 | 1.923 | 21_bit_twopoint_parametric_dominant |
| 1 | 38,759,631.00 | rs4147130 | 2.1542 | 22_bit_twopoint_parametric_dominant |
| 1 | 40,789,715.00 | rs12723678 | 2.2184 | 22_bit_twopoint_parametric_dominant |
| 1 | 42,805,257.00 | rs323719 | 2.2077 | 22_bit_twopoint_parametric_dominant |
| 1 | 42,805,362.00 | rs323720 | 1.8943 | 22_bit_twopoint_parametric_dominant |
| 1 | 44,133,088.00 | rs2254746 | 2.2842 | 22_bit_twopoint_parametric_dominant |
| 1 | 54,951,102.00 | rs12016659 | 1.9341 | 21_bit_twopoint_parametric_dominant |
| 1 | 54,954,446.00 | GSA-rs11206483 | 1.9708 | 22_bit_twopoint_parametric_dominant |
| 1 | 60,097,852.00 | rs12071650 | 2.1456 | 21_bit_twopoint_parametric_recessive |
| 1 | 60,097,852.00 | rs12071650 | 2.7274 | 22_bit_twopoint_parametric_recessive |
| 1 | 61,876,558.00 | GSA-rs6685516 | 2.1042 | 22_bit_twopoint_parametric_dominant |
| 1 | 63,288,065.00 | GSA-rs4915891 | 1.8822 | 22_bit_twopoint_parametric_dominant |
| 1 | 69,619,114.00 | rs11809230 | 2.1078 | 22_bit_twopoint_parametric_recessive |
| 1 | 69,759,657.00 | rs7523521 | 1.961 | 21_bit_twopoint_parametric_recessive |
| 1 | 74,426,653.00 | 1:74892337-A-G | 2.0575 | 21_bit_twopoint_parametric_recessive |
| 1 | 76,800,991.00 | rs7534641 | 2.0772 | 21_bit_twopoint_parametric_recessive |
| 1 | 77,414,503.00 | rs7512897 | 2.0736 | 22_bit_twopoint_parametric_recessive |
| 1 | 77,414,503.00 | rs7512897 | 2.0736 | 22_bit_twopoint_parametric_recessive |
| 1 | 79,824,153.00 | rs11162830 | 2.4376 | 21_bit_twopoint_parametric_dominant |
| 1 | 81,005,711.00 | rs34026797 | 2.0125 | 21_bit_twopoint_parametric_recessive |
| 1 | 81,005,711.00 | rs34026797 | 2.1318 | 22_bit_twopoint_parametric_recessive |
| 1 | 81,413,747.00 | rs74094108 | 2.3141 | 21_bit_twopoint_parametric_recessive |
| 1 | 88,650,617.00 | rs7534720 | 2.1333 | 22_bit_twopoint_parametric_recessive |
| 1 | 88,650,617.00 | rs7534720 | 2.1333 | 22_bit_twopoint_parametric_recessive |
| 1 | 91,153,209.00 | rs347027 | 2.7507 | 22_bit_twopoint_parametric_dominant |
| 1 | 94,741,968.00 | rs10493875 | 2.2351 | 22_bit_twopoint_parametric_dominant |
| 1 | 103,570,536.00 | GSA-rs12132911 | 2.2582 | 22_bit_twopoint_parametric_dominant |
| 1 | 107,367,256.00 | rs6664221 | 1.8802 | 21_bit_twopoint_parametric_dominant |
| 1 | 107,721,589.00 | rs12059570 | 3.2508 | 21_bit_twopoint_parametric_dominant |
| 1 | 107,744,349.00 | GSA-rs6583045 | 2.0423 | 22_bit_twopoint_parametric_recessive |
| 1 | 108,695,523.00 | GSA-rs6703128 | 2.0839 | 21_bit_twopoint_parametric_dominant |
| 1 | 110,727,983.00 | GSA-rs343609 | 2.0121 | 22_bit_twopoint_parametric_dominant |
| 1 | 110,727,983.00 | GSA-rs343609 | 2.0121 | 22_bit_twopoint_parametric_dominant |
| 1 | 110,727,983.00 | GSA-rs343609 | 2.1319 | 21_bit_twopoint_parametric_dominant |
| 1 | 110,727,983.00 | GSA-rs343609 | 2.1319 | 21_bit_twopoint_parametric_dominant |
| 1 | 110,753,326.00 | rs440156 | 2.4127 | 22_bit_twopoint_parametric_dominant |
| 1 | 110,753,326.00 | rs440156 | 2.4127 | 22_bit_twopoint_parametric_dominant |
| 1 | 110,753,326.00 | rs440156 | 2.669 | 21_bit_twopoint_parametric_dominant |
| 1 | 110,753,326.00 | rs440156 | 2.669 | 21_bit_twopoint_parametric_dominant |
| 1 | 110,759,001.00 | rs343791 | 2.0434 | 22_bit_twopoint_parametric_dominant |
| 1 | 110,759,001.00 | rs343791 | 2.0434 | 22_bit_twopoint_parametric_dominant |
| 1 | 110,827,750.00 | rs343769 | 2.9569 | 22_bit_twopoint_parametric_recessive |
| 1 | 110,827,750.00 | rs343769 | 2.9569 | 22_bit_twopoint_parametric_recessive |
| 1 | 112,496,173.00 | rs351354 | 2.3533 | 22_bit_twopoint_parametric_recessive |
| 1 | 112,968,134.00 | GSA-rs41283094 | 2.2121 | 22_bit_twopoint_parametric_recessive |
| 1 | 112,968,134.00 | GSA-rs41283094 | 2.6861 | 21_bit_twopoint_parametric_recessive |
| 1 | 113,126,295.00 | GSA-rs10494151 | 2.0044 | 22_bit_twopoint_parametric_recessive |
| 1 | 113,126,295.00 | GSA-rs10494151 | 2.6117 | 21_bit_twopoint_parametric_recessive |
| 1 | 114,133,007.00 | rs670495 | 2.5334 | 21_bit_twopoint_parametric_dominant |
| 1 | 114,632,222.00 | rs9308267 | 1.9055 | 21_bit_twopoint_parametric_recessive |
| 1 | 115,595,568.00 | rs112383816 | 2.1247 | 21_bit_twopoint_parametric_dominant |
| 1 | 115,597,499.00 | rs57714835 | 2.1335 | 21_bit_twopoint_parametric_dominant |
| 1 | 118,670,738.00 | GSA-rs56259287 | 1.8642 | 21_bit_twopoint_parametric_dominant |
| 1 | 118,929,285.00 | GSA-rs2645290 | 2.3175 | 22_bit_twopoint_parametric_recessive |
| 1 | 118,929,285.00 | GSA-rs2645290 | 3.1585 | 22_bit_twopoint_parametric_dominant |
| 1 | 151,399,662.00 | rs7172 | 1.9089 | 21_bit_twopoint_parametric_dominant |
| 1 | 151,399,662.00 | rs7172 | 1.9635 | 21_bit_twopoint_parametric_recessive |
| 1 | 151,401,549.00 | GSA-rs4603 | 1.9414 | 21_bit_twopoint_parametric_dominant |
| 1 | 151,401,549.00 | GSA-rs4603 | 1.9736 | 21_bit_twopoint_parametric_recessive |
| 1 | 151,473,276.00 | JHU_1.151445751 | 1.861 | 21_bit_twopoint_parametric_dominant |
| 1 | 152,698,909.00 | GSA-rs80271068 | 2.075 | 21_bit_twopoint_parametric_dominant |
| 1 | 152,698,909.00 | GSA-rs80271068 | 2.0995 | 21_bit_twopoint_parametric_recessive |
| 1 | 152,709,127.00 | GSA-rs41268472 | 2.0406 | 21_bit_twopoint_parametric_dominant |
| 1 | 153,390,031.00 | exm2273336 | 2.2634 | 21_bit_twopoint_parametric_dominant |
| 1 | 154,654,737.00 | rs10752605 | 1.8858 | 22_bit_twopoint_parametric_dominant |
| 1 | 154,842,336.00 | 1:154814812-C-T | 1.8908 | 22_bit_twopoint_parametric_dominant |
| 1 | 154,845,927.00 | 1:154818403-A-G | 1.8993 | 22_bit_twopoint_parametric_dominant |
| 1 | 154,861,707.00 | rs1218582 | 1.8919 | 22_bit_twopoint_parametric_recessive |
| 1 | 154,861,707.00 | rs1218582 | 2.0918 | 21_bit_twopoint_parametric_dominant |
| 1 | 157,092,904.00 | rs1176537 | 3.2563 | 22_bit_twopoint_parametric_recessive |
| 1 | 157,095,551.00 | JHU_1.157065342 | 2.3466 | 22_bit_twopoint_parametric_recessive |
| 1 | 157,160,928.00 | rs6657097 | 2.4997 | 22_bit_twopoint_parametric_recessive |
| 1 | 157,858,809.00 | rs34900478 | 2.4048 | 21_bit_twopoint_parametric_recessive |
| 1 | 159,205,095.00 | rs7550207 | 2.0363 | 22_bit_twopoint_parametric_recessive |
| 1 | 159,226,975.00 | exm-rs10489849 | 1.9276 | 22_bit_twopoint_parametric_dominant |
| 1 | 159,245,497.00 | rs12072644 | 1.865 | 22_bit_twopoint_parametric_dominant |
| 1 | 159,744,359.00 | rs4285692 | 2.4216 | 21_bit_twopoint_parametric_recessive |
| 1 | 162,001,915.00 | rs55681245 | 2.2489 | 21_bit_twopoint_parametric_recessive |
| 1 | 162,827,901.00 | rs2684874 | 2.4365 | 22_bit_twopoint_parametric_recessive |
| 1 | 165,425,645.00 | exm-rs466639 | 2.1454 | 22_bit_twopoint_parametric_recessive |
| 1 | 165,469,675.00 | rs10800102 | 3.1557 | 21_bit_twopoint_parametric_recessive |
| 1 | 165,478,920.00 | GSA-rs10918196 | 1.897 | 21_bit_twopoint_NPL |
| 1 | 165,478,920.00 | GSA-rs10918196 | 3.2149 | 22_bit_twopoint_parametric_recessive |
| 1 | 165,704,121.00 | JHU_1.165673357 | 1.8686 | 22_bit_twopoint_parametric_dominant |
| 1 | 165,993,535.00 | GSA-rs4656471 | 2.8811 | 22_bit_twopoint_parametric_recessive |
| 1 | 166,530,786.00 | rs269676 | 1.9174 | 22_bit_twopoint_parametric_recessive |
| 1 | 166,530,786.00 | rs269676 | 2.2223 | 21_bit_twopoint_parametric_recessive |
| 1 | 166,530,786.00 | rs269676 | 3.1108 | 21_bit_twopoint_parametric_dominant |
| 1 | 168,241,767.00 | rs58386753 | 1.8923 | 22_bit_twopoint_parametric_recessive |
| 1 | 168,241,767.00 | rs58386753 | 1.8938 | 21_bit_twopoint_parametric_dominant |
| 1 | 168,953,344.00 | rs12048750 | 2.8759 | 22_bit_twopoint_parametric_dominant |
| 1 | 168,953,344.00 | rs12048750 | 2.8759 | 22_bit_twopoint_parametric_dominant |
| 1 | 168,953,344.00 | rs12048750 | 3.0608 | 22_bit_twopoint_parametric_recessive |
| 1 | 168,953,344.00 | rs12048750 | 3.0608 | 22_bit_twopoint_parametric_recessive |
| 1 | 169,556,570.00 | 1:169525808-G-A | 1.9002 | 21_bit_twopoint_parametric_dominant |
| 1 | 169,582,444.00 | rs6028 | 1.9978 | 21_bit_twopoint_parametric_dominant |
| 1 | 170,026,661.00 | exm-rs1541160 | 1.9939 | 21_bit_twopoint_parametric_dominant |
| 1 | 171,715,366.00 | GSA-rs1001373 | 1.8735 | 21_bit_twopoint_parametric_recessive |
| 1 | 172,529,076.00 | rs10489279 | 1.9908 | 21_bit_twopoint_parametric_recessive |
| 1 | 173,422,069.00 | rs6702339 | 3.0038 | 22_bit_twopoint_parametric_recessive |
| 1 | 173,439,178.00 | GSA-rs12085083 | 3.0038 | 22_bit_twopoint_parametric_recessive |
| 1 | 173,492,069.00 | JHU_1.173461207 | 2.893 | 21_bit_twopoint_parametric_recessive |
| 1 | 173,561,887.00 | JHU_1.173531025 | 1.9546 | 21_bit_twopoint_parametric_recessive |
| 1 | 175,327,312.00 | exm-rs3766680 | 2.4503 | 22_bit_twopoint_parametric_dominant |
| 1 | 176,556,058.00 | rs2294654 | 2.1804 | 21_bit_twopoint_parametric_dominant |
| 1 | 177,936,334.00 | exm126209 | 2.3582 | 22_bit_twopoint_parametric_recessive |
| 1 | 177,936,334.00 | exm126209 | 2.9767 | 21_bit_twopoint_parametric_recessive |
| 1 | 178,750,806.00 | GSA-rs115104841 | 2.0027 | 21_bit_twopoint_parametric_recessive |
| 1 | 179,037,327.00 | rs6425513 | 2.2112 | 21_bit_twopoint_parametric_dominant |
| 1 | 179,638,147.00 | JHU_1.179607281 | 3.0458 | 21_bit_twopoint_parametric_dominant |
| 1 | 182,078,797.00 | rs12030839 | 2.3834 | 21_bit_twopoint_parametric_dominant |
| 1 | 198,605,772.00 | rs16843430 | 1.8919 | 21_bit_twopoint_parametric_recessive |
| 1 | 199,021,964.00 | rs2186188 | 2.4339 | 21_bit_twopoint_parametric_dominant |
| 1 | 202,113,933.00 | rs4463734 | 2.5742 | 21_bit_twopoint_parametric_dominant |
| 1 | 202,250,043.00 | rs6685457 | 1.9982 | 21_bit_twopoint_parametric_dominant |
| 1 | 202,250,043.00 | rs6685457 | 2.3398 | 21_bit_twopoint_parametric_recessive |
| 1 | 203,242,850.00 | GSA-rs2494289 | 1.9174 | 21_bit_twopoint_parametric_dominant |
| 1 | 206,108,197.00 | JHU_1.206233133 | 2.4624 | 21_bit_twopoint_parametric_dominant |
| 1 | 207,875,732.00 | rs2952 | 2.0169 | 21_bit_twopoint_parametric_dominant |
| 1 | 208,337,460.00 | rs4844419 | 1.8679 | 21_bit_twopoint_parametric_dominant |
| 1 | 208,337,460.00 | rs4844419 | 2.4203 | 22_bit_twopoint_parametric_dominant |
| 1 | 209,458,859.00 | rs7526722 | 1.8738 | 22_bit_twopoint_parametric_dominant |
| 1 | 209,608,998.00 | JHU_1.209782342 | 2.0036 | 22_bit_twopoint_parametric_recessive |
| 1 | 209,608,998.00 | JHU_1.209782342 | 2.7617 | 21_bit_twopoint_parametric_dominant |
| 1 | 209,608,998.00 | JHU_1.209782342 | 3.0369 | 21_bit_twopoint_parametric_recessive |
| 1 | 210,925,008.00 | rs1354112 | 2.8397 | 22_bit_twopoint_parametric_recessive |
| 1 | 211,014,321.00 | rs924568 | 2.1448 | 22_bit_twopoint_parametric_recessive |
| 1 | 211,014,321.00 | rs924568 | 2.1881 | 21_bit_twopoint_parametric_recessive |
| 1 | 211,084,819.00 | rs946143 | 2.0375 | 22_bit_twopoint_parametric_recessive |
| 1 | 211,516,124.00 | GSA-rs11800588 | 3.0097 | 21_bit_twopoint_parametric_dominant |
| 1 | 211,518,364.00 | rs7553035 | 3.0097 | 21_bit_twopoint_parametric_dominant |
| 1 | 211,518,935.00 | rs75906549 | 2.0318 | 21_bit_twopoint_parametric_dominant |
| 1 | 211,518,935.00 | rs75906549 | 2.086 | 22_bit_twopoint_parametric_dominant |
| 1 | 211,521,166.00 | rs17017668 | 3.0097 | 21_bit_twopoint_parametric_dominant |
| 1 | 214,151,875.00 | rs1470141 | 2.0897 | 21_bit_twopoint_parametric_recessive |
| 1 | 219,809,672.00 | rs72738768 | 2.3557 | 22_bit_twopoint_parametric_dominant |
| 1 | 221,914,491.00 | rs3828088 | 1.9026 | 21_bit_twopoint_parametric_recessive |
| 1 | 221,914,491.00 | rs3828088 | 1.9026 | 21_bit_twopoint_parametric_recessive |
| 1 | 223,658,173.00 | rs6668723 | 1.8653 | 21_bit_twopoint_parametric_dominant |
| 1 | 224,173,752.00 | 1:224361454-A-G | 1.8665 | 22_bit_twopoint_parametric_recessive |
| 1 | 230,071,686.00 | rs1474925 | 2.0546 | 21_bit_twopoint_parametric_dominant |
| 1 | 230,385,285.00 | rs11806158 | 1.8959 | 21_bit_twopoint_parametric_dominant |
| 1 | 230,385,285.00 | rs11806158 | 1.9197 | 21_bit_multipoint_parametric_dominant |
| 1 | 232,032,754.00 | rs9726024 | 2.2383 | 21_bit_twopoint_parametric_dominant |
| 1 | 232,178,848.00 | JHU_1.232314593 | 2.7447 | 22_bit_twopoint_parametric_dominant |
| 1 | 232,180,856.00 | GSA-rs1984027 | 2.6455 | 22_bit_twopoint_parametric_dominant |
| 1 | 232,183,045.00 | rs10864724 | 2.7116 | 22_bit_twopoint_parametric_dominant |
| 1 | 240,682,824.00 | rs4659996 | 2.0757 | 21_bit_twopoint_parametric_dominant |
| 1 | 246,776,641.00 | rs6685364 | 2.2273 | 22_bit_twopoint_parametric_dominant |
| 2 | 212,106.00 | rs2952790 | 2.0112 | 22_bit_twopoint_parametric_dominant |
| 2 | 1,363,739.00 | rs11894739 | 1.9081 | 21_bit_twopoint_parametric_dominant |
| 2 | 1,363,739.00 | rs11894739 | 3.0626 | 22_bit_twopoint_parametric_dominant |
| 2 | 1,911,525.00 | rs1368233 | 2.5664 | 22_bit_twopoint_parametric_dominant |
| 2 | 1,954,258.00 | rs10191292 | 2.2803 | 22_bit_twopoint_parametric_dominant |
| 2 | 2,223,863.00 | rs1862113 | 1.9209 | 22_bit_twopoint_parametric_dominant |
| 2 | 2,515,741.00 | exm-rs10199521 | 2.7235 | 22_bit_twopoint_parametric_dominant |
| 2 | 2,517,529.00 | rs2668843 | 2.6423 | 22_bit_twopoint_parametric_dominant |
| 2 | 4,632,434.00 | rs9808088 | 1.9188 | 22_bit_twopoint_parametric_recessive |
| 2 | 4,632,434.00 | rs9808088 | 2.1053 | 22_bit_twopoint_parametric_dominant |
| 2 | 6,514,490.00 | rs6431804 | 2.1465 | 21_bit_twopoint_parametric_dominant |
| 2 | 6,541,346.00 | rs17802554 | 3.0158 | 21_bit_twopoint_parametric_dominant |
| 2 | 7,654,805.00 | GSA-rs11682922 | 1.9005 | 21_bit_twopoint_parametric_dominant |
| 2 | 7,654,805.00 | GSA-rs11682922 | 1.9005 | 21_bit_twopoint_parametric_dominant |
| 2 | 9,924,321.00 | rs4669496 | 1.9687 | 21_bit_twopoint_parametric_dominant |
| 2 | 10,274,597.00 | GSA-rs6710746 | 2.0413 | 21_bit_twopoint_parametric_dominant |
| 2 | 11,581,956.00 | 2:11722082-C-T | 2.2603 | 22_bit_twopoint_parametric_recessive |
| 2 | 12,173,560.00 | JHU_2.12313685 | 1.9157 | 21_bit_twopoint_parametric_dominant |
| 2 | 12,173,560.00 | JHU_2.12313685 | 1.98 | 21_bit_twopoint_parametric_recessive |
| 2 | 12,173,560.00 | JHU_2.12313685 | 2.2936 | 22_bit_twopoint_parametric_recessive |
| 2 | 12,195,176.00 | rs78734409 | 1.8839 | 21_bit_twopoint_parametric_dominant |
| 2 | 12,195,176.00 | rs78734409 | 1.9415 | 21_bit_twopoint_parametric_recessive |
| 2 | 12,195,176.00 | rs78734409 | 2.2473 | 22_bit_twopoint_parametric_recessive |
| 2 | 12,478,983.00 | rs1547587 | 1.9454 | 21_bit_twopoint_parametric_dominant |
| 2 | 13,326,420.00 | GSA-rs114752265 | 2.0061 | 21_bit_twopoint_parametric_dominant |
| 2 | 13,326,420.00 | GSA-rs114752265 | 2.1111 | 22_bit_twopoint_parametric_dominant |
| 2 | 15,311,650.00 | GSA-rs10173491 | 2.0322 | 22_bit_twopoint_parametric_recessive |
| 2 | 15,582,505.00 | JHU_2.15722628 | 1.8664 | 21_bit_twopoint_parametric_dominant |
| 2 | 16,634,738.00 | rs13417603 | 2.1605 | 22_bit_twopoint_parametric_recessive |
| 2 | 16,634,738.00 | rs13417603 | 2.1982 | 21_bit_twopoint_parametric_recessive |
| 2 | 17,095,787.00 | GSA-rs12615585 | 2.0552 | 21_bit_twopoint_parametric_recessive |
| 2 | 18,192,765.00 | GSA-rs35959511 | 2.2379 | 21_bit_twopoint_parametric_recessive |
| 2 | 18,556,522.00 | rs2346135 | 1.9103 | 21_bit_twopoint_parametric_dominant |
| 2 | 18,960,982.00 | GSA-rs6531137 | 2.4019 | 21_bit_twopoint_parametric_dominant |
| 2 | 19,429,543.00 | rs13398721 | 1.8703 | 22_bit_twopoint_parametric_recessive |
| 2 | 19,429,543.00 | rs13398721 | 1.9189 | 21_bit_twopoint_parametric_recessive |
| 2 | 19,429,543.00 | rs13398721 | 2.1967 | 21_bit_twopoint_parametric_dominant |
| 2 | 19,429,543.00 | rs13398721 | 2.2818 | 22_bit_twopoint_parametric_dominant |
| 2 | 23,121,941.00 | JHU_2.23344811 | 1.9613 | 21_bit_twopoint_parametric_recessive |
| 2 | 23,245,436.00 | rs1037301 | 1.87 | 21_bit_twopoint_parametric_recessive |
| 2 | 27,101,801.00 | GSA-rs61753362 | 2.1305 | 21_bit_twopoint_parametric_recessive |
| 2 | 37,313,917.00 | GSA-rs4670688 | 2.4285 | 22_bit_twopoint_parametric_dominant |
| 2 | 45,318,733.00 | rs12995732 | 2.2049 | 22_bit_twopoint_parametric_dominant |
| 2 | 47,536,756.00 | rs748780 | 2.1407 | 22_bit_twopoint_parametric_dominant |
| 2 | 53,431,568.00 | GSA-rs72806786 | 1.9605 | 21_bit_twopoint_parametric_recessive |
| 2 | 53,431,568.00 | GSA-rs72806786 | 2.01 | 22_bit_twopoint_parametric_recessive |
| 2 | 53,431,568.00 | GSA-rs72806786 | 2.9001 | 21_bit_twopoint_parametric_dominant |
| 2 | 53,576,884.00 | GSA-rs2357486 | 2.0779 | 22_bit_twopoint_parametric_dominant |
| 2 | 53,607,946.00 | rs736085 | 2.4562 | 22_bit_twopoint_parametric_recessive |
| 2 | 55,972,675.00 | JHU_2.56199809 | 2.2118 | 22_bit_twopoint_parametric_recessive |
| 2 | 55,972,675.00 | JHU_2.56199809 | 2.3102 | 22_bit_twopoint_parametric_dominant |
| 2 | 55,974,981.00 | JHU_2.56202115 | 2.2118 | 22_bit_twopoint_parametric_recessive |
| 2 | 55,974,981.00 | JHU_2.56202115 | 2.3102 | 22_bit_twopoint_parametric_dominant |
| 2 | 56,046,569.00 | rs1368243 | 2.4944 | 22_bit_twopoint_parametric_recessive |
| 2 | 58,809,781.00 | rs6719884 | 2.2647 | 22_bit_twopoint_parametric_recessive |
| 2 | 58,812,863.00 | GSA-rs12617233 | 2.2647 | 22_bit_twopoint_parametric_recessive |
| 2 | 59,725,711.00 | GSA-rs6545747 | 1.9654 | 22_bit_twopoint_parametric_recessive |
| 2 | 61,001,159.00 | rs17039790 | 2.1058 | 21_bit_twopoint_parametric_recessive |
| 2 | 62,405,445.00 | GSA-rs79973408 | 2.82 | 21_bit_twopoint_parametric_recessive |
| 2 | 67,514,660.00 | rs1402440 | 1.8987 | 22_bit_twopoint_parametric_recessive |
| 2 | 68,335,369.00 | JHU_2.68562500 | 2.3723 | 22_bit_twopoint_parametric_recessive |
| 2 | 68,335,369.00 | JHU_2.68562500 | 2.3723 | 22_bit_twopoint_parametric_recessive |
| 2 | 68,345,342.00 | rs17035333 | 2.3843 | 22_bit_twopoint_parametric_recessive |
| 2 | 68,345,342.00 | rs17035333 | 2.3843 | 22_bit_twopoint_parametric_recessive |
| 2 | 68,351,885.00 | GSA-rs17035342 | 2.7702 | 22_bit_twopoint_parametric_recessive |
| 2 | 68,351,885.00 | GSA-rs17035342 | 2.7702 | 22_bit_twopoint_parametric_recessive |
| 2 | 68,351,885.00 | GSA-rs17035342 | 3.0206 | 22_bit_twopoint_parametric_dominant |
| 2 | 68,351,885.00 | GSA-rs17035342 | 3.0206 | 22_bit_twopoint_parametric_dominant |
| 2 | 68,395,858.00 | rs6725743 | 2.3985 | 22_bit_twopoint_parametric_recessive |
| 2 | 68,395,858.00 | rs6725743 | 2.3985 | 22_bit_twopoint_parametric_recessive |
| 2 | 68,424,631.00 | rs7557828 | 2.1162 | 22_bit_twopoint_parametric_recessive |
| 2 | 68,424,631.00 | rs7557828 | 2.1162 | 22_bit_twopoint_parametric_recessive |
| 2 | 70,133,130.00 | rs12992553 | 1.9281 | 21_bit_twopoint_parametric_dominant |
| 2 | 71,506,811.00 | rs9309457 | 1.9287 | 21_bit_twopoint_parametric_recessive |
| 2 | 71,837,757.00 | JHU_2.72064886 | 2.063 | 22_bit_twopoint_parametric_dominant |
| 2 | 71,862,807.00 | rs10208677 | 2.061 | 22_bit_twopoint_parametric_recessive |
| 2 | 71,933,282.00 | rs6753449 | 1.8951 | 22_bit_twopoint_parametric_dominant |
| 2 | 74,131,774.00 | rs2122290 | 2.3894 | 21_bit_twopoint_parametric_dominant |
| 2 | 75,099,624.00 | rs12713829 | 2.0083 | 21_bit_twopoint_parametric_recessive |
| 2 | 76,204,151.00 | exm2269286 | 2.2462 | 21_bit_twopoint_parametric_recessive |
| 2 | 76,204,151.00 | exm2269286 | 2.5471 | 21_bit_twopoint_parametric_dominant |
| 2 | 76,218,022.00 | rs9789473 | 2.4414 | 21_bit_twopoint_parametric_dominant |
| 2 | 76,261,438.00 | rs2860732 | 2.1258 | 21_bit_twopoint_parametric_recessive |
| 2 | 76,263,680.00 | GSA-rs13390508 | 1.9389 | 21_bit_twopoint_parametric_dominant |
| 2 | 76,762,442.00 | rs9917184 | 1.9582 | 21_bit_twopoint_parametric_dominant |
| 2 | 77,057,991.00 | rs1982340 | 2.0167 | 21_bit_twopoint_parametric_dominant |
| 2 | 78,854,459.00 | rs2861676 | 2.4509 | 22_bit_twopoint_parametric_dominant |
| 2 | 78,854,459.00 | rs2861676 | 2.4735 | 22_bit_twopoint_parametric_recessive |
| 2 | 78,854,459.00 | rs2861676 | 2.8033 | 21_bit_twopoint_parametric_dominant |
| 2 | 78,854,459.00 | rs2861676 | 3.2798 | 21_bit_twopoint_parametric_recessive |
| 2 | 78,873,098.00 | rs1402906 | 1.886 | 22_bit_twopoint_NPL |
| 2 | 78,873,098.00 | rs1402906 | 2.418 | 21_bit_twopoint_NPL |
| 2 | 78,873,098.00 | rs1402906 | 3.0225 | 22_bit_twopoint_parametric_recessive |
| 2 | 78,951,004.00 | rs11126689 | 2.1732 | 22_bit_twopoint_parametric_recessive |
| 2 | 84,159,993.00 | JHU_2.84387116 | 1.8773 | 22_bit_twopoint_parametric_recessive |
| 2 | 85,684,216.00 | rs7355681 | 1.9492 | 21_bit_twopoint_parametric_recessive |
| 2 | 88,575,373.00 | GSA-rs1805165 | 2.4703 | 22_bit_twopoint_parametric_dominant |
| 2 | 88,595,605.00 | exm209282 | 2.5991 | 22_bit_twopoint_parametric_dominant |
| 2 | 88,595,833.00 | exm-rs7571971 | 2.4185 | 22_bit_twopoint_parametric_dominant |
| 2 | 88,613,755.00 | exm209289 | 2.4804 | 22_bit_twopoint_parametric_dominant |
| 2 | 88,625,104.00 | rs11684404 | 2.7092 | 22_bit_twopoint_parametric_dominant |
| 2 | 88,728,129.00 | rs335115 | 2.6441 | 22_bit_twopoint_parametric_dominant |
| 2 | 101,598,767.00 | GSA-rs11884391 | 2.847 | 21_bit_twopoint_parametric_dominant |
| 2 | 101,617,269.00 | rs6543081 | 2.2804 | 21_bit_twopoint_parametric_recessive |
| 2 | 101,617,269.00 | rs6543081 | 2.3033 | 22_bit_twopoint_parametric_dominant |
| 2 | 104,891,131.00 | rs6719286 | 2.1016 | 21_bit_twopoint_parametric_dominant |
| 2 | 105,192,205.00 | GSA-rs2576791 | 1.9176 | 21_bit_twopoint_parametric_dominant |
| 2 | 105,566,496.00 | rs6729061 | 2.2256 | 22_bit_twopoint_parametric_dominant |
| 2 | 106,829,251.00 | GSA-rs1868256 | 1.8683 | 21_bit_twopoint_parametric_dominant |
| 2 | 106,887,913.00 | rs10166025 | 2.1906 | 22_bit_twopoint_parametric_dominant |
| 2 | 106,898,067.00 | GSA-rs13392153 | 2.1729 | 22_bit_twopoint_parametric_dominant |
| 2 | 109,285,291.00 | rs62152179 | 2.9871 | 22_bit_twopoint_parametric_dominant |
| 2 | 110,928,212.00 | GSA-rs4849255 | 2.1073 | 21_bit_twopoint_parametric_dominant |
| 2 | 110,928,212.00 | GSA-rs4849255 | 2.1073 | 21_bit_twopoint_parametric_dominant |
| 2 | 114,453,065.00 | rs1430090 | 2.1953 | 22_bit_twopoint_parametric_dominant |
| 2 | 114,513,945.00 | rs10201941 | 1.948 | 21_bit_twopoint_parametric_dominant |
| 2 | 117,811,620.00 | rs17507637 | 2.3905 | 22_bit_twopoint_parametric_recessive |
| 2 | 123,772,928.00 | rs17010489 | 2.2754 | 21_bit_twopoint_parametric_recessive |
| 2 | 123,772,928.00 | rs17010489 | 2.2754 | 21_bit_twopoint_parametric_recessive |
| 2 | 123,772,928.00 | rs17010489 | 2.7831 | 21_bit_twopoint_parametric_dominant |
| 2 | 123,772,928.00 | rs17010489 | 2.7831 | 21_bit_twopoint_parametric_dominant |
| 2 | 123,890,101.00 | rs6726456 | 2.275 | 21_bit_twopoint_parametric_recessive |
| 2 | 123,890,101.00 | rs6726456 | 2.275 | 21_bit_twopoint_parametric_recessive |
| 2 | 123,890,101.00 | rs6726456 | 2.7832 | 21_bit_twopoint_parametric_dominant |
| 2 | 123,890,101.00 | rs6726456 | 2.7832 | 21_bit_twopoint_parametric_dominant |
| 2 | 124,043,954.00 | rs1213952 | 2.4082 | 22_bit_twopoint_parametric_dominant |
| 2 | 124,043,954.00 | rs1213952 | 2.4082 | 22_bit_twopoint_parametric_dominant |
| 2 | 124,064,811.00 | rs1213946 | 2.3619 | 22_bit_twopoint_parametric_dominant |
| 2 | 124,064,811.00 | rs1213946 | 2.3619 | 22_bit_twopoint_parametric_dominant |
| 2 | 124,148,375.00 | GSA-rs17391285 | 3.05 | 22_bit_twopoint_parametric_dominant |
| 2 | 124,148,375.00 | GSA-rs17391285 | 3.05 | 22_bit_twopoint_parametric_dominant |
| 2 | 126,039,513.00 | rs11687421 | 2.4494 | 21_bit_twopoint_parametric_dominant |
| 2 | 126,039,513.00 | rs11687421 | 2.4782 | 21_bit_multipoint_parametric_dominant |
| 2 | 132,884,038.00 | JHU_2.133641610 | 2.8292 | 21_bit_twopoint_parametric_dominant |
| 2 | 132,891,267.00 | rs6731366 | 2.0348 | 22_bit_twopoint_parametric_dominant |
| 2 | 133,886,738.00 | rs13429789 | 1.9799 | 22_bit_twopoint_parametric_recessive |
| 2 | 136,842,079.00 | rs778193 | 2.4981 | 22_bit_twopoint_parametric_dominant |
| 2 | 141,401,398.00 | rs4485511 | 2.1633 | 21_bit_twopoint_parametric_dominant |
| 2 | 141,401,398.00 | rs4485511 | 2.6173 | 22_bit_twopoint_parametric_dominant |
| 2 | 147,362,495.00 | GSA-rs1522663 | 2.2211 | 22_bit_twopoint_parametric_dominant |
| 2 | 147,362,495.00 | GSA-rs1522663 | 2.2211 | 22_bit_twopoint_parametric_dominant |
| 2 | 147,362,495.00 | GSA-rs1522663 | 2.6116 | 21_bit_twopoint_parametric_dominant |
| 2 | 147,362,495.00 | GSA-rs1522663 | 2.6116 | 21_bit_twopoint_parametric_dominant |
| 2 | 147,380,514.00 | exm2261081 | 2.6593 | 21_bit_twopoint_parametric_dominant |
| 2 | 147,380,514.00 | exm2261081 | 2.6593 | 21_bit_twopoint_parametric_dominant |
| 2 | 149,163,815.00 | rs229335 | 2.1102 | 21_bit_twopoint_parametric_dominant |
| 2 | 158,248,684.00 | GSA-rs1402867 | 1.8641 | 22_bit_twopoint_parametric_recessive |
| 2 | 168,317,121.00 | rs4629128 | 2.8843 | 21_bit_twopoint_parametric_dominant |
| 2 | 170,163,424.00 | rs2161917 | 1.9865 | 21_bit_twopoint_parametric_recessive |
| 2 | 170,163,424.00 | rs2161917 | 2.4269 | 22_bit_twopoint_parametric_recessive |
| 2 | 170,169,674.00 | rs10930407 | 1.9789 | 22_bit_twopoint_parametric_recessive |
| 2 | 179,513,526.00 | rs10210118 | 1.8873 | 21_bit_twopoint_parametric_dominant |
| 2 | 184,870,872.00 | GSA-rs7588907 | 2.0734 | 22_bit_twopoint_parametric_dominant |
| 2 | 187,139,942.00 | rs1487400 | 1.9156 | 22_bit_twopoint_parametric_dominant |
| 2 | 192,333,788.00 | rs144524409 | 1.8764 | 22_bit_twopoint_parametric_dominant |
| 2 | 205,440,432.00 | exm259037 | 2.2692 | 21_bit_twopoint_parametric_dominant |
| 2 | 205,453,869.00 | exm-rs11884476 | 2.2692 | 21_bit_twopoint_parametric_dominant |
| 2 | 211,419,896.00 | rs3791704 | 1.8606 | 22_bit_twopoint_parametric_dominant |
| 2 | 213,010,402.00 | rs11676331 | 1.9834 | 22_bit_twopoint_parametric_recessive |
| 2 | 213,010,402.00 | rs11676331 | 2.3792 | 21_bit_twopoint_parametric_recessive |
| 2 | 217,056,046.00 | JHU_2.217920768 | 1.9837 | 22_bit_twopoint_parametric_dominant |
| 2 | 217,789,365.00 | rs2571460 | 2.358 | 21_bit_twopoint_parametric_dominant |
| 2 | 220,961,311.00 | rs1550068 | 2.349 | 22_bit_twopoint_parametric_recessive |
| 2 | 226,377,128.00 | GSA-rs10202706 | 1.9973 | 21_bit_twopoint_parametric_recessive |
| 2 | 230,179,337.00 | JHU_2.231044052 | 2.0728 | 22_bit_twopoint_parametric_recessive |
| 2 | 232,643,967.00 | exm2269235 | 2.0656 | 21_bit_twopoint_parametric_dominant |
| 2 | 233,602,042.00 | rs2741019 | 2.2559 | 22_bit_twopoint_parametric_recessive |
| 2 | 233,602,042.00 | rs2741019 | 2.4766 | 22_bit_twopoint_parametric_dominant |
| 2 | 234,084,919.00 | GSA-rs250956 | 2.0134 | 22_bit_twopoint_parametric_recessive |
| 2 | 234,088,241.00 | GSA-rs250945 | 1.9957 | 22_bit_twopoint_parametric_recessive |
| 2 | 236,021,729.00 | JHU_2.236930372 | 2.0325 | 21_bit_twopoint_parametric_recessive |
| 2 | 239,893,308.00 | rs13408710 | 2.3012 | 22_bit_twopoint_parametric_dominant |
| 2 | 239,903,292.00 | rs78219974 | 2.1538 | 22_bit_twopoint_parametric_dominant |
| 2 | 240,041,845.00 | exm281381 | 2.0438 | 22_bit_twopoint_parametric_recessive |
| 3 | 4,426,396.00 | rs17685501 | 1.877 | 22_bit_twopoint_parametric_recessive |
| 3 | 4,426,396.00 | rs17685501 | 1.8827 | 22_bit_multipoint_parametric_recessive |
| 3 | 6,052,686.00 | rs1607193 | 2.3289 | 22_bit_twopoint_parametric_dominant |
| 3 | 9,618,380.00 | rs6443251 | 2.139 | 22_bit_twopoint_parametric_dominant |
| 3 | 14,264,614.00 | rs900186 | 1.8936 | 22_bit_twopoint_parametric_dominant |
| 3 | 18,138,495.00 | rs9828987 | 2.5414 | 21_bit_twopoint_parametric_dominant |
| 3 | 18,138,495.00 | rs9828987 | 2.5414 | 21_bit_twopoint_parametric_dominant |
| 3 | 20,067,024.00 | rs13090341 | 2.0545 | 22_bit_twopoint_parametric_dominant |
| 3 | 21,156,351.00 | rs7635122 | 1.8917 | 21_bit_twopoint_parametric_dominant |
| 3 | 21,156,351.00 | rs7635122 | 1.8917 | 21_bit_twopoint_parametric_dominant |
| 3 | 21,156,351.00 | rs7635122 | 1.8917 | 21_bit_twopoint_parametric_dominant |
| 3 | 21,156,351.00 | rs7635122 | 1.8917 | 21_bit_twopoint_parametric_dominant |
| 3 | 21,156,351.00 | rs7635122 | 2.4891 | 21_bit_twopoint_parametric_recessive |
| 3 | 21,156,351.00 | rs7635122 | 2.4891 | 21_bit_twopoint_parametric_recessive |
| 3 | 21,156,351.00 | rs7635122 | 2.4891 | 21_bit_twopoint_parametric_recessive |
| 3 | 21,156,351.00 | rs7635122 | 2.4891 | 21_bit_twopoint_parametric_recessive |
| 3 | 21,256,203.00 | GSA-rs9860254 | 2.438 | 21_bit_twopoint_parametric_dominant |
| 3 | 21,256,203.00 | GSA-rs9860254 | 2.438 | 21_bit_twopoint_parametric_dominant |
| 3 | 21,256,203.00 | GSA-rs9860254 | 2.438 | 21_bit_twopoint_parametric_dominant |
| 3 | 21,256,203.00 | GSA-rs9860254 | 2.438 | 21_bit_twopoint_parametric_dominant |
| 3 | 21,483,646.00 | rs9883288 | 2.0666 | 22_bit_twopoint_parametric_recessive |
| 3 | 21,483,646.00 | rs9883288 | 2.0666 | 22_bit_twopoint_parametric_recessive |
| 3 | 21,502,604.00 | rs78506212 | 2.0603 | 22_bit_twopoint_parametric_recessive |
| 3 | 21,502,604.00 | rs78506212 | 2.0603 | 22_bit_twopoint_parametric_recessive |
| 3 | 22,435,282.00 | rs6789629 | 2.6201 | 22_bit_twopoint_parametric_dominant |
| 3 | 22,435,969.00 | GSA-rs76920506 | 2.6201 | 22_bit_twopoint_parametric_dominant |
| 3 | 22,444,183.00 | rs79010602 | 1.914 | 22_bit_twopoint_parametric_dominant |
| 3 | 22,890,243.00 | rs580227 | 2.7798 | 22_bit_twopoint_parametric_dominant |
| 3 | 22,985,397.00 | rs680930 | 2.4188 | 22_bit_twopoint_parametric_recessive |
| 3 | 22,985,397.00 | rs680930 | 2.8983 | 22_bit_twopoint_parametric_dominant |
| 3 | 22,990,734.00 | rs680110 | 2.3892 | 22_bit_twopoint_parametric_recessive |
| 3 | 23,013,017.00 | GSA-rs537660 | 2.0235 | 22_bit_twopoint_parametric_recessive |
| 3 | 23,086,636.00 | JHU_3.23128126 | 2.2796 | 22_bit_twopoint_parametric_recessive |
| 3 | 23,105,281.00 | GSA-rs6807204 | 2.1115 | 22_bit_twopoint_parametric_recessive |
| 3 | 23,108,817.00 | GSA-rs9826624 | 2.5054 | 22_bit_twopoint_parametric_dominant |
| 3 | 23,111,582.00 | rs6550726 | 2.6954 | 22_bit_twopoint_parametric_recessive |
| 3 | 23,120,749.00 | rs4390954 | 2.2461 | 22_bit_twopoint_parametric_dominant |
| 3 | 23,184,807.00 | rs6550742 | 1.9765 | 22_bit_twopoint_parametric_recessive |
| 3 | 24,854,674.00 | rs75931154 | 2.2562 | 22_bit_twopoint_parametric_dominant |
| 3 | 24,876,741.00 | rs2362772 | 2.0811 | 22_bit_twopoint_parametric_recessive |
| 3 | 26,522,418.00 | rs6763409 | 2.8339 | 22_bit_twopoint_parametric_recessive |
| 3 | 26,572,165.00 | rs9868148 | 2.4315 | 22_bit_twopoint_parametric_recessive |
| 3 | 27,521,122.00 | rs2643825 | 2.3424 | 21_bit_twopoint_parametric_recessive |
| 3 | 27,521,122.00 | rs2643825 | 2.3424 | 21_bit_twopoint_parametric_recessive |
| 3 | 27,521,122.00 | rs2643825 | 3.2395 | 22_bit_twopoint_parametric_recessive |
| 3 | 27,531,219.00 | rs1845700 | 2.4129 | 22_bit_twopoint_parametric_recessive |
| 3 | 27,664,884.00 | rs2643845 | 1.8734 | 21_bit_twopoint_parametric_recessive |
| 3 | 27,664,884.00 | rs2643845 | 1.8734 | 21_bit_twopoint_parametric_recessive |
| 3 | 27,717,913.00 | rs2581183 | 1.8642 | 22_bit_twopoint_parametric_dominant |
| 3 | 29,099,352.00 | rs1303912 | 1.9505 | 22_bit_twopoint_parametric_dominant |
| 3 | 29,277,718.00 | JHU_3.29319208 | 1.9167 | 22_bit_twopoint_parametric_recessive |
| 3 | 29,277,718.00 | JHU_3.29319208 | 1.9906 | 21_bit_twopoint_parametric_recessive |
| 3 | 29,277,718.00 | JHU_3.29319208 | 1.9906 | 21_bit_twopoint_parametric_recessive |
| 3 | 29,356,995.00 | rs6549930 | 2.0032 | 21_bit_twopoint_parametric_dominant |
| 3 | 29,356,995.00 | rs6549930 | 2.0032 | 21_bit_twopoint_parametric_dominant |
| 3 | 31,494,268.00 | GSA-rs9854271 | 2.6435 | 21_bit_twopoint_parametric_recessive |
| 3 | 31,494,268.00 | GSA-rs9854271 | 2.6435 | 21_bit_twopoint_parametric_recessive |
| 3 | 38,021,881.00 | rs6599099 | 2.5266 | 22_bit_twopoint_parametric_dominant |
| 3 | 38,524,085.00 | exm300987 | 1.8738 | 21_bit_twopoint_parametric_recessive |
| 3 | 38,524,085.00 | exm300987 | 1.8738 | 21_bit_twopoint_parametric_recessive |
| 3 | 38,524,085.00 | exm300987 | 1.9021 | 22_bit_twopoint_parametric_recessive |
| 3 | 39,252,313.00 | GSA-rs9861255 | 2.1415 | 22_bit_twopoint_parametric_recessive |
| 3 | 39,252,313.00 | GSA-rs9861255 | 3.0104 | 22_bit_twopoint_parametric_dominant |
| 3 | 42,077,987.00 | rs1909116 | 1.9682 | 21_bit_twopoint_parametric_dominant |
| 3 | 42,077,987.00 | rs1909116 | 1.9682 | 21_bit_twopoint_parametric_dominant |
| 3 | 46,680,172.00 | GSA-rs6803988 | 1.9073 | 22_bit_twopoint_parametric_dominant |
| 3 | 54,452,780.00 | JHU_3.54486806 | 2.68 | 22_bit_twopoint_parametric_recessive |
| 3 | 54,563,802.00 | rs2139683 | 2.0965 | 22_bit_twopoint_parametric_dominant |
| 3 | 54,563,802.00 | rs2139683 | 2.3444 | 22_bit_twopoint_parametric_recessive |
| 3 | 54,564,167.00 | GSA-rs7433850 | 2.219 | 22_bit_twopoint_parametric_dominant |
| 3 | 54,564,167.00 | GSA-rs7433850 | 2.3345 | 22_bit_twopoint_parametric_recessive |
| 3 | 54,587,973.00 | rs1851046 | 2.3684 | 22_bit_twopoint_parametric_dominant |
| 3 | 54,587,973.00 | rs1851046 | 2.3684 | 22_bit_twopoint_parametric_dominant |
| 3 | 54,588,132.00 | rs11130428 | 2.2483 | 22_bit_twopoint_parametric_dominant |
| 3 | 54,588,132.00 | rs11130428 | 2.2483 | 22_bit_twopoint_parametric_dominant |
| 3 | 55,403,787.00 | JHU_3.55437814 | 1.868 | 22_bit_twopoint_parametric_recessive |
| 3 | 55,404,166.00 | GSA-rs1829555 | 1.868 | 22_bit_twopoint_parametric_recessive |
| 3 | 56,383,379.00 | GSA-rs77804667 | 2.3275 | 21_bit_twopoint_parametric_recessive |
| 3 | 56,383,379.00 | GSA-rs77804667 | 2.3275 | 21_bit_twopoint_parametric_recessive |
| 3 | 56,840,005.00 | rs7636889 | 1.976 | 21_bit_twopoint_parametric_recessive |
| 3 | 56,840,005.00 | rs7636889 | 1.976 | 21_bit_twopoint_parametric_recessive |
| 3 | 58,085,917.00 | rs4681784 | 2.1906 | 22_bit_twopoint_parametric_dominant |
| 3 | 59,490,010.00 | exm2265456 | 1.9246 | 21_bit_twopoint_parametric_recessive |
| 3 | 59,490,010.00 | exm2265456 | 1.9246 | 21_bit_twopoint_parametric_recessive |
| 3 | 59,490,010.00 | exm2265456 | 2.0851 | 22_bit_twopoint_parametric_recessive |
| 3 | 60,367,889.00 | rs12489534 | 2.4031 | 22_bit_twopoint_parametric_dominant |
| 3 | 61,754,110.00 | rs636624 | 2.7462 | 21_bit_twopoint_parametric_dominant |
| 3 | 61,754,110.00 | rs636624 | 2.7462 | 21_bit_twopoint_parametric_dominant |
| 3 | 62,663,484.00 | GSA-rs11925415 | 1.9312 | 22_bit_twopoint_parametric_recessive |
| 3 | 65,294,624.00 | rs13084546 | 2.5806 | 22_bit_twopoint_parametric_recessive |
| 3 | 66,653,261.00 | rs9862866 | 1.9663 | 22_bit_twopoint_parametric_dominant |
| 3 | 66,717,635.00 | rs11128202 | 2.3475 | 22_bit_multipoint_parametric_recessive |
| 3 | 66,717,635.00 | rs11128202 | 2.3475 | 22_bit_twopoint_parametric_recessive |
| 3 | 66,717,635.00 | rs11128202 | 2.5581 | 21_bit_multipoint_parametric_recessive |
| 3 | 66,717,635.00 | rs11128202 | 2.5581 | 21_bit_twopoint_parametric_recessive |
| 3 | 66,717,635.00 | rs11128202 | 2.5581 | 21_bit_twopoint_parametric_recessive |
| 3 | 67,405,034.00 | rs4443191 | 1.9586 | 21_bit_twopoint_parametric_dominant |
| 3 | 67,405,034.00 | rs4443191 | 1.9586 | 21_bit_twopoint_parametric_dominant |
| 3 | 67,405,034.00 | rs4443191 | 2.0047 | 22_bit_twopoint_parametric_recessive |
| 3 | 67,405,034.00 | rs4443191 | 2.8743 | 21_bit_twopoint_parametric_recessive |
| 3 | 67,405,034.00 | rs4443191 | 2.8743 | 21_bit_twopoint_parametric_recessive |
| 3 | 70,577,209.00 | rs7628219 | 2.162 | 21_bit_twopoint_parametric_dominant |
| 3 | 70,577,209.00 | rs7628219 | 2.162 | 21_bit_twopoint_parametric_dominant |
| 3 | 71,714,953.00 | rs6807109 | 1.9241 | 21_bit_twopoint_parametric_dominant |
| 3 | 71,714,953.00 | rs6807109 | 1.9241 | 21_bit_twopoint_parametric_dominant |
| 3 | 71,714,953.00 | rs6807109 | 1.9241 | 21_bit_twopoint_parametric_dominant |
| 3 | 71,714,953.00 | rs6807109 | 1.9241 | 21_bit_twopoint_parametric_dominant |
| 3 | 72,239,594.00 | rs12632003 | 2.0578 | 22_bit_twopoint_parametric_dominant |
| 3 | 72,267,803.00 | rs9830272 | 2.1574 | 22_bit_twopoint_parametric_recessive |
| 3 | 72,326,461.00 | rs4677145 | 2.1423 | 22_bit_twopoint_parametric_recessive |
| 3 | 72,463,751.00 | rs2028241 | 2.0674 | 22_bit_twopoint_parametric_recessive |
| 3 | 72,463,751.00 | rs2028241 | 2.2958 | 21_bit_twopoint_parametric_recessive |
| 3 | 72,463,751.00 | rs2028241 | 2.2958 | 21_bit_twopoint_parametric_recessive |
| 3 | 73,617,665.00 | rs4676940 | 2.5676 | 22_bit_twopoint_parametric_dominant |
| 3 | 73,805,965.00 | JHU_3.73855115 | 2.1592 | 22_bit_twopoint_parametric_dominant |
| 3 | 73,805,965.00 | JHU_3.73855115 | 2.1737 | 22_bit_multipoint_parametric_dominant |
| 3 | 75,873,904.00 | rs9834641 | 1.9289 | 21_bit_twopoint_parametric_dominant |
| 3 | 75,873,904.00 | rs9834641 | 1.9289 | 21_bit_twopoint_parametric_dominant |
| 3 | 75,873,904.00 | rs9834641 | 1.9904 | 22_bit_twopoint_parametric_recessive |
| 3 | 75,873,904.00 | rs9834641 | 2.5067 | 21_bit_twopoint_parametric_recessive |
| 3 | 75,873,904.00 | rs9834641 | 2.5067 | 21_bit_twopoint_parametric_recessive |
| 3 | 77,036,099.00 | rs9862848 | 1.9974 | 21_bit_twopoint_parametric_dominant |
| 3 | 77,036,099.00 | rs9862848 | 1.9974 | 21_bit_twopoint_parametric_dominant |
| 3 | 82,114,084.00 | rs12638629 | 2.2768 | 21_bit_twopoint_parametric_recessive |
| 3 | 82,114,084.00 | rs12638629 | 2.2768 | 21_bit_twopoint_parametric_recessive |
| 3 | 82,114,084.00 | rs12638629 | 2.4945 | 22_bit_twopoint_parametric_dominant |
| 3 | 82,114,084.00 | rs12638629 | 3.1625 | 22_bit_twopoint_parametric_recessive |
| 3 | 82,190,248.00 | rs4561854 | 2.168 | 21_bit_twopoint_NPL |
| 3 | 82,190,248.00 | rs4561854 | 2.6915 | 22_bit_twopoint_parametric_recessive |
| 3 | 82,314,045.00 | rs7649805 | 2.113 | 22_bit_twopoint_parametric_dominant |
| 3 | 82,314,045.00 | rs7649805 | 2.5891 | 22_bit_twopoint_parametric_recessive |
| 3 | 82,436,793.00 | JHU_3.82485943 | 2.1781 | 22_bit_twopoint_parametric_dominant |
| 3 | 87,749,292.00 | JHU_3.87798441 | 1.8765 | 21_bit_twopoint_parametric_recessive |
| 3 | 87,749,292.00 | JHU_3.87798441 | 1.8765 | 21_bit_twopoint_parametric_recessive |
| 3 | 87,766,987.00 | rs4546169 | 1.9108 | 21_bit_twopoint_parametric_recessive |
| 3 | 87,766,987.00 | rs4546169 | 1.9108 | 21_bit_twopoint_parametric_recessive |
| 3 | 87,766,987.00 | rs4546169 | 2.5643 | 22_bit_twopoint_parametric_recessive |
| 3 | 87,930,059.00 | rs2938258 | 1.8813 | 21_bit_twopoint_parametric_recessive |
| 3 | 87,930,059.00 | rs2938258 | 1.8813 | 21_bit_twopoint_parametric_recessive |
| 3 | 94,597,274.00 | GSA-rs7642961 | 2.0736 | 21_bit_twopoint_parametric_recessive |
| 3 | 94,597,274.00 | GSA-rs7642961 | 2.0736 | 21_bit_twopoint_parametric_recessive |
| 3 | 99,669,059.00 | rs9859600 | 2.0467 | 21_bit_twopoint_parametric_recessive |
| 3 | 99,669,059.00 | rs9859600 | 2.0467 | 21_bit_twopoint_parametric_recessive |
| 3 | 100,165,665.00 | rs9875878 | 2.1716 | 22_bit_twopoint_parametric_dominant |
| 3 | 100,167,818.00 | exm333608 | 2.0151 | 22_bit_twopoint_parametric_dominant |
| 3 | 100,185,939.00 | GSA-rs12629505 | 1.949 | 21_bit_twopoint_NPL |
| 3 | 100,185,939.00 | GSA-rs12629505 | 1.981 | 22_bit_twopoint_NPL |
| 3 | 100,185,939.00 | GSA-rs12629505 | 2.1727 | 21_bit_twopoint_parametric_recessive |
| 3 | 100,185,939.00 | GSA-rs12629505 | 2.1727 | 21_bit_twopoint_parametric_recessive |
| 3 | 100,185,939.00 | GSA-rs12629505 | 2.378 | 21_bit_twopoint_parametric_dominant |
| 3 | 100,185,939.00 | GSA-rs12629505 | 2.378 | 21_bit_twopoint_parametric_dominant |
| 3 | 100,185,939.00 | GSA-rs12629505 | 2.4897 | 22_bit_twopoint_parametric_dominant |
| 3 | 100,188,149.00 | rs9290003 | 1.959 | 21_bit_twopoint_NPL |
| 3 | 100,760,540.00 | rs7642041 | 1.9032 | 22_bit_twopoint_parametric_dominant |
| 3 | 104,042,294.00 | rs1920329 | 1.952 | 22_bit_twopoint_parametric_dominant |
| 3 | 104,224,854.00 | rs2895264 | 2.7727 | 22_bit_twopoint_parametric_recessive |
| 3 | 105,281,625.00 | JHU_3.105000468 | 2.4854 | 21_bit_twopoint_parametric_recessive |
| 3 | 105,281,625.00 | JHU_3.105000468 | 2.4854 | 21_bit_twopoint_parametric_recessive |
| 3 | 105,281,625.00 | JHU_3.105000468 | 2.4854 | 21_bit_twopoint_parametric_recessive |
| 3 | 105,281,625.00 | JHU_3.105000468 | 2.4854 | 21_bit_twopoint_parametric_recessive |
| 3 | 105,281,625.00 | JHU_3.105000468 | 2.731 | 22_bit_twopoint_parametric_recessive |
| 3 | 105,281,625.00 | JHU_3.105000468 | 2.731 | 22_bit_twopoint_parametric_recessive |
| 3 | 105,976,371.00 | rs2618317 | 2.181 | 22_bit_twopoint_parametric_recessive |
| 3 | 106,022,997.00 | rs6437639 | 1.9197 | 22_bit_twopoint_parametric_recessive |
| 3 | 106,022,997.00 | rs6437639 | 2.909 | 21_bit_twopoint_parametric_recessive |
| 3 | 106,022,997.00 | rs6437639 | 2.909 | 21_bit_twopoint_parametric_recessive |
| 3 | 108,251,237.00 | GSA-rs1531191 | 2.007 | 22_bit_twopoint_NPL |
| 3 | 108,251,237.00 | GSA-rs1531191 | 3.2829 | 21_bit_twopoint_parametric_recessive |
| 3 | 108,251,237.00 | GSA-rs1531191 | 3.2829 | 21_bit_twopoint_parametric_recessive |
| 3 | 110,346,485.00 | rs13091887 | 1.9324 | 21_bit_twopoint_parametric_dominant |
| 3 | 110,346,485.00 | rs13091887 | 1.9324 | 21_bit_twopoint_parametric_dominant |
| 3 | 110,346,485.00 | rs13091887 | 2.37 | 22_bit_twopoint_parametric_dominant |
| 3 | 110,364,664.00 | exm2269506 | 2.3104 | 21_bit_twopoint_parametric_dominant |
| 3 | 110,364,664.00 | exm2269506 | 2.3104 | 21_bit_twopoint_parametric_dominant |
| 3 | 110,364,664.00 | exm2269506 | 2.9994 | 22_bit_twopoint_parametric_dominant |
| 3 | 112,735,768.00 | JHU_3.112454614 | 2.2238 | 21_bit_twopoint_parametric_dominant |
| 3 | 112,735,768.00 | JHU_3.112454614 | 2.2238 | 21_bit_twopoint_parametric_dominant |
| 3 | 112,735,768.00 | JHU_3.112454614 | 2.3308 | 22_bit_twopoint_parametric_recessive |
| 3 | 114,611,114.00 | rs6438213 | 2.054 | 21_bit_twopoint_parametric_dominant |
| 3 | 114,611,114.00 | rs6438213 | 2.054 | 21_bit_twopoint_parametric_dominant |
| 3 | 115,534,400.00 | JHU_3.115253246 | 2.0834 | 21_bit_twopoint_parametric_recessive |
| 3 | 115,534,400.00 | JHU_3.115253246 | 2.0834 | 21_bit_twopoint_parametric_recessive |
| 3 | 115,534,400.00 | JHU_3.115253246 | 2.7772 | 21_bit_twopoint_parametric_dominant |
| 3 | 115,534,400.00 | JHU_3.115253246 | 2.7772 | 21_bit_twopoint_parametric_dominant |
| 3 | 115,560,136.00 | rs9857602 | 2.102 | 21_bit_twopoint_parametric_recessive |
| 3 | 115,560,136.00 | rs9857602 | 2.102 | 21_bit_twopoint_parametric_recessive |
| 3 | 115,560,136.00 | rs9857602 | 2.8493 | 21_bit_twopoint_parametric_dominant |
| 3 | 115,560,136.00 | rs9857602 | 2.8493 | 21_bit_twopoint_parametric_dominant |
| 3 | 115,930,974.00 | rs7638681 | 3.0642 | 21_bit_twopoint_parametric_dominant |
| 3 | 115,930,974.00 | rs7638681 | 3.0642 | 21_bit_twopoint_parametric_dominant |
| 3 | 116,568,971.00 | rs72954002 | 2.5427 | 22_bit_twopoint_parametric_recessive |
| 3 | 117,062,319.00 | rs9840382 | 2.6631 | 22_bit_twopoint_parametric_recessive |
| 3 | 117,062,319.00 | rs9840382 | 2.6631 | 22_bit_twopoint_parametric_recessive |
| 3 | 117,254,561.00 | rs938170 | 3.1979 | 22_bit_twopoint_parametric_recessive |
| 3 | 117,254,561.00 | rs938170 | 3.1979 | 22_bit_twopoint_parametric_recessive |
| 3 | 117,295,218.00 | rs4855941 | 2.5281 | 22_bit_twopoint_parametric_dominant |
| 3 | 117,295,218.00 | rs4855941 | 2.5281 | 22_bit_twopoint_parametric_dominant |
| 3 | 117,596,959.00 | GSA-rs11719399 | 2.7334 | 22_bit_twopoint_parametric_dominant |
| 3 | 117,596,959.00 | GSA-rs11719399 | 2.7334 | 22_bit_twopoint_parametric_dominant |
| 3 | 117,820,961.00 | rs72645715 | 2.1922 | 22_bit_twopoint_parametric_recessive |
| 3 | 117,929,467.00 | rs77855782 | 1.8881 | 22_bit_twopoint_parametric_recessive |
| 3 | 117,954,317.00 | rs61355450 | 1.8881 | 22_bit_twopoint_parametric_recessive |
| 3 | 119,237,041.00 | rs3806698 | 1.9104 | 21_bit_twopoint_parametric_recessive |
| 3 | 119,237,041.00 | rs3806698 | 1.9104 | 21_bit_twopoint_parametric_recessive |
| 3 | 119,237,041.00 | rs3806698 | 2.4984 | 22_bit_twopoint_parametric_dominant |
| 3 | 119,841,759.00 | rs6805251 | 1.8957 | 22_bit_twopoint_parametric_dominant |
| 3 | 119,841,759.00 | rs6805251 | 2.0033 | 21_bit_twopoint_parametric_dominant |
| 3 | 119,841,759.00 | rs6805251 | 2.0033 | 21_bit_twopoint_parametric_dominant |
| 3 | 119,891,946.00 | GSA-rs6782799 | 1.9899 | 22_bit_twopoint_parametric_dominant |
| 3 | 119,891,946.00 | GSA-rs6782799 | 2.0603 | 21_bit_twopoint_parametric_dominant |
| 3 | 119,891,946.00 | GSA-rs6782799 | 2.0603 | 21_bit_twopoint_parametric_dominant |
| 3 | 119,912,967.00 | 3:119631814-A-G | 1.9881 | 22_bit_twopoint_parametric_dominant |
| 3 | 119,912,967.00 | 3:119631814-A-G | 2.062 | 21_bit_twopoint_parametric_dominant |
| 3 | 119,912,967.00 | 3:119631814-A-G | 2.062 | 21_bit_twopoint_parametric_dominant |
| 3 | 120,094,435.00 | 3:119813282-A-G | 1.9752 | 21_bit_twopoint_parametric_dominant |
| 3 | 120,094,435.00 | 3:119813282-A-G | 1.9752 | 21_bit_twopoint_parametric_dominant |
| 3 | 120,641,484.00 | rs804971 | 1.8984 | 21_bit_twopoint_parametric_dominant |
| 3 | 120,641,484.00 | rs804971 | 1.8984 | 21_bit_twopoint_parametric_dominant |
| 3 | 120,641,484.00 | rs804971 | 2.0586 | 21_bit_twopoint_parametric_recessive |
| 3 | 120,641,484.00 | rs804971 | 2.0586 | 21_bit_twopoint_parametric_recessive |
| 3 | 122,414,474.00 | rs6777596 | 1.9225 | 22_bit_twopoint_parametric_recessive |
| 3 | 123,438,630.00 | GSA-rs9841477 | 2.4 | 22_bit_twopoint_parametric_dominant |
| 3 | 123,438,630.00 | GSA-rs9841477 | 3.0045 | 21_bit_twopoint_parametric_dominant |
| 3 | 123,438,630.00 | GSA-rs9841477 | 3.0045 | 21_bit_twopoint_parametric_dominant |
| 3 | 123,477,972.00 | JHU_3.123196818 | 2.4469 | 21_bit_twopoint_parametric_dominant |
| 3 | 123,477,972.00 | JHU_3.123196818 | 2.4469 | 21_bit_twopoint_parametric_dominant |
| 3 | 127,277,932.00 | GSA-rs9860652 | 2.3822 | 21_bit_twopoint_parametric_dominant |
| 3 | 127,277,932.00 | GSA-rs9860652 | 2.3822 | 21_bit_twopoint_parametric_dominant |
| 3 | 127,277,932.00 | GSA-rs9860652 | 3.156 | 22_bit_twopoint_parametric_dominant |
| 3 | 128,449,424.00 | rs13061811 | 2.1707 | 22_bit_twopoint_parametric_dominant |
| 3 | 128,449,424.00 | rs13061811 | 2.1707 | 22_bit_twopoint_parametric_dominant |
| 3 | 132,340,395.00 | rs2310230 | 1.9389 | 22_bit_twopoint_parametric_dominant |
| 3 | 133,286,769.00 | GSA-rs6776275 | 1.9408 | 22_bit_twopoint_parametric_recessive |
| 3 | 133,733,600.00 | rs4241356 | 2.1868 | 21_bit_twopoint_parametric_dominant |
| 3 | 133,733,600.00 | rs4241356 | 2.1868 | 21_bit_twopoint_parametric_dominant |
| 3 | 133,733,600.00 | rs4241356 | 2.5939 | 22_bit_twopoint_parametric_dominant |
| 3 | 133,994,816.00 | 3:133713660 | 1.9242 | 22_bit_twopoint_parametric_dominant |
| 3 | 134,653,644.00 | rs9883955 | 3.1078 | 22_bit_twopoint_parametric_recessive |
| 3 | 139,812,052.00 | rs6789124 | 2.1557 | 22_bit_twopoint_parametric_recessive |
| 3 | 139,812,052.00 | rs6789124 | 2.1557 | 22_bit_twopoint_parametric_recessive |
| 3 | 140,445,341.00 | rs2114860 | 2.7272 | 21_bit_twopoint_parametric_dominant |
| 3 | 140,445,341.00 | rs2114860 | 2.7272 | 21_bit_twopoint_parametric_dominant |
| 3 | 140,445,341.00 | rs2114860 | 2.7272 | 21_bit_twopoint_parametric_dominant |
| 3 | 140,445,341.00 | rs2114860 | 2.7272 | 21_bit_twopoint_parametric_dominant |
| 3 | 140,461,721.00 | rs6439928 | 2.5807 | 21_bit_twopoint_parametric_dominant |
| 3 | 140,461,721.00 | rs6439928 | 2.5807 | 21_bit_twopoint_parametric_dominant |
| 3 | 140,461,721.00 | rs6439928 | 2.5807 | 21_bit_twopoint_parametric_dominant |
| 3 | 140,461,721.00 | rs6439928 | 2.5807 | 21_bit_twopoint_parametric_dominant |
| 3 | 145,166,545.00 | rs7622285 | 2.8279 | 21_bit_twopoint_parametric_recessive |
| 3 | 145,166,545.00 | rs7622285 | 2.8279 | 21_bit_twopoint_parametric_recessive |
| 3 | 146,806,630.00 | rs6781142 | 2.2144 | 22_bit_twopoint_parametric_recessive |
| 3 | 147,308,854.00 | rs9831738 | 2.0802 | 22_bit_twopoint_parametric_dominant |
| 3 | 147,308,854.00 | rs9831738 | 2.829 | 21_bit_twopoint_parametric_recessive |
| 3 | 147,308,854.00 | rs9831738 | 2.829 | 21_bit_twopoint_parametric_recessive |
| 3 | 147,317,800.00 | rs6440477 | 1.938 | 22_bit_twopoint_parametric_recessive |
| 3 | 147,317,800.00 | rs6440477 | 2.3592 | 22_bit_twopoint_parametric_dominant |
| 3 | 147,317,800.00 | rs6440477 | 2.9609 | 21_bit_twopoint_parametric_recessive |
| 3 | 147,317,800.00 | rs6440477 | 2.9609 | 21_bit_twopoint_parametric_recessive |
| 3 | 148,329,558.00 | GSA-rs62275230 | 1.8915 | 22_bit_twopoint_parametric_recessive |
| 3 | 148,329,558.00 | GSA-rs62275230 | 1.8915 | 22_bit_twopoint_parametric_recessive |
| 3 | 148,629,955.00 | rs275685 | 1.8961 | 22_bit_twopoint_parametric_recessive |
| 3 | 148,629,955.00 | rs275685 | 1.8961 | 22_bit_twopoint_parametric_recessive |
| 3 | 150,823,576.00 | rs10935819 | 2.3177 | 21_bit_twopoint_parametric_dominant |
| 3 | 150,823,576.00 | rs10935819 | 2.3177 | 21_bit_twopoint_parametric_dominant |
| 3 | 156,617,924.00 | rs2176618 | 2.2599 | 22_bit_twopoint_parametric_dominant |
| 3 | 157,428,114.00 | rs7632702 | 1.8907 | 22_bit_twopoint_parametric_dominant |
| 3 | 157,437,525.00 | GSA-rs3816527 | 2.3815 | 22_bit_twopoint_parametric_dominant |
| 3 | 157,448,334.00 | GSA-rs4680367 | 2.3639 | 22_bit_twopoint_parametric_dominant |
| 3 | 157,614,622.00 | rs10936094 | 2.1295 | 22_bit_twopoint_parametric_dominant |
| 3 | 168,651,587.00 | rs10804826 | 1.896 | 22_bit_twopoint_NPL |
| 3 | 168,651,587.00 | rs10804826 | 2.2168 | 21_bit_twopoint_parametric_dominant |
| 3 | 168,651,587.00 | rs10804826 | 2.2168 | 21_bit_twopoint_parametric_dominant |
| 3 | 168,651,587.00 | rs10804826 | 2.2494 | 21_bit_twopoint_parametric_recessive |
| 3 | 168,651,587.00 | rs10804826 | 2.2494 | 21_bit_twopoint_parametric_recessive |
| 3 | 168,651,587.00 | rs10804826 | 3.0013 | 22_bit_twopoint_parametric_recessive |
| 3 | 168,721,570.00 | rs6444823 | 1.8945 | 21_bit_twopoint_parametric_recessive |
| 3 | 168,721,570.00 | rs6444823 | 1.8945 | 21_bit_twopoint_parametric_recessive |
| 3 | 168,721,570.00 | rs6444823 | 2.5164 | 22_bit_twopoint_parametric_recessive |
| 3 | 168,721,570.00 | rs6444823 | 2.9059 | 22_bit_twopoint_parametric_dominant |
| 3 | 168,749,887.00 | JHU_3.168467674 | 2.3282 | 22_bit_twopoint_parametric_recessive |
| 3 | 168,749,887.00 | JHU_3.168467674 | 2.9383 | 22_bit_twopoint_parametric_dominant |
| 3 | 168,803,055.00 | rs496741 | 2.0285 | 22_bit_twopoint_parametric_dominant |
| 3 | 176,976,778.00 | rs41333946 | 2.1796 | 21_bit_twopoint_parametric_recessive |
| 3 | 176,976,778.00 | rs41333946 | 2.1796 | 21_bit_twopoint_parametric_recessive |
| 3 | 177,402,515.00 | rs7639599 | 2.1096 | 21_bit_twopoint_parametric_recessive |
| 3 | 177,402,515.00 | rs7639599 | 2.1096 | 21_bit_twopoint_parametric_recessive |
| 3 | 177,402,515.00 | rs7639599 | 2.4457 | 21_bit_twopoint_parametric_dominant |
| 3 | 177,402,515.00 | rs7639599 | 2.4457 | 21_bit_twopoint_parametric_dominant |
| 3 | 177,511,191.00 | rs1353899 | 1.9963 | 22_bit_twopoint_parametric_dominant |
| 3 | 178,826,529.00 | rs4624543 | 1.9734 | 21_bit_twopoint_parametric_dominant |
| 3 | 178,826,529.00 | rs4624543 | 1.9734 | 21_bit_twopoint_parametric_dominant |
| 3 | 178,826,529.00 | rs4624543 | 1.9734 | 21_bit_twopoint_parametric_dominant |
| 3 | 178,826,529.00 | rs4624543 | 1.9734 | 21_bit_twopoint_parametric_dominant |
| 3 | 180,651,495.00 | chr3-180369283 | 2.4556 | 22_bit_twopoint_parametric_recessive |
| 3 | 180,651,495.00 | chr3-180369283 | 2.6131 | 22_bit_twopoint_parametric_dominant |
| 3 | 182,270,994.00 | rs12634003 | 1.919 | 22_bit_twopoint_parametric_recessive |
| 3 | 182,270,994.00 | rs12634003 | 2.9134 | 21_bit_twopoint_parametric_dominant |
| 3 | 182,270,994.00 | rs12634003 | 2.9134 | 21_bit_twopoint_parametric_dominant |
| 3 | 182,270,994.00 | rs12634003 | 3.0729 | 22_bit_twopoint_parametric_dominant |
| 3 | 182,726,631.00 | rs2687806 | 2.3506 | 21_bit_twopoint_parametric_recessive |
| 3 | 182,726,631.00 | rs2687806 | 2.3506 | 21_bit_twopoint_parametric_recessive |
| 3 | 182,922,639.00 | JHU_3.182640426 | 2.0421 | 22_bit_twopoint_parametric_recessive |
| 3 | 183,153,676.00 | JHU_3.182871463 | 1.9055 | 22_bit_twopoint_parametric_recessive |
| 3 | 183,247,486.00 | rs16857199 | 3.021 | 22_bit_twopoint_parametric_dominant |
| 3 | 183,247,486.00 | rs16857199 | 3.1094 | 22_bit_twopoint_parametric_recessive |
| 3 | 183,247,486.00 | rs16857199 | 3.1323 | 21_bit_twopoint_parametric_recessive |
| 3 | 183,247,486.00 | rs16857199 | 3.1323 | 21_bit_twopoint_parametric_recessive |
| 3 | 183,247,486.00 | rs16857199 | 3.1518 | 21_bit_twopoint_parametric_dominant |
| 3 | 183,247,486.00 | rs16857199 | 3.1518 | 21_bit_twopoint_parametric_dominant |
| 3 | 183,868,095.00 | rs953419 | 2.2546 | 21_bit_twopoint_parametric_dominant |
| 3 | 183,868,095.00 | rs953419 | 2.2546 | 21_bit_twopoint_parametric_dominant |
| 3 | 185,463,024.00 | rs9878704 | 2.0742 | 22_bit_twopoint_parametric_dominant |
| 3 | 185,484,919.00 | rs2293206 | 2.2961 | 22_bit_twopoint_parametric_dominant |
| 3 | 186,159,593.00 | rs7627157 | 2.457 | 22_bit_twopoint_parametric_dominant |
| 3 | 187,539,752.00 | GSA-rs67560135 | 2.113 | 22_bit_twopoint_parametric_recessive |
| 3 | 187,539,752.00 | GSA-rs67560135 | 2.113 | 22_bit_twopoint_parametric_recessive |
| 3 | 187,577,896.00 | GSA-rs60376301 | 1.9886 | 22_bit_twopoint_parametric_recessive |
| 3 | 187,577,896.00 | GSA-rs60376301 | 1.9886 | 22_bit_twopoint_parametric_recessive |
| 3 | 187,825,554.00 | rs10513821 | 1.9833 | 22_bit_twopoint_parametric_dominant |
| 3 | 187,825,554.00 | rs10513821 | 1.9833 | 22_bit_twopoint_parametric_dominant |
| 3 | 187,912,939.00 | rs11711595 | 2.3316 | 22_bit_twopoint_parametric_recessive |
| 3 | 187,912,939.00 | rs11711595 | 2.3316 | 22_bit_twopoint_parametric_recessive |
| 3 | 187,912,939.00 | rs11711595 | 2.4259 | 22_bit_twopoint_parametric_dominant |
| 3 | 187,912,939.00 | rs11711595 | 2.4259 | 22_bit_twopoint_parametric_dominant |
| 3 | 188,253,260.00 | rs9844029 | 2.4367 | 22_bit_twopoint_parametric_dominant |
| 3 | 188,253,260.00 | rs9844029 | 2.4375 | 22_bit_multipoint_parametric_dominant |
| 3 | 191,205,804.00 | rs9843668 | 2.045 | 22_bit_twopoint_parametric_dominant |
| 3 | 192,155,435.00 | rs10937534 | 2.2536 | 22_bit_twopoint_parametric_recessive |
| 3 | 192,155,435.00 | rs10937534 | 3.2113 | 22_bit_twopoint_parametric_dominant |
| 3 | 192,361,098.00 | rs11711838 | 2.4408 | 21_bit_twopoint_parametric_dominant |
| 3 | 192,361,098.00 | rs11711838 | 2.4408 | 21_bit_twopoint_parametric_dominant |
| 3 | 192,554,936.00 | JHU_3.192272724 | 1.9473 | 21_bit_twopoint_parametric_dominant |
| 3 | 192,554,936.00 | JHU_3.192272724 | 1.9473 | 21_bit_twopoint_parametric_dominant |
| 3 | 192,554,936.00 | JHU_3.192272724 | 2.0751 | 21_bit_twopoint_parametric_recessive |
| 3 | 192,554,936.00 | JHU_3.192272724 | 2.0751 | 21_bit_twopoint_parametric_recessive |
| 3 | 192,555,978.00 | GSA-rs2669297 | 1.9473 | 21_bit_twopoint_parametric_dominant |
| 3 | 192,555,978.00 | GSA-rs2669297 | 1.9473 | 21_bit_twopoint_parametric_dominant |
| 3 | 192,555,978.00 | GSA-rs2669297 | 2.0751 | 21_bit_twopoint_parametric_recessive |
| 3 | 192,555,978.00 | GSA-rs2669297 | 2.0751 | 21_bit_twopoint_parametric_recessive |
| 3 | 192,556,302.00 | JHU_3.192274090 | 1.8968 | 21_bit_twopoint_parametric_recessive |
| 3 | 192,556,302.00 | JHU_3.192274090 | 1.8968 | 21_bit_twopoint_parametric_recessive |
| 3 | 193,394,076.00 | rs902192 | 2.2479 | 21_bit_twopoint_parametric_dominant |
| 3 | 193,394,076.00 | rs902192 | 2.2479 | 21_bit_twopoint_parametric_dominant |
| 3 | 193,953,547.00 | rs7624193 | 2.1793 | 22_bit_twopoint_parametric_dominant |
| 3 | 193,953,547.00 | rs7624193 | 2.2679 | 21_bit_twopoint_parametric_dominant |
| 3 | 193,953,547.00 | rs7624193 | 2.2679 | 21_bit_twopoint_parametric_dominant |
| 3 | 194,294,387.00 | rs11922293 | 1.8994 | 21_bit_twopoint_parametric_dominant |
| 3 | 194,294,387.00 | rs11922293 | 1.8994 | 21_bit_twopoint_parametric_dominant |
| 3 | 194,294,387.00 | rs11922293 | 1.8994 | 21_bit_twopoint_parametric_dominant |
| 3 | 194,294,387.00 | rs11922293 | 1.8994 | 21_bit_twopoint_parametric_dominant |
| 3 | 194,675,200.00 | GSA-rs7612967 | 1.9893 | 21_bit_twopoint_parametric_dominant |
| 3 | 194,675,200.00 | GSA-rs7612967 | 1.9893 | 21_bit_twopoint_parametric_dominant |
| 3 | 194,684,212.00 | rs9846429 | 2.2377 | 21_bit_twopoint_parametric_dominant |
| 3 | 194,684,212.00 | rs9846429 | 2.2377 | 21_bit_twopoint_parametric_dominant |
| 3 | 194,818,321.00 | JHU_3.194539049 | 1.9308 | 21_bit_twopoint_parametric_dominant |
| 3 | 194,818,321.00 | JHU_3.194539049 | 1.9308 | 21_bit_twopoint_parametric_dominant |
| 3 | 194,818,321.00 | JHU_3.194539049 | 2.0328 | 21_bit_twopoint_parametric_recessive |
| 3 | 194,818,321.00 | JHU_3.194539049 | 2.0328 | 21_bit_twopoint_parametric_recessive |
| 3 | 194,852,883.00 | rs9825236 | 2.2752 | 22_bit_multipoint_parametric_dominant |
| 3 | 194,852,883.00 | rs9825236 | 2.2987 | 22_bit_twopoint_parametric_dominant |
| 3 | 194,856,113.00 | rs4677766 | 2.6568 | 22_bit_twopoint_parametric_recessive |
| 3 | 194,856,113.00 | rs4677766 | 3.1931 | 21_bit_twopoint_parametric_recessive |
| 3 | 194,856,113.00 | rs4677766 | 3.1931 | 21_bit_twopoint_parametric_recessive |
| 3 | 195,012,736.00 | JHU_3.194733464 | 1.8724 | 22_bit_twopoint_parametric_recessive |
| 3 | 195,012,736.00 | JHU_3.194733464 | 2.4276 | 21_bit_twopoint_parametric_recessive |
| 3 | 195,012,736.00 | JHU_3.194733464 | 2.4276 | 21_bit_twopoint_parametric_recessive |
| 3 | 195,802,897.00 | GSA-rs6791220 | 2.3767 | 21_bit_twopoint_parametric_recessive |
| 3 | 195,802,897.00 | GSA-rs6791220 | 2.3767 | 21_bit_twopoint_parametric_recessive |
| 3 | 196,472,333.00 | GSA-rs3796129 | 2.0654 | 21_bit_twopoint_parametric_dominant |
| 3 | 196,472,333.00 | GSA-rs3796129 | 2.0654 | 21_bit_twopoint_parametric_dominant |
| 3 | 196,472,333.00 | GSA-rs3796129 | 2.1847 | 22_bit_twopoint_parametric_dominant |
| 3 | 196,483,937.00 | rs7646174 | 2.1271 | 21_bit_twopoint_parametric_recessive |
| 3 | 196,483,937.00 | rs7646174 | 2.1271 | 21_bit_twopoint_parametric_recessive |
| 3 | 196,483,937.00 | rs7646174 | 2.3751 | 21_bit_twopoint_parametric_dominant |
| 3 | 196,483,937.00 | rs7646174 | 2.3751 | 21_bit_twopoint_parametric_dominant |
| 3 | 197,054,254.00 | JHU_3.196781124 | 2.1786 | 22_bit_twopoint_parametric_dominant |
| 3 | 197,375,038.00 | GSA-rs9325388 | 2.0033 | 22_bit_twopoint_parametric_recessive |
| 3 | 197,381,482.00 | rs4132331 | 2.1855 | 21_bit_twopoint_parametric_recessive |
| 3 | 197,381,482.00 | rs4132331 | 2.1855 | 21_bit_twopoint_parametric_recessive |
| 3 | 197,381,482.00 | rs4132331 | 2.906 | 22_bit_twopoint_parametric_recessive |
| 3 | 197,381,482.00 | rs4132331 | 3.1217 | 21_bit_twopoint_parametric_dominant |
| 3 | 197,381,482.00 | rs4132331 | 3.1217 | 21_bit_twopoint_parametric_dominant |
| 3 | 197,381,482.00 | rs4132331 | 3.1926 | 22_bit_twopoint_parametric_dominant |
| 4 | 1,165,203.00 | rs17802266 | 2.2046 | 21_bit_twopoint_parametric_dominant |
| 4 | 3,719,542.00 | rs7377501 | 2.1586 | 22_bit_twopoint_parametric_recessive |
| 4 | 3,900,815.00 | rs28767603 | 1.8655 | 22_bit_twopoint_parametric_dominant |
| 4 | 4,742,310.00 | GSA-rs10516168 | 2.0648 | 22_bit_twopoint_parametric_dominant |
| 4 | 4,742,310.00 | GSA-rs10516168 | 2.2891 | 22_bit_twopoint_parametric_recessive |
| 4 | 4,880,423.00 | rs10937898 | 2.1393 | 21_bit_twopoint_parametric_dominant |
| 4 | 5,458,300.00 | rs6446363 | 1.932 | 22_bit_twopoint_parametric_recessive |
| 4 | 5,811,247.00 | rs16837692 | 2.2434 | 22_bit_twopoint_parametric_recessive |
| 4 | 7,302,927.00 | rs1546267 | 2.1261 | 22_bit_twopoint_parametric_dominant |
| 4 | 7,302,927.00 | rs1546267 | 2.1261 | 22_bit_twopoint_parametric_dominant |
| 4 | 7,865,562.00 | GSA-rs7376648 | 1.8638 | 22_bit_twopoint_parametric_recessive |
| 4 | 7,985,133.00 | rs7677869 | 2.771 | 22_bit_twopoint_parametric_recessive |
| 4 | 7,991,059.00 | rs4696727 | 1.8977 | 22_bit_twopoint_parametric_recessive |
| 4 | 8,565,578.00 | rs13108246 | 2.1531 | 22_bit_twopoint_parametric_dominant |
| 4 | 8,735,473.00 | JHU_4.8737198 | 1.9275 | 22_bit_twopoint_parametric_recessive |
| 4 | 8,878,494.00 | rs78923138 | 2.0527 | 22_bit_twopoint_parametric_dominant |
| 4 | 8,878,494.00 | rs78923138 | 2.1964 | 22_bit_twopoint_parametric_recessive |
| 4 | 10,697,503.00 | JHU_4.10699126 | 1.9098 | 21_bit_twopoint_parametric_dominant |
| 4 | 10,697,503.00 | JHU_4.10699126 | 2.3562 | 21_bit_twopoint_parametric_recessive |
| 4 | 11,540,164.00 | JHU_4.11541787 | 1.9812 | 21_bit_twopoint_parametric_recessive |
| 4 | 11,980,248.00 | rs13133185 | 2.1221 | 22_bit_twopoint_parametric_recessive |
| 4 | 12,605,299.00 | rs7680227 | 2.6308 | 22_bit_twopoint_parametric_dominant |
| 4 | 14,381,799.00 | JHU_4.14383422 | 2.4901 | 22_bit_twopoint_parametric_recessive |
| 4 | 14,383,713.00 | rs717355 | 2.4467 | 22_bit_twopoint_parametric_recessive |
| 4 | 14,383,713.00 | rs717355 | 2.448 | 22_bit_multipoint_parametric_recessive |
| 4 | 14,937,765.00 | rs34479500 | 2.3772 | 22_bit_twopoint_parametric_recessive |
| 4 | 14,937,765.00 | rs34479500 | 2.3772 | 22_bit_twopoint_parametric_recessive |
| 4 | 15,101,263.00 | rs1455208 | 2.1986 | 22_bit_twopoint_parametric_recessive |
| 4 | 15,101,263.00 | rs1455208 | 2.1986 | 22_bit_twopoint_parametric_recessive |
| 4 | 15,485,808.00 | rs4698112 | 2.2168 | 22_bit_twopoint_parametric_recessive |
| 4 | 15,485,808.00 | rs4698112 | 2.2168 | 22_bit_twopoint_parametric_recessive |
| 4 | 15,734,884.00 | rs955411 | 2.622 | 22_bit_twopoint_parametric_dominant |
| 4 | 15,734,884.00 | rs955411 | 2.622 | 22_bit_twopoint_parametric_dominant |
| 4 | 15,737,875.00 | rs997250 | 2.125 | 22_bit_twopoint_parametric_dominant |
| 4 | 15,737,875.00 | rs997250 | 2.125 | 22_bit_twopoint_parametric_dominant |
| 4 | 15,737,875.00 | rs997250 | 2.4092 | 22_bit_twopoint_parametric_recessive |
| 4 | 15,737,875.00 | rs997250 | 2.4092 | 22_bit_twopoint_parametric_recessive |
| 4 | 15,941,459.00 | JHU_4.15943081 | 2.0506 | 22_bit_twopoint_parametric_recessive |
| 4 | 16,143,965.00 | rs11730743 | 1.941 | 21_bit_twopoint_parametric_dominant |
| 4 | 16,596,887.00 | GSA-rs2173074 | 1.9559 | 21_bit_twopoint_parametric_recessive |
| 4 | 17,274,681.00 | GSA-rs12507442 | 2.4375 | 22_bit_twopoint_parametric_recessive |
| 4 | 17,296,384.00 | rs7690593 | 1.9888 | 21_bit_twopoint_parametric_recessive |
| 4 | 17,329,137.00 | rs13107521 | 2.015 | 21_bit_twopoint_NPL |
| 4 | 17,329,137.00 | rs13107521 | 2.0823 | 22_bit_twopoint_parametric_recessive |
| 4 | 17,329,137.00 | rs13107521 | 2.9557 | 21_bit_twopoint_parametric_dominant |
| 4 | 17,365,081.00 | rs17513992 | 2.5017 | 21_bit_twopoint_parametric_recessive |
| 4 | 17,383,928.00 | rs2122574 | 2.9348 | 22_bit_twopoint_parametric_recessive |
| 4 | 17,688,449.00 | GSA-rs61741063 | 2.4391 | 22_bit_twopoint_parametric_recessive |
| 4 | 18,129,622.00 | rs1503872 | 1.9883 | 21_bit_twopoint_parametric_dominant |
| 4 | 18,743,493.00 | GSA-rs78678947 | 2.0883 | 22_bit_twopoint_parametric_dominant |
| 4 | 18,743,493.00 | GSA-rs78678947 | 2.49 | 22_bit_twopoint_parametric_recessive |
| 4 | 19,005,140.00 | rs929587 | 2.2288 | 22_bit_twopoint_parametric_dominant |
| 4 | 19,408,554.00 | GSA-rs10023012 | 1.9209 | 22_bit_twopoint_parametric_recessive |
| 4 | 19,631,715.00 | rs3846388 | 1.9319 | 22_bit_twopoint_parametric_recessive |
| 4 | 19,716,002.00 | rs9291405 | 1.9426 | 22_bit_twopoint_parametric_recessive |
| 4 | 19,831,372.00 | rs4261984 | 2.4715 | 22_bit_twopoint_parametric_dominant |
| 4 | 19,831,372.00 | rs4261984 | 2.4756 | 22_bit_twopoint_parametric_recessive |
| 4 | 19,939,345.00 | JHU_4.19940967 | 2.3352 | 22_bit_twopoint_parametric_recessive |
| 4 | 20,283,892.00 | rs76498824 | 2.2748 | 22_bit_twopoint_parametric_recessive |
| 4 | 21,454,254.00 | rs1460485 | 1.9549 | 22_bit_twopoint_parametric_recessive |
| 4 | 21,454,254.00 | rs1460485 | 2.9461 | 22_bit_twopoint_parametric_dominant |
| 4 | 21,979,430.00 | rs6827204 | 2.3167 | 22_bit_twopoint_parametric_recessive |
| 4 | 22,133,701.00 | rs16872414 | 2.0294 | 22_bit_twopoint_parametric_recessive |
| 4 | 22,300,729.00 | GSA-rs1392592 | 1.8872 | 22_bit_twopoint_parametric_recessive |
| 4 | 22,300,729.00 | GSA-rs1392592 | 1.9724 | 21_bit_twopoint_parametric_recessive |
| 4 | 22,792,978.00 | rs358254 | 1.9682 | 21_bit_twopoint_parametric_dominant |
| 4 | 23,446,043.00 | JHU_4.23447665 | 2.0971 | 22_bit_twopoint_parametric_recessive |
| 4 | 23,950,225.00 | rs73243633 | 1.9324 | 22_bit_twopoint_parametric_dominant |
| 4 | 24,047,088.00 | rs625365 | 2.0416 | 21_bit_twopoint_parametric_recessive |
| 4 | 24,050,199.00 | JHU_4.24051821 | 2.2853 | 21_bit_twopoint_parametric_recessive |
| 4 | 24,228,700.00 | rs6448251 | 1.9138 | 21_bit_twopoint_parametric_dominant |
| 4 | 24,228,700.00 | rs6448251 | 2.0546 | 22_bit_twopoint_parametric_recessive |
| 4 | 24,397,079.00 | rs11723070 | 2.3471 | 22_bit_twopoint_parametric_recessive |
| 4 | 24,397,079.00 | rs11723070 | 2.3471 | 22_bit_twopoint_parametric_recessive |
| 4 | 24,398,535.00 | GSA-rs1989170 | 2.1932 | 22_bit_twopoint_parametric_recessive |
| 4 | 24,398,535.00 | GSA-rs1989170 | 2.1932 | 22_bit_twopoint_parametric_recessive |
| 4 | 24,734,635.00 | rs1476880 | 2.0504 | 21_bit_twopoint_parametric_recessive |
| 4 | 24,734,635.00 | rs1476880 | 2.0504 | 21_bit_twopoint_parametric_recessive |
| 4 | 24,799,693.00 | exm392513 | 2.1488 | 21_bit_twopoint_parametric_recessive |
| 4 | 24,799,693.00 | exm392513 | 2.1488 | 21_bit_twopoint_parametric_recessive |
| 4 | 24,887,527.00 | rs11732285 | 2.1536 | 21_bit_twopoint_parametric_dominant |
| 4 | 24,887,527.00 | rs11732285 | 2.1536 | 21_bit_twopoint_parametric_dominant |
| 4 | 25,039,054.00 | rs4235328 | 2.141 | 22_bit_twopoint_parametric_recessive |
| 4 | 25,039,054.00 | rs4235328 | 2.141 | 22_bit_twopoint_parametric_recessive |
| 4 | 25,039,891.00 | GSA-rs2324649 | 2.11 | 22_bit_twopoint_parametric_recessive |
| 4 | 25,039,891.00 | GSA-rs2324649 | 2.11 | 22_bit_twopoint_parametric_recessive |
| 4 | 25,046,914.00 | GSA-rs13124493 | 2.4227 | 22_bit_multipoint_parametric_dominant |
| 4 | 25,046,914.00 | GSA-rs13124493 | 2.4489 | 22_bit_twopoint_parametric_dominant |
| 4 | 25,046,914.00 | GSA-rs13124493 | 2.4489 | 22_bit_twopoint_parametric_dominant |
| 4 | 25,453,782.00 | rs922054 | 2.0061 | 22_bit_twopoint_parametric_recessive |
| 4 | 25,620,389.00 | rs11728892 | 2.1431 | 22_bit_twopoint_parametric_dominant |
| 4 | 25,835,688.00 | rs6847559 | 2.8152 | 22_bit_twopoint_parametric_dominant |
| 4 | 26,017,068.00 | rs6847589 | 2.0176 | 22_bit_twopoint_parametric_recessive |
| 4 | 26,842,847.00 | rs7688662 | 1.9534 | 21_bit_twopoint_parametric_recessive |
| 4 | 26,850,816.00 | rs7660174 | 1.9603 | 22_bit_twopoint_parametric_recessive |
| 4 | 26,850,816.00 | rs7660174 | 2.4894 | 21_bit_twopoint_parametric_recessive |
| 4 | 27,084,444.00 | rs13144846 | 2.0138 | 22_bit_twopoint_parametric_recessive |
| 4 | 27,084,444.00 | rs13144846 | 2.0514 | 21_bit_twopoint_parametric_recessive |
| 4 | 27,124,491.00 | JHU_4.27126112 | 3.2336 | 21_bit_twopoint_parametric_dominant |
| 4 | 27,673,788.00 | rs6846226 | 2.5993 | 22_bit_twopoint_parametric_recessive |
| 4 | 27,831,877.00 | rs6832303 | 2.6629 | 22_bit_twopoint_parametric_recessive |
| 4 | 29,363,058.00 | rs195444 | 1.8621 | 22_bit_twopoint_parametric_dominant |
| 4 | 31,161,845.00 | rs7685902 | 1.9361 | 21_bit_twopoint_parametric_recessive |
| 4 | 31,313,027.00 | rs4525934 | 2.4805 | 21_bit_twopoint_parametric_recessive |
| 4 | 35,247,225.00 | rs7686175 | 1.9669 | 22_bit_twopoint_parametric_dominant |
| 4 | 35,247,225.00 | rs7686175 | 1.9669 | 22_bit_twopoint_parametric_dominant |
| 4 | 35,247,225.00 | rs7686175 | 2.5613 | 22_bit_twopoint_parametric_recessive |
| 4 | 35,247,225.00 | rs7686175 | 2.5613 | 22_bit_twopoint_parametric_recessive |
| 4 | 35,911,270.00 | rs1030700 | 1.9137 | 22_bit_twopoint_parametric_recessive |
| 4 | 35,911,270.00 | rs1030700 | 1.9137 | 22_bit_twopoint_parametric_recessive |
| 4 | 36,285,659.00 | rs6854387 | 2.1844 | 22_bit_twopoint_parametric_dominant |
| 4 | 36,285,659.00 | rs6854387 | 2.1844 | 22_bit_twopoint_parametric_dominant |
| 4 | 37,788,637.00 | rs955868 | 2.1711 | 22_bit_twopoint_parametric_recessive |
| 4 | 38,108,679.00 | GSA-rs11096923 | 2.5843 | 21_bit_twopoint_parametric_recessive |
| 4 | 38,108,679.00 | GSA-rs11096923 | 2.8789 | 22_bit_twopoint_parametric_recessive |
| 4 | 38,122,295.00 | rs2067557 | 2.0295 | 21_bit_twopoint_parametric_recessive |
| 4 | 38,122,295.00 | rs2067557 | 2.7709 | 22_bit_twopoint_parametric_recessive |
| 4 | 38,189,490.00 | rs62295510 | 2.5275 | 22_bit_twopoint_parametric_recessive |
| 4 | 38,300,069.00 | rs2890608 | 2.2118 | 21_bit_twopoint_parametric_recessive |
| 4 | 38,929,374.00 | JHU_4.38930994 | 2.0311 | 22_bit_twopoint_parametric_recessive |
| 4 | 38,929,476.00 | rs13150445 | 2.0478 | 22_bit_twopoint_parametric_recessive |
| 4 | 38,960,524.00 | rs6837941 | 2.0921 | 21_bit_twopoint_parametric_dominant |
| 4 | 40,226,051.00 | rs11729556 | 2.0365 | 21_bit_twopoint_parametric_recessive |
| 4 | 40,226,051.00 | rs11729556 | 2.2265 | 22_bit_twopoint_parametric_recessive |
| 4 | 40,226,051.00 | rs11729556 | 2.8878 | 22_bit_twopoint_parametric_dominant |
| 4 | 41,179,144.00 | GSA-rs7659645 | 2.2965 | 22_bit_twopoint_parametric_recessive |
| 4 | 41,239,650.00 | rs62410048 | 1.9672 | 21_bit_twopoint_parametric_recessive |
| 4 | 41,287,988.00 | rs57896726 | 1.9725 | 21_bit_twopoint_parametric_recessive |
| 4 | 41,344,791.00 | rs11721727 | 2.0505 | 21_bit_twopoint_parametric_dominant |
| 4 | 42,193,360.00 | rs34809764 | 1.8764 | 22_bit_twopoint_parametric_recessive |
| 4 | 42,649,987.00 | exm2256615 | 1.9971 | 21_bit_twopoint_parametric_recessive |
| 4 | 42,712,609.00 | GSA-rs988600 | 2.519 | 21_bit_twopoint_parametric_recessive |
| 4 | 43,482,233.00 | rs7694215 | 1.8734 | 21_bit_twopoint_parametric_dominant |
| 4 | 43,482,233.00 | rs7694215 | 2.1454 | 22_bit_twopoint_parametric_recessive |
| 4 | 43,482,233.00 | rs7694215 | 2.456 | 21_bit_twopoint_parametric_recessive |
| 4 | 43,518,573.00 | rs4861284 | 2.7931 | 22_bit_twopoint_parametric_recessive |
| 4 | 43,524,791.00 | GSA-rs9992804 | 1.9353 | 21_bit_twopoint_parametric_dominant |
| 4 | 43,524,791.00 | GSA-rs9992804 | 2.4038 | 22_bit_twopoint_parametric_recessive |
| 4 | 43,524,791.00 | GSA-rs9992804 | 2.8732 | 21_bit_twopoint_parametric_recessive |
| 4 | 43,562,814.00 | rs6816987 | 2.0834 | 22_bit_twopoint_parametric_recessive |
| 4 | 43,562,814.00 | rs6816987 | 2.5663 | 21_bit_twopoint_parametric_recessive |
| 4 | 43,593,429.00 | GSA-rs6447283 | 2.0285 | 21_bit_twopoint_parametric_dominant |
| 4 | 43,593,429.00 | GSA-rs6447283 | 2.7054 | 22_bit_twopoint_parametric_recessive |
| 4 | 43,593,429.00 | GSA-rs6447283 | 3.0028 | 21_bit_twopoint_parametric_recessive |
| 4 | 43,602,104.00 | rs77999098 | 2.5274 | 22_bit_twopoint_parametric_recessive |
| 4 | 43,602,104.00 | rs77999098 | 2.9673 | 21_bit_twopoint_parametric_recessive |
| 4 | 43,664,122.00 | rs13137462 | 1.905 | 21_bit_twopoint_parametric_dominant |
| 4 | 43,681,990.00 | rs6842669 | 3.1106 | 21_bit_twopoint_parametric_recessive |
| 4 | 43,681,990.00 | rs6842669 | 3.1866 | 22_bit_twopoint_parametric_recessive |
| 4 | 44,930,234.00 | GSA-rs1390911 | 1.912 | 21_bit_twopoint_parametric_recessive |
| 4 | 44,930,234.00 | GSA-rs1390911 | 1.912 | 21_bit_twopoint_parametric_recessive |
| 4 | 45,173,674.00 | rs13130484 | 1.9615 | 21_bit_twopoint_parametric_recessive |
| 4 | 45,173,674.00 | rs13130484 | 1.9615 | 21_bit_twopoint_parametric_recessive |
| 4 | 45,173,787.00 | rs16858082 | 2.4577 | 21_bit_twopoint_parametric_recessive |
| 4 | 45,173,787.00 | rs16858082 | 2.4577 | 21_bit_twopoint_parametric_recessive |
| 4 | 45,182,425.00 | rs348495 | 2.5192 | 21_bit_twopoint_parametric_recessive |
| 4 | 45,182,425.00 | rs348495 | 2.5192 | 21_bit_twopoint_parametric_recessive |
| 4 | 47,099,206.00 | GSA-rs1866990 | 2.0929 | 22_bit_twopoint_parametric_recessive |
| 4 | 47,101,098.00 | rs971354 | 2.8012 | 22_bit_twopoint_parametric_dominant |
| 4 | 47,490,096.00 | rs10938488 | 1.995 | 21_bit_twopoint_parametric_recessive |
| 4 | 55,906,554.00 | rs4352536 | 2.6081 | 21_bit_twopoint_parametric_recessive |
| 4 | 55,906,554.00 | rs4352536 | 2.8438 | 22_bit_twopoint_parametric_recessive |
| 4 | 55,928,791.00 | rs1133506 | 2.6187 | 22_bit_twopoint_parametric_recessive |
| 4 | 56,044,892.00 | rs2702334 | 2.0686 | 22_bit_twopoint_parametric_recessive |
| 4 | 56,260,212.00 | rs598210 | 1.8768 | 21_bit_twopoint_parametric_dominant |
| 4 | 56,267,797.00 | JHU_4.57133962 | 2.0276 | 21_bit_twopoint_parametric_dominant |
| 4 | 56,758,743.00 | rs1564400 | 2.3923 | 22_bit_twopoint_parametric_recessive |
| 4 | 56,830,752.00 | rs2127495 | 2.3174 | 21_bit_twopoint_parametric_dominant |
| 4 | 56,972,417.00 | rs17087335 | 2.1973 | 21_bit_twopoint_parametric_recessive |
| 4 | 57,804,828.00 | rs951232 | 2.473 | 22_bit_twopoint_parametric_recessive |
| 4 | 57,804,828.00 | rs951232 | 2.6585 | 21_bit_twopoint_parametric_recessive |
| 4 | 58,849,252.00 | rs9998639 | 2.1811 | 22_bit_twopoint_parametric_recessive |
| 4 | 58,925,999.00 | JHU_4.59791665 | 2.1106 | 22_bit_twopoint_parametric_recessive |
| 4 | 59,133,545.00 | GSA-rs79222519 | 1.8654 | 21_bit_twopoint_parametric_recessive |
| 4 | 59,133,545.00 | GSA-rs79222519 | 1.8654 | 21_bit_twopoint_parametric_recessive |
| 4 | 59,263,450.00 | rs7663467 | 2.0242 | 21_bit_twopoint_parametric_recessive |
| 4 | 59,263,450.00 | rs7663467 | 2.0242 | 21_bit_twopoint_parametric_recessive |
| 4 | 59,263,450.00 | rs7663467 | 2.4264 | 22_bit_twopoint_parametric_recessive |
| 4 | 59,263,450.00 | rs7663467 | 2.4264 | 22_bit_twopoint_parametric_recessive |
| 4 | 62,415,997.00 | rs17828720 | 1.9767 | 22_bit_twopoint_parametric_recessive |
| 4 | 62,627,586.00 | rs11940300 | 2.0581 | 22_bit_twopoint_parametric_dominant |
| 4 | 62,973,078.00 | rs10025992 | 2.4632 | 22_bit_twopoint_parametric_dominant |
| 4 | 63,051,469.00 | JHU_4.63917186 | 1.9265 | 21_bit_twopoint_parametric_recessive |
| 4 | 63,089,548.00 | GSA-rs111732351 | 2.0373 | 21_bit_twopoint_parametric_recessive |
| 4 | 63,231,948.00 | JHU_4.64097665 | 1.8735 | 21_bit_twopoint_parametric_recessive |
| 4 | 64,882,949.00 | GSA-rs6849205 | 1.9362 | 22_bit_twopoint_parametric_dominant |
| 4 | 64,990,793.00 | JHU_4.65856510 | 1.9933 | 22_bit_twopoint_parametric_recessive |
| 4 | 65,037,879.00 | GSA-rs13152245 | 2.0799 | 22_bit_twopoint_parametric_recessive |
| 4 | 67,852,768.00 | rs1872581 | 1.9326 | 22_bit_twopoint_parametric_recessive |
| 4 | 69,103,288.00 | rs7439152 | 1.8643 | 21_bit_twopoint_parametric_recessive |
| 4 | 69,589,760.00 | GSA-rs2288741 | 2.6576 | 21_bit_twopoint_parametric_recessive |
| 4 | 69,619,909.00 | rs6824830 | 3.1272 | 21_bit_twopoint_parametric_recessive |
| 4 | 69,647,992.00 | GSA-rs1432329 | 2.9596 | 21_bit_twopoint_parametric_recessive |
| 4 | 70,133,902.00 | rs1399252 | 1.9913 | 22_bit_twopoint_parametric_dominant |
| 4 | 70,485,853.00 | JHU_4.71351569 | 2.3507 | 21_bit_twopoint_parametric_dominant |
| 4 | 70,589,968.00 | rs28660083 | 2.3951 | 21_bit_twopoint_parametric_dominant |
| 4 | 71,838,702.00 | GSA-rs1491714 | 1.9776 | 21_bit_twopoint_parametric_recessive |
| 4 | 71,838,702.00 | GSA-rs1491714 | 1.9776 | 21_bit_twopoint_parametric_recessive |
| 4 | 71,926,132.00 | rs1520499 | 2.0848 | 21_bit_twopoint_parametric_recessive |
| 4 | 71,926,132.00 | rs1520499 | 2.0848 | 21_bit_twopoint_parametric_recessive |
| 4 | 72,857,260.00 | rs6822170 | 1.8735 | 21_bit_twopoint_parametric_recessive |
| 4 | 72,857,260.00 | rs6822170 | 1.8735 | 21_bit_twopoint_parametric_recessive |
| 4 | 72,857,260.00 | rs6822170 | 1.9478 | 22_bit_twopoint_parametric_recessive |
| 4 | 72,857,260.00 | rs6822170 | 1.9478 | 22_bit_twopoint_parametric_recessive |
| 4 | 76,238,547.00 | rs1441922 | 2.4531 | 22_bit_twopoint_parametric_recessive |
| 4 | 78,236,974.00 | rs11725082 | 2.5339 | 21_bit_twopoint_parametric_dominant |
| 4 | 79,624,561.00 | rs1371990 | 2.0059 | 22_bit_twopoint_parametric_recessive |
| 4 | 79,928,220.00 | JHU_4.80849373 | 2.0893 | 21_bit_twopoint_parametric_dominant |
| 4 | 81,766,369.00 | rs4693005 | 1.8626 | 21_bit_twopoint_parametric_dominant |
| 4 | 81,799,178.00 | rs523329 | 2.3869 | 21_bit_twopoint_parametric_dominant |
| 4 | 81,831,568.00 | rs685815 | 2.0825 | 22_bit_twopoint_parametric_dominant |
| 4 | 81,890,940.00 | rs7663795 | 1.9434 | 21_bit_twopoint_parametric_dominant |
| 4 | 87,612,388.00 | exm411985 | 2.1801 | 21_bit_twopoint_parametric_recessive |
| 4 | 87,612,388.00 | exm411985 | 3.0694 | 22_bit_twopoint_parametric_recessive |
| 4 | 87,981,540.00 | rs4754 | 2.6378 | 21_bit_twopoint_parametric_dominant |
| 4 | 87,992,786.00 | JHU_4.88913937 | 2.6378 | 21_bit_twopoint_parametric_dominant |
| 4 | 88,030,924.00 | rs2725225 | 2.4157 | 22_bit_twopoint_parametric_dominant |
| 4 | 88,249,578.00 | rs17013965 | 2.673 | 22_bit_twopoint_parametric_recessive |
| 4 | 89,757,390.00 | exm-rs2736990 | 1.908 | 21_bit_twopoint_parametric_dominant |
| 4 | 90,104,699.00 | rs34001515 | 1.9181 | 22_bit_twopoint_parametric_dominant |
| 4 | 94,623,011.00 | rs4699312 | 1.9598 | 22_bit_twopoint_parametric_dominant |
| 4 | 100,853,044.00 | GSA-rs6847657 | 2.1495 | 22_bit_twopoint_parametric_dominant |
| 4 | 104,163,654.00 | GSA-rs795135 | 2.0395 | 21_bit_twopoint_parametric_dominant |
| 4 | 109,096,050.00 | rs4632736 | 1.9006 | 21_bit_twopoint_parametric_dominant |
| 4 | 110,832,659.00 | rs3866838 | 2.22 | 22_bit_twopoint_parametric_dominant |
| 4 | 110,840,566.00 | rs56339597 | 2.22 | 22_bit_twopoint_parametric_dominant |
| 4 | 110,878,261.00 | rs6843250 | 2.0083 | 21_bit_twopoint_parametric_dominant |
| 4 | 113,047,902.00 | JHU_4.113969057 | 1.9318 | 22_bit_twopoint_parametric_dominant |
| 4 | 113,054,962.00 | rs35308370 | 1.861 | 21_bit_multipoint_NPL |
| 4 | 113,159,084.00 | rs17590572 | 2.3687 | 21_bit_twopoint_parametric_dominant |
| 4 | 117,868,560.00 | rs7696793 | 1.9488 | 22_bit_twopoint_parametric_recessive |
| 4 | 117,868,560.00 | rs7696793 | 2.3037 | 22_bit_twopoint_parametric_dominant |
| 4 | 122,660,926.00 | rs6825988 | 1.9916 | 22_bit_twopoint_parametric_dominant |
| 4 | 122,680,542.00 | GSA-rs6836610 | 2.0053 | 22_bit_twopoint_parametric_dominant |
| 4 | 125,445,779.00 | rs17009673 | 2.8151 | 21_bit_twopoint_parametric_dominant |
| 4 | 132,175,788.00 | rs2135175 | 2.0862 | 22_bit_twopoint_parametric_recessive |
| 4 | 134,125,081.00 | rs1494984 | 2.233 | 22_bit_twopoint_parametric_recessive |
| 4 | 134,199,911.00 | exm424839 | 2.233 | 22_bit_twopoint_parametric_recessive |
| 4 | 134,200,446.00 | exm424850 | 2.233 | 22_bit_twopoint_parametric_recessive |
| 4 | 135,570,389.00 | rs931121 | 1.872 | 21_bit_twopoint_parametric_dominant |
| 4 | 135,570,389.00 | rs931121 | 2.3013 | 21_bit_twopoint_parametric_recessive |
| 4 | 135,620,307.00 | rs72713003 | 1.9577 | 21_bit_twopoint_parametric_recessive |
| 4 | 136,532,868.00 | rs4315856 | 2.6133 | 22_bit_twopoint_parametric_dominant |
| 4 | 136,532,868.00 | rs4315856 | 2.6133 | 22_bit_twopoint_parametric_dominant |
| 4 | 136,559,338.00 | rs11733040 | 2.3729 | 22_bit_twopoint_parametric_dominant |
| 4 | 136,559,338.00 | rs11733040 | 2.3729 | 22_bit_twopoint_parametric_dominant |
| 4 | 137,474,628.00 | rs12643295 | 2.4873 | 22_bit_twopoint_parametric_dominant |
| 4 | 137,769,969.00 | rs7675201 | 2.5094 | 22_bit_twopoint_parametric_recessive |
| 4 | 139,616,579.00 | rs2602233 | 1.918 | 21_bit_twopoint_parametric_recessive |
| 4 | 139,616,579.00 | rs2602233 | 2.4017 | 21_bit_twopoint_parametric_dominant |
| 4 | 142,976,517.00 | rs4690741 | 2.1813 | 22_bit_twopoint_parametric_recessive |
| 4 | 144,234,103.00 | rs6537265 | 1.908 | 21_bit_twopoint_parametric_recessive |
| 4 | 151,373,577.00 | rs6535791 | 2.9507 | 22_bit_twopoint_parametric_dominant |
| 4 | 151,454,651.00 | GSA-rs6845067 | 3.2983 | 22_bit_twopoint_parametric_dominant |
| 4 | 151,566,923.00 | GSA-rs7695412 | 2.0091 | 21_bit_twopoint_parametric_dominant |
| 4 | 151,669,185.00 | GSA-rs17027780 | 1.9424 | 21_bit_twopoint_parametric_dominant |
| 4 | 151,689,166.00 | GSA-rs6833277 | 2.1965 | 22_bit_twopoint_parametric_dominant |
| 4 | 151,689,797.00 | GSA-rs10009423 | 2.2369 | 22_bit_twopoint_parametric_dominant |
| 4 | 151,699,998.00 | rs4696288 | 3.1997 | 21_bit_twopoint_parametric_dominant |
| 4 | 151,760,894.00 | exm428747 | 2.1957 | 22_bit_twopoint_parametric_dominant |
| 4 | 151,790,820.00 | rs6833558 | 2.1962 | 22_bit_twopoint_parametric_dominant |
| 4 | 151,806,147.00 | rs11099829 | 2.1504 | 21_bit_twopoint_parametric_dominant |
| 4 | 151,976,909.00 | rs360912 | 2.269 | 22_bit_twopoint_parametric_dominant |
| 4 | 153,132,106.00 | rs4323085 | 1.9664 | 21_bit_twopoint_parametric_dominant |
| 4 | 153,269,813.00 | rs72729610 | 1.8642 | 22_bit_twopoint_parametric_dominant |
| 4 | 154,014,422.00 | rs6824634 | 2.2996 | 22_bit_twopoint_parametric_dominant |
| 4 | 155,634,698.00 | rs79716105 | 2.3714 | 22_bit_twopoint_parametric_dominant |
| 4 | 155,806,225.00 | rs17033572 | 1.9109 | 21_bit_twopoint_parametric_dominant |
| 4 | 155,944,334.00 | rs10019161 | 1.9314 | 22_bit_twopoint_parametric_dominant |
| 4 | 157,857,687.00 | rs7693369 | 2.1026 | 21_bit_twopoint_parametric_recessive |
| 4 | 158,396,290.00 | rs10857312 | 2.0338 | 21_bit_twopoint_parametric_dominant |
| 4 | 158,575,554.00 | rs62351164 | 2.7051 | 21_bit_twopoint_parametric_dominant |
| 4 | 161,842,862.00 | rs1440588 | 2.2345 | 22_bit_twopoint_parametric_dominant |
| 4 | 162,328,145.00 | JHU_4.163249296 | 2.5051 | 21_bit_twopoint_parametric_dominant |
| 4 | 162,622,793.00 | rs6849368 | 2.634 | 22_bit_twopoint_parametric_dominant |
| 4 | 162,659,119.00 | GSA-rs195937 | 2.6989 | 22_bit_twopoint_parametric_dominant |
| 4 | 163,649,779.00 | JHU_4.164570930 | 2.0077 | 22_bit_twopoint_parametric_recessive |
| 4 | 165,729,292.00 | rs1466351 | 2.1034 | 21_bit_twopoint_parametric_dominant |
| 4 | 165,882,376.00 | GSA-rs9637646 | 2.0968 | 22_bit_twopoint_parametric_dominant |
| 4 | 168,417,369.00 | rs1963569 | 2.0414 | 22_bit_multipoint_parametric_dominant |
| 4 | 168,417,369.00 | rs1963569 | 2.0904 | 22_bit_twopoint_parametric_dominant |
| 4 | 168,498,698.00 | GSA-rs2712135 | 1.9804 | 22_bit_twopoint_parametric_dominant |
| 4 | 168,542,983.00 | rs6836618 | 2.3816 | 21_bit_twopoint_parametric_dominant |
| 4 | 168,609,188.00 | rs4389538 | 2.0788 | 22_bit_twopoint_parametric_dominant |
| 4 | 168,675,498.00 | rs13137200 | 1.8744 | 22_bit_twopoint_parametric_dominant |
| 4 | 171,552,299.00 | exm2265795 | 2.123 | 21_bit_twopoint_parametric_dominant |
| 4 | 171,887,243.00 | GSA-rs6553597 | 1.9008 | 22_bit_twopoint_parametric_dominant |
| 4 | 171,887,243.00 | GSA-rs6553597 | 2.0657 | 21_bit_twopoint_parametric_dominant |
| 4 | 171,891,338.00 | rs4124044 | 1.8973 | 22_bit_twopoint_parametric_dominant |
| 4 | 173,041,274.00 | rs2019683 | 2.3809 | 22_bit_twopoint_parametric_recessive |
| 4 | 173,041,274.00 | rs2019683 | 2.7236 | 22_bit_twopoint_parametric_dominant |
| 4 | 173,743,414.00 | rs60258086 | 2.5105 | 21_bit_twopoint_parametric_recessive |
| 4 | 173,743,414.00 | rs60258086 | 3.0578 | 22_bit_twopoint_parametric_recessive |
| 4 | 176,078,836.00 | GSA-rs114546144 | 1.8939 | 22_bit_twopoint_parametric_dominant |
| 4 | 180,087,014.00 | rs1689004 | 1.8633 | 22_bit_twopoint_parametric_dominant |
| 4 | 183,537,886.00 | JHU_4.184459038 | 1.9354 | 22_bit_twopoint_parametric_dominant |
| 4 | 184,948,062.00 | rs4370185 | 1.911 | 21_bit_twopoint_parametric_recessive |
| 4 | 184,948,062.00 | rs4370185 | 2.0289 | 22_bit_twopoint_parametric_recessive |
| 4 | 184,956,289.00 | GSA-rs13143382 | 2.3926 | 21_bit_twopoint_parametric_recessive |
| 4 | 185,691,521.00 | rs4376189 | 2.2319 | 21_bit_twopoint_parametric_recessive |
| 4 | 185,714,356.00 | JHU_4.186635509 | 2.3542 | 21_bit_twopoint_parametric_recessive |
| 4 | 186,157,863.00 | rs1877321 | 3.0981 | 21_bit_twopoint_parametric_recessive |
| 4 | 186,264,231.00 | rs3756008 | 1.9054 | 21_bit_twopoint_parametric_recessive |
| 4 | 186,274,397.00 | rs1593 | 2.4778 | 22_bit_twopoint_parametric_recessive |
| 4 | 186,785,240.00 | rs325029 | 2.684 | 22_bit_twopoint_parametric_dominant |
| 4 | 186,786,644.00 | rs6847347 | 2.3721 | 22_bit_twopoint_parametric_dominant |
| 4 | 187,809,555.00 | GSA-rs10017989 | 2.1526 | 21_bit_twopoint_parametric_recessive |
| 4 | 187,813,461.00 | rs9996150 | 1.9872 | 21_bit_twopoint_parametric_recessive |
| 5 | 1,764,635.00 | rs2353591 | 2.2908 | 21_bit_twopoint_parametric_dominant |
| 5 | 1,915,681.00 | JHU_5.1915794 | 1.9903 | 21_bit_twopoint_parametric_dominant |
| 5 | 3,132,408.00 | rs468944 | 2.1957 | 21_bit_twopoint_parametric_dominant |
| 5 | 4,486,692.00 | rs73737898 | 2.105 | 22_bit_twopoint_parametric_recessive |
| 5 | 4,486,692.00 | rs73737898 | 2.4708 | 21_bit_twopoint_parametric_recessive |
| 5 | 4,520,107.00 | rs12517115 | 2.076 | 21_bit_twopoint_parametric_recessive |
| 5 | 4,520,107.00 | rs12517115 | 3.1826 | 21_bit_twopoint_parametric_dominant |
| 5 | 5,140,028.00 | rs270210 | 2.227 | 21_bit_twopoint_parametric_dominant |
| 5 | 5,914,704.00 | JHU_5.5914816 | 2.1567 | 22_bit_twopoint_parametric_dominant |
| 5 | 5,914,704.00 | JHU_5.5914816 | 2.1567 | 22_bit_twopoint_parametric_dominant |
| 5 | 6,676,818.00 | rs1651071 | 2.4414 | 21_bit_twopoint_parametric_recessive |
| 5 | 6,676,818.00 | rs1651071 | 2.5232 | 21_bit_twopoint_parametric_dominant |
| 5 | 6,772,480.00 | GSA-rs11947998 | 2.1059 | 22_bit_twopoint_parametric_dominant |
| 5 | 10,941,876.00 | rs852563 | 1.8867 | 22_bit_twopoint_parametric_recessive |
| 5 | 10,954,074.00 | rs860434 | 2.1842 | 21_bit_twopoint_parametric_recessive |
| 5 | 10,954,074.00 | rs860434 | 2.4593 | 22_bit_twopoint_parametric_recessive |
| 5 | 11,918,791.00 | rs7713607 | 2.0659 | 22_bit_twopoint_parametric_dominant |
| 5 | 11,918,791.00 | rs7713607 | 2.225 | 22_bit_twopoint_parametric_recessive |
| 5 | 13,149,565.00 | rs32534 | 2.3727 | 21_bit_twopoint_parametric_dominant |
| 5 | 14,902,855.00 | rs3006064 | 1.9699 | 22_bit_twopoint_parametric_dominant |
| 5 | 15,723,524.00 | GSA-rs11748790 | 2.0782 | 22_bit_twopoint_parametric_recessive |
| 5 | 17,254,917.00 | rs11133892 | 1.9158 | 21_bit_twopoint_parametric_dominant |
| 5 | 18,848,989.00 | JHU_5.18849097 | 2.7479 | 22_bit_twopoint_parametric_recessive |
| 5 | 21,629,319.00 | GSA-rs12658292 | 2.5272 | 22_bit_twopoint_parametric_recessive |
| 5 | 21,633,089.00 | rs77746646 | 2.1568 | 22_bit_twopoint_parametric_recessive |
| 5 | 21,637,306.00 | GSA-rs13161353 | 1.9094 | 22_bit_twopoint_parametric_dominant |
| 5 | 21,637,306.00 | GSA-rs13161353 | 2.9439 | 22_bit_twopoint_parametric_recessive |
| 5 | 21,674,531.00 | rs516235 | 1.9924 | 22_bit_twopoint_parametric_recessive |
| 5 | 21,676,560.00 | GSA-rs586654 | 2.3084 | 22_bit_twopoint_parametric_recessive |
| 5 | 22,755,365.00 | rs2119887 | 2.4327 | 21_bit_twopoint_parametric_dominant |
| 5 | 22,991,736.00 | rs1896710 | 1.8965 | 22_bit_twopoint_parametric_recessive |
| 5 | 23,484,263.00 | rs2934789 | 2.1542 | 22_bit_twopoint_parametric_dominant |
| 5 | 28,315,441.00 | JHU_5.28315547 | 2.4004 | 22_bit_twopoint_parametric_recessive |
| 5 | 30,916,400.00 | rs73757878 | 3.2325 | 21_bit_twopoint_parametric_recessive |
| 5 | 31,695,215.00 | rs10472779 | 1.868 | 21_bit_twopoint_NPL |
| 5 | 31,695,215.00 | rs10472779 | 1.868 | 21_bit_twopoint_NPL |
| 5 | 33,011,880.00 | rs4867131 | 2.1483 | 22_bit_twopoint_parametric_recessive |
| 5 | 33,336,169.00 | rs76162202 | 2.3536 | 21_bit_twopoint_parametric_recessive |
| 5 | 33,400,315.00 | JHU_5.33400420 | 2.856 | 22_bit_twopoint_parametric_dominant |
| 5 | 33,400,315.00 | JHU_5.33400420 | 3.0842 | 21_bit_twopoint_parametric_dominant |
| 5 | 33,410,601.00 | GSA-rs17550908 | 2.0532 | 21_bit_twopoint_parametric_dominant |
| 5 | 35,010,767.00 | rs11960801 | 1.8729 | 21_bit_twopoint_parametric_dominant |
| 5 | 35,209,237.00 | rs4235652 | 2.04 | 22_bit_twopoint_parametric_dominant |
| 5 | 35,254,507.00 | rs75977988 | 2.5281 | 22_bit_twopoint_parametric_dominant |
| 5 | 35,292,824.00 | rs78127368 | 2.0155 | 22_bit_twopoint_parametric_dominant |
| 5 | 36,157,713.00 | GSA-rs33678 | 1.9256 | 22_bit_twopoint_parametric_dominant |
| 5 | 36,157,713.00 | GSA-rs33678 | 1.9256 | 22_bit_twopoint_parametric_dominant |
| 5 | 36,163,368.00 | rs3815779 | 2.017 | 22_bit_twopoint_parametric_dominant |
| 5 | 36,163,368.00 | rs3815779 | 2.017 | 22_bit_twopoint_parametric_dominant |
| 5 | 41,564,460.00 | rs687174 | 3.1089 | 21_bit_twopoint_parametric_dominant |
| 5 | 41,603,338.00 | rs670365 | 1.923 | 22_bit_twopoint_NPL |
| 5 | 41,603,338.00 | rs670365 | 2.8327 | 22_bit_twopoint_parametric_recessive |
| 5 | 41,603,883.00 | JHU_5.41603984 | 1.9625 | 22_bit_twopoint_parametric_dominant |
| 5 | 41,603,883.00 | JHU_5.41603984 | 2.037 | 22_bit_twopoint_NPL |
| 5 | 41,603,883.00 | JHU_5.41603984 | 2.7339 | 22_bit_twopoint_parametric_recessive |
| 5 | 41,615,582.00 | JHU_5.41615683 | 1.9614 | 22_bit_twopoint_parametric_dominant |
| 5 | 41,615,582.00 | JHU_5.41615683 | 1.999 | 22_bit_twopoint_NPL |
| 5 | 41,615,582.00 | JHU_5.41615683 | 2.6574 | 22_bit_twopoint_parametric_recessive |
| 5 | 42,015,432.00 | rs80097998 | 2.5697 | 21_bit_twopoint_parametric_dominant |
| 5 | 61,185,581.00 | rs34609 | 2.4436 | 21_bit_twopoint_parametric_recessive |
| 5 | 67,110,315.00 | rs2545394 | 2.4458 | 22_bit_twopoint_parametric_dominant |
| 5 | 67,110,315.00 | rs2545394 | 2.4458 | 22_bit_twopoint_parametric_dominant |
| 5 | 68,949,463.00 | rs10056163 | 1.9089 | 22_bit_twopoint_parametric_dominant |
| 5 | 68,965,684.00 | JHU_5.68261510 | 1.9089 | 22_bit_twopoint_parametric_dominant |
| 5 | 71,374,944.00 | JHU_5.70670770 | 2.0812 | 22_bit_twopoint_parametric_dominant |
| 5 | 71,383,799.00 | kgp9926352 | 2.0647 | 22_bit_twopoint_parametric_dominant |
| 5 | 72,757,879.00 | rs6882763 | 1.8752 | 21_bit_twopoint_parametric_dominant |
| 5 | 73,113,560.00 | 5:72409387-G-A | 2.0386 | 21_bit_twopoint_parametric_dominant |
| 5 | 75,712,368.00 | GSA-rs2047059 | 2.562 | 21_bit_twopoint_parametric_recessive |
| 5 | 75,998,585.00 | rs10214163 | 1.9971 | 22_bit_twopoint_parametric_dominant |
| 5 | 77,035,624.00 | exm462394 | 2.044 | 21_bit_twopoint_parametric_recessive |
| 5 | 77,646,664.00 | rs1895254 | 1.9831 | 22_bit_twopoint_parametric_dominant |
| 5 | 81,113,707.00 | exm464793 | 2.0618 | 21_bit_twopoint_parametric_recessive |
| 5 | 81,113,707.00 | exm464793 | 2.7665 | 21_bit_twopoint_parametric_dominant |
| 5 | 81,212,851.00 | GSA-rs11958318 | 1.8632 | 21_bit_twopoint_parametric_dominant |
| 5 | 81,298,527.00 | rs10061143 | 2.2304 | 21_bit_twopoint_parametric_dominant |
| 5 | 81,302,186.00 | GSA-rs10052929 | 2.5908 | 21_bit_twopoint_parametric_dominant |
| 5 | 81,323,227.00 | rs10056565 | 2.1724 | 21_bit_twopoint_parametric_dominant |
| 5 | 82,673,044.00 | rs9791094 | 1.9648 | 21_bit_twopoint_parametric_dominant |
| 5 | 82,862,939.00 | rs12522257 | 2.8767 | 21_bit_twopoint_parametric_dominant |
| 5 | 83,003,254.00 | rs376689 | 1.9644 | 22_bit_twopoint_parametric_dominant |
| 5 | 83,054,991.00 | GSA-rs255561 | 2.9006 | 22_bit_twopoint_parametric_dominant |
| 5 | 83,076,846.00 | 5:82372665-G-T | 3.0549 | 22_bit_twopoint_parametric_dominant |
| 5 | 83,081,185.00 | rs1478486 | 2.8812 | 22_bit_twopoint_parametric_dominant |
| 5 | 83,662,984.00 | JHU_5.82958802 | 1.9763 | 22_bit_twopoint_parametric_dominant |
| 5 | 83,975,811.00 | rs72774740 | 1.8891 | 22_bit_twopoint_parametric_dominant |
| 5 | 83,975,811.00 | rs72774740 | 2.1501 | 22_bit_twopoint_parametric_recessive |
| 5 | 84,080,563.00 | rs1559064 | 2.2584 | 21_bit_twopoint_parametric_recessive |
| 5 | 86,019,682.00 | JHU_5.85315499 | 1.968 | 22_bit_twopoint_parametric_recessive |
| 5 | 86,019,682.00 | JHU_5.85315499 | 2.5488 | 21_bit_twopoint_parametric_recessive |
| 5 | 97,183,088.00 | GSA-rs3734010 | 1.9432 | 21_bit_twopoint_parametric_recessive |
| 5 | 105,384,997.00 | GSA-rs13153357 | 2.3354 | 22_bit_twopoint_parametric_dominant |
| 5 | 105,477,051.00 | rs4703342 | 2.398 | 22_bit_twopoint_parametric_dominant |
| 5 | 105,501,572.00 | GSA-rs6596615 | 2.396 | 22_bit_twopoint_parametric_dominant |
| 5 | 105,511,996.00 | GSA-rs28593492 | 2.398 | 22_bit_twopoint_parametric_dominant |
| 5 | 106,972,307.00 | rs4382150 | 2.0971 | 21_bit_twopoint_parametric_recessive |
| 5 | 106,972,307.00 | rs4382150 | 2.6805 | 21_bit_twopoint_parametric_dominant |
| 5 | 106,972,307.00 | rs4382150 | 2.9216 | 22_bit_twopoint_parametric_recessive |
| 5 | 107,030,138.00 | rs4073517 | 1.9774 | 22_bit_twopoint_parametric_dominant |
| 5 | 107,030,138.00 | rs4073517 | 2.1033 | 21_bit_twopoint_parametric_dominant |
| 5 | 107,030,138.00 | rs4073517 | 2.1935 | 21_bit_twopoint_parametric_recessive |
| 5 | 109,287,918.00 | rs2416205 | 2.5618 | 22_bit_twopoint_parametric_dominant |
| 5 | 109,287,918.00 | rs2416205 | 2.5677 | 22_bit_twopoint_parametric_recessive |
| 5 | 111,560,277.00 | rs247544 | 2.3605 | 21_bit_twopoint_parametric_recessive |
| 5 | 111,560,277.00 | rs247544 | 2.5122 | 22_bit_twopoint_parametric_recessive |
| 5 | 112,110,768.00 | JHU_5.111446464 | 1.8799 | 21_bit_twopoint_parametric_recessive |
| 5 | 113,013,600.00 | exm2273410 | 1.9431 | 21_bit_twopoint_parametric_recessive |
| 5 | 114,391,352.00 | rs13187753 | 1.9073 | 22_bit_twopoint_parametric_recessive |
| 5 | 114,391,352.00 | rs13187753 | 2.0087 | 21_bit_twopoint_parametric_recessive |
| 5 | 116,941,141.00 | rs4920862 | 1.8859 | 22_bit_twopoint_parametric_recessive |
| 5 | 116,941,141.00 | rs4920862 | 2.1969 | 21_bit_twopoint_parametric_recessive |
| 5 | 118,018,671.00 | rs10037996 | 2.0853 | 21_bit_twopoint_parametric_dominant |
| 5 | 119,181,218.00 | rs7727906 | 1.9037 | 22_bit_twopoint_parametric_dominant |
| 5 | 119,347,094.00 | rs6878879 | 2.3892 | 22_bit_twopoint_parametric_recessive |
| 5 | 119,356,193.00 | GSA-rs3797343 | 2.5653 | 22_bit_twopoint_parametric_recessive |
| 5 | 119,564,907.00 | rs92631 | 1.8637 | 21_bit_twopoint_parametric_recessive |
| 5 | 119,564,907.00 | rs92631 | 2.0621 | 21_bit_twopoint_parametric_dominant |
| 5 | 123,271,329.00 | rs255628 | 2.0576 | 21_bit_twopoint_parametric_dominant |
| 5 | 123,271,329.00 | rs255628 | 2.2608 | 21_bit_twopoint_parametric_recessive |
| 5 | 124,662,814.00 | JHU_5.123998506 | 1.8786 | 22_bit_twopoint_parametric_recessive |
| 5 | 124,673,164.00 | rs28611427 | 1.8724 | 22_bit_twopoint_parametric_recessive |
| 5 | 126,946,554.00 | rs11949302 | 2.8423 | 21_bit_twopoint_parametric_dominant |
| 5 | 127,134,238.00 | GSA-rs6884988 | 2.9138 | 21_bit_twopoint_parametric_dominant |
| 5 | 127,139,880.00 | rs13178353 | 1.8658 | 21_bit_twopoint_parametric_dominant |
| 5 | 127,811,690.00 | rs10051423 | 1.8732 | 22_bit_twopoint_parametric_dominant |
| 5 | 127,811,690.00 | rs10051423 | 2.1846 | 21_bit_twopoint_parametric_dominant |
| 5 | 127,816,284.00 | rs1421746 | 2.226 | 22_bit_twopoint_parametric_dominant |
| 5 | 127,816,284.00 | rs1421746 | 2.5622 | 21_bit_twopoint_parametric_dominant |
| 5 | 132,872,671.00 | rs803223 | 1.9455 | 21_bit_twopoint_parametric_recessive |
| 5 | 133,225,776.00 | exm477889 | 1.9809 | 22_bit_twopoint_parametric_recessive |
| 5 | 133,225,776.00 | exm477889 | 2.1883 | 21_bit_twopoint_parametric_recessive |
| 5 | 133,720,353.00 | rs17643897 | 2.3325 | 22_bit_twopoint_parametric_recessive |
| 5 | 134,996,626.00 | rs13182891 | 2.8608 | 22_bit_twopoint_parametric_recessive |
| 5 | 134,996,626.00 | rs13182891 | 3.1466 | 21_bit_twopoint_parametric_recessive |
| 5 | 141,793,380.00 | rs496327 | 2.0373 | 22_bit_twopoint_parametric_dominant |
| 5 | 141,831,188.00 | rs166040 | 2.0084 | 21_bit_twopoint_parametric_recessive |
| 5 | 141,831,188.00 | rs166040 | 2.5229 | 22_bit_twopoint_parametric_recessive |
| 5 | 143,606,706.00 | rs246537 | 2.7999 | 21_bit_twopoint_parametric_recessive |
| 5 | 145,840,192.00 | rs11745791 | 1.9739 | 22_bit_twopoint_parametric_recessive |
| 5 | 146,956,824.00 | rs6580446 | 1.9214 | 22_bit_twopoint_parametric_recessive |
| 5 | 146,956,824.00 | rs6580446 | 2.447 | 21_bit_twopoint_parametric_recessive |
| 5 | 146,958,131.00 | JHU_5.146337693 | 1.947 | 22_bit_twopoint_parametric_recessive |
| 5 | 146,958,131.00 | JHU_5.146337693 | 2.5805 | 21_bit_twopoint_parametric_recessive |
| 5 | 151,687,037.00 | rs4958281 | 2.3105 | 21_bit_twopoint_parametric_dominant |
| 5 | 154,055,937.00 | rs11958069 | 2.2559 | 22_bit_twopoint_parametric_dominant |
| 5 | 155,078,345.00 | rs1319694 | 2.8336 | 22_bit_twopoint_parametric_recessive |
| 5 | 155,079,894.00 | rs9324802 | 2.7128 | 21_bit_twopoint_parametric_recessive |
| 5 | 155,079,894.00 | rs9324802 | 2.9788 | 22_bit_twopoint_parametric_recessive |
| 5 | 155,952,989.00 | rs10062650 | 2.5534 | 21_bit_twopoint_parametric_recessive |
| 5 | 156,025,720.00 | rs17536707 | 2.0911 | 21_bit_twopoint_parametric_dominant |
| 5 | 157,138,361.00 | rs3797851 | 2.3326 | 22_bit_twopoint_parametric_dominant |
| 5 | 157,158,143.00 | GSA-rs17054294 | 3.263 | 22_bit_twopoint_parametric_dominant |
| 5 | 158,104,961.00 | rs4704916 | 2.0437 | 21_bit_twopoint_parametric_recessive |
| 5 | 167,636,509.00 | rs10516034 | 2.5645 | 21_bit_twopoint_parametric_recessive |
| 5 | 167,645,438.00 | rs279397 | 2.846 | 22_bit_twopoint_parametric_recessive |
| 5 | 168,199,691.00 | GSA-rs3797719 | 2.2014 | 22_bit_twopoint_parametric_recessive |
| 5 | 168,199,691.00 | GSA-rs3797719 | 2.7354 | 21_bit_twopoint_parametric_recessive |
| 5 | 168,530,549.00 | GSA-rs3733977 | 2.7817 | 21_bit_twopoint_parametric_dominant |
| 5 | 168,557,716.00 | rs13185224 | 2.3173 | 21_bit_twopoint_parametric_dominant |
| 5 | 170,981,958.00 | rs10037007 | 2.1271 | 22_bit_twopoint_parametric_dominant |
| 5 | 171,095,902.00 | rs2446046 | 2.1271 | 22_bit_twopoint_parametric_dominant |
| 5 | 175,009,973.00 | rs17799508 | 2.0426 | 21_bit_twopoint_parametric_dominant |
| 5 | 175,042,206.00 | rs72809097 | 1.9515 | 22_bit_twopoint_parametric_dominant |
| 5 | 176,704,464.00 | JHU_5.176131464 | 2.066 | 21_bit_twopoint_parametric_recessive |
| 5 | 178,835,798.00 | rs10479432 | 2.0046 | 22_bit_twopoint_parametric_dominant |
| 5 | 179,207,546.00 | JHU_5.178634546 | 1.8897 | 21_bit_twopoint_parametric_dominant |
| 5 | 179,222,282.00 | rs13173873 | 1.8641 | 21_bit_twopoint_parametric_recessive |
| 5 | 179,222,282.00 | rs13173873 | 2.0702 | 22_bit_twopoint_parametric_dominant |
| 5 | 180,192,222.00 | rs7737438 | 2.004 | 21_bit_multipoint_NPL |
| 5 | 180,796,261.00 | GSA-rs10464106 | 2.0967 | 22_bit_twopoint_parametric_dominant |
| 5 | 180,813,754.00 | GSA-rs115052250 | 1.8656 | 21_bit_twopoint_parametric_dominant |
| 5 | 180,887,337.00 | JHU_5.180314336 | 2.3098 | 21_bit_twopoint_parametric_recessive |
| 5 | 180,887,337.00 | JHU_5.180314336 | 2.6778 | 21_bit_twopoint_parametric_dominant |
| 6 | 792,771.00 | JHU_6.792770 | 2.7937 | 22_bit_twopoint_parametric_recessive |
| 6 | 792,771.00 | JHU_6.792770 | 3.1939 | 21_bit_twopoint_parametric_recessive |
| 6 | 2,248,591.00 | rs335963 | 1.967 | 21_bit_twopoint_parametric_recessive |
| 6 | 2,447,465.00 | rs742487 | 1.8732 | 21_bit_twopoint_parametric_recessive |
| 6 | 2,447,465.00 | rs742487 | 2.142 | 22_bit_twopoint_parametric_recessive |
| 6 | 5,952,708.00 | rs80081488 | 1.9143 | 21_bit_twopoint_parametric_dominant |
| 6 | 6,051,822.00 | rs9405897 | 1.905 | 21_bit_twopoint_parametric_dominant |
| 6 | 6,289,624.00 | rs1267914 | 2.2324 | 22_bit_twopoint_parametric_dominant |
| 6 | 9,341,769.00 | JHU_6.9342001 | 1.9292 | 21_bit_twopoint_parametric_recessive |
| 6 | 9,956,329.00 | rs13217591 | 1.8695 | 22_bit_twopoint_parametric_recessive |
| 6 | 9,956,329.00 | rs13217591 | 2.1345 | 21_bit_twopoint_parametric_recessive |
| 6 | 10,337,018.00 | rs6908920 | 2.0563 | 21_bit_twopoint_parametric_dominant |
| 6 | 10,738,580.00 | rs529124 | 2.0586 | 22_bit_twopoint_parametric_dominant |
| 6 | 10,744,833.00 | rs545019 | 2.3977 | 22_bit_twopoint_parametric_dominant |
| 6 | 10,886,352.00 | rs12211124 | 2.0014 | 22_bit_twopoint_parametric_dominant |
| 6 | 10,898,587.00 | rs12190074 | 2.4543 | 22_bit_twopoint_parametric_dominant |
| 6 | 10,905,921.00 | rs9357021 | 1.9773 | 22_bit_twopoint_parametric_dominant |
| 6 | 11,357,745.00 | GSA-rs7775768 | 2.0601 | 21_bit_twopoint_parametric_dominant |
| 6 | 11,444,709.00 | rs77728288 | 2.3125 | 21_bit_twopoint_parametric_recessive |
| 6 | 11,444,709.00 | rs77728288 | 2.4131 | 22_bit_twopoint_parametric_recessive |
| 6 | 12,296,022.00 | exm517095 | 2.1771 | 22_bit_twopoint_parametric_dominant |
| 6 | 12,429,630.00 | rs13203590 | 2.61 | 22_bit_twopoint_parametric_dominant |
| 6 | 12,903,203.00 | rs4714955 | 2.1786 | 22_bit_twopoint_parametric_recessive |
| 6 | 12,911,733.00 | 6:12911965-A-G | 2.1786 | 22_bit_twopoint_parametric_recessive |
| 6 | 12,922,502.00 | 6:12922734-C-T | 2.3485 | 22_bit_twopoint_parametric_recessive |
| 6 | 12,923,535.00 | 6:12923767-C-A | 2.3129 | 22_bit_twopoint_parametric_recessive |
| 6 | 14,323,819.00 | 6:14324050 | 2.4763 | 22_bit_twopoint_parametric_recessive |
| 6 | 14,647,533.00 | rs7747013 | 2.5273 | 22_bit_multipoint_parametric_recessive |
| 6 | 14,647,533.00 | rs7747013 | 2.535 | 22_bit_twopoint_parametric_recessive |
| 6 | 16,144,383.00 | rs2056937 | 2.4815 | 22_bit_twopoint_parametric_dominant |
| 6 | 16,508,238.00 | rs4716077 | 2.5025 | 22_bit_twopoint_parametric_dominant |
| 6 | 20,210,143.00 | rs1327665 | 2.1329 | 21_bit_twopoint_parametric_dominant |
| 6 | 21,903,302.00 | rs7760611 | 2.1721 | 22_bit_twopoint_parametric_recessive |
| 6 | 21,903,302.00 | rs7760611 | 2.1721 | 22_bit_twopoint_parametric_recessive |
| 6 | 21,903,302.00 | rs7760611 | 2.2759 | 21_bit_twopoint_parametric_recessive |
| 6 | 21,903,302.00 | rs7760611 | 2.2759 | 21_bit_twopoint_parametric_recessive |
| 6 | 21,909,772.00 | JHU_6.21910002 | 1.8726 | 22_bit_twopoint_parametric_recessive |
| 6 | 21,909,772.00 | JHU_6.21910002 | 1.8726 | 22_bit_twopoint_parametric_recessive |
| 6 | 21,909,772.00 | JHU_6.21910002 | 2.1048 | 21_bit_twopoint_parametric_recessive |
| 6 | 21,909,772.00 | JHU_6.21910002 | 2.1048 | 21_bit_twopoint_parametric_recessive |
| 6 | 21,986,618.00 | rs6909060 | 2.2022 | 22_bit_twopoint_parametric_recessive |
| 6 | 21,986,618.00 | rs6909060 | 2.2022 | 22_bit_twopoint_parametric_recessive |
| 6 | 25,515,898.00 | rs301379 | 1.8676 | 21_bit_twopoint_parametric_recessive |
| 6 | 25,515,898.00 | rs301379 | 2.1669 | 21_bit_twopoint_parametric_dominant |
| 6 | 25,686,177.00 | GSA-rs9366627 | 2.3193 | 22_bit_twopoint_parametric_dominant |
| 6 | 26,276,422.00 | rs9393692 | 1.94 | 22_bit_twopoint_parametric_dominant |
| 6 | 26,295,458.00 | rs2893820 | 2.5449 | 22_bit_twopoint_parametric_dominant |
| 6 | 29,704,388.00 | JHU_6.29672164 | 3.1172 | 22_bit_twopoint_parametric_dominant |
| 6 | 32,388,031.00 | JHU_6.32355807 | 2.1162 | 22_bit_twopoint_parametric_dominant |
| 6 | 32,460,338.00 | rs9268835 | 1.9481 | 22_bit_twopoint_parametric_dominant |
| 6 | 32,460,938.00 | rs9268838 | 1.9477 | 22_bit_twopoint_parametric_dominant |
| 6 | 33,821,425.00 | GSA-rs73747323 | 2.0156 | 21_bit_twopoint_parametric_recessive |
| 6 | 33,821,425.00 | GSA-rs73747323 | 2.0156 | 21_bit_twopoint_parametric_recessive |
| 6 | 33,821,425.00 | GSA-rs73747323 | 2.2385 | 22_bit_twopoint_parametric_recessive |
| 6 | 33,821,425.00 | GSA-rs73747323 | 2.2385 | 22_bit_twopoint_parametric_recessive |
| 6 | 34,504,230.00 | rs4713805 | 1.8904 | 21_bit_twopoint_parametric_dominant |
| 6 | 35,408,229.00 | rs9462081 | 1.9337 | 22_bit_twopoint_parametric_recessive |
| 6 | 35,438,376.00 | rs4713859 | 2.1253 | 21_bit_twopoint_parametric_recessive |
| 6 | 35,438,376.00 | rs4713859 | 2.2948 | 22_bit_twopoint_parametric_recessive |
| 6 | 36,777,803.00 | JHU_6.36745579 | 2.1698 | 21_bit_twopoint_parametric_dominant |
| 6 | 39,315,030.00 | exm544617 | 1.9089 | 21_bit_twopoint_parametric_dominant |
| 6 | 41,500,122.00 | exm2270315 | 2.2913 | 21_bit_twopoint_parametric_dominant |
| 6 | 41,500,122.00 | exm2270315 | 2.2913 | 21_bit_twopoint_parametric_dominant |
| 6 | 41,500,942.00 | rs2495228 | 2.2369 | 21_bit_twopoint_parametric_dominant |
| 6 | 41,500,942.00 | rs2495228 | 2.2369 | 21_bit_twopoint_parametric_dominant |
| 6 | 43,931,040.00 | rs833623 | 2.274 | 21_bit_twopoint_parametric_dominant |
| 6 | 43,931,040.00 | rs833623 | 2.3052 | 21_bit_twopoint_parametric_recessive |
| 6 | 44,183,753.00 | exm551126 | 1.9389 | 21_bit_twopoint_parametric_dominant |
| 6 | 44,344,390.00 | JHU_6.44312126 | 1.947 | 21_bit_twopoint_parametric_dominant |
| 6 | 44,344,390.00 | JHU_6.44312126 | 1.9581 | 21_bit_twopoint_parametric_recessive |
| 6 | 44,978,726.00 | GSA-rs2396369 | 2.3981 | 22_bit_twopoint_parametric_dominant |
| 6 | 45,552,982.00 | rs9472501 | 2.232 | 22_bit_twopoint_parametric_dominant |
| 6 | 45,602,126.00 | rs9472521 | 1.9844 | 22_bit_twopoint_parametric_dominant |
| 6 | 46,024,596.00 | rs13328271 | 2.0783 | 21_bit_twopoint_parametric_dominant |
| 6 | 46,789,539.00 | JHU_6.46757275 | 2.2649 | 21_bit_twopoint_parametric_dominant |
| 6 | 47,356,881.00 | rs9473079 | 2.5295 | 21_bit_twopoint_parametric_dominant |
| 6 | 47,686,162.00 | rs9296571 | 2.2028 | 21_bit_twopoint_parametric_dominant |
| 6 | 47,757,304.00 | rs9473171 | 2.1706 | 21_bit_twopoint_parametric_dominant |
| 6 | 47,787,551.00 | GSA-rs9473176 | 1.9108 | 22_bit_twopoint_parametric_recessive |
| 6 | 47,899,741.00 | rs9968847 | 1.8709 | 21_bit_twopoint_parametric_recessive |
| 6 | 47,899,741.00 | rs9968847 | 1.9426 | 21_bit_twopoint_parametric_dominant |
| 6 | 48,052,095.00 | GSA-rs167976 | 2.2373 | 22_bit_twopoint_parametric_dominant |
| 6 | 50,747,694.00 | GSA-rs9349553 | 2.0467 | 21_bit_twopoint_parametric_dominant |
| 6 | 53,446,041.00 | exm-rs9367532 | 1.8932 | 21_bit_twopoint_parametric_recessive |
| 6 | 55,202,829.00 | rs6937878 | 2.0271 | 21_bit_twopoint_parametric_dominant |
| 6 | 55,242,469.00 | GSA-rs9382471 | 1.9268 | 21_bit_twopoint_parametric_dominant |
| 6 | 55,472,264.00 | rs9475303 | 1.8636 | 22_bit_twopoint_parametric_dominant |
| 6 | 65,024,465.00 | rs7738129 | 1.9143 | 21_bit_twopoint_parametric_dominant |
| 6 | 65,027,022.00 | GSA-rs13216649 | 1.9203 | 21_bit_twopoint_parametric_dominant |
| 6 | 65,415,882.00 | rs9363358 | 1.9541 | 21_bit_twopoint_parametric_recessive |
| 6 | 65,415,882.00 | rs9363358 | 2.3583 | 22_bit_twopoint_parametric_recessive |
| 6 | 68,374,440.00 | GSA-rs2341485 | 2.0289 | 21_bit_twopoint_parametric_dominant |
| 6 | 68,374,440.00 | GSA-rs2341485 | 2.531 | 21_bit_twopoint_parametric_recessive |
| 6 | 73,136,526.00 | JHU_6.73846248 | 2.2161 | 21_bit_twopoint_parametric_dominant |
| 6 | 76,224,151.00 | JHU_6.76933867 | 1.8935 | 21_bit_twopoint_parametric_dominant |
| 6 | 76,290,692.00 | GSA-rs6918782 | 1.8935 | 21_bit_twopoint_parametric_dominant |
| 6 | 76,404,952.00 | JHU_6.77114668 | 1.9167 | 21_bit_twopoint_parametric_dominant |
| 6 | 77,098,077.00 | rs10943414 | 2.5554 | 21_bit_twopoint_parametric_dominant |
| 6 | 78,243,529.00 | rs6911371 | 2.3914 | 22_bit_twopoint_parametric_dominant |
| 6 | 79,745,114.00 | rs2092745 | 1.888 | 22_bit_twopoint_parametric_recessive |
| 6 | 79,745,114.00 | rs2092745 | 2.5609 | 21_bit_twopoint_parametric_recessive |
| 6 | 82,059,713.00 | rs1361682 | 1.8725 | 22_bit_twopoint_parametric_dominant |
| 6 | 82,059,713.00 | rs1361682 | 1.8725 | 22_bit_twopoint_parametric_dominant |
| 6 | 82,084,877.00 | rs1538138 | 2.5026 | 22_bit_twopoint_parametric_dominant |
| 6 | 82,084,877.00 | rs1538138 | 2.5026 | 22_bit_twopoint_parametric_dominant |
| 6 | 82,084,877.00 | rs1538138 | 3.1873 | 21_bit_twopoint_parametric_dominant |
| 6 | 82,084,877.00 | rs1538138 | 3.1873 | 21_bit_twopoint_parametric_dominant |
| 6 | 82,090,155.00 | rs7775215 | 2.2495 | 22_bit_twopoint_parametric_dominant |
| 6 | 82,090,155.00 | rs7775215 | 2.2495 | 22_bit_twopoint_parametric_dominant |
| 6 | 82,110,675.00 | rs1933023 | 2.3917 | 21_bit_twopoint_parametric_dominant |
| 6 | 82,110,675.00 | rs1933023 | 2.3917 | 21_bit_twopoint_parametric_dominant |
| 6 | 82,113,053.00 | rs7741402 | 2.4924 | 22_bit_twopoint_parametric_dominant |
| 6 | 82,113,053.00 | rs7741402 | 2.4924 | 22_bit_twopoint_parametric_dominant |
| 6 | 82,113,053.00 | rs7741402 | 2.9273 | 21_bit_twopoint_parametric_dominant |
| 6 | 82,113,053.00 | rs7741402 | 2.9273 | 21_bit_twopoint_parametric_dominant |
| 6 | 82,142,827.00 | GSA-rs9341911 | 3.0402 | 22_bit_twopoint_parametric_dominant |
| 6 | 82,142,827.00 | GSA-rs9341911 | 3.0402 | 22_bit_twopoint_parametric_dominant |
| 6 | 82,142,827.00 | GSA-rs9341911 | 3.1395 | 21_bit_twopoint_parametric_dominant |
| 6 | 82,142,827.00 | GSA-rs9341911 | 3.1395 | 21_bit_twopoint_parametric_dominant |
| 6 | 82,901,120.00 | exm2262180 | 1.9508 | 22_bit_twopoint_parametric_dominant |
| 6 | 82,901,120.00 | exm2262180 | 1.9508 | 22_bit_twopoint_parametric_dominant |
| 6 | 82,907,632.00 | rs58578814 | 1.9334 | 22_bit_twopoint_parametric_dominant |
| 6 | 82,907,632.00 | rs58578814 | 1.9334 | 22_bit_twopoint_parametric_dominant |
| 6 | 83,718,561.00 | rs586661 | 2.2843 | 22_bit_twopoint_parametric_recessive |
| 6 | 83,718,561.00 | rs586661 | 2.4841 | 21_bit_twopoint_parametric_recessive |
| 6 | 84,272,490.00 | rs12174666 | 2.1394 | 21_bit_twopoint_parametric_dominant |
| 6 | 90,035,123.00 | GSA-rs9342217 | 1.9514 | 22_bit_twopoint_parametric_dominant |
| 6 | 90,046,734.00 | rs9342219 | 1.9577 | 22_bit_twopoint_parametric_dominant |
| 6 | 98,861,388.00 | rs76358326 | 2.3418 | 21_bit_twopoint_parametric_dominant |
| 6 | 102,044,379.00 | rs1413966 | 2.0728 | 21_bit_twopoint_parametric_recessive |
| 6 | 102,044,379.00 | rs1413966 | 3.0761 | 22_bit_twopoint_parametric_recessive |
| 6 | 103,187,245.00 | rs12213111 | 1.9657 | 21_bit_twopoint_parametric_recessive |
| 6 | 103,187,245.00 | rs12213111 | 2.1479 | 21_bit_twopoint_parametric_dominant |
| 6 | 105,587,909.00 | rs9399925 | 2.2407 | 21_bit_twopoint_parametric_dominant |
| 6 | 105,587,909.00 | rs9399925 | 2.4736 | 22_bit_twopoint_parametric_dominant |
| 6 | 105,592,738.00 | GSA-rs2013092 | 2.142 | 22_bit_twopoint_parametric_dominant |
| 6 | 105,592,738.00 | GSA-rs2013092 | 2.647 | 21_bit_twopoint_parametric_dominant |
| 6 | 105,600,810.00 | rs34621793 | 2.1245 | 21_bit_twopoint_parametric_dominant |
| 6 | 110,438,805.00 | exm571670 | 1.9975 | 21_bit_twopoint_parametric_dominant |
| 6 | 110,492,725.00 | rs2428168 | 1.8626 | 22_bit_twopoint_parametric_dominant |
| 6 | 111,149,813.00 | rs62421925 | 2.2795 | 22_bit_twopoint_parametric_recessive |
| 6 | 113,221,496.00 | rs706922 | 2.5154 | 22_bit_twopoint_parametric_dominant |
| 6 | 113,237,600.00 | JHU_6.113558801 | 2.3383 | 22_bit_twopoint_parametric_dominant |
| 6 | 113,284,630.00 | rs9488132 | 2.0128 | 22_bit_twopoint_parametric_dominant |
| 6 | 114,914,441.00 | GSA-rs9372420 | 2.1174 | 22_bit_twopoint_parametric_recessive |
| 6 | 115,527,965.00 | JHU_6.115849128 | 2.3208 | 22_bit_twopoint_parametric_recessive |
| 6 | 122,106,037.00 | GSA-rs2816076 | 2.8229 | 22_bit_multipoint_parametric_dominant |
| 6 | 122,106,037.00 | GSA-rs2816076 | 2.8547 | 22_bit_twopoint_parametric_dominant |
| 6 | 122,801,319.00 | exm575754 | 1.9678 | 21_bit_twopoint_parametric_dominant |
| 6 | 122,806,452.00 | rs13192569 | 1.9334 | 21_bit_twopoint_parametric_dominant |
| 6 | 123,857,135.00 | rs6569368 | 1.8896 | 21_bit_twopoint_parametric_dominant |
| 6 | 127,450,471.00 | GSA-rs41285270 | 2.8657 | 22_bit_twopoint_parametric_recessive |
| 6 | 127,985,973.00 | rs4341027 | 2.8787 | 21_bit_twopoint_parametric_recessive |
| 6 | 132,630,708.00 | JHU_6.132951846 | 2.1147 | 22_bit_twopoint_parametric_dominant |
| 6 | 132,630,708.00 | JHU_6.132951846 | 2.5516 | 22_bit_twopoint_parametric_recessive |
| 6 | 132,752,286.00 | rs35229014 | 1.9356 | 21_bit_twopoint_parametric_recessive |
| 6 | 134,143,635.00 | rs6941979 | 1.8943 | 22_bit_twopoint_parametric_recessive |
| 6 | 134,143,635.00 | rs6941979 | 2.2446 | 21_bit_twopoint_parametric_recessive |
| 6 | 139,311,722.00 | rs6570330 | 2.4177 | 21_bit_twopoint_parametric_dominant |
| 6 | 148,581,015.00 | rs208699 | 1.9538 | 22_bit_twopoint_parametric_dominant |
| 6 | 152,316,543.00 | rs2013767 | 2.3943 | 22_bit_twopoint_parametric_dominant |
| 6 | 155,421,711.00 | rs2235667 | 2.1756 | 21_bit_twopoint_parametric_dominant |
| 6 | 156,048,595.00 | rs78686837 | 2.0285 | 22_bit_twopoint_parametric_recessive |
| 6 | 156,048,595.00 | rs78686837 | 2.0394 | 22_bit_twopoint_parametric_dominant |
| 6 | 156,175,326.00 | rs7742923 | 2.6398 | 21_bit_twopoint_parametric_recessive |
| 6 | 157,656,955.00 | rs117565536 | 1.9093 | 21_bit_twopoint_parametric_recessive |
| 6 | 157,656,955.00 | rs117565536 | 1.9093 | 21_bit_twopoint_parametric_recessive |
| 6 | 157,656,955.00 | rs117565536 | 2.145 | 22_bit_twopoint_parametric_recessive |
| 6 | 157,656,955.00 | rs117565536 | 2.145 | 22_bit_twopoint_parametric_recessive |
| 6 | 160,430,734.00 | rs376563 | 2.2905 | 22_bit_twopoint_parametric_dominant |
| 6 | 160,548,706.00 | rs10755578 | 1.9344 | 22_bit_twopoint_parametric_dominant |
| 6 | 161,896,237.00 | rs9458413 | 2.3039 | 21_bit_twopoint_parametric_recessive |
| 6 | 162,232,577.00 | rs10945805 | 2.5821 | 21_bit_twopoint_parametric_dominant |
| 6 | 163,925,930.00 | rs73024969 | 2.3298 | 21_bit_twopoint_parametric_dominant |
| 6 | 163,929,055.00 | rs2322177 | 2.3248 | 21_bit_twopoint_parametric_dominant |
| 6 | 164,849,469.00 | rs73030892 | 2.1555 | 21_bit_twopoint_parametric_dominant |
| 6 | 164,849,469.00 | rs73030892 | 2.1555 | 21_bit_twopoint_parametric_dominant |
| 6 | 165,146,847.00 | GSA-rs6926303 | 2.3349 | 21_bit_twopoint_parametric_dominant |
| 6 | 165,146,847.00 | GSA-rs6926303 | 2.3349 | 21_bit_twopoint_parametric_dominant |
| 6 | 166,153,857.00 | rs6914547 | 1.8858 | 22_bit_twopoint_parametric_recessive |
| 6 | 166,297,799.00 | rs999234 | 2.3085 | 21_bit_twopoint_parametric_dominant |
| 6 | 170,038,190.00 | GSA-rs116988566 | 1.8744 | 22_bit_twopoint_parametric_recessive |
| 7 | 534,590.00 | rs9801615 | 1.9516 | 22_bit_twopoint_parametric_dominant |
| 7 | 1,529,782.00 | rs3814477 | 1.9473 | 22_bit_twopoint_parametric_dominant |
| 7 | 1,534,767.00 | rs4720833 | 2.4058 | 22_bit_twopoint_parametric_dominant |
| 7 | 1,543,652.00 | rs892523 | 2.2508 | 22_bit_twopoint_parametric_dominant |
| 7 | 3,205,266.00 | rs1404874 | 2.1404 | 22_bit_twopoint_parametric_recessive |
| 7 | 4,860,464.00 | exm-rs1553960 | 2.4407 | 22_bit_twopoint_parametric_dominant |
| 7 | 7,325,375.00 | rs4724972 | 1.9099 | 21_bit_twopoint_parametric_recessive |
| 7 | 7,325,375.00 | rs4724972 | 1.9099 | 21_bit_twopoint_parametric_recessive |
| 7 | 7,325,375.00 | rs4724972 | 2.4079 | 22_bit_twopoint_parametric_recessive |
| 7 | 7,325,375.00 | rs4724972 | 2.4079 | 22_bit_twopoint_parametric_recessive |
| 7 | 7,359,543.00 | rs4724976 | 2.0439 | 22_bit_twopoint_parametric_recessive |
| 7 | 7,359,543.00 | rs4724976 | 2.0439 | 22_bit_twopoint_parametric_recessive |
| 7 | 7,359,773.00 | GSA-rs10486158 | 2.0439 | 22_bit_twopoint_parametric_recessive |
| 7 | 7,359,773.00 | GSA-rs10486158 | 2.0439 | 22_bit_twopoint_parametric_recessive |
| 7 | 7,362,258.00 | JHU_7.7401888 | 2.0175 | 22_bit_twopoint_parametric_recessive |
| 7 | 7,362,258.00 | JHU_7.7401888 | 2.0175 | 22_bit_twopoint_parametric_recessive |
| 7 | 7,702,425.00 | rs2881961 | 1.9999 | 21_bit_twopoint_parametric_recessive |
| 7 | 7,702,425.00 | rs2881961 | 1.9999 | 21_bit_twopoint_parametric_recessive |
| 7 | 7,702,425.00 | rs2881961 | 2.1112 | 21_bit_twopoint_parametric_dominant |
| 7 | 7,702,425.00 | rs2881961 | 2.1112 | 21_bit_twopoint_parametric_dominant |
| 7 | 7,840,966.00 | rs758994 | 2.0396 | 22_bit_twopoint_parametric_recessive |
| 7 | 7,840,966.00 | rs758994 | 2.0396 | 22_bit_twopoint_parametric_recessive |
| 7 | 8,140,216.00 | rs887848 | 1.8897 | 22_bit_twopoint_parametric_dominant |
| 7 | 8,204,478.00 | rs12670131 | 2.2695 | 21_bit_twopoint_parametric_dominant |
| 7 | 8,324,586.00 | rs17147407 | 1.8889 | 22_bit_twopoint_parametric_recessive |
| 7 | 10,862,754.00 | rs6952827 | 2.046 | 22_bit_twopoint_parametric_dominant |
| 7 | 10,968,594.00 | rs1640705 | 2.0572 | 21_bit_twopoint_parametric_dominant |
| 7 | 11,141,729.00 | GSA-rs73065407 | 2.2161 | 21_bit_twopoint_parametric_dominant |
| 7 | 11,504,983.00 | GSA-rs6943841 | 1.9376 | 21_bit_twopoint_parametric_dominant |
| 7 | 13,653,435.00 | rs17167319 | 2.1259 | 21_bit_twopoint_parametric_dominant |
| 7 | 16,087,672.00 | rs12539174 | 1.9467 | 21_bit_twopoint_parametric_recessive |
| 7 | 18,585,177.00 | rs2073974 | 2.2078 | 21_bit_twopoint_parametric_dominant |
| 7 | 21,775,270.00 | rs74418216 | 1.962 | 21_bit_twopoint_parametric_recessive |
| 7 | 21,911,487.00 | rs11978167 | 2.0513 | 21_bit_twopoint_parametric_dominant |
| 7 | 21,948,266.00 | rs1534771 | 1.8856 | 22_bit_twopoint_parametric_recessive |
| 7 | 22,564,700.00 | rs10275628 | 2.0778 | 21_bit_twopoint_parametric_dominant |
| 7 | 22,623,834.00 | GSA-rs2961287 | 2.0062 | 21_bit_twopoint_parametric_dominant |
| 7 | 22,856,292.00 | rs62449832 | 2.1885 | 22_bit_twopoint_parametric_dominant |
| 7 | 24,976,244.00 | rs6461832 | 1.8814 | 22_bit_twopoint_parametric_recessive |
| 7 | 24,976,244.00 | rs6461832 | 2.0068 | 21_bit_twopoint_parametric_recessive |
| 7 | 26,363,328.00 | rs2698719 | 2.118 | 21_bit_twopoint_parametric_dominant |
| 7 | 27,192,143.00 | rs4722672 | 2.7027 | 22_bit_twopoint_parametric_dominant |
| 7 | 28,649,970.00 | rs41327 | 1.9094 | 21_bit_twopoint_parametric_dominant |
| 7 | 29,193,383.00 | rs245923 | 2.2327 | 22_bit_twopoint_parametric_recessive |
| 7 | 29,193,383.00 | rs245923 | 2.4949 | 21_bit_twopoint_parametric_recessive |
| 7 | 29,416,389.00 | rs2057739 | 1.9648 | 22_bit_twopoint_parametric_recessive |
| 7 | 29,827,466.00 | rs174929 | 2.8716 | 22_bit_twopoint_parametric_dominant |
| 7 | 31,048,278.00 | rs35863573 | 2.1713 | 22_bit_twopoint_parametric_recessive |
| 7 | 31,310,930.00 | rs7799218 | 1.8807 | 22_bit_twopoint_parametric_recessive |
| 7 | 31,465,355.00 | rs218092 | 1.9155 | 22_bit_twopoint_parametric_dominant |
| 7 | 34,134,561.00 | rs6961966 | 2.3735 | 22_bit_twopoint_parametric_recessive |
| 7 | 34,134,561.00 | rs6961966 | 2.5545 | 22_bit_twopoint_parametric_dominant |
| 7 | 36,516,987.00 | rs17170619 | 2.1417 | 22_bit_twopoint_parametric_recessive |
| 7 | 36,516,987.00 | rs17170619 | 2.9744 | 22_bit_twopoint_parametric_dominant |
| 7 | 37,872,933.00 | rs2722309 | 1.8794 | 21_bit_twopoint_parametric_dominant |
| 7 | 39,486,744.00 | rs17713347 | 1.963 | 22_bit_twopoint_parametric_recessive |
| 7 | 43,004,132.00 | rs75899742 | 2.3096 | 22_bit_twopoint_parametric_recessive |
| 7 | 43,412,111.00 | rs4724203 | 2.0979 | 21_bit_twopoint_parametric_dominant |
| 7 | 44,189,469.00 | exm-rs1799884 | 2.9327 | 22_bit_twopoint_parametric_recessive |
| 7 | 44,189,469.00 | exm-rs1799884 | 2.9618 | 21_bit_twopoint_parametric_recessive |
| 7 | 44,192,179.00 | GSA-rs2971669 | 2.002 | 21_bit_twopoint_parametric_recessive |
| 7 | 44,195,138.00 | 7:44234737-C-T | 2.9592 | 22_bit_twopoint_parametric_recessive |
| 7 | 44,195,138.00 | 7:44234737-C-T | 3.0177 | 21_bit_twopoint_parametric_recessive |
| 7 | 44,196,069.00 | exm-rs4607517 | 2.4464 | 22_bit_twopoint_parametric_recessive |
| 7 | 44,196,069.00 | exm-rs4607517 | 2.4521 | 21_bit_twopoint_parametric_recessive |
| 7 | 44,200,808.00 | GSA-rs1004558 | 2.9327 | 22_bit_twopoint_parametric_recessive |
| 7 | 44,200,808.00 | GSA-rs1004558 | 2.9618 | 21_bit_twopoint_parametric_recessive |
| 7 | 44,205,461.00 | GSA-rs2971667 | 2.9488 | 22_bit_twopoint_parametric_recessive |
| 7 | 44,205,461.00 | GSA-rs2971667 | 3.2107 | 21_bit_twopoint_parametric_recessive |
| 7 | 44,222,379.00 | 7:44261978-G-A | 2.4893 | 22_bit_twopoint_parametric_recessive |
| 7 | 44,222,379.00 | 7:44261978-G-A | 2.8854 | 21_bit_twopoint_parametric_recessive |
| 7 | 44,920,337.00 | rs4724334 | 1.9772 | 21_bit_twopoint_parametric_dominant |
| 7 | 47,941,638.00 | rs2686828 | 1.8661 | 22_bit_twopoint_parametric_dominant |
| 7 | 48,459,713.00 | rs4083233 | 2.0019 | 22_bit_twopoint_parametric_dominant |
| 7 | 49,697,573.00 | rs2162178 | 1.8869 | 22_bit_twopoint_parametric_recessive |
| 7 | 50,286,121.00 | rs716719 | 1.9084 | 22_bit_twopoint_parametric_dominant |
| 7 | 50,295,636.00 | rs4917017 | 2.5233 | 22_bit_twopoint_parametric_dominant |
| 7 | 50,306,538.00 | rs7781977 | 2.5258 | 22_bit_twopoint_parametric_dominant |
| 7 | 68,070,177.00 | rs11509864 | 2.1489 | 22_bit_twopoint_parametric_recessive |
| 7 | 68,070,177.00 | rs11509864 | 2.6451 | 21_bit_twopoint_parametric_recessive |
| 7 | 69,151,123.00 | rs1195227 | 2.1101 | 21_bit_twopoint_parametric_dominant |
| 7 | 85,742,070.00 | JHU_7.85371385 | 1.9182 | 22_bit_twopoint_parametric_recessive |
| 7 | 85,780,871.00 | rs73387717 | 2.4612 | 22_bit_twopoint_parametric_recessive |
| 7 | 85,902,146.00 | rs17339155 | 2.7651 | 22_bit_twopoint_parametric_dominant |
| 7 | 85,929,062.00 | rs717708 | 1.8839 | 22_bit_twopoint_parametric_dominant |
| 7 | 85,929,062.00 | rs717708 | 2.1223 | 21_bit_twopoint_parametric_recessive |
| 7 | 85,929,062.00 | rs717708 | 2.6543 | 22_bit_twopoint_parametric_recessive |
| 7 | 86,670,917.00 | rs12668286 | 1.9353 | 21_bit_twopoint_parametric_dominant |
| 7 | 87,391,707.00 | exm631536 | 1.9031 | 21_bit_twopoint_parametric_recessive |
| 7 | 87,398,843.00 | kgp12073594 | 1.9176 | 21_bit_twopoint_parametric_recessive |
| 7 | 87,408,758.00 | rs17149539 | 1.971 | 21_bit_twopoint_parametric_recessive |
| 7 | 87,417,154.00 | rs4148829 | 1.971 | 21_bit_twopoint_parametric_recessive |
| 7 | 88,100,063.00 | JHU_7.87729377 | 2.0924 | 22_bit_twopoint_parametric_dominant |
| 7 | 88,125,600.00 | GSA-rs17255978 | 2.0924 | 22_bit_twopoint_parametric_dominant |
| 7 | 88,138,859.00 | rs10268574 | 1.8903 | 22_bit_twopoint_parametric_dominant |
| 7 | 88,145,842.00 | rs10238607 | 2.0924 | 22_bit_twopoint_parametric_dominant |
| 7 | 88,983,797.00 | rs2214628 | 2.346 | 22_bit_twopoint_parametric_dominant |
| 7 | 90,469,589.00 | rs10268928 | 2.6204 | 21_bit_twopoint_parametric_recessive |
| 7 | 90,543,735.00 | GSA-rs17875046 | 1.8747 | 21_bit_twopoint_parametric_recessive |
| 7 | 90,543,735.00 | GSA-rs17875046 | 2.5871 | 22_bit_twopoint_parametric_recessive |
| 7 | 94,373,948.00 | rs6465411 | 2.2671 | 21_bit_twopoint_parametric_recessive |
| 7 | 94,373,948.00 | rs6465411 | 3.1164 | 21_bit_twopoint_parametric_dominant |
| 7 | 105,433,341.00 | rs955056 | 1.8818 | 22_bit_twopoint_parametric_dominant |
| 7 | 105,436,530.00 | GSA-rs6970417 | 2.1728 | 22_bit_twopoint_parametric_dominant |
| 7 | 111,832,984.00 | rs10238664 | 1.9353 | 21_bit_twopoint_parametric_dominant |
| 7 | 111,832,984.00 | rs10238664 | 2.2051 | 22_bit_twopoint_parametric_dominant |
| 7 | 111,846,970.00 | rs10275038 | 1.9817 | 21_bit_twopoint_parametric_dominant |
| 7 | 111,846,970.00 | rs10275038 | 2.286 | 22_bit_twopoint_parametric_dominant |
| 7 | 111,860,497.00 | 7:111500553-C-T | 2.1938 | 21_bit_twopoint_parametric_recessive |
| 7 | 111,860,766.00 | 7:111500822-G-A | 2.1938 | 21_bit_twopoint_parametric_recessive |
| 7 | 111,860,880.00 | 7:111500936-G-A | 2.1938 | 21_bit_twopoint_parametric_recessive |
| 7 | 111,863,604.00 | 7:111503660-G-A | 2.1938 | 21_bit_twopoint_parametric_recessive |
| 7 | 111,886,263.00 | 7:111526319-T-C | 2.1222 | 21_bit_twopoint_parametric_recessive |
| 7 | 111,887,504.00 | rs10255299 | 2.1246 | 21_bit_twopoint_parametric_recessive |
| 7 | 111,893,464.00 | 7:111533520-T-C | 2.2451 | 21_bit_twopoint_parametric_recessive |
| 7 | 111,897,412.00 | 7:111537468-T-C | 2.1246 | 21_bit_twopoint_parametric_recessive |
| 7 | 111,899,490.00 | 7:111539546-G-A | 2.1046 | 21_bit_twopoint_parametric_recessive |
| 7 | 111,900,776.00 | 7:111540832-A-G | 2.1246 | 21_bit_twopoint_parametric_recessive |
| 7 | 111,940,111.00 | GSA-rs144867634 | 1.968 | 21_bit_twopoint_parametric_recessive |
| 7 | 111,940,111.00 | GSA-rs144867634 | 2.0535 | 22_bit_twopoint_parametric_recessive |
| 7 | 112,442,867.00 | rs3109107 | 1.9269 | 21_bit_twopoint_parametric_dominant |
| 7 | 114,659,719.00 | rs72603568 | 2.7221 | 21_bit_twopoint_parametric_recessive |
| 7 | 115,086,036.00 | GSA-rs76277946 | 2.201 | 21_bit_twopoint_parametric_recessive |
| 7 | 115,132,748.00 | rs13229627 | 1.9161 | 21_bit_twopoint_parametric_recessive |
| 7 | 115,159,855.00 | GSA-rs2464884 | 1.9342 | 21_bit_twopoint_parametric_recessive |
| 7 | 120,765,281.00 | GSA-rs10239799 | 2.4151 | 21_bit_twopoint_parametric_dominant |
| 7 | 120,765,281.00 | GSA-rs10239799 | 2.4151 | 21_bit_twopoint_parametric_dominant |
| 7 | 121,268,768.00 | rs2536149 | 2.0629 | 22_bit_twopoint_parametric_recessive |
| 7 | 121,268,768.00 | rs2536149 | 2.0629 | 22_bit_twopoint_parametric_recessive |
| 7 | 121,268,768.00 | rs2536149 | 2.2036 | 22_bit_twopoint_parametric_dominant |
| 7 | 121,268,768.00 | rs2536149 | 2.2036 | 22_bit_twopoint_parametric_dominant |
| 7 | 121,974,272.00 | rs1916872 | 1.9721 | 22_bit_twopoint_parametric_recessive |
| 7 | 121,974,272.00 | rs1916872 | 1.9721 | 22_bit_twopoint_parametric_recessive |
| 7 | 122,028,266.00 | rs1147499 | 2.2554 | 22_bit_twopoint_parametric_recessive |
| 7 | 123,993,179.00 | rs12668777 | 2.2622 | 21_bit_twopoint_parametric_recessive |
| 7 | 124,020,482.00 | JHU_7.123660535 | 1.9462 | 22_bit_twopoint_parametric_recessive |
| 7 | 124,020,482.00 | JHU_7.123660535 | 2.634 | 21_bit_twopoint_parametric_recessive |
| 7 | 124,020,547.00 | rs75030752 | 1.9462 | 22_bit_twopoint_parametric_recessive |
| 7 | 124,020,547.00 | rs75030752 | 2.634 | 21_bit_twopoint_parametric_recessive |
| 7 | 124,068,042.00 | JHU_7.123708095 | 2.411 | 21_bit_twopoint_parametric_recessive |
| 7 | 124,088,981.00 | JHU_7.123729034 | 2.0714 | 21_bit_twopoint_parametric_recessive |
| 7 | 124,278,587.00 | rs13233306 | 2.1998 | 21_bit_twopoint_parametric_recessive |
| 7 | 124,760,651.00 | rs3807544 | 1.9325 | 22_bit_twopoint_parametric_dominant |
| 7 | 125,769,644.00 | rs7780223 | 2.2193 | 22_bit_twopoint_parametric_recessive |
| 7 | 127,217,186.00 | rs2299555 | 2.1442 | 22_bit_twopoint_parametric_dominant |
| 7 | 130,200,151.00 | JHU_7.129839990 | 2.2609 | 22_bit_twopoint_parametric_recessive |
| 7 | 130,937,536.00 | JHU_7.130622294 | 2.1452 | 22_bit_twopoint_parametric_recessive |
| 7 | 135,261,427.00 | rs292639 | 1.95 | 22_bit_twopoint_parametric_recessive |
| 7 | 135,943,622.00 | GSA-rs73162998 | 2.143 | 22_bit_twopoint_parametric_recessive |
| 7 | 135,997,671.00 | GSA-rs1994464 | 2.0324 | 22_bit_twopoint_parametric_dominant |
| 7 | 136,073,809.00 | rs6967877 | 2.1816 | 22_bit_twopoint_parametric_recessive |
| 7 | 136,077,324.00 | rs10229361 | 2.0905 | 22_bit_twopoint_parametric_recessive |
| 7 | 136,153,780.00 | rs1002292 | 2.2052 | 21_bit_twopoint_parametric_dominant |
| 7 | 136,183,082.00 | JHU_7.135867829 | 1.8848 | 22_bit_twopoint_parametric_recessive |
| 7 | 136,958,947.00 | rs1824024 | 1.9233 | 21_bit_twopoint_parametric_recessive |
| 7 | 136,958,947.00 | rs1824024 | 2.0072 | 22_bit_twopoint_parametric_recessive |
| 7 | 137,060,207.00 | rs11977730 | 1.9142 | 22_bit_twopoint_parametric_recessive |
| 7 | 137,979,155.00 | rs59191429 | 1.9378 | 22_bit_twopoint_parametric_recessive |
| 7 | 138,052,800.00 | rs6467729 | 2.1649 | 22_bit_twopoint_parametric_recessive |
| 7 | 139,267,954.00 | rs17160838 | 1.8889 | 22_bit_twopoint_parametric_recessive |
| 7 | 139,278,618.00 | rs7796255 | 2.2781 | 21_bit_twopoint_parametric_recessive |
| 7 | 139,278,618.00 | rs7796255 | 2.862 | 22_bit_twopoint_parametric_recessive |
| 7 | 139,996,449.00 | rs2267705 | 2.118 | 21_bit_twopoint_NPL |
| 7 | 139,996,449.00 | rs2267705 | 2.4133 | 21_bit_twopoint_parametric_dominant |
| 7 | 140,004,763.00 | rs11761051 | 1.871 | 21_bit_twopoint_parametric_dominant |
| 7 | 140,004,763.00 | rs11761051 | 2.744 | 21_bit_twopoint_parametric_recessive |
| 7 | 140,007,340.00 | rs740204 | 2.0179 | 22_bit_twopoint_parametric_dominant |
| 7 | 140,007,340.00 | rs740204 | 2.3554 | 21_bit_twopoint_parametric_dominant |
| 7 | 140,101,977.00 | rs1062277 | 2.0148 | 22_bit_twopoint_parametric_recessive |
| 7 | 140,324,252.00 | rs12703744 | 1.9873 | 22_bit_twopoint_parametric_recessive |
| 7 | 141,410,346.00 | JHU_7.141110145 | 2.4661 | 22_bit_twopoint_parametric_recessive |
| 7 | 141,410,346.00 | JHU_7.141110145 | 3.1771 | 22_bit_twopoint_parametric_dominant |
| 7 | 141,916,706.00 | GSA-rs745162 | 1.8627 | 22_bit_twopoint_parametric_recessive |
| 7 | 142,802,725.00 | rs11327 | 2.1856 | 21_bit_multipoint_parametric_recessive |
| 7 | 142,802,725.00 | rs11327 | 2.1856 | 21_bit_twopoint_parametric_recessive |
| 7 | 142,802,725.00 | rs11327 | 2.1856 | 21_bit_twopoint_parametric_recessive |
| 7 | 143,053,854.00 | GSA-rs1506403 | 2.0718 | 21_bit_twopoint_parametric_recessive |
| 7 | 143,053,854.00 | GSA-rs1506403 | 2.0718 | 21_bit_twopoint_parametric_recessive |
| 7 | 143,491,644.00 | rs1525107 | 1.9465 | 21_bit_twopoint_parametric_recessive |
| 7 | 143,491,644.00 | rs1525107 | 1.9465 | 21_bit_twopoint_parametric_recessive |
| 7 | 145,186,603.00 | rs7783785 | 1.9557 | 21_bit_twopoint_parametric_recessive |
| 7 | 145,557,932.00 | JHU_7.145255024 | 2.416 | 21_bit_twopoint_parametric_recessive |
| 7 | 146,078,801.00 | rs55822162 | 2.2134 | 22_bit_twopoint_parametric_recessive |
| 7 | 146,078,801.00 | rs55822162 | 2.9656 | 21_bit_twopoint_parametric_recessive |
| 7 | 146,120,170.00 | kgp8369373 | 2.2114 | 21_bit_twopoint_parametric_recessive |
| 7 | 146,742,777.00 | rs7790238 | 2.4066 | 22_bit_twopoint_parametric_recessive |
| 7 | 147,103,014.00 | rs6962089 | 1.927 | 21_bit_twopoint_parametric_recessive |
| 7 | 147,103,014.00 | rs6962089 | 2.0493 | 22_bit_twopoint_parametric_recessive |
| 7 | 148,238,607.00 | rs6464862 | 1.8978 | 22_bit_twopoint_parametric_recessive |
| 7 | 148,239,711.00 | GSA-rs1534702 | 1.8993 | 22_bit_twopoint_parametric_recessive |
| 7 | 148,277,563.00 | JHU_7.147974654 | 2.004 | 21_bit_twopoint_NPL |
| 7 | 148,277,563.00 | JHU_7.147974654 | 2.2343 | 22_bit_twopoint_parametric_recessive |
| 7 | 148,655,941.00 | rs6963551 | 1.9096 | 21_bit_twopoint_parametric_recessive |
| 7 | 148,655,941.00 | rs6963551 | 2.1978 | 21_bit_twopoint_parametric_dominant |
| 7 | 148,658,364.00 | rs2041074 | 2.07 | 21_bit_twopoint_parametric_dominant |
| 7 | 148,783,898.00 | rs10271133 | 2.3734 | 21_bit_twopoint_parametric_recessive |
| 7 | 149,236,851.00 | rs7779475 | 2.5262 | 21_bit_twopoint_parametric_recessive |
| 7 | 149,236,851.00 | rs7779475 | 2.6065 | 21_bit_twopoint_parametric_dominant |
| 7 | 149,448,048.00 | rs6973105 | 2.7457 | 22_bit_twopoint_parametric_recessive |
| 7 | 149,475,803.00 | rs62491927 | 1.987 | 21_bit_twopoint_parametric_recessive |
| 7 | 149,912,859.00 | JHU_7.149609947 | 1.8949 | 21_bit_twopoint_parametric_dominant |
| 7 | 149,912,859.00 | JHU_7.149609947 | 1.9174 | 22_bit_twopoint_parametric_dominant |
| 7 | 150,793,288.00 | exm671163 | 3.2582 | 22_bit_twopoint_parametric_recessive |
| 7 | 150,793,996.00 | GSA-rs3173833 | 2.1999 | 22_bit_twopoint_parametric_recessive |
| 7 | 151,193,154.00 | rs2487151 | 1.9584 | 21_bit_twopoint_parametric_recessive |
| 7 | 151,260,831.00 | GSA-rs78461635 | 2.0103 | 21_bit_twopoint_parametric_dominant |
| 7 | 151,260,831.00 | GSA-rs78461635 | 2.1224 | 21_bit_twopoint_parametric_recessive |
| 7 | 151,262,740.00 | GSA-rs10487756 | 2.0371 | 21_bit_twopoint_parametric_dominant |
| 7 | 151,262,740.00 | GSA-rs10487756 | 2.0863 | 21_bit_twopoint_parametric_recessive |
| 7 | 153,006,311.00 | rs520556 | 1.9673 | 21_bit_twopoint_parametric_dominant |
| 7 | 153,021,857.00 | rs567778 | 1.9648 | 22_bit_twopoint_parametric_dominant |
| 7 | 153,325,289.00 | rs422283 | 1.9207 | 22_bit_twopoint_parametric_dominant |
| 7 | 153,372,026.00 | rs7793529 | 2.1997 | 21_bit_twopoint_parametric_recessive |
| 7 | 154,125,963.00 | rs12533045 | 2.446 | 22_bit_twopoint_parametric_dominant |
| 7 | 154,149,745.00 | JHU_7.153846829 | 2.0438 | 22_bit_twopoint_parametric_dominant |
| 7 | 154,262,770.00 | rs7384911 | 2.0176 | 22_bit_multipoint_parametric_recessive |
| 7 | 154,262,770.00 | rs7384911 | 2.0356 | 22_bit_twopoint_parametric_recessive |
| 7 | 154,289,990.00 | rs9655634 | 2.6129 | 22_bit_twopoint_parametric_recessive |
| 7 | 154,298,322.00 | rs7787841 | 1.9896 | 21_bit_twopoint_parametric_recessive |
| 7 | 154,298,322.00 | rs7787841 | 2.7358 | 22_bit_twopoint_parametric_recessive |
| 7 | 154,478,870.00 | rs12535735 | 1.9499 | 22_bit_twopoint_parametric_dominant |
| 7 | 155,188,458.00 | GSA-rs7790169 | 2.2379 | 21_bit_twopoint_parametric_dominant |
| 7 | 155,476,577.00 | JHU_7.155269271 | 1.9175 | 22_bit_twopoint_parametric_recessive |
| 7 | 155,476,577.00 | JHU_7.155269271 | 2.948 | 22_bit_twopoint_parametric_dominant |
| 7 | 155,905,561.00 | rs6459961 | 1.9157 | 21_bit_twopoint_parametric_recessive |
| 7 | 156,144,923.00 | JHU_7.155937616 | 2.6288 | 21_bit_twopoint_parametric_recessive |
| 7 | 156,198,119.00 | rs737681 | 1.8733 | 22_bit_twopoint_parametric_recessive |
| 7 | 158,911,363.00 | GSA-rs3815215 | 2.0994 | 21_bit_twopoint_parametric_recessive |
| 7 | 158,911,363.00 | GSA-rs3815215 | 2.1979 | 22_bit_twopoint_parametric_recessive |
| 7 | 158,926,562.00 | GSA-rs3815214 | 2.3306 | 22_bit_twopoint_parametric_recessive |
| 7 | 158,926,562.00 | GSA-rs3815214 | 2.5034 | 21_bit_twopoint_parametric_recessive |
| 7 | 158,928,108.00 | GSA-rs3793183 | 2.1985 | 21_bit_twopoint_parametric_recessive |
| 7 | 158,928,108.00 | GSA-rs3793183 | 2.628 | 22_bit_twopoint_parametric_recessive |
| 7 | 159,161,907.00 | rs11489218 | 1.8829 | 22_bit_twopoint_parametric_dominant |
| 7 | 159,162,201.00 | JHU_7.158954891 | 1.9333 | 22_bit_twopoint_parametric_dominant |
| 8 | 952,064.00 | rs17667613 | 2.1188 | 21_bit_twopoint_parametric_dominant |
| 8 | 2,035,440.00 | rs7814914 | 2.2979 | 21_bit_twopoint_parametric_dominant |
| 8 | 2,605,071.00 | JHU_8.2462161 | 1.9087 | 21_bit_twopoint_parametric_recessive |
| 8 | 2,605,071.00 | JHU_8.2462161 | 2.0133 | 22_bit_twopoint_parametric_recessive |
| 8 | 3,633,300.00 | rs10089138 | 2.7167 | 22_bit_twopoint_parametric_dominant |
| 8 | 4,576,646.00 | rs7843036 | 2.6958 | 21_bit_twopoint_parametric_dominant |
| 8 | 4,576,646.00 | rs7843036 | 2.6958 | 21_bit_twopoint_parametric_dominant |
| 8 | 5,687,568.00 | rs7831044 | 2.0496 | 21_bit_twopoint_parametric_recessive |
| 8 | 8,614,994.00 | rs9329273 | 2.0624 | 22_bit_twopoint_parametric_recessive |
| 8 | 8,614,994.00 | rs9329273 | 2.1341 | 22_bit_twopoint_parametric_dominant |
| 8 | 8,638,567.00 | rs11250215 | 2.1878 | 21_bit_twopoint_parametric_dominant |
| 8 | 8,732,273.00 | rs11781985 | 2.0956 | 21_bit_twopoint_parametric_recessive |
| 8 | 8,732,273.00 | rs11781985 | 3.2689 | 21_bit_twopoint_parametric_dominant |
| 8 | 8,732,273.00 | rs11781985 | 3.2781 | 22_bit_twopoint_parametric_dominant |
| 8 | 9,003,920.00 | GSA-rs13252797 | 2.0239 | 21_bit_twopoint_parametric_dominant |
| 8 | 9,003,920.00 | GSA-rs13252797 | 2.9775 | 22_bit_twopoint_parametric_dominant |
| 8 | 9,003,920.00 | GSA-rs13252797 | 3.127 | 22_bit_twopoint_parametric_recessive |
| 8 | 9,122,490.00 | rs10092662 | 2.4886 | 21_bit_twopoint_parametric_dominant |
| 8 | 9,133,067.00 | rs189798 | 2.2219 | 22_bit_twopoint_parametric_dominant |
| 8 | 9,293,716.00 | rs719409 | 1.9471 | 22_bit_twopoint_parametric_recessive |
| 8 | 9,293,716.00 | rs719409 | 2.1203 | 21_bit_twopoint_parametric_dominant |
| 8 | 9,293,716.00 | rs719409 | 2.599 | 22_bit_twopoint_parametric_dominant |
| 8 | 10,391,970.00 | rs17765901 | 1.8967 | 21_bit_twopoint_parametric_dominant |
| 8 | 10,391,970.00 | rs17765901 | 1.8967 | 21_bit_twopoint_parametric_dominant |
| 8 | 10,875,482.00 | JHU_8.10732991 | 2.1138 | 21_bit_twopoint_parametric_dominant |
| 8 | 11,182,707.00 | rs73198970 | 2.2063 | 22_bit_twopoint_parametric_dominant |
| 8 | 11,182,707.00 | rs73198970 | 2.3369 | 22_bit_twopoint_parametric_recessive |
| 8 | 11,278,465.00 | GSA-rs7004362 | 2.1098 | 21_bit_twopoint_parametric_dominant |
| 8 | 11,285,763.00 | GSA-rs3808518 | 2.279 | 21_bit_twopoint_parametric_dominant |
| 8 | 11,582,510.00 | rs4841567 | 2.2824 | 21_bit_twopoint_parametric_dominant |
| 8 | 13,990,451.00 | rs11780201 | 2.1797 | 21_bit_twopoint_parametric_dominant |
| 8 | 14,344,164.00 | GSA-rs113291523 | 2.499 | 21_bit_twopoint_parametric_recessive |
| 8 | 14,768,147.00 | GSA-rs2255678 | 2.8233 | 21_bit_twopoint_parametric_dominant |
| 8 | 15,170,138.00 | JHU_8.15027646 | 1.8757 | 21_bit_twopoint_parametric_dominant |
| 8 | 15,182,355.00 | rs7820870 | 1.8783 | 21_bit_twopoint_parametric_dominant |
| 8 | 18,507,264.00 | GSA-rs901938 | 2.032 | 22_bit_twopoint_parametric_recessive |
| 8 | 18,864,802.00 | rs768073 | 2.1366 | 21_bit_twopoint_parametric_dominant |
| 8 | 18,924,266.00 | rs11779965 | 2.2063 | 21_bit_twopoint_parametric_recessive |
| 8 | 21,247,834.00 | rs2446614 | 1.8733 | 22_bit_twopoint_parametric_dominant |
| 8 | 26,434,628.00 | rs13252617 | 1.9441 | 22_bit_twopoint_parametric_recessive |
| 8 | 26,434,628.00 | rs13252617 | 2.3 | 22_bit_twopoint_NPL |
| 8 | 31,632,958.00 | rs4733263 | 2.9116 | 22_bit_twopoint_parametric_dominant |
| 8 | 31,638,065.00 | 8:31495581-C-T | 2.4904 | 22_bit_twopoint_parametric_dominant |
| 8 | 31,859,446.00 | rs776401 | 2.0338 | 21_bit_twopoint_parametric_dominant |
| 8 | 37,337,037.00 | rs462931 | 2.3204 | 22_bit_twopoint_parametric_dominant |
| 8 | 40,031,908.00 | rs75386109 | 2.7838 | 21_bit_twopoint_parametric_dominant |
| 8 | 40,031,908.00 | rs75386109 | 3.2267 | 22_bit_twopoint_parametric_dominant |
| 8 | 56,568,278.00 | rs2670040 | 1.8759 | 22_bit_twopoint_parametric_recessive |
| 8 | 56,910,161.00 | GSA-rs11992437 | 1.9433 | 22_bit_twopoint_parametric_dominant |
| 8 | 61,067,314.00 | rs4738861 | 2.0043 | 21_bit_twopoint_parametric_dominant |
| 8 | 61,067,314.00 | rs4738861 | 2.0043 | 21_bit_twopoint_parametric_dominant |
| 8 | 61,090,192.00 | rs1985815 | 3.1651 | 21_bit_twopoint_parametric_dominant |
| 8 | 61,090,192.00 | rs1985815 | 3.1651 | 21_bit_twopoint_parametric_dominant |
| 8 | 67,656,369.00 | JHU_8.68568603 | 2.1572 | 22_bit_twopoint_parametric_recessive |
| 8 | 71,306,382.00 | rs11993876 | 1.8871 | 21_bit_twopoint_parametric_dominant |
| 8 | 71,606,939.00 | rs1544844 | 2.6005 | 22_bit_twopoint_parametric_dominant |
| 8 | 71,735,564.00 | rs9694975 | 2.2493 | 22_bit_twopoint_parametric_dominant |
| 8 | 71,755,173.00 | JHU_8.72667407 | 1.9673 | 22_bit_twopoint_parametric_dominant |
| 8 | 73,251,543.00 | rs2925444 | 2.1203 | 22_bit_twopoint_parametric_dominant |
| 8 | 73,251,543.00 | rs2925444 | 2.2988 | 21_bit_twopoint_parametric_dominant |
| 8 | 78,130,096.00 | rs10504670 | 1.8758 | 21_bit_twopoint_parametric_recessive |
| 8 | 81,377,603.00 | rs1450790 | 1.9957 | 22_bit_twopoint_parametric_dominant |
| 8 | 83,087,868.00 | rs4739920 | 2.325 | 21_bit_twopoint_parametric_dominant |
| 8 | 89,354,598.00 | rs2086343 | 2.8485 | 21_bit_twopoint_parametric_dominant |
| 8 | 94,055,714.00 | GSA-rs7013178 | 2.2387 | 21_bit_twopoint_parametric_dominant |
| 8 | 94,056,770.00 | GSA-rs3018892 | 1.8969 | 21_bit_twopoint_parametric_dominant |
| 8 | 104,072,113.00 | exm2271006 | 1.9718 | 21_bit_twopoint_parametric_recessive |
| 8 | 104,072,113.00 | exm2271006 | 2.8934 | 21_bit_twopoint_parametric_dominant |
| 8 | 104,280,337.00 | rs16871199 | 2.0153 | 22_bit_twopoint_parametric_recessive |
| 8 | 105,546,199.00 | rs117044043 | 2.0242 | 22_bit_twopoint_parametric_recessive |
| 8 | 105,546,199.00 | rs117044043 | 2.1425 | 22_bit_twopoint_parametric_dominant |
| 8 | 106,411,834.00 | rs10109171 | 2.1575 | 22_bit_twopoint_parametric_dominant |
| 8 | 114,852,842.00 | JHU_8.115865070 | 2.3805 | 22_bit_twopoint_parametric_dominant |
| 8 | 117,758,326.00 | rs4876741 | 1.86 | 21_bit_twopoint_NPL |
| 8 | 117,758,326.00 | rs4876741 | 2.9056 | 21_bit_twopoint_parametric_dominant |
| 8 | 117,794,184.00 | rs4876754 | 2.2388 | 21_bit_twopoint_parametric_dominant |
| 8 | 118,337,292.00 | JHU_8.119349530 | 2.0553 | 22_bit_twopoint_parametric_dominant |
| 8 | 121,662,004.00 | GSA-rs7008033 | 2.0762 | 22_bit_twopoint_parametric_dominant |
| 8 | 122,698,904.00 | rs7018013 | 1.9085 | 22_bit_twopoint_parametric_recessive |
| 8 | 125,762,070.00 | rs1444003 | 1.9995 | 22_bit_twopoint_parametric_recessive |
| 8 | 125,794,291.00 | rs1023794 | 2.1931 | 22_bit_twopoint_parametric_recessive |
| 8 | 126,004,315.00 | rs4483112 | 2.0628 | 21_bit_twopoint_parametric_dominant |
| 8 | 127,310,936.00 | exm-rs445114 | 2.1953 | 21_bit_twopoint_parametric_recessive |
| 8 | 131,244,270.00 | GSA-rs78971670 | 2.3262 | 22_bit_twopoint_parametric_recessive |
| 8 | 131,244,270.00 | GSA-rs78971670 | 2.3262 | 22_bit_twopoint_parametric_recessive |
| 8 | 131,713,484.00 | rs587639 | 1.9585 | 21_bit_twopoint_parametric_dominant |
| 8 | 133,403,614.00 | rs79792639 | 2.3121 | 22_bit_twopoint_parametric_dominant |
| 8 | 133,697,656.00 | rs10956719 | 2.2299 | 22_bit_multipoint_parametric_dominant |
| 8 | 133,697,656.00 | rs10956719 | 2.2304 | 22_bit_twopoint_parametric_dominant |
| 8 | 134,217,751.00 | GSA-rs16905126 | 2.2039 | 22_bit_twopoint_parametric_recessive |
| 8 | 134,232,588.00 | rs59410498 | 1.8941 | 22_bit_twopoint_parametric_recessive |
| 8 | 135,099,991.00 | rs12155615 | 1.8887 | 21_bit_twopoint_parametric_recessive |
| 8 | 135,099,991.00 | rs12155615 | 1.892 | 21_bit_multipoint_parametric_recessive |
| 8 | 135,188,316.00 | rs7002465 | 1.9776 | 22_bit_twopoint_parametric_dominant |
| 8 | 135,191,691.00 | rs7820620 | 1.9787 | 22_bit_twopoint_parametric_dominant |
| 8 | 135,226,451.00 | GSA-rs6578225 | 2.1854 | 22_bit_twopoint_parametric_recessive |
| 8 | 135,258,230.00 | rs7004726 | 1.9708 | 21_bit_twopoint_parametric_recessive |
| 8 | 135,281,563.00 | rs10095580 | 2.0802 | 21_bit_twopoint_parametric_recessive |
| 8 | 135,292,909.00 | JHU_8.136305151 | 2.0004 | 22_bit_twopoint_parametric_recessive |
| 8 | 135,292,909.00 | JHU_8.136305151 | 2.3196 | 21_bit_twopoint_parametric_dominant |
| 8 | 135,318,578.00 | rs2925046 | 1.8932 | 21_bit_twopoint_parametric_dominant |
| 8 | 135,693,732.00 | rs6984541 | 2.2403 | 21_bit_twopoint_parametric_dominant |
| 8 | 135,693,732.00 | rs6984541 | 2.2745 | 22_bit_twopoint_parametric_recessive |
| 8 | 135,693,732.00 | rs6984541 | 2.3684 | 21_bit_twopoint_parametric_recessive |
| 8 | 135,693,732.00 | rs6984541 | 2.5736 | 22_bit_twopoint_parametric_dominant |
| 8 | 135,920,274.00 | GSA-rs13275610 | 2.0448 | 22_bit_twopoint_parametric_recessive |
| 8 | 136,330,904.00 | GSA-rs7822138 | 2.1413 | 21_bit_twopoint_parametric_recessive |
| 8 | 136,339,446.00 | rs16905672 | 2.3837 | 21_bit_twopoint_parametric_dominant |
| 8 | 136,339,446.00 | rs16905672 | 2.4825 | 21_bit_twopoint_parametric_recessive |
| 8 | 139,804,949.00 | rs4736007 | 1.9022 | 21_bit_multipoint_parametric_dominant |
| 8 | 139,804,949.00 | rs4736007 | 1.9059 | 21_bit_twopoint_parametric_dominant |
| 8 | 139,804,949.00 | rs4736007 | 2.0105 | 22_bit_twopoint_parametric_dominant |
| 8 | 139,804,949.00 | rs4736007 | 2.0359 | 22_bit_multipoint_parametric_dominant |
| 8 | 141,195,033.00 | exm2266702 | 2.1202 | 21_bit_multipoint_parametric_recessive |
| 8 | 141,195,033.00 | exm2266702 | 2.1202 | 21_bit_twopoint_parametric_recessive |
| 8 | 141,195,865.00 | rs10102728 | 1.9184 | 21_bit_twopoint_parametric_recessive |
| 8 | 141,505,299.00 | rs2748414 | 2.1473 | 21_bit_twopoint_parametric_recessive |
| 8 | 141,546,705.00 | rs1566080 | 1.8608 | 22_bit_twopoint_parametric_dominant |
| 8 | 141,578,318.00 | rs72687459 | 2.288 | 21_bit_twopoint_parametric_recessive |
| 8 | 143,043,095.00 | rs11136299 | 2.5697 | 21_bit_twopoint_parametric_dominant |
| 8 | 143,055,202.00 | JHU_8.144136618 | 2.3557 | 22_bit_twopoint_parametric_dominant |
| 8 | 143,122,435.00 | rs7820178 | 2.2972 | 21_bit_twopoint_parametric_dominant |
| 8 | 143,123,456.00 | JHU_8.144204872 | 2.2931 | 21_bit_twopoint_parametric_dominant |
| 9 | 477,236.00 | GSA-rs62531956 | 2.0641 | 21_bit_twopoint_parametric_recessive |
| 9 | 477,236.00 | GSA-rs62531956 | 2.1056 | 22_bit_twopoint_parametric_dominant |
| 9 | 478,530.00 | GSA-rs2282038 | 2.2117 | 22_bit_twopoint_parametric_dominant |
| 9 | 479,157.00 | GSA-rs10974741 | 2.3804 | 22_bit_twopoint_parametric_dominant |
| 9 | 1,047,343.00 | GSA-rs4740448 | 2.5366 | 21_bit_twopoint_parametric_dominant |
| 9 | 1,238,454.00 | GSA-rs72689507 | 2.418 | 21_bit_multipoint_NPL |
| 9 | 2,198,627.00 | GSA-rs10965177 | 2.2015 | 22_bit_twopoint_parametric_dominant |
| 9 | 2,678,077.00 | GSA-rs6475927 | 2.1182 | 22_bit_twopoint_parametric_recessive |
| 9 | 2,938,757.00 | GSA-rs7025901 | 1.9098 | 21_bit_twopoint_parametric_dominant |
| 9 | 3,671,251.00 | GSA-rs10511449 | 1.862 | 22_bit_twopoint_parametric_recessive |
| 9 | 4,267,209.00 | rs1571583 | 1.9444 | 21_bit_twopoint_parametric_dominant |
| 9 | 4,604,807.00 | GSA-rs301453 | 3.2131 | 21_bit_twopoint_parametric_recessive |
| 9 | 5,844,943.00 | GSA-rs16923551 | 2.4033 | 22_bit_twopoint_parametric_recessive |
| 9 | 5,844,943.00 | GSA-rs16923551 | 2.4033 | 22_bit_twopoint_parametric_recessive |
| 9 | 5,856,209.00 | GSA-rs6477005 | 1.9992 | 22_bit_twopoint_parametric_recessive |
| 9 | 5,856,209.00 | GSA-rs6477005 | 1.9992 | 22_bit_twopoint_parametric_recessive |
| 9 | 5,859,564.00 | GSA-rs7041526 | 2.0176 | 21_bit_twopoint_parametric_recessive |
| 9 | 5,859,564.00 | GSA-rs7041526 | 2.0176 | 21_bit_twopoint_parametric_recessive |
| 9 | 5,859,564.00 | GSA-rs7041526 | 2.4049 | 22_bit_twopoint_parametric_recessive |
| 9 | 5,859,564.00 | GSA-rs7041526 | 2.4049 | 22_bit_twopoint_parametric_recessive |
| 9 | 5,861,170.00 | GSA-rs7026604 | 2.1515 | 21_bit_twopoint_parametric_recessive |
| 9 | 5,861,170.00 | GSA-rs7026604 | 2.1515 | 21_bit_twopoint_parametric_recessive |
| 9 | 5,864,988.00 | GSA-rs1418742 | 2.6846 | 22_bit_twopoint_parametric_recessive |
| 9 | 5,864,988.00 | GSA-rs1418742 | 2.6846 | 22_bit_twopoint_parametric_recessive |
| 9 | 9,020,223.00 | GSA-rs378363 | 2.2323 | 21_bit_twopoint_parametric_dominant |
| 9 | 9,441,229.00 | GSA-rs56409001 | 1.9076 | 22_bit_twopoint_parametric_dominant |
| 9 | 9,464,561.00 | GSA-rs10491613 | 2.0664 | 22_bit_twopoint_parametric_dominant |
| 9 | 12,227,068.00 | GSA-rs10960537 | 2.3263 | 22_bit_twopoint_parametric_recessive |
| 9 | 12,227,068.00 | GSA-rs10960537 | 2.3263 | 22_bit_twopoint_parametric_recessive |
| 9 | 12,227,068.00 | GSA-rs10960537 | 3.0809 | 21_bit_twopoint_parametric_dominant |
| 9 | 12,227,068.00 | GSA-rs10960537 | 3.0809 | 21_bit_twopoint_parametric_dominant |
| 9 | 12,267,132.00 | GSA-rs10809744 | 1.9382 | 22_bit_twopoint_parametric_recessive |
| 9 | 12,267,132.00 | GSA-rs10809744 | 1.9382 | 22_bit_twopoint_parametric_recessive |
| 9 | 12,267,132.00 | GSA-rs10809744 | 2.4997 | 21_bit_twopoint_parametric_dominant |
| 9 | 12,267,132.00 | GSA-rs10809744 | 2.4997 | 21_bit_twopoint_parametric_dominant |
| 9 | 12,272,705.00 | GSA-rs6474694 | 1.9248 | 21_bit_twopoint_parametric_dominant |
| 9 | 12,272,705.00 | GSA-rs6474694 | 1.9248 | 21_bit_twopoint_parametric_dominant |
| 9 | 14,556,661.00 | GSA-rs10756576 | 1.94 | 22_bit_twopoint_parametric_recessive |
| 9 | 14,832,351.00 | GSA-rs9792613 | 2.2008 | 22_bit_twopoint_parametric_dominant |
| 9 | 17,655,332.00 | GSA-rs1536064 | 2.471 | 21_bit_twopoint_parametric_dominant |
| 9 | 17,674,078.00 | GSA-rs7020158 | 2.3896 | 21_bit_twopoint_parametric_dominant |
| 9 | 17,763,134.00 | GSA-rs3808673 | 1.9097 | 21_bit_twopoint_parametric_dominant |
| 9 | 18,515,349.00 | GSA-rs73421042 | 2.762 | 22_bit_twopoint_parametric_dominant |
| 9 | 18,516,582.00 | GSA-rs11792817 | 2.4137 | 22_bit_twopoint_parametric_dominant |
| 9 | 18,653,326.00 | GSA-rs702203 | 2.27 | 21_bit_twopoint_parametric_dominant |
| 9 | 23,050,747.00 | GSA-rs1331597 | 2.0902 | 21_bit_twopoint_parametric_dominant |
| 9 | 23,546,463.00 | GSA-rs7028484 | 1.9762 | 21_bit_twopoint_parametric_recessive |
| 9 | 24,925,472.00 | GSA-rs7023908 | 1.9353 | 22_bit_twopoint_parametric_recessive |
| 9 | 24,961,154.00 | GSA-rs35963392 | 2.0041 | 22_bit_twopoint_parametric_recessive |
| 9 | 26,507,213.00 | GSA-rs12339423 | 2.342 | 21_bit_twopoint_parametric_dominant |
| 9 | 27,342,059.00 | GSA-rs12376262 | 2.1598 | 21_bit_twopoint_parametric_dominant |
| 9 | 30,804,874.00 | GSA-rs2502921 | 2.0062 | 21_bit_twopoint_parametric_dominant |
| 9 | 31,680,288.00 | GSA-rs67477866 | 2.0461 | 21_bit_twopoint_parametric_recessive |
| 9 | 31,811,671.00 | GSA-rs1928091 | 1.8883 | 21_bit_twopoint_parametric_recessive |
| 9 | 32,784,840.00 | GSA-rs1810807 | 2.2194 | 22_bit_twopoint_parametric_recessive |
| 9 | 35,815,359.00 | exm2271184 | 2.2807 | 21_bit_twopoint_parametric_recessive |
| 9 | 36,958,585.00 | 9:36958582-A-G | 2.6125 | 22_bit_twopoint_parametric_recessive |
| 9 | 37,731,010.00 | GSA-rs2274324 | 2.4757 | 21_bit_twopoint_parametric_dominant |
| 9 | 37,731,010.00 | GSA-rs2274324 | 2.5321 | 22_bit_twopoint_parametric_dominant |
| 9 | 38,372,180.00 | GSA-rs12351689 | 2.0869 | 22_bit_twopoint_parametric_recessive |
| 9 | 38,380,392.00 | GSA-rs10973770 | 2.3415 | 22_bit_twopoint_parametric_recessive |
| 9 | 38,380,392.00 | GSA-rs10973770 | 2.5648 | 21_bit_twopoint_parametric_recessive |
| 9 | 38,727,088.00 | GSA-rs10125906 | 2.1157 | 22_bit_twopoint_parametric_recessive |
| 9 | 38,727,088.00 | GSA-rs10125906 | 2.1157 | 22_bit_twopoint_parametric_recessive |
| 9 | 68,867,372.00 | GSA-rs2152647 | 2.0305 | 21_bit_twopoint_parametric_recessive |
| 9 | 71,630,352.00 | GSA-rs6560217 | 2.0683 | 22_bit_twopoint_parametric_dominant |
| 9 | 73,641,820.00 | GSA-rs1352514 | 1.8971 | 21_bit_twopoint_parametric_recessive |
| 9 | 74,685,003.00 | GSA-rs77600772 | 2.6238 | 21_bit_twopoint_parametric_dominant |
| 9 | 74,685,003.00 | GSA-rs77600772 | 3.2465 | 22_bit_twopoint_parametric_recessive |
| 9 | 74,696,783.00 | GSA-rs79485647 | 1.8684 | 21_bit_twopoint_parametric_dominant |
| 9 | 74,696,783.00 | GSA-rs79485647 | 2.4829 | 22_bit_twopoint_parametric_recessive |
| 9 | 74,702,207.00 | GSA-rs11791579 | 2.6049 | 22_bit_twopoint_parametric_recessive |
| 9 | 75,471,698.00 | GSA-rs74500225 | 1.9102 | 22_bit_twopoint_parametric_recessive |
| 9 | 75,471,698.00 | GSA-rs74500225 | 2.1617 | 22_bit_twopoint_parametric_dominant |
| 9 | 75,652,183.00 | GSA-rs7024078 | 2.134 | 21_bit_twopoint_parametric_dominant |
| 9 | 75,975,753.00 | GSA-rs10781322 | 1.9571 | 22_bit_twopoint_parametric_dominant |
| 9 | 77,515,630.00 | GSA-rs17195438 | 2.2887 | 21_bit_twopoint_parametric_dominant |
| 9 | 80,433,088.00 | GSA-rs11138659 | 1.9844 | 21_bit_twopoint_parametric_dominant |
| 9 | 80,433,088.00 | GSA-rs11138659 | 2.0407 | 21_bit_twopoint_parametric_recessive |
| 9 | 80,565,943.00 | GSA-rs11138734 | 2.4491 | 21_bit_twopoint_parametric_recessive |
| 9 | 81,567,319.00 | GSA-rs2777765 | 2.3855 | 21_bit_twopoint_parametric_dominant |
| 9 | 81,576,352.00 | GSA-rs7027019 | 2.1687 | 21_bit_twopoint_parametric_dominant |
| 9 | 84,349,741.00 | GSA-rs13298157 | 2.2554 | 22_bit_twopoint_parametric_dominant |
| 9 | 84,958,341.00 | GSA-rs56405676 | 1.982 | 22_bit_twopoint_NPL |
| 9 | 84,958,341.00 | GSA-rs56405676 | 2.161 | 22_bit_twopoint_parametric_recessive |
| 9 | 84,958,431.00 | GSA-rs28435670 | 1.861 | 22_bit_twopoint_NPL |
| 9 | 85,388,753.00 | GSA-rs895606 | 2.0208 | 21_bit_twopoint_parametric_dominant |
| 9 | 85,388,753.00 | GSA-rs895606 | 2.211 | 22_bit_twopoint_parametric_dominant |
| 9 | 86,414,788.00 | GSA-rs187117 | 2.2305 | 22_bit_twopoint_parametric_recessive |
| 9 | 86,814,766.00 | GSA-rs4877337 | 2.0069 | 22_bit_twopoint_parametric_recessive |
| 9 | 88,193,667.00 | GSA-rs4877127 | 1.9292 | 21_bit_twopoint_parametric_dominant |
| 9 | 92,984,654.00 | GSA-rs10992564 | 2.2587 | 22_bit_twopoint_parametric_dominant |
| 9 | 92,988,449.00 | GSA-rs10992566 | 2.2587 | 22_bit_twopoint_parametric_dominant |
| 9 | 93,143,270.00 | GSA-rs4743894 | 1.921 | 21_bit_twopoint_parametric_dominant |
| 9 | 99,583,747.00 | GSA-rs10819687 | 2.1608 | 22_bit_twopoint_parametric_recessive |
| 9 | 99,818,277.00 | GSA-rs56002013 | 1.8615 | 22_bit_twopoint_parametric_recessive |
| 9 | 99,818,277.00 | GSA-rs56002013 | 2.5908 | 21_bit_twopoint_parametric_recessive |
| 9 | 102,985,343.00 | GSA-rs78350186 | 1.9197 | 22_bit_twopoint_parametric_dominant |
| 9 | 104,782,875.00 | GSA-rs4149339 | 1.8932 | 21_bit_twopoint_parametric_dominant |
| 9 | 107,673,587.00 | GSA-rs10816550 | 1.9548 | 22_bit_twopoint_parametric_dominant |
| 9 | 107,673,587.00 | GSA-rs10816550 | 1.9548 | 22_bit_twopoint_parametric_dominant |
| 9 | 110,127,035.00 | GSA-rs914355 | 2.0152 | 21_bit_twopoint_parametric_dominant |
| 9 | 113,243,503.00 | GSA-rs10817465 | 2.8794 | 22_bit_twopoint_parametric_dominant |
| 9 | 114,318,108.00 | GSA-rs2636881 | 2.115 | 21_bit_twopoint_parametric_dominant |
| 9 | 114,408,030.00 | GSA-rs2274160 | 2.0354 | 21_bit_twopoint_parametric_dominant |
| 9 | 114,691,388.00 | GSA-rs16930907 | 2.0053 | 22_bit_twopoint_parametric_recessive |
| 9 | 115,778,789.00 | GSA-rs2418412 | 2.104 | 22_bit_twopoint_parametric_dominant |
| 9 | 115,778,789.00 | GSA-rs2418412 | 2.104 | 22_bit_twopoint_parametric_dominant |
| 9 | 116,522,865.00 | GSA-rs4837587 | 2.189 | 21_bit_twopoint_parametric_dominant |
| 9 | 122,365,436.00 | GSA-rs10818682 | 2.0261 | 21_bit_twopoint_parametric_dominant |
| 9 | 122,407,651.00 | GSA-rs10818684 | 1.8741 | 21_bit_twopoint_parametric_dominant |
| 9 | 122,723,877.00 | GSA-rs16912140 | 1.8783 | 22_bit_twopoint_parametric_recessive |
| 9 | 124,021,541.00 | GSA-rs60483758 | 2.0913 | 21_bit_twopoint_parametric_recessive |
| 9 | 124,021,541.00 | GSA-rs60483758 | 2.4216 | 22_bit_twopoint_parametric_recessive |
| 9 | 126,442,268.00 | GSA-rs10987278 | 2.3723 | 21_bit_twopoint_parametric_dominant |
| 9 | 126,442,268.00 | GSA-rs10987278 | 2.3723 | 21_bit_twopoint_parametric_dominant |
| 9 | 129,867,508.00 | GSA-rs2417157 | 2.1549 | 22_bit_twopoint_parametric_dominant |
| 9 | 130,678,606.00 | GSA-rs7021384 | 1.9143 | 22_bit_twopoint_parametric_recessive |
| 9 | 130,678,606.00 | GSA-rs7021384 | 2.1369 | 22_bit_twopoint_parametric_dominant |
| 9 | 130,982,735.00 | GSA-rs2178107 | 2.0855 | 21_bit_twopoint_parametric_dominant |
| 9 | 133,028,610.00 | GSA-rs623489 | 2.402 | 21_bit_twopoint_parametric_recessive |
| 9 | 133,038,096.00 | GSA-rs675167 | 3.2393 | 21_bit_twopoint_parametric_dominant |
| 9 | 133,174,393.00 | rs6597604 | 2.1821 | 21_bit_twopoint_parametric_dominant |
| 10 | 2,059,060.00 | rs17809678 | 1.8648 | 21_bit_twopoint_parametric_recessive |
| 10 | 2,059,060.00 | rs17809678 | 2.2003 | 21_bit_twopoint_parametric_dominant |
| 10 | 2,921,468.00 | GSA-rs1574052 | 1.889 | 22_bit_twopoint_parametric_dominant |
| 10 | 3,055,020.00 | exm2249192 | 1.8745 | 22_bit_twopoint_parametric_dominant |
| 10 | 3,825,904.00 | JHU_10.3868095 | 1.9128 | 22_bit_twopoint_parametric_recessive |
| 10 | 4,983,904.00 | rs7909151 | 2.1968 | 22_bit_twopoint_parametric_dominant |
| 10 | 4,994,249.00 | rs9804392 | 2.0052 | 22_bit_twopoint_parametric_recessive |
| 10 | 5,033,301.00 | rs12249281 | 2.0013 | 22_bit_twopoint_parametric_dominant |
| 10 | 5,065,603.00 | rs12570598 | 2.6343 | 22_bit_twopoint_parametric_dominant |
| 10 | 6,982,737.00 | GSA-rs2486871 | 2.0545 | 21_bit_multipoint_parametric_dominant |
| 10 | 6,982,737.00 | GSA-rs2486871 | 2.0623 | 21_bit_twopoint_parametric_dominant |
| 10 | 8,066,629.00 | rs569421 | 2.1452 | 21_bit_twopoint_parametric_recessive |
| 10 | 8,414,044.00 | rs11255686 | 2.1094 | 21_bit_twopoint_parametric_dominant |
| 10 | 11,121,888.00 | rs7894817 | 1.8623 | 22_bit_twopoint_parametric_recessive |
| 10 | 11,121,888.00 | rs7894817 | 2.1117 | 22_bit_twopoint_parametric_dominant |
| 10 | 11,727,587.00 | rs72779878 | 1.9613 | 21_bit_twopoint_parametric_recessive |
| 10 | 11,738,838.00 | rs2055040 | 2.2128 | 22_bit_twopoint_parametric_dominant |
| 10 | 12,416,033.00 | rs2399860 | 1.9871 | 21_bit_twopoint_parametric_dominant |
| 10 | 12,457,861.00 | rs2399849 | 1.9746 | 21_bit_twopoint_parametric_recessive |
| 10 | 12,457,861.00 | rs2399849 | 2.6822 | 21_bit_twopoint_parametric_dominant |
| 10 | 12,576,351.00 | rs56956284 | 2.2196 | 21_bit_twopoint_parametric_dominant |
| 10 | 12,576,351.00 | rs56956284 | 2.2471 | 22_bit_twopoint_parametric_dominant |
| 10 | 12,592,504.00 | rs9787594 | 2.4174 | 22_bit_twopoint_parametric_dominant |
| 10 | 12,592,504.00 | rs9787594 | 2.4842 | 21_bit_twopoint_parametric_dominant |
| 10 | 12,597,168.00 | rs57519070 | 2.4174 | 22_bit_twopoint_parametric_dominant |
| 10 | 12,597,168.00 | rs57519070 | 2.4842 | 21_bit_twopoint_parametric_dominant |
| 10 | 13,661,065.00 | rs9787417 | 1.9128 | 21_bit_twopoint_parametric_dominant |
| 10 | 14,384,966.00 | rs673875 | 2.1565 | 21_bit_twopoint_parametric_dominant |
| 10 | 16,080,780.00 | rs77813541 | 2.3019 | 21_bit_twopoint_parametric_dominant |
| 10 | 16,159,902.00 | GSA-rs1890848 | 2.2435 | 21_bit_twopoint_parametric_dominant |
| 10 | 16,213,054.00 | GSA-rs1340141 | 1.8775 | 21_bit_twopoint_parametric_dominant |
| 10 | 16,232,906.00 | rs7092247 | 2.121 | 21_bit_twopoint_parametric_dominant |
| 10 | 16,255,599.00 | rs1340132 | 2.6346 | 21_bit_twopoint_parametric_dominant |
| 10 | 16,658,059.00 | rs111338993 | 2.3126 | 22_bit_twopoint_parametric_dominant |
| 10 | 17,814,445.00 | JHU_10.17856443 | 2.4887 | 21_bit_twopoint_parametric_dominant |
| 10 | 18,505,144.00 | rs10828803 | 2.4421 | 22_bit_twopoint_parametric_dominant |
| 10 | 18,546,425.00 | GSA-rs10828879 | 2.1503 | 21_bit_twopoint_parametric_dominant |
| 10 | 18,617,006.00 | GSA-rs7895709 | 2.2847 | 21_bit_twopoint_parametric_dominant |
| 10 | 19,882,841.00 | rs7087684 | 2.3916 | 21_bit_twopoint_parametric_recessive |
| 10 | 19,882,841.00 | rs7087684 | 2.3916 | 21_bit_twopoint_parametric_recessive |
| 10 | 19,882,841.00 | rs7087684 | 3.071 | 21_bit_twopoint_parametric_dominant |
| 10 | 19,882,841.00 | rs7087684 | 3.071 | 21_bit_twopoint_parametric_dominant |
| 10 | 22,047,222.00 | rs6482198 | 1.9184 | 21_bit_twopoint_parametric_dominant |
| 10 | 22,581,015.00 | rs72816826 | 2.3006 | 22_bit_twopoint_parametric_recessive |
| 10 | 25,673,004.00 | rs7897614 | 2.8622 | 21_bit_twopoint_parametric_dominant |
| 10 | 25,690,862.00 | rs7914182 | 2.3226 | 22_bit_twopoint_parametric_dominant |
| 10 | 30,751,163.00 | rs7924137 | 1.9166 | 22_bit_twopoint_parametric_dominant |
| 10 | 44,358,954.00 | JHU_10.44854401 | 1.8805 | 22_bit_twopoint_parametric_dominant |
| 10 | 48,670,042.00 | JHU_10.49878086 | 3.1683 | 22_bit_twopoint_parametric_dominant |
| 10 | 52,924,823.00 | JHU_10.54684582 | 2.2624 | 21_bit_twopoint_parametric_dominant |
| 10 | 55,254,831.00 | rs2589425 | 2.7323 | 22_bit_twopoint_parametric_dominant |
| 10 | 55,258,273.00 | rs10763190 | 2.2325 | 22_bit_twopoint_parametric_dominant |
| 10 | 65,966,756.00 | GSA-rs2105702 | 2.2928 | 21_bit_twopoint_parametric_dominant |
| 10 | 65,972,905.00 | JHU_10.67732662 | 1.9949 | 21_bit_twopoint_parametric_dominant |
| 10 | 75,636,068.00 | rs4745785 | 1.9197 | 22_bit_twopoint_parametric_recessive |
| 10 | 78,736,719.00 | rs17459521 | 2.6619 | 21_bit_twopoint_parametric_dominant |
| 10 | 78,736,719.00 | rs17459521 | 2.6619 | 21_bit_twopoint_parametric_dominant |
| 10 | 78,742,262.00 | JHU_10.80502018 | 2.535 | 21_bit_twopoint_parametric_dominant |
| 10 | 78,742,262.00 | JHU_10.80502018 | 2.535 | 21_bit_twopoint_parametric_dominant |
| 10 | 81,312,946.00 | GSA-rs653266 | 2.27 | 22_bit_twopoint_parametric_dominant |
| 10 | 93,434,954.00 | rs7091625 | 2.021 | 21_bit_twopoint_parametric_recessive |
| 10 | 93,434,954.00 | rs7091625 | 2.0669 | 21_bit_twopoint_parametric_dominant |
| 10 | 93,434,954.00 | rs7091625 | 2.1182 | 22_bit_twopoint_parametric_recessive |
| 10 | 93,962,115.00 | rs11599818 | 2.1034 | 22_bit_twopoint_parametric_dominant |
| 10 | 93,977,298.00 | rs11187749 | 2.0825 | 21_bit_twopoint_parametric_dominant |
| 10 | 93,977,298.00 | rs11187749 | 2.1278 | 22_bit_twopoint_parametric_dominant |
| 10 | 95,234,069.00 | rs11188236 | 1.932 | 22_bit_twopoint_parametric_recessive |
| 10 | 95,297,886.00 | rs7900898 | 1.9145 | 22_bit_twopoint_parametric_dominant |
| 10 | 95,359,752.00 | GSA-rs17110667 | 2.4389 | 21_bit_twopoint_parametric_dominant |
| 10 | 95,938,465.00 | GSA-rs1336457 | 2.1243 | 22_bit_twopoint_parametric_dominant |
| 10 | 100,630,970.00 | rs2489015 | 1.9027 | 21_bit_multipoint_parametric_dominant |
| 10 | 100,630,970.00 | rs2489015 | 1.9027 | 21_bit_twopoint_parametric_dominant |
| 10 | 104,278,136.00 | rs157077 | 2.2696 | 22_bit_twopoint_parametric_dominant |
| 10 | 104,522,837.00 | rs10884015 | 2.7686 | 21_bit_twopoint_parametric_dominant |
| 10 | 105,158,092.00 | GSA-rs7912305 | 2.3922 | 22_bit_twopoint_parametric_recessive |
| 10 | 105,260,573.00 | GSA-rs11192382 | 2.3834 | 22_bit_twopoint_parametric_recessive |
| 10 | 105,276,843.00 | JHU_10.107036600 | 2.3806 | 22_bit_twopoint_parametric_recessive |
| 10 | 105,276,861.00 | GSA-rs12250309 | 2.3806 | 22_bit_twopoint_parametric_recessive |
| 10 | 107,678,614.00 | rs2444215 | 1.8787 | 21_bit_twopoint_parametric_dominant |
| 10 | 107,838,085.00 | rs1946304 | 1.9382 | 21_bit_twopoint_parametric_dominant |
| 10 | 109,571,062.00 | rs11194725 | 2.5343 | 22_bit_twopoint_parametric_recessive |
| 10 | 109,571,062.00 | rs11194725 | 2.7597 | 22_bit_twopoint_parametric_dominant |
| 10 | 117,170,472.00 | rs1638438 | 2.0369 | 22_bit_twopoint_parametric_dominant |
| 10 | 118,089,633.00 | rs1932644 | 2.3021 | 21_bit_twopoint_parametric_dominant |
| 10 | 120,014,338.00 | GSA-rs6585571 | 1.9572 | 22_bit_twopoint_parametric_dominant |
| 10 | 121,052,848.00 | rs11596249 | 2.0556 | 22_bit_twopoint_parametric_recessive |
| 10 | 121,052,848.00 | rs11596249 | 2.0556 | 22_bit_twopoint_parametric_recessive |
| 10 | 121,345,609.00 | GSA-rs7903175 | 1.9981 | 22_bit_twopoint_parametric_dominant |
| 10 | 121,824,677.00 | rs7476932 | 1.9398 | 21_bit_twopoint_parametric_dominant |
| 10 | 121,876,368.00 | rs11200206 | 2.0746 | 22_bit_twopoint_parametric_dominant |
| 10 | 122,030,862.00 | rs2947603 | 2.1794 | 22_bit_twopoint_parametric_dominant |
| 10 | 122,070,326.00 | rs10788235 | 2.4427 | 22_bit_twopoint_parametric_dominant |
| 10 | 122,237,461.00 | exm861322 | 1.9482 | 22_bit_twopoint_parametric_dominant |
| 10 | 122,653,484.00 | rs9663139 | 1.881 | 21_bit_twopoint_NPL |
| 10 | 122,658,384.00 | GSA-rs2421030 | 3.242 | 21_bit_twopoint_parametric_dominant |
| 10 | 122,659,607.00 | GSA-rs7918540 | 3.242 | 21_bit_twopoint_parametric_dominant |
| 10 | 122,744,759.00 | JHU_10.124504274 | 2.1129 | 22_bit_twopoint_parametric_recessive |
| 10 | 122,744,759.00 | JHU_10.124504274 | 2.372 | 21_bit_twopoint_parametric_recessive |
| 10 | 123,466,662.00 | rs705145 | 2.0066 | 21_bit_twopoint_parametric_dominant |
| 10 | 123,502,407.00 | GSA-rs10751760 | 1.8921 | 22_bit_twopoint_parametric_recessive |
| 10 | 123,890,033.00 | rs28627761 | 2.6786 | 22_bit_twopoint_parametric_dominant |
| 10 | 123,943,586.00 | rs7071108 | 1.9099 | 22_bit_twopoint_parametric_recessive |
| 10 | 123,943,586.00 | rs7071108 | 1.9959 | 21_bit_twopoint_parametric_recessive |
| 10 | 124,043,787.00 | rs7085454 | 2.1647 | 22_bit_twopoint_parametric_dominant |
| 10 | 124,235,125.00 | rs4282943 | 2.5084 | 21_bit_twopoint_parametric_dominant |
| 10 | 124,578,221.00 | GSA-rs4962661 | 2.5337 | 21_bit_twopoint_parametric_dominant |
| 10 | 124,783,137.00 | rs113801716 | 2.6036 | 22_bit_twopoint_parametric_recessive |
| 10 | 126,425,016.00 | rs3812678 | 1.9247 | 21_bit_twopoint_parametric_recessive |
| 10 | 126,425,016.00 | rs3812678 | 2.071 | 21_bit_twopoint_parametric_dominant |
| 10 | 127,162,722.00 | rs1413592 | 2.0262 | 22_bit_twopoint_parametric_dominant |
| 10 | 127,324,769.00 | exm2267062 | 2.1387 | 21_bit_twopoint_parametric_dominant |
| 10 | 127,330,053.00 | rs9663318 | 2.8744 | 21_bit_twopoint_parametric_dominant |
| 10 | 127,330,720.00 | GSA-rs10764970 | 2.2112 | 21_bit_twopoint_parametric_dominant |
| 10 | 127,355,124.00 | rs73384660 | 2.6408 | 22_bit_twopoint_parametric_recessive |
| 10 | 127,355,161.00 | JHU_10.129153424 | 2.6408 | 22_bit_twopoint_parametric_recessive |
| 10 | 128,061,728.00 | rs4462251 | 2.7089 | 21_bit_twopoint_parametric_dominant |
| 10 | 128,066,251.00 | rs10764743 | 2.5514 | 21_bit_twopoint_parametric_dominant |
| 10 | 128,271,495.00 | JHU_10.130069758 | 2.7456 | 21_bit_twopoint_parametric_dominant |
| 10 | 129,647,467.00 | rs1008982 | 1.8851 | 21_bit_twopoint_parametric_recessive |
| 10 | 131,263,415.00 | rs10829963 | 2.117 | 21_bit_multipoint_NPL |
| 10 | 131,315,181.00 | rs74766946 | 2.1645 | 22_bit_twopoint_parametric_dominant |
| 10 | 132,260,382.00 | rs11146274 | 3.1904 | 22_bit_twopoint_parametric_dominant |
| 10 | 132,278,005.00 | rs7094372 | 2.5475 | 22_bit_twopoint_parametric_dominant |
| 10 | 132,709,769.00 | rs34459585 | 1.8869 | 21_bit_twopoint_parametric_recessive |
| 11 | 309,127.00 | 11:309127-AG | 2.112 | 22_bit_twopoint_parametric_dominant |
| 11 | 443,587.00 | rs72848748 | 1.9978 | 22_bit_twopoint_parametric_recessive |
| 11 | 463,577.00 | JHU_11.463576 | 2.0022 | 22_bit_twopoint_parametric_recessive |
| 11 | 621,634.00 | rs2740373 | 2.5534 | 21_bit_twopoint_parametric_dominant |
| 11 | 621,634.00 | rs2740373 | 2.7962 | 22_bit_twopoint_parametric_dominant |
| 11 | 2,408,387.00 | GSA-rs800343 | 2.7411 | 21_bit_twopoint_parametric_dominant |
| 11 | 2,816,395.00 | GSA-rs233446 | 1.8699 | 21_bit_twopoint_parametric_recessive |
| 11 | 2,816,395.00 | GSA-rs233446 | 2.6943 | 21_bit_twopoint_parametric_dominant |
| 11 | 2,820,135.00 | rs151293 | 2.3238 | 21_bit_twopoint_parametric_dominant |
| 11 | 3,895,276.00 | GSA-rs10835262 | 2.3768 | 22_bit_twopoint_parametric_dominant |
| 11 | 3,897,371.00 | rs4622250 | 2.7796 | 22_bit_twopoint_parametric_dominant |
| 11 | 5,364,702.00 | GSA-rs2647608 | 1.9149 | 21_bit_twopoint_parametric_dominant |
| 11 | 5,364,702.00 | GSA-rs2647608 | 1.9488 | 22_bit_twopoint_parametric_dominant |
| 11 | 5,365,438.00 | GSA-rs10500635 | 1.9149 | 21_bit_twopoint_parametric_dominant |
| 11 | 5,365,438.00 | GSA-rs10500635 | 1.9488 | 22_bit_twopoint_parametric_dominant |
| 11 | 5,423,831.00 | rs12291806 | 2.9103 | 21_bit_twopoint_parametric_dominant |
| 11 | 5,604,483.00 | rs10769118 | 2.1584 | 22_bit_twopoint_parametric_dominant |
| 11 | 7,235,892.00 | rs74053911 | 2.0945 | 22_bit_twopoint_parametric_dominant |
| 11 | 7,460,995.00 | rs7107529 | 1.9114 | 21_bit_twopoint_parametric_recessive |
| 11 | 11,210,503.00 | JHU_11.11232049 | 1.8699 | 21_bit_twopoint_parametric_dominant |
| 11 | 11,210,503.00 | JHU_11.11232049 | 2.7501 | 22_bit_twopoint_parametric_dominant |
| 11 | 11,210,713.00 | rs4910278 | 2.149 | 21_bit_twopoint_parametric_dominant |
| 11 | 11,210,713.00 | rs4910278 | 2.728 | 22_bit_twopoint_parametric_dominant |
| 11 | 11,296,401.00 | rs7114163 | 2.0863 | 22_bit_twopoint_parametric_dominant |
| 11 | 11,451,681.00 | rs12417203 | 2.6279 | 21_bit_twopoint_parametric_dominant |
| 11 | 12,640,391.00 | rs35098412 | 2.1208 | 21_bit_twopoint_parametric_recessive |
| 11 | 12,640,391.00 | rs35098412 | 2.1208 | 21_bit_twopoint_parametric_recessive |
| 11 | 12,972,565.00 | rs12295316 | 2.556 | 21_bit_twopoint_parametric_recessive |
| 11 | 12,972,565.00 | rs12295316 | 2.556 | 21_bit_twopoint_parametric_recessive |
| 11 | 12,972,565.00 | rs12295316 | 2.6939 | 22_bit_twopoint_parametric_recessive |
| 11 | 12,972,565.00 | rs12295316 | 2.6939 | 22_bit_twopoint_parametric_recessive |
| 11 | 13,913,381.00 | exm2267254 | 2.003 | 22_bit_twopoint_parametric_recessive |
| 11 | 14,102,542.00 | rs1609340 | 2.7507 | 21_bit_twopoint_parametric_recessive |
| 11 | 16,821,828.00 | JHU_11.16843374 | 1.9626 | 21_bit_twopoint_parametric_dominant |
| 11 | 16,821,828.00 | JHU_11.16843374 | 2.7425 | 21_bit_twopoint_parametric_recessive |
| 11 | 16,821,828.00 | JHU_11.16843374 | 2.8249 | 22_bit_twopoint_parametric_recessive |
| 11 | 19,396,808.00 | JHU_11.19418354 | 2.5426 | 22_bit_twopoint_parametric_recessive |
| 11 | 21,184,744.00 | JHU_11.21206289 | 2.4134 | 22_bit_twopoint_parametric_dominant |
| 11 | 21,184,744.00 | JHU_11.21206289 | 2.4134 | 22_bit_twopoint_parametric_dominant |
| 11 | 21,212,477.00 | rs11025965 | 2.7053 | 22_bit_twopoint_parametric_dominant |
| 11 | 21,212,477.00 | rs11025965 | 2.7053 | 22_bit_twopoint_parametric_dominant |
| 11 | 23,514,691.00 | rs12291079 | 2.2144 | 21_bit_twopoint_parametric_recessive |
| 11 | 25,088,602.00 | JHU_11.25110147 | 2.0078 | 21_bit_twopoint_parametric_dominant |
| 11 | 26,800,045.00 | JHU_11.26821591 | 2.1639 | 21_bit_multipoint_parametric_dominant |
| 11 | 26,800,045.00 | JHU_11.26821591 | 2.2355 | 21_bit_twopoint_parametric_dominant |
| 11 | 26,853,033.00 | GSA-rs7124231 | 1.9566 | 22_bit_twopoint_parametric_recessive |
| 11 | 27,937,273.00 | rs2726832 | 1.9494 | 22_bit_twopoint_parametric_recessive |
| 11 | 30,144,349.00 | rs930321 | 1.9438 | 21_bit_twopoint_parametric_dominant |
| 11 | 30,144,917.00 | JHU_11.30166463 | 1.9437 | 21_bit_twopoint_parametric_dominant |
| 11 | 31,567,652.00 | GSA-rs2996464 | 2.1126 | 22_bit_twopoint_parametric_recessive |
| 11 | 32,273,766.00 | rs7130589 | 2.8481 | 21_bit_twopoint_parametric_dominant |
| 11 | 32,273,766.00 | rs7130589 | 2.8481 | 21_bit_twopoint_parametric_dominant |
| 11 | 36,006,925.00 | rs1322737 | 1.8891 | 21_bit_twopoint_parametric_dominant |
| 11 | 36,126,094.00 | rs7117000 | 2.1879 | 21_bit_twopoint_parametric_dominant |
| 11 | 36,354,471.00 | rs10501149 | 1.9465 | 22_bit_twopoint_parametric_recessive |
| 11 | 36,946,187.00 | JHU_11.36967736 | 1.8822 | 21_bit_twopoint_parametric_recessive |
| 11 | 37,569,649.00 | rs7932763 | 2.1262 | 21_bit_twopoint_parametric_dominant |
| 11 | 37,644,731.00 | GSA-rs1381441 | 2.6998 | 22_bit_twopoint_parametric_dominant |
| 11 | 38,556,236.00 | GSA-rs7934284 | 1.9567 | 22_bit_twopoint_parametric_dominant |
| 11 | 38,571,292.00 | GSA-rs11034859 | 2.2145 | 22_bit_twopoint_parametric_dominant |
| 11 | 39,440,468.00 | JHU_11.39462017 | 1.8973 | 22_bit_twopoint_parametric_recessive |
| 11 | 39,440,468.00 | JHU_11.39462017 | 2.0991 | 21_bit_twopoint_parametric_recessive |
| 11 | 40,058,505.00 | GSA-rs11035665 | 2.1354 | 21_bit_twopoint_parametric_dominant |
| 11 | 43,538,441.00 | rs10400240 | 1.999 | 22_bit_multipoint_parametric_dominant |
| 11 | 43,538,441.00 | rs10400240 | 2.0217 | 22_bit_twopoint_parametric_dominant |
| 11 | 43,538,441.00 | rs10400240 | 2.2175 | 21_bit_multipoint_parametric_dominant |
| 11 | 43,538,441.00 | rs10400240 | 2.2412 | 21_bit_twopoint_parametric_dominant |
| 11 | 44,500,979.00 | rs1560293 | 1.9494 | 21_bit_twopoint_parametric_dominant |
| 11 | 44,500,979.00 | rs1560293 | 2.3554 | 22_bit_twopoint_parametric_dominant |
| 11 | 45,657,647.00 | rs895729 | 2.1185 | 21_bit_twopoint_parametric_dominant |
| 11 | 56,892,262.00 | GSA-rs1792500 | 1.9418 | 21_bit_twopoint_parametric_dominant |
| 11 | 59,863,363.00 | GSA-rs2000613 | 1.8692 | 22_bit_twopoint_parametric_recessive |
| 11 | 61,017,880.00 | rs1050922 | 2.1367 | 22_bit_twopoint_parametric_dominant |
| 11 | 61,017,880.00 | rs1050922 | 2.1367 | 22_bit_twopoint_parametric_dominant |
| 11 | 64,040,591.00 | rs320145 | 2.9024 | 22_bit_twopoint_parametric_dominant |
| 11 | 75,487,586.00 | rs61897413 | 1.9759 | 21_bit_twopoint_parametric_dominant |
| 11 | 81,431,636.00 | rs10792563 | 1.908 | 22_bit_twopoint_parametric_dominant |
| 11 | 81,431,636.00 | rs10792563 | 1.908 | 22_bit_twopoint_parametric_dominant |
| 11 | 81,431,636.00 | rs10792563 | 2.041 | 22_bit_twopoint_NPL |
| 11 | 82,961,481.00 | rs35198520 | 1.907 | 21_bit_twopoint_parametric_recessive |
| 11 | 82,961,481.00 | rs35198520 | 2.4253 | 22_bit_twopoint_parametric_recessive |
| 11 | 83,081,483.00 | GSA-rs6592098 | 2.014 | 21_bit_twopoint_NPL |
| 11 | 83,081,483.00 | GSA-rs6592098 | 2.8862 | 21_bit_twopoint_parametric_recessive |
| 11 | 95,504,759.00 | GSA-rs4237573 | 2.0901 | 21_bit_twopoint_parametric_dominant |
| 11 | 100,295,058.00 | exm2271640 | 2.6722 | 22_bit_twopoint_parametric_dominant |
| 11 | 111,431,036.00 | rs7931130 | 2.0088 | 21_bit_twopoint_parametric_dominant |
| 11 | 112,781,047.00 | JHU_11.112651769 | 1.9244 | 22_bit_twopoint_parametric_dominant |
| 11 | 114,262,697.00 | rs694539 | 2.7799 | 22_bit_twopoint_parametric_dominant |
| 11 | 114,288,721.00 | rs10400393 | 2.7757 | 22_bit_twopoint_parametric_dominant |
| 11 | 116,314,203.00 | rs10450623 | 2.3172 | 22_bit_twopoint_parametric_recessive |
| 11 | 123,508,679.00 | rs2714070 | 1.9095 | 22_bit_twopoint_parametric_dominant |
| 11 | 124,515,560.00 | GSA-rs7950818 | 2.9415 | 21_bit_twopoint_parametric_dominant |
| 11 | 124,516,247.00 | JHU_11.124386142 | 2.9415 | 21_bit_twopoint_parametric_dominant |
| 11 | 124,521,234.00 | rs75862655 | 2.9415 | 21_bit_twopoint_parametric_dominant |
| 11 | 128,774,512.00 | rs603021 | 2.3727 | 21_bit_twopoint_parametric_recessive |
| 11 | 128,774,512.00 | rs603021 | 2.4029 | 21_bit_twopoint_parametric_dominant |
| 11 | 130,870,529.00 | rs11222368 | 2.3972 | 22_bit_twopoint_parametric_recessive |
| 11 | 130,870,529.00 | rs11222368 | 3.1762 | 21_bit_twopoint_parametric_recessive |
| 11 | 131,219,459.00 | rs1540228 | 2.001 | 21_bit_twopoint_parametric_recessive |
| 11 | 131,219,459.00 | rs1540228 | 2.5974 | 22_bit_twopoint_parametric_recessive |
| 11 | 131,231,885.00 | rs12794558 | 2.5675 | 22_bit_twopoint_parametric_recessive |
| 11 | 131,237,428.00 | rs7124776 | 2.0854 | 21_bit_twopoint_parametric_recessive |
| 11 | 131,237,428.00 | rs7124776 | 2.1247 | 22_bit_twopoint_parametric_recessive |
| 11 | 131,237,428.00 | rs7124776 | 2.2646 | 22_bit_twopoint_parametric_dominant |
| 11 | 131,424,538.00 | rs4570588 | 1.9473 | 22_bit_twopoint_parametric_recessive |
| 11 | 131,455,973.00 | rs10894394 | 2.0054 | 22_bit_twopoint_parametric_recessive |
| 11 | 132,422,574.00 | rs10894558 | 2.1685 | 22_bit_twopoint_parametric_recessive |
| 11 | 132,436,509.00 | GSA-rs2298479 | 2.1685 | 22_bit_twopoint_parametric_recessive |
| 11 | 133,088,279.00 | rs66628883 | 2.0152 | 21_bit_twopoint_parametric_recessive |
| 11 | 133,564,530.00 | GSA-rs11223511 | 1.9373 | 22_bit_twopoint_parametric_dominant |
| 12 | 110,822.00 | rs2011738 | 2.0156 | 21_bit_twopoint_parametric_recessive |
| 12 | 200,587.00 | rs7967868 | 1.8624 | 21_bit_twopoint_parametric_recessive |
| 12 | 200,587.00 | rs7967868 | 2.0482 | 22_bit_twopoint_parametric_recessive |
| 12 | 200,587.00 | rs7967868 | 2.071 | 21_bit_twopoint_NPL |
| 12 | 1,546,669.00 | rs12581146 | 2.0171 | 22_bit_twopoint_parametric_dominant |
| 12 | 1,546,669.00 | rs12581146 | 2.3392 | 22_bit_twopoint_parametric_recessive |
| 12 | 3,452,878.00 | rs16930499 | 2.4384 | 21_bit_twopoint_parametric_dominant |
| 12 | 4,211,789.00 | rs10849015 | 2.1225 | 22_bit_twopoint_parametric_recessive |
| 12 | 4,938,539.00 | rs12423672 | 2.2469 | 22_bit_twopoint_parametric_recessive |
| 12 | 5,022,387.00 | rs16932667 | 1.9807 | 22_bit_twopoint_parametric_recessive |
| 12 | 5,022,387.00 | rs16932667 | 2.3102 | 22_bit_twopoint_parametric_dominant |
| 12 | 5,030,207.00 | rs9919705 | 3.2258 | 21_bit_twopoint_parametric_dominant |
| 12 | 5,031,816.00 | rs11063476 | 1.9364 | 22_bit_twopoint_parametric_dominant |
| 12 | 5,531,947.00 | rs2193160 | 2.0254 | 21_bit_twopoint_parametric_recessive |
| 12 | 5,531,947.00 | rs2193160 | 2.0452 | 22_bit_twopoint_parametric_dominant |
| 12 | 5,531,947.00 | rs2193160 | 2.0666 | 22_bit_twopoint_parametric_recessive |
| 12 | 5,987,525.00 | JHU_12.6096690 | 1.9182 | 22_bit_twopoint_parametric_recessive |
| 12 | 5,987,525.00 | JHU_12.6096690 | 1.9182 | 22_bit_twopoint_parametric_recessive |
| 12 | 6,231,222.00 | rs2268009 | 2.2786 | 22_bit_twopoint_parametric_dominant |
| 12 | 6,238,980.00 | JHU_12.6348145 | 1.868 | 21_bit_twopoint_NPL |
| 12 | 6,238,980.00 | JHU_12.6348145 | 2.216 | 22_bit_twopoint_parametric_dominant |
| 12 | 6,238,980.00 | JHU_12.6348145 | 2.24 | 22_bit_twopoint_parametric_recessive |
| 12 | 6,296,412.00 | rs6489709 | 1.8638 | 22_bit_twopoint_parametric_recessive |
| 12 | 6,296,412.00 | rs6489709 | 2.4356 | 21_bit_twopoint_parametric_dominant |
| 12 | 6,296,412.00 | rs6489709 | 2.5191 | 22_bit_twopoint_parametric_dominant |
| 12 | 6,787,028.00 | GSA-rs1922452 | 2.1796 | 22_bit_twopoint_parametric_dominant |
| 12 | 9,459,608.00 | JHU_12.9612203 | 2.1001 | 21_bit_twopoint_parametric_recessive |
| 12 | 10,118,223.00 | JHU_12.10270821 | 2.1443 | 21_bit_twopoint_parametric_recessive |
| 12 | 10,118,488.00 | exm983328 | 2.0461 | 21_bit_twopoint_parametric_recessive |
| 12 | 10,356,376.00 | rs6488284 | 1.8633 | 21_bit_twopoint_parametric_dominant |
| 12 | 10,550,245.00 | GSA-rs111782036 | 2.5879 | 21_bit_twopoint_parametric_dominant |
| 12 | 12,748,034.00 | rs7970677 | 2.1452 | 22_bit_twopoint_parametric_recessive |
| 12 | 12,748,034.00 | rs7970677 | 2.5354 | 21_bit_twopoint_parametric_recessive |
| 12 | 12,749,077.00 | rs4763870 | 2.1452 | 22_bit_twopoint_parametric_recessive |
| 12 | 12,749,077.00 | rs4763870 | 2.5354 | 21_bit_twopoint_parametric_recessive |
| 12 | 13,417,162.00 | JHU_12.13570095 | 2.0803 | 22_bit_twopoint_parametric_dominant |
| 12 | 13,417,162.00 | JHU_12.13570095 | 2.1622 | 22_bit_twopoint_parametric_recessive |
| 12 | 13,420,968.00 | rs11055415 | 1.8987 | 22_bit_twopoint_parametric_dominant |
| 12 | 13,420,968.00 | rs11055415 | 2.1551 | 22_bit_twopoint_parametric_recessive |
| 12 | 13,564,574.00 | 12:13717508-G-A | 2.1003 | 21_bit_twopoint_parametric_dominant |
| 12 | 13,835,936.00 | rs2284424 | 2.327 | 22_bit_twopoint_parametric_dominant |
| 12 | 13,835,936.00 | rs2284424 | 2.327 | 22_bit_twopoint_parametric_dominant |
| 12 | 15,820,338.00 | rs11056659 | 2.2472 | 21_bit_twopoint_parametric_dominant |
| 12 | 16,146,667.00 | rs7137096 | 2.0979 | 21_bit_twopoint_parametric_dominant |
| 12 | 20,776,059.00 | rs117826517 | 2.498 | 21_bit_twopoint_parametric_dominant |
| 12 | 22,307,485.00 | rs7295402 | 2.0819 | 22_bit_twopoint_parametric_dominant |
| 12 | 23,667,060.00 | JHU_12.23819993 | 2.257 | 22_bit_twopoint_parametric_dominant |
| 12 | 23,667,060.00 | JHU_12.23819993 | 2.257 | 22_bit_twopoint_parametric_dominant |
| 12 | 23,700,135.00 | rs725124 | 2.04 | 21_bit_twopoint_parametric_dominant |
| 12 | 23,700,135.00 | rs725124 | 2.04 | 21_bit_twopoint_parametric_dominant |
| 12 | 24,205,319.00 | rs12581837 | 2.0801 | 21_bit_twopoint_parametric_recessive |
| 12 | 25,300,180.00 | rs7295713 | 2.2606 | 21_bit_twopoint_parametric_recessive |
| 12 | 25,300,180.00 | rs7295713 | 2.6009 | 22_bit_twopoint_parametric_recessive |
| 12 | 25,300,180.00 | rs7295713 | 2.9876 | 21_bit_twopoint_parametric_dominant |
| 12 | 27,851,953.00 | rs258394 | 2.5595 | 21_bit_twopoint_parametric_dominant |
| 12 | 29,716,110.00 | rs4931213 | 1.864 | 21_bit_twopoint_parametric_dominant |
| 12 | 30,929,390.00 | rs11613187 | 1.8616 | 22_bit_twopoint_parametric_dominant |
| 12 | 30,929,390.00 | rs11613187 | 1.8616 | 22_bit_twopoint_parametric_dominant |
| 12 | 30,930,098.00 | rs74689507 | 1.8956 | 22_bit_twopoint_parametric_dominant |
| 12 | 30,930,098.00 | rs74689507 | 1.8956 | 22_bit_twopoint_parametric_dominant |
| 12 | 31,969,279.00 | rs4930999 | 2.3189 | 21_bit_twopoint_parametric_dominant |
| 12 | 31,981,704.00 | exm992988 | 2.2999 | 21_bit_twopoint_parametric_dominant |
| 12 | 32,333,125.00 | rs10844187 | 2.6787 | 22_bit_twopoint_parametric_recessive |
| 12 | 32,333,125.00 | rs10844187 | 3.0627 | 22_bit_twopoint_parametric_dominant |
| 12 | 33,382,354.00 | GSA-rs10844571 | 2.0511 | 22_bit_twopoint_parametric_recessive |
| 12 | 33,492,074.00 | rs1905240 | 2.0983 | 22_bit_twopoint_parametric_recessive |
| 12 | 34,301,366.00 | rs7314457 | 1.88 | 22_bit_twopoint_parametric_dominant |
| 12 | 39,618,495.00 | rs10877201 | 2.4262 | 22_bit_twopoint_parametric_dominant |
| 12 | 39,681,847.00 | GSA-rs10747853 | 1.8729 | 22_bit_twopoint_parametric_dominant |
| 12 | 39,724,447.00 | GSA-rs4388963 | 1.8729 | 22_bit_twopoint_parametric_dominant |
| 12 | 39,752,596.00 | rs4255600 | 1.8729 | 22_bit_twopoint_parametric_dominant |
| 12 | 40,479,449.00 | GSA-rs2588400 | 2.0789 | 22_bit_twopoint_parametric_dominant |
| 12 | 40,568,134.00 | rs7971126 | 2.0891 | 22_bit_twopoint_parametric_dominant |
| 12 | 42,663,926.00 | rs4556609 | 3.1544 | 22_bit_twopoint_parametric_recessive |
| 12 | 43,175,563.00 | GSA-rs7310014 | 1.8748 | 22_bit_twopoint_parametric_recessive |
| 12 | 43,562,591.00 | rs6582473 | 2.9725 | 21_bit_twopoint_parametric_recessive |
| 12 | 43,580,150.00 | rs7956765 | 1.9138 | 22_bit_multipoint_parametric_recessive |
| 12 | 43,580,150.00 | rs7956765 | 1.9156 | 22_bit_twopoint_parametric_recessive |
| 12 | 43,580,150.00 | rs7956765 | 2.9449 | 21_bit_multipoint_parametric_recessive |
| 12 | 43,580,150.00 | rs7956765 | 2.9495 | 21_bit_twopoint_parametric_recessive |
| 12 | 43,587,700.00 | JHU_12.43981502 | 1.9392 | 21_bit_twopoint_parametric_recessive |
| 12 | 43,780,963.00 | rs4251580 | 1.8614 | 21_bit_twopoint_parametric_recessive |
| 12 | 43,786,492.00 | exm995911 | 2.2948 | 22_bit_twopoint_parametric_dominant |
| 12 | 44,019,363.00 | exm2267432 | 2.1816 | 22_bit_twopoint_parametric_dominant |
| 12 | 51,191,552.00 | rs10783419 | 1.9878 | 22_bit_twopoint_parametric_dominant |
| 12 | 51,191,552.00 | rs10783419 | 2.221 | 21_bit_twopoint_parametric_dominant |
| 12 | 51,191,552.00 | rs10783419 | 2.6129 | 22_bit_twopoint_parametric_recessive |
| 12 | 51,191,552.00 | rs10783419 | 2.6447 | 21_bit_twopoint_parametric_recessive |
| 12 | 52,823,166.00 | GSA-rs73102423 | 1.913 | 22_bit_twopoint_parametric_recessive |
| 12 | 54,727,409.00 | rs11495473 | 2.0315 | 22_bit_twopoint_parametric_recessive |
| 12 | 58,916,359.00 | rs61921902 | 2.0054 | 22_bit_twopoint_parametric_dominant |
| 12 | 58,916,359.00 | rs61921902 | 2.0054 | 22_bit_twopoint_parametric_dominant |
| 12 | 63,311,190.00 | rs1566303 | 2.1122 | 21_bit_twopoint_parametric_dominant |
| 12 | 66,971,564.00 | exm2271747 | 2.3776 | 21_bit_twopoint_parametric_recessive |
| 12 | 67,894,127.00 | rs7959346 | 3.0549 | 21_bit_twopoint_parametric_dominant |
| 12 | 75,724,167.00 | JHU_12.76117946 | 1.936 | 21_bit_multipoint_NPL |
| 12 | 84,712,848.00 | rs61530084 | 2.2497 | 21_bit_twopoint_parametric_dominant |
| 12 | 86,053,035.00 | rs10779227 | 1.9065 | 21_bit_twopoint_parametric_dominant |
| 12 | 90,883,726.00 | exm2267380 | 1.9001 | 21_bit_twopoint_parametric_dominant |
| 12 | 90,986,856.00 | exm2260140 | 1.8615 | 21_bit_twopoint_parametric_dominant |
| 12 | 92,705,980.00 | rs1394585 | 2.1632 | 22_bit_twopoint_parametric_dominant |
| 12 | 93,051,361.00 | rs78156351 | 1.982 | 22_bit_twopoint_parametric_recessive |
| 12 | 94,646,236.00 | rs17023008 | 2.0369 | 21_bit_twopoint_parametric_recessive |
| 12 | 94,646,236.00 | rs17023008 | 2.0369 | 21_bit_twopoint_parametric_recessive |
| 12 | 104,574,876.00 | rs834723 | 1.9545 | 21_bit_twopoint_parametric_recessive |
| 12 | 108,399,400.00 | rs10861911 | 2.3542 | 22_bit_twopoint_parametric_dominant |
| 12 | 116,639,556.00 | rs66614092 | 2.0162 | 21_bit_twopoint_parametric_dominant |
| 12 | 118,576,703.00 | rs2682753 | 2.3481 | 21_bit_twopoint_parametric_dominant |
| 12 | 118,576,703.00 | rs2682753 | 2.7929 | 22_bit_twopoint_parametric_dominant |
| 12 | 121,392,140.00 | rs7961855 | 3.1678 | 22_bit_twopoint_parametric_dominant |
| 12 | 121,392,140.00 | rs7961855 | 3.1678 | 22_bit_twopoint_parametric_dominant |
| 12 | 122,690,196.00 | rs601339 | 2.1219 | 22_bit_twopoint_parametric_recessive |
| 12 | 122,695,514.00 | GSA-rs1798219 | 2.0924 | 22_bit_twopoint_parametric_recessive |
| 12 | 124,417,653.00 | GSA-rs1702339 | 3.0098 | 22_bit_twopoint_parametric_dominant |
| 12 | 124,435,245.00 | rs2660376 | 1.9132 | 22_bit_twopoint_parametric_recessive |
| 12 | 124,972,605.00 | rs12320413 | 2.2441 | 22_bit_twopoint_parametric_dominant |
| 12 | 126,226,772.00 | rs10773250 | 1.8955 | 22_bit_twopoint_parametric_dominant |
| 12 | 126,466,790.00 | rs1029783 | 2.145 | 22_bit_twopoint_parametric_dominant |
| 12 | 126,466,790.00 | rs1029783 | 2.1597 | 22_bit_multipoint_parametric_dominant |
| 12 | 128,083,027.00 | rs10773481 | 2.0602 | 21_bit_twopoint_parametric_dominant |
| 12 | 128,096,322.00 | rs9668398 | 2.0208 | 21_bit_twopoint_parametric_recessive |
| 12 | 128,101,018.00 | rs61939151 | 1.9902 | 21_bit_twopoint_parametric_recessive |
| 12 | 128,583,014.00 | JHU_12.129067558 | 2.1078 | 22_bit_twopoint_parametric_dominant |
| 12 | 128,583,014.00 | JHU_12.129067558 | 2.1078 | 22_bit_twopoint_parametric_dominant |
| 12 | 128,595,838.00 | rs11059785 | 2.273 | 22_bit_twopoint_parametric_recessive |
| 12 | 128,595,838.00 | rs11059785 | 2.273 | 22_bit_twopoint_parametric_recessive |
| 12 | 128,840,393.00 | rs10744391 | 1.9251 | 22_bit_twopoint_parametric_recessive |
| 12 | 128,840,393.00 | rs10744391 | 1.9251 | 22_bit_twopoint_parametric_recessive |
| 12 | 129,100,568.00 | rs10847765 | 1.8652 | 21_bit_twopoint_parametric_dominant |
| 12 | 129,107,989.00 | rs10773597 | 2.2405 | 21_bit_twopoint_parametric_dominant |
| 12 | 129,146,355.00 | rs7138570 | 2.2134 | 22_bit_twopoint_parametric_dominant |
| 12 | 129,454,564.00 | rs4759943 | 2.4972 | 22_bit_twopoint_parametric_dominant |
| 12 | 129,561,524.00 | rs922970 | 2.3692 | 21_bit_twopoint_parametric_dominant |
| 12 | 129,568,601.00 | rs155699 | 2.3774 | 21_bit_twopoint_parametric_recessive |
| 12 | 129,578,973.00 | rs265639 | 2.0309 | 21_bit_twopoint_parametric_recessive |
| 12 | 130,056,478.00 | rs11060677 | 2.1081 | 22_bit_twopoint_parametric_dominant |
| 12 | 130,056,478.00 | rs11060677 | 2.3882 | 22_bit_twopoint_parametric_recessive |
| 12 | 130,733,140.00 | rs7953586 | 2.9875 | 22_bit_twopoint_parametric_dominant |
| 12 | 131,422,798.00 | rs6486680 | 2.0137 | 22_bit_twopoint_parametric_dominant |
| 12 | 131,847,692.00 | rs11613757 | 2.0069 | 22_bit_twopoint_parametric_dominant |
| 12 | 132,193,544.00 | rs10902506 | 2.5065 | 21_bit_twopoint_parametric_recessive |
| 12 | 132,193,544.00 | rs10902506 | 3.0753 | 21_bit_twopoint_parametric_dominant |
| 12 | 132,207,620.00 | exm2267408 | 1.9053 | 21_bit_twopoint_parametric_recessive |
| 12 | 132,207,620.00 | exm2267408 | 2.3296 | 21_bit_twopoint_parametric_dominant |
| 12 | 132,432,615.00 | rs11610986 | 2.5754 | 22_bit_twopoint_parametric_dominant |
| 12 | 132,785,257.00 | rs10870511 | 1.9749 | 22_bit_twopoint_parametric_dominant |
| 12 | 132,801,854.00 | rs2291258 | 1.9021 | 22_bit_twopoint_parametric_dominant |
| 13 | 20,307,116.00 | rs9550629 | 2.2722 | 21_bit_twopoint_parametric_dominant |
| 13 | 20,439,856.00 | rs9552182 | 2.0785 | 21_bit_twopoint_parametric_dominant |
| 13 | 20,985,148.00 | rs649363 | 2.0547 | 21_bit_twopoint_parametric_dominant |
| 13 | 21,229,882.00 | rs9509607 | 2.6077 | 21_bit_twopoint_parametric_dominant |
| 13 | 21,743,051.00 | exm2271909 | 2.0825 | 22_bit_twopoint_parametric_dominant |
| 13 | 21,747,551.00 | rs544400 | 2.2969 | 22_bit_twopoint_parametric_dominant |
| 13 | 21,747,551.00 | rs544400 | 2.3226 | 22_bit_multipoint_parametric_dominant |
| 13 | 21,977,280.00 | rs7327237 | 2.2232 | 22_bit_twopoint_parametric_dominant |
| 13 | 21,977,280.00 | rs7327237 | 2.6971 | 21_bit_twopoint_parametric_dominant |
| 13 | 23,109,739.00 | rs885301 | 1.9823 | 22_bit_twopoint_parametric_dominant |
| 13 | 23,109,739.00 | rs885301 | 2.0036 | 21_bit_twopoint_parametric_dominant |
| 13 | 23,214,773.00 | rs7319164 | 2.8543 | 22_bit_twopoint_parametric_dominant |
| 13 | 23,417,237.00 | GSA-rs10747165 | 2.1165 | 21_bit_twopoint_parametric_dominant |
| 13 | 25,032,281.00 | JHU_13.25606418 | 2.0427 | 21_bit_twopoint_parametric_dominant |
| 13 | 27,517,320.00 | rs74960394 | 1.8872 | 22_bit_twopoint_parametric_dominant |
| 13 | 27,634,286.00 | rs616662 | 1.983 | 21_bit_twopoint_parametric_dominant |
| 13 | 30,225,133.00 | rs605095 | 2.0667 | 22_bit_twopoint_parametric_dominant |
| 13 | 33,194,210.00 | GSA-rs797199 | 2.3612 | 22_bit_twopoint_parametric_dominant |
| 13 | 36,239,385.00 | rs1170932 | 1.8613 | 21_bit_twopoint_parametric_recessive |
| 13 | 36,570,711.00 | rs9547615 | 1.8611 | 21_bit_twopoint_parametric_recessive |
| 13 | 36,875,096.00 | rs2762137 | 2.3793 | 22_bit_multipoint_parametric_dominant |
| 13 | 36,875,096.00 | rs2762137 | 2.4006 | 22_bit_twopoint_parametric_dominant |
| 13 | 37,194,877.00 | rs9532012 | 2.1659 | 22_bit_twopoint_parametric_dominant |
| 13 | 38,184,046.00 | rs612986 | 2.2169 | 21_bit_twopoint_parametric_recessive |
| 13 | 38,184,046.00 | rs612986 | 2.4356 | 22_bit_twopoint_parametric_recessive |
| 13 | 39,890,802.00 | rs17238355 | 2.04 | 21_bit_twopoint_parametric_recessive |
| 13 | 40,074,447.00 | rs12864585 | 1.8623 | 21_bit_twopoint_parametric_dominant |
| 13 | 40,074,447.00 | rs12864585 | 2.6264 | 22_bit_twopoint_parametric_dominant |
| 13 | 40,292,060.00 | rs11616421 | 2.1129 | 22_bit_twopoint_parametric_dominant |
| 13 | 40,293,411.00 | rs9549153 | 1.8869 | 21_bit_twopoint_parametric_recessive |
| 13 | 40,293,411.00 | rs9549153 | 1.9565 | 22_bit_twopoint_parametric_dominant |
| 13 | 41,548,489.00 | rs447270 | 1.898 | 22_bit_twopoint_parametric_recessive |
| 13 | 41,696,169.00 | rs9634767 | 1.9103 | 22_bit_twopoint_parametric_recessive |
| 13 | 42,933,898.00 | rs12868745 | 2.2096 | 21_bit_twopoint_parametric_recessive |
| 13 | 42,933,898.00 | rs12868745 | 2.2096 | 21_bit_twopoint_parametric_recessive |
| 13 | 42,933,898.00 | rs12868745 | 2.4379 | 21_bit_twopoint_parametric_dominant |
| 13 | 42,933,898.00 | rs12868745 | 2.4379 | 21_bit_twopoint_parametric_dominant |
| 13 | 43,649,762.00 | rs9525816 | 2.1312 | 22_bit_twopoint_parametric_dominant |
| 13 | 45,786,543.00 | rs7997707 | 2.1415 | 22_bit_twopoint_parametric_dominant |
| 13 | 47,288,752.00 | rs6561366 | 2.2788 | 21_bit_twopoint_parametric_recessive |
| 13 | 47,288,752.00 | rs6561366 | 2.9531 | 22_bit_twopoint_parametric_recessive |
| 13 | 47,563,408.00 | rs9670039 | 2.6174 | 22_bit_twopoint_parametric_recessive |
| 13 | 47,592,132.00 | rs1326468 | 2.362 | 22_bit_twopoint_parametric_recessive |
| 13 | 48,622,381.00 | rs7985015 | 2.0425 | 22_bit_twopoint_parametric_dominant |
| 13 | 48,622,381.00 | rs7985015 | 2.0686 | 22_bit_twopoint_parametric_recessive |
| 13 | 48,623,947.00 | rs80279557 | 2.7423 | 22_bit_twopoint_parametric_dominant |
| 13 | 48,623,947.00 | rs80279557 | 3.1283 | 22_bit_twopoint_parametric_recessive |
| 13 | 48,632,611.00 | JHU_13.49206746 | 2.7423 | 22_bit_twopoint_parametric_dominant |
| 13 | 48,632,611.00 | JHU_13.49206746 | 3.1283 | 22_bit_twopoint_parametric_recessive |
| 13 | 48,709,659.00 | rs2407249 | 1.9626 | 22_bit_twopoint_parametric_dominant |
| 13 | 48,709,659.00 | rs2407249 | 2.9976 | 22_bit_twopoint_parametric_recessive |
| 13 | 49,836,367.00 | rs4942875 | 2.3078 | 22_bit_twopoint_parametric_dominant |
| 13 | 51,107,878.00 | exm2260216 | 1.871 | 21_bit_twopoint_parametric_dominant |
| 13 | 52,987,433.00 | GSA-rs9596742 | 2.0556 | 22_bit_twopoint_parametric_recessive |
| 13 | 52,987,433.00 | GSA-rs9596742 | 2.0556 | 22_bit_twopoint_parametric_recessive |
| 13 | 53,077,540.00 | GSA-rs75610212 | 2.5306 | 22_bit_twopoint_parametric_recessive |
| 13 | 60,650,236.00 | rs7338215 | 2.2127 | 21_bit_twopoint_parametric_dominant |
| 13 | 60,650,236.00 | rs7338215 | 2.2206 | 22_bit_twopoint_parametric_dominant |
| 13 | 60,663,474.00 | GSA-rs4556691 | 2.0221 | 22_bit_twopoint_parametric_dominant |
| 13 | 60,663,474.00 | GSA-rs4556691 | 2.696 | 21_bit_twopoint_parametric_dominant |
| 13 | 60,673,071.00 | rs34500959 | 2.0221 | 22_bit_twopoint_parametric_dominant |
| 13 | 60,673,071.00 | rs34500959 | 2.696 | 21_bit_twopoint_parametric_dominant |
| 13 | 62,298,832.00 | rs4146191 | 2.2522 | 22_bit_twopoint_parametric_dominant |
| 13 | 62,361,771.00 | rs9598441 | 2.1575 | 21_bit_twopoint_parametric_dominant |
| 13 | 62,680,306.00 | GSA-rs75140026 | 1.8987 | 22_bit_twopoint_parametric_recessive |
| 13 | 63,629,611.00 | rs9570958 | 2.262 | 22_bit_twopoint_parametric_dominant |
| 13 | 63,629,611.00 | rs9570958 | 2.3162 | 21_bit_twopoint_parametric_dominant |
| 13 | 63,631,552.00 | rs1373303 | 2.2372 | 21_bit_twopoint_parametric_dominant |
| 13 | 63,631,552.00 | rs1373303 | 2.2518 | 22_bit_twopoint_parametric_dominant |
| 13 | 64,334,710.00 | rs4883735 | 2.1821 | 21_bit_twopoint_parametric_recessive |
| 13 | 64,334,710.00 | rs4883735 | 2.91 | 22_bit_twopoint_parametric_recessive |
| 13 | 64,362,111.00 | GSA-rs9571178 | 2.5986 | 21_bit_twopoint_parametric_recessive |
| 13 | 64,362,111.00 | GSA-rs9571178 | 3.2995 | 22_bit_twopoint_parametric_recessive |
| 13 | 64,851,792.00 | GSA-rs34251537 | 2.0525 | 21_bit_twopoint_parametric_recessive |
| 13 | 64,895,655.00 | rs1949203 | 2.2036 | 21_bit_twopoint_parametric_recessive |
| 13 | 65,841,224.00 | rs1460654 | 2.3774 | 22_bit_twopoint_parametric_recessive |
| 13 | 65,841,224.00 | rs1460654 | 2.3774 | 22_bit_twopoint_parametric_recessive |
| 13 | 66,443,150.00 | rs8001417 | 2.2022 | 21_bit_twopoint_parametric_recessive |
| 13 | 66,443,150.00 | rs8001417 | 2.2022 | 21_bit_twopoint_parametric_recessive |
| 13 | 66,443,150.00 | rs8001417 | 2.3451 | 22_bit_twopoint_parametric_recessive |
| 13 | 66,443,150.00 | rs8001417 | 2.3451 | 22_bit_twopoint_parametric_recessive |
| 13 | 66,447,963.00 | GSA-rs3904582 | 1.9158 | 21_bit_twopoint_parametric_dominant |
| 13 | 66,447,963.00 | GSA-rs3904582 | 1.9158 | 21_bit_twopoint_parametric_dominant |
| 13 | 66,447,963.00 | GSA-rs3904582 | 2.7245 | 22_bit_twopoint_parametric_recessive |
| 13 | 66,447,963.00 | GSA-rs3904582 | 2.7245 | 22_bit_twopoint_parametric_recessive |
| 13 | 66,447,963.00 | GSA-rs3904582 | 2.8497 | 21_bit_twopoint_parametric_recessive |
| 13 | 66,447,963.00 | GSA-rs3904582 | 2.8497 | 21_bit_twopoint_parametric_recessive |
| 13 | 67,366,186.00 | rs1123862 | 1.903 | 22_bit_twopoint_parametric_recessive |
| 13 | 68,103,705.00 | rs7320823 | 1.8707 | 22_bit_twopoint_parametric_recessive |
| 13 | 68,999,619.00 | rs1954147 | 1.9343 | 21_bit_twopoint_parametric_dominant |
| 13 | 70,307,533.00 | GSA-rs9599608 | 2.0852 | 21_bit_twopoint_parametric_dominant |
| 13 | 70,342,154.00 | rs9592702 | 2.0905 | 21_bit_twopoint_parametric_recessive |
| 13 | 70,568,082.00 | rs9542404 | 2.049 | 22_bit_twopoint_parametric_recessive |
| 13 | 71,293,401.00 | rs11838796 | 1.9113 | 22_bit_twopoint_parametric_recessive |
| 13 | 73,004,718.00 | rs9600057 | 2.26 | 21_bit_twopoint_parametric_dominant |
| 13 | 74,168,157.00 | 13:74742294-C-A | 1.8648 | 21_bit_twopoint_parametric_dominant |
| 13 | 74,193,635.00 | 13:74767772-C-T | 2.0199 | 21_bit_twopoint_parametric_dominant |
| 13 | 74,690,435.00 | GSA-rs7334899 | 2.392 | 21_bit_twopoint_parametric_dominant |
| 13 | 74,690,435.00 | GSA-rs7334899 | 2.392 | 21_bit_twopoint_parametric_dominant |
| 13 | 74,696,206.00 | JHU_13.75270342 | 2.8981 | 21_bit_twopoint_parametric_dominant |
| 13 | 74,696,206.00 | JHU_13.75270342 | 2.8981 | 21_bit_twopoint_parametric_dominant |
| 13 | 74,714,407.00 | rs2328906 | 2.3147 | 21_bit_twopoint_parametric_dominant |
| 13 | 74,714,407.00 | rs2328906 | 2.3147 | 21_bit_twopoint_parametric_dominant |
| 13 | 76,809,661.00 | rs9530599 | 1.9542 | 21_bit_twopoint_parametric_dominant |
| 13 | 81,489,657.00 | rs6563216 | 2.4551 | 22_bit_twopoint_parametric_dominant |
| 13 | 81,513,970.00 | rs9318781 | 2.1918 | 21_bit_twopoint_parametric_dominant |
| 13 | 81,517,128.00 | GSA-rs1819529 | 1.9845 | 21_bit_twopoint_parametric_dominant |
| 13 | 89,905,578.00 | rs1341651 | 1.86 | 21_bit_multipoint_NPL |
| 13 | 93,104,866.00 | JHU_13.93757118 | 2.8171 | 21_bit_twopoint_parametric_recessive |
| 13 | 93,557,435.00 | rs3848059 | 1.8945 | 21_bit_twopoint_parametric_recessive |
| 13 | 95,358,827.00 | rs13313300 | 2.4294 | 21_bit_twopoint_parametric_dominant |
| 13 | 101,310,600.00 | rs518657 | 1.8655 | 21_bit_twopoint_parametric_dominant |
| 13 | 101,770,118.00 | rs9557735 | 2.0334 | 21_bit_twopoint_parametric_dominant |
| 13 | 101,862,411.00 | rs17625232 | 1.962 | 21_bit_twopoint_parametric_dominant |
| 13 | 101,948,233.00 | rs11069468 | 2.1537 | 22_bit_twopoint_parametric_dominant |
| 13 | 102,455,075.00 | rs7328480 | 1.9729 | 22_bit_twopoint_parametric_dominant |
| 13 | 102,455,075.00 | rs7328480 | 2.2391 | 22_bit_twopoint_parametric_recessive |
| 13 | 103,252,741.00 | rs4132332 | 1.9446 | 22_bit_twopoint_parametric_recessive |
| 13 | 107,096,315.00 | rs746950 | 2.2125 | 22_bit_twopoint_parametric_dominant |
| 13 | 107,429,364.00 | exm2271991 | 2.2187 | 22_bit_twopoint_parametric_dominant |
| 13 | 109,164,340.00 | rs9301354 | 2.5488 | 21_bit_twopoint_parametric_recessive |
| 13 | 109,203,973.00 | rs927606 | 2.1311 | 21_bit_twopoint_parametric_dominant |
| 13 | 114,107,730.00 | rs9314891 | 2.1281 | 21_bit_twopoint_parametric_dominant |
| 13 | 114,107,730.00 | rs9314891 | 2.5836 | 22_bit_twopoint_parametric_dominant |
| 14 | 22,313,945.00 | exm2260295 | 1.8776 | 21_bit_twopoint_parametric_recessive |
| 14 | 22,313,945.00 | exm2260295 | 1.8776 | 21_bit_twopoint_parametric_recessive |
| 14 | 22,313,945.00 | exm2260295 | 1.8998 | 21_bit_twopoint_parametric_dominant |
| 14 | 22,313,945.00 | exm2260295 | 1.8998 | 21_bit_twopoint_parametric_dominant |
| 14 | 22,796,339.00 | rs8008926 | 2.0266 | 22_bit_twopoint_parametric_dominant |
| 14 | 24,361,644.00 | exm-rs1950500 | 2.0182 | 22_bit_twopoint_parametric_recessive |
| 14 | 35,506,135.00 | rs9322955 | 1.9456 | 21_bit_twopoint_parametric_dominant |
| 14 | 35,506,135.00 | rs9322955 | 1.9456 | 21_bit_twopoint_parametric_dominant |
| 14 | 35,506,135.00 | rs9322955 | 1.9456 | 21_bit_twopoint_parametric_dominant |
| 14 | 35,506,135.00 | rs9322955 | 1.9456 | 21_bit_twopoint_parametric_dominant |
| 14 | 47,267,091.00 | rs2148509 | 1.885 | 22_bit_twopoint_parametric_recessive |
| 14 | 48,990,971.00 | rs2352918 | 1.9328 | 22_bit_twopoint_parametric_dominant |
| 14 | 48,994,841.00 | GSA-rs73260922 | 1.9328 | 22_bit_twopoint_parametric_dominant |
| 14 | 50,853,532.00 | rs7159869 | 2.1397 | 22_bit_twopoint_parametric_dominant |
| 14 | 51,449,542.00 | rs2356914 | 1.8711 | 21_bit_twopoint_parametric_recessive |
| 14 | 51,449,542.00 | rs2356914 | 1.8711 | 21_bit_twopoint_parametric_recessive |
| 14 | 51,449,542.00 | rs2356914 | 2.0961 | 21_bit_twopoint_parametric_dominant |
| 14 | 51,449,542.00 | rs2356914 | 2.0961 | 21_bit_twopoint_parametric_dominant |
| 14 | 52,225,867.00 | rs1557191 | 2.0098 | 22_bit_twopoint_parametric_dominant |
| 14 | 52,225,867.00 | rs1557191 | 2.9213 | 22_bit_twopoint_parametric_recessive |
| 14 | 52,225,867.00 | rs1557191 | 3.1107 | 21_bit_twopoint_parametric_recessive |
| 14 | 52,225,867.00 | rs1557191 | 3.1107 | 21_bit_twopoint_parametric_recessive |
| 14 | 56,950,709.00 | rs709993 | 2.022 | 21_bit_twopoint_parametric_dominant |
| 14 | 56,950,709.00 | rs709993 | 2.022 | 21_bit_twopoint_parametric_dominant |
| 14 | 56,950,709.00 | rs709993 | 2.022 | 21_bit_twopoint_parametric_dominant |
| 14 | 56,950,709.00 | rs709993 | 2.022 | 21_bit_twopoint_parametric_dominant |
| 14 | 56,987,314.00 | rs73290435 | 2.1391 | 21_bit_twopoint_parametric_dominant |
| 14 | 56,987,314.00 | rs73290435 | 2.1391 | 21_bit_twopoint_parametric_dominant |
| 14 | 56,987,314.00 | rs73290435 | 2.1391 | 21_bit_twopoint_parametric_dominant |
| 14 | 56,987,314.00 | rs73290435 | 2.1391 | 21_bit_twopoint_parametric_dominant |
| 14 | 58,782,576.00 | GSA-rs856221 | 2.0013 | 22_bit_twopoint_parametric_dominant |
| 14 | 62,518,248.00 | rs2000250 | 2.0346 | 22_bit_twopoint_parametric_dominant |
| 14 | 62,518,248.00 | rs2000250 | 2.0699 | 22_bit_multipoint_parametric_dominant |
| 14 | 63,174,699.00 | rs7152965 | 2.0935 | 21_bit_multipoint_parametric_dominant |
| 14 | 63,174,699.00 | rs7152965 | 2.1021 | 21_bit_twopoint_parametric_dominant |
| 14 | 63,174,699.00 | rs7152965 | 2.1021 | 21_bit_twopoint_parametric_dominant |
| 14 | 64,912,429.00 | 14:65379147-T-C | 2.3357 | 21_bit_twopoint_parametric_dominant |
| 14 | 64,912,429.00 | 14:65379147-T-C | 2.3357 | 21_bit_twopoint_parametric_dominant |
| 14 | 64,912,429.00 | 14:65379147-T-C | 2.768 | 22_bit_twopoint_parametric_dominant |
| 14 | 70,872,905.00 | rs10148840 | 2.163 | 22_bit_twopoint_parametric_dominant |
| 14 | 71,606,705.00 | JHU_14.72073421 | 1.8967 | 21_bit_twopoint_parametric_recessive |
| 14 | 71,606,705.00 | JHU_14.72073421 | 1.8967 | 21_bit_twopoint_parametric_recessive |
| 14 | 76,116,515.00 | JHU_14.76582857 | 1.9192 | 21_bit_twopoint_parametric_dominant |
| 14 | 76,116,515.00 | JHU_14.76582857 | 1.9192 | 21_bit_twopoint_parametric_dominant |
| 14 | 76,116,515.00 | JHU_14.76582857 | 2.1984 | 22_bit_twopoint_parametric_dominant |
| 14 | 76,175,361.00 | JHU_14.76641703 | 1.9899 | 21_bit_twopoint_parametric_recessive |
| 14 | 76,175,361.00 | JHU_14.76641703 | 1.9899 | 21_bit_twopoint_parametric_recessive |
| 14 | 77,530,316.00 | GSA-rs112381399 | 1.9914 | 22_bit_twopoint_parametric_dominant |
| 14 | 79,229,925.00 | rs4363787 | 2.0203 | 21_bit_twopoint_parametric_recessive |
| 14 | 79,229,925.00 | rs4363787 | 2.0203 | 21_bit_twopoint_parametric_recessive |
| 14 | 83,757,040.00 | GSA-rs7157077 | 1.884 | 21_bit_twopoint_parametric_dominant |
| 14 | 83,757,040.00 | GSA-rs7157077 | 1.884 | 21_bit_twopoint_parametric_dominant |
| 14 | 86,463,582.00 | rs2022771 | 2.2505 | 21_bit_twopoint_parametric_dominant |
| 14 | 86,463,582.00 | rs2022771 | 2.2505 | 21_bit_twopoint_parametric_dominant |
| 14 | 87,814,065.00 | rs10148173 | 2.4495 | 22_bit_twopoint_parametric_dominant |
| 14 | 87,814,065.00 | rs10148173 | 2.4495 | 22_bit_twopoint_parametric_dominant |
| 14 | 88,165,977.00 | exm2272129 | 1.9443 | 21_bit_twopoint_parametric_dominant |
| 14 | 88,165,977.00 | exm2272129 | 1.9443 | 21_bit_twopoint_parametric_dominant |
| 14 | 88,165,977.00 | exm2272129 | 1.9443 | 21_bit_twopoint_parametric_dominant |
| 14 | 88,165,977.00 | exm2272129 | 1.9443 | 21_bit_twopoint_parametric_dominant |
| 14 | 89,629,101.00 | rs100016 | 2.2686 | 21_bit_twopoint_parametric_dominant |
| 14 | 89,629,101.00 | rs100016 | 2.2686 | 21_bit_twopoint_parametric_dominant |
| 14 | 89,671,082.00 | rs8006303 | 1.9216 | 22_bit_twopoint_parametric_dominant |
| 14 | 89,672,967.00 | rs998564 | 2.9088 | 21_bit_twopoint_parametric_dominant |
| 14 | 89,672,967.00 | rs998564 | 2.9088 | 21_bit_twopoint_parametric_dominant |
| 14 | 89,672,967.00 | rs998564 | 3.2049 | 22_bit_twopoint_parametric_dominant |
| 14 | 91,166,817.00 | rs12590057 | 2.059 | 22_bit_twopoint_parametric_dominant |
| 14 | 91,660,483.00 | GSA-rs2756166 | 1.8924 | 21_bit_twopoint_parametric_dominant |
| 14 | 91,660,483.00 | GSA-rs2756166 | 1.8924 | 21_bit_twopoint_parametric_dominant |
| 14 | 92,410,493.00 | rs4904900 | 1.9037 | 22_bit_twopoint_parametric_dominant |
| 14 | 93,866,491.00 | rs8017732 | 2.6188 | 21_bit_twopoint_parametric_dominant |
| 14 | 93,866,491.00 | rs8017732 | 2.6188 | 21_bit_twopoint_parametric_dominant |
| 14 | 94,290,413.00 | exm1124090 | 1.9219 | 21_bit_twopoint_parametric_recessive |
| 14 | 94,290,413.00 | exm1124090 | 1.9219 | 21_bit_twopoint_parametric_recessive |
| 14 | 94,290,413.00 | exm1124090 | 2.0273 | 21_bit_twopoint_parametric_dominant |
| 14 | 94,290,413.00 | exm1124090 | 2.0273 | 21_bit_twopoint_parametric_dominant |
| 14 | 94,396,220.00 | rs28694574 | 2.0721 | 22_bit_twopoint_parametric_recessive |
| 14 | 95,024,715.00 | rs80152868 | 2.4481 | 22_bit_twopoint_parametric_recessive |
| 14 | 95,024,715.00 | rs80152868 | 2.4481 | 22_bit_twopoint_parametric_recessive |
| 14 | 97,763,150.00 | rs4900375 | 1.8674 | 21_bit_twopoint_parametric_recessive |
| 14 | 97,763,150.00 | rs4900375 | 1.8674 | 21_bit_twopoint_parametric_recessive |
| 14 | 97,763,150.00 | rs4900375 | 1.9274 | 22_bit_twopoint_parametric_recessive |
| 14 | 97,783,065.00 | rs1462266 | 1.9884 | 21_bit_twopoint_parametric_recessive |
| 14 | 97,783,065.00 | rs1462266 | 1.9884 | 21_bit_twopoint_parametric_recessive |
| 14 | 97,783,065.00 | rs1462266 | 2.0601 | 22_bit_twopoint_parametric_recessive |
| 14 | 97,795,732.00 | rs1973150 | 2.8334 | 21_bit_twopoint_parametric_dominant |
| 14 | 97,795,732.00 | rs1973150 | 2.8334 | 21_bit_twopoint_parametric_dominant |
| 14 | 99,133,644.00 | JHU_14.99599980 | 2.4856 | 22_bit_twopoint_parametric_dominant |
| 14 | 99,564,805.00 | rs7159751 | 2.1294 | 22_bit_twopoint_parametric_recessive |
| 14 | 99,564,805.00 | rs7159751 | 2.2296 | 22_bit_twopoint_parametric_dominant |
| 14 | 101,183,096.00 | JHU_14.101649432 | 1.9703 | 22_bit_twopoint_parametric_dominant |
| 14 | 101,459,939.00 | rs34227869 | 2.1532 | 22_bit_twopoint_parametric_dominant |
| 14 | 101,687,061.00 | rs7155050 | 2.4003 | 22_bit_twopoint_parametric_recessive |
| 14 | 101,687,061.00 | rs7155050 | 2.8737 | 21_bit_twopoint_parametric_recessive |
| 14 | 101,687,061.00 | rs7155050 | 2.8737 | 21_bit_twopoint_parametric_recessive |
| 14 | 101,881,696.00 | rs1677991 | 1.8915 | 22_bit_twopoint_parametric_recessive |
| 15 | 22,945,901.00 | rs11634023 | 1.9104 | 21_bit_multipoint_parametric_dominant |
| 15 | 22,945,901.00 | rs11634023 | 1.9363 | 21_bit_twopoint_parametric_dominant |
| 15 | 23,485,976.00 | GSA-rs1526175 | 1.864 | 22_bit_twopoint_parametric_recessive |
| 15 | 23,519,300.00 | rs7174702 | 1.9447 | 22_bit_twopoint_parametric_recessive |
| 15 | 23,923,803.00 | rs9783733 | 2.1963 | 22_bit_twopoint_parametric_recessive |
| 15 | 24,212,440.00 | rs4778426 | 2.4378 | 22_bit_twopoint_parametric_dominant |
| 15 | 24,591,454.00 | GSA-rs7402866 | 2.2248 | 22_bit_twopoint_parametric_recessive |
| 15 | 24,829,620.00 | rs12905653 | 2.2741 | 21_bit_twopoint_parametric_dominant |
| 15 | 24,859,922.00 | rs7359196 | 2.2338 | 21_bit_twopoint_parametric_dominant |
| 15 | 24,859,922.00 | rs7359196 | 2.8863 | 21_bit_twopoint_parametric_recessive |
| 15 | 24,925,023.00 | rs12592731 | 1.9404 | 21_bit_twopoint_parametric_dominant |
| 15 | 25,125,517.00 | rs2356290 | 2.2936 | 22_bit_twopoint_parametric_recessive |
| 15 | 25,738,782.00 | rs4906631 | 2.1062 | 21_bit_twopoint_parametric_recessive |
| 15 | 25,869,511.00 | rs1553893 | 1.9212 | 22_bit_twopoint_parametric_recessive |
| 15 | 27,723,352.00 | rs8031442 | 1.9319 | 22_bit_twopoint_parametric_dominant |
| 15 | 27,953,421.00 | rs921221 | 1.8676 | 21_bit_twopoint_parametric_dominant |
| 15 | 29,941,315.00 | rs62014517 | 2.1666 | 21_bit_twopoint_parametric_dominant |
| 15 | 29,941,315.00 | rs62014517 | 2.7315 | 21_bit_twopoint_parametric_recessive |
| 15 | 33,426,743.00 | rs2596227 | 1.9904 | 22_bit_twopoint_parametric_dominant |
| 15 | 35,256,267.00 | GSA-rs2622756 | 1.9865 | 22_bit_twopoint_parametric_dominant |
| 15 | 35,259,173.00 | rs1867523 | 1.9618 | 22_bit_twopoint_parametric_dominant |
| 15 | 37,568,492.00 | rs12593354 | 2.0259 | 21_bit_twopoint_parametric_dominant |
| 15 | 38,015,558.00 | rs8027027 | 2.0347 | 22_bit_multipoint_parametric_recessive |
| 15 | 38,015,558.00 | rs8027027 | 2.039 | 22_bit_twopoint_parametric_recessive |
| 15 | 39,998,280.00 | JHU_15.40290480 | 2.4124 | 21_bit_twopoint_parametric_dominant |
| 15 | 47,766,287.00 | JHU_15.48058483 | 2.3341 | 22_bit_twopoint_parametric_recessive |
| 15 | 47,778,954.00 | rs512255 | 1.934 | 22_bit_twopoint_parametric_recessive |
| 15 | 47,836,699.00 | rs8029881 | 2.249 | 22_bit_twopoint_parametric_recessive |
| 15 | 47,836,699.00 | rs8029881 | 2.3139 | 22_bit_twopoint_parametric_dominant |
| 15 | 50,206,855.00 | rs7161822 | 2.1332 | 21_bit_twopoint_parametric_dominant |
| 15 | 51,358,154.00 | GSA-rs2470184 | 1.8901 | 22_bit_twopoint_parametric_dominant |
| 15 | 54,095,122.00 | rs12594549 | 2.7579 | 21_bit_twopoint_parametric_dominant |
| 15 | 54,095,122.00 | rs12594549 | 2.7579 | 21_bit_twopoint_parametric_dominant |
| 15 | 54,344,334.00 | rs8030739 | 2.0647 | 22_bit_twopoint_parametric_dominant |
| 15 | 54,344,334.00 | rs8030739 | 2.0647 | 22_bit_twopoint_parametric_dominant |
| 15 | 56,637,026.00 | rs72744943 | 2.1268 | 22_bit_twopoint_parametric_recessive |
| 15 | 58,322,330.00 | rs1663242 | 2.2905 | 22_bit_twopoint_parametric_recessive |
| 15 | 58,322,330.00 | rs1663242 | 2.352 | 21_bit_twopoint_parametric_recessive |
| 15 | 61,022,397.00 | GSA-rs4774381 | 2.0541 | 21_bit_twopoint_parametric_recessive |
| 15 | 61,803,374.00 | rs4775435 | 1.9091 | 22_bit_twopoint_parametric_dominant |
| 15 | 76,348,089.00 | rs2460161 | 2.6013 | 22_bit_twopoint_parametric_recessive |
| 15 | 76,451,961.00 | rs284904 | 2.6013 | 22_bit_twopoint_parametric_recessive |
| 15 | 76,479,721.00 | rs2469249 | 2.2575 | 22_bit_twopoint_parametric_dominant |
| 15 | 77,454,848.00 | rs7178572 | 1.9219 | 21_bit_twopoint_parametric_recessive |
| 15 | 78,836,157.00 | GSA-rs4539564 | 2.401 | 21_bit_twopoint_parametric_dominant |
| 15 | 78,892,045.00 | rs9806693 | 1.9604 | 22_bit_twopoint_parametric_dominant |
| 15 | 79,241,030.00 | rs4779031 | 1.9374 | 22_bit_twopoint_parametric_dominant |
| 15 | 79,814,193.00 | rs2035027 | 2.3198 | 22_bit_twopoint_parametric_recessive |
| 15 | 79,814,193.00 | rs2035027 | 2.7496 | 22_bit_twopoint_parametric_dominant |
| 15 | 79,865,241.00 | rs16971460 | 2.0132 | 22_bit_twopoint_parametric_dominant |
| 15 | 95,457,300.00 | rs7165561 | 1.9337 | 22_bit_twopoint_parametric_dominant |
| 15 | 97,121,424.00 | rs7180063 | 2.0585 | 21_bit_twopoint_parametric_dominant |
| 15 | 97,121,424.00 | rs7180063 | 2.8728 | 21_bit_twopoint_parametric_recessive |
| 15 | 97,121,424.00 | rs7180063 | 2.9777 | 22_bit_twopoint_parametric_recessive |
| 15 | 97,225,192.00 | exm2267854 | 2.1896 | 21_bit_twopoint_parametric_recessive |
| 15 | 97,710,064.00 | rs11633855 | 2.4335 | 22_bit_twopoint_parametric_recessive |
| 15 | 97,710,064.00 | rs11633855 | 2.4335 | 22_bit_twopoint_parametric_recessive |
| 15 | 97,711,897.00 | GSA-rs520140 | 2.6215 | 22_bit_twopoint_parametric_dominant |
| 15 | 97,711,897.00 | GSA-rs520140 | 2.6215 | 22_bit_twopoint_parametric_dominant |
| 15 | 98,956,125.00 | rs41508349 | 2.0143 | 22_bit_twopoint_parametric_dominant |
| 15 | 101,178,722.00 | exm1194215 | 1.9319 | 22_bit_twopoint_parametric_dominant |
| 15 | 101,301,841.00 | rs4965376 | 2.0899 | 22_bit_twopoint_parametric_dominant |
| 16 | 615,323.00 | rs11643412 | 1.9005 | 21_bit_twopoint_parametric_dominant |
| 16 | 1,299,928.00 | rs4984803 | 2.0227 | 21_bit_twopoint_parametric_recessive |
| 16 | 2,061,778.00 | rs17135764 | 2.9861 | 22_bit_twopoint_parametric_dominant |
| 16 | 3,925,155.00 | rs8044196 | 2.0203 | 21_bit_twopoint_parametric_dominant |
| 16 | 5,055,364.00 | rs741167 | 1.944 | 21_bit_twopoint_NPL |
| 16 | 5,055,364.00 | rs741167 | 2.1498 | 22_bit_twopoint_parametric_recessive |
| 16 | 5,055,364.00 | rs741167 | 2.222 | 21_bit_twopoint_parametric_dominant |
| 16 | 5,055,364.00 | rs741167 | 2.6101 | 21_bit_twopoint_parametric_recessive |
| 16 | 5,844,781.00 | rs10500329 | 2.3051 | 22_bit_twopoint_parametric_dominant |
| 16 | 5,993,105.00 | rs9929481 | 1.9653 | 22_bit_twopoint_parametric_dominant |
| 16 | 7,642,184.00 | rs2079753 | 1.9901 | 22_bit_twopoint_parametric_dominant |
| 16 | 9,538,071.00 | rs8062824 | 2.1377 | 21_bit_twopoint_parametric_dominant |
| 16 | 10,372,098.00 | rs116020376 | 2.0152 | 22_bit_twopoint_parametric_dominant |
| 16 | 10,566,337.00 | rs57889968 | 2.0616 | 21_bit_twopoint_parametric_recessive |
| 16 | 10,566,337.00 | rs57889968 | 2.7128 | 21_bit_twopoint_parametric_dominant |
| 16 | 10,577,178.00 | rs56092102 | 2.4359 | 21_bit_twopoint_parametric_recessive |
| 16 | 10,593,336.00 | rs17684113 | 1.9562 | 22_bit_twopoint_parametric_dominant |
| 16 | 10,593,336.00 | rs17684113 | 2.3568 | 21_bit_twopoint_parametric_dominant |
| 16 | 10,593,336.00 | rs17684113 | 2.644 | 22_bit_twopoint_parametric_recessive |
| 16 | 10,593,336.00 | rs17684113 | 2.9129 | 21_bit_twopoint_parametric_recessive |
| 16 | 11,179,602.00 | GSA-rs62023563 | 2.4879 | 22_bit_twopoint_parametric_recessive |
| 16 | 12,632,406.00 | rs7194011 | 1.8611 | 21_bit_twopoint_parametric_dominant |
| 16 | 12,632,406.00 | rs7194011 | 1.8611 | 21_bit_twopoint_parametric_dominant |
| 16 | 17,089,102.00 | rs1497984 | 1.9025 | 22_bit_twopoint_parametric_recessive |
| 16 | 17,089,102.00 | rs1497984 | 2.1542 | 22_bit_twopoint_parametric_dominant |
| 16 | 23,325,044.00 | rs17256755 | 2.3372 | 22_bit_twopoint_parametric_dominant |
| 16 | 24,852,386.00 | rs274115 | 1.8811 | 22_bit_twopoint_parametric_recessive |
| 16 | 24,887,651.00 | rs274068 | 2.0258 | 22_bit_twopoint_parametric_recessive |
| 16 | 25,759,427.00 | rs11074714 | 2.263 | 22_bit_twopoint_parametric_recessive |
| 16 | 27,205,918.00 | rs6498001 | 1.8671 | 22_bit_twopoint_parametric_recessive |
| 16 | 27,205,918.00 | rs6498001 | 2.3966 | 22_bit_twopoint_parametric_dominant |
| 16 | 27,967,397.00 | rs9941164 | 2.0055 | 21_bit_twopoint_parametric_recessive |
| 16 | 27,985,046.00 | rs953395 | 1.8664 | 22_bit_twopoint_parametric_recessive |
| 16 | 27,985,046.00 | rs953395 | 2.604 | 22_bit_twopoint_parametric_dominant |
| 16 | 27,994,979.00 | rs1121813 | 2.0341 | 22_bit_twopoint_parametric_dominant |
| 16 | 29,209,997.00 | rs252308 | 2.4763 | 21_bit_twopoint_parametric_dominant |
| 16 | 31,096,606.00 | 16:31107927-T-C | 1.8966 | 21_bit_twopoint_parametric_recessive |
| 16 | 48,451,004.00 | rs9928557 | 2.1592 | 21_bit_twopoint_parametric_dominant |
| 16 | 49,814,901.00 | rs4077247 | 2.0218 | 21_bit_twopoint_parametric_dominant |
| 16 | 49,814,901.00 | rs4077247 | 2.0218 | 21_bit_twopoint_parametric_dominant |
| 16 | 50,823,782.00 | rs1861760 | 2.053 | 21_bit_twopoint_parametric_recessive |
| 16 | 51,129,495.00 | rs1007883 | 1.9091 | 21_bit_twopoint_parametric_recessive |
| 16 | 51,129,495.00 | rs1007883 | 2.1492 | 22_bit_twopoint_parametric_dominant |
| 16 | 51,129,495.00 | rs1007883 | 2.3346 | 22_bit_twopoint_parametric_recessive |
| 16 | 52,070,835.00 | rs42771 | 1.8723 | 22_bit_twopoint_parametric_recessive |
| 16 | 52,920,896.00 | rs2387879 | 2.1551 | 22_bit_twopoint_parametric_dominant |
| 16 | 53,162,130.00 | rs1073454 | 2.1557 | 21_bit_twopoint_parametric_dominant |
| 16 | 54,644,393.00 | rs13380763 | 2.047 | 21_bit_twopoint_parametric_recessive |
| 16 | 55,035,449.00 | rs4783863 | 2.0185 | 22_bit_twopoint_parametric_recessive |
| 16 | 56,839,583.00 | rs1138295 | 2.1226 | 22_bit_twopoint_parametric_dominant |
| 16 | 56,862,818.00 | rs3829502 | 2.2675 | 22_bit_twopoint_parametric_dominant |
| 16 | 56,862,818.00 | rs3829502 | 2.2803 | 22_bit_multipoint_parametric_dominant |
| 16 | 57,034,448.00 | rs4783966 | 1.9396 | 22_bit_twopoint_parametric_dominant |
| 16 | 57,034,448.00 | rs4783966 | 1.9396 | 22_bit_twopoint_parametric_dominant |
| 16 | 57,034,448.00 | rs4783966 | 2.3937 | 22_bit_twopoint_parametric_recessive |
| 16 | 57,034,448.00 | rs4783966 | 2.3937 | 22_bit_twopoint_parametric_recessive |
| 16 | 57,667,338.00 | GSA-rs11076198 | 1.9965 | 21_bit_twopoint_parametric_dominant |
| 16 | 57,667,338.00 | GSA-rs11076198 | 1.9965 | 21_bit_twopoint_parametric_dominant |
| 16 | 58,998,224.00 | rs74023021 | 1.9543 | 22_bit_twopoint_parametric_dominant |
| 16 | 60,542,392.00 | GSA-rs112626221 | 1.9639 | 22_bit_twopoint_parametric_dominant |
| 16 | 62,354,780.00 | rs11647822 | 2.0478 | 21_bit_twopoint_parametric_dominant |
| 16 | 65,492,282.00 | JHU_16.65526184 | 2.5233 | 21_bit_twopoint_parametric_dominant |
| 16 | 69,048,914.00 | rs8062856 | 2.0048 | 21_bit_twopoint_parametric_dominant |
| 16 | 69,048,914.00 | rs8062856 | 2.0048 | 21_bit_twopoint_parametric_dominant |
| 16 | 69,048,914.00 | rs8062856 | 2.2194 | 21_bit_twopoint_parametric_recessive |
| 16 | 69,048,914.00 | rs8062856 | 2.2194 | 21_bit_twopoint_parametric_recessive |
| 16 | 70,036,216.00 | JHU_16.70070118 | 2.2228 | 21_bit_twopoint_parametric_dominant |
| 16 | 70,072,296.00 | GSA-rs4985392 | 2.1883 | 21_bit_twopoint_parametric_dominant |
| 16 | 70,662,723.00 | JHU_16.70696625 | 2.4874 | 22_bit_twopoint_parametric_dominant |
| 16 | 71,576,373.00 | GSA-rs74344827 | 1.9283 | 21_bit_twopoint_parametric_recessive |
| 16 | 71,576,373.00 | GSA-rs74344827 | 1.9469 | 22_bit_twopoint_parametric_recessive |
| 16 | 71,649,815.00 | exm1256052 | 2.0014 | 22_bit_twopoint_parametric_dominant |
| 16 | 75,232,709.00 | rs9934251 | 2.0543 | 21_bit_twopoint_parametric_dominant |
| 16 | 75,717,066.00 | GSA-rs72789466 | 1.9329 | 21_bit_twopoint_parametric_dominant |
| 16 | 75,985,634.00 | JHU_16.76019531 | 3.0298 | 21_bit_twopoint_parametric_dominant |
| 16 | 76,196,054.00 | rs3851748 | 2.3731 | 22_bit_twopoint_parametric_recessive |
| 16 | 76,201,812.00 | rs3851752 | 2.4444 | 22_bit_twopoint_parametric_recessive |
| 16 | 76,480,817.00 | rs8057325 | 1.961 | 22_bit_twopoint_parametric_recessive |
| 16 | 76,498,495.00 | GSA-rs12932943 | 2.179 | 21_bit_twopoint_parametric_recessive |
| 16 | 76,498,495.00 | GSA-rs12932943 | 2.4971 | 22_bit_twopoint_parametric_recessive |
| 16 | 76,510,861.00 | rs1506826 | 2.2854 | 21_bit_twopoint_parametric_recessive |
| 16 | 76,510,861.00 | rs1506826 | 2.5389 | 22_bit_twopoint_parametric_recessive |
| 16 | 76,515,449.00 | JHU_16.76549345 | 2.0422 | 22_bit_twopoint_parametric_dominant |
| 16 | 77,385,344.00 | rs2454935 | 2.4241 | 21_bit_twopoint_parametric_dominant |
| 16 | 77,554,998.00 | rs28576941 | 2.7896 | 22_bit_twopoint_parametric_recessive |
| 16 | 80,033,987.00 | rs12051191 | 2.4562 | 22_bit_twopoint_parametric_recessive |
| 16 | 80,353,046.00 | rs4889140 | 1.904 | 21_bit_twopoint_parametric_recessive |
| 16 | 80,353,046.00 | rs4889140 | 2.0081 | 22_bit_twopoint_parametric_recessive |
| 16 | 82,803,101.00 | rs2199430 | 2.0034 | 22_bit_twopoint_parametric_dominant |
| 16 | 83,105,029.00 | rs67972857 | 2.1564 | 22_bit_twopoint_parametric_dominant |
| 16 | 83,105,029.00 | rs67972857 | 2.1564 | 22_bit_twopoint_parametric_dominant |
| 16 | 83,105,029.00 | rs67972857 | 2.2346 | 22_bit_twopoint_parametric_recessive |
| 16 | 83,105,029.00 | rs67972857 | 2.2346 | 22_bit_twopoint_parametric_recessive |
| 16 | 85,471,076.00 | rs876056 | 1.8817 | 21_bit_twopoint_parametric_dominant |
| 16 | 86,993,749.00 | rs7194455 | 2.8513 | 22_bit_twopoint_parametric_dominant |
| 16 | 86,993,749.00 | rs7194455 | 2.8513 | 22_bit_twopoint_parametric_dominant |
| 16 | 88,351,032.00 | GSA-rs62048984 | 2.3106 | 21_bit_twopoint_parametric_dominant |
| 16 | 89,080,306.00 | GSA-rs9921637 | 1.9731 | 22_bit_twopoint_parametric_dominant |
| 16 | 90,044,542.00 | 16:90110950-CT | 1.9496 | 21_bit_twopoint_parametric_dominant |
| 17 | 958,168.00 | JHU_17.861407 | 2.0285 | 21_bit_twopoint_parametric_dominant |
| 17 | 1,907,306.00 | rs75314133 | 1.9385 | 21_bit_twopoint_parametric_recessive |
| 17 | 1,935,058.00 | JHU_17.1838351 | 2.2761 | 22_bit_twopoint_parametric_dominant |
| 17 | 2,525,214.00 | exm-rs9891572 | 2.0562 | 21_bit_twopoint_parametric_recessive |
| 17 | 3,716,844.00 | GSA-rs2293881 | 2.0002 | 21_bit_twopoint_parametric_dominant |
| 17 | 4,487,906.00 | rs2291742 | 2.2959 | 21_bit_multipoint_parametric_dominant |
| 17 | 4,487,906.00 | rs2291742 | 2.2959 | 21_bit_twopoint_parametric_dominant |
| 17 | 5,457,755.00 | GSA-rs3865353 | 1.9392 | 22_bit_twopoint_parametric_dominant |
| 17 | 5,510,073.00 | JHU_17.5413392 | 1.8904 | 22_bit_twopoint_parametric_dominant |
| 17 | 5,511,967.00 | rs4790267 | 2.6277 | 22_bit_twopoint_parametric_dominant |
| 17 | 5,512,364.00 | JHU_17.5415683 | 1.966 | 21_bit_twopoint_parametric_dominant |
| 17 | 5,512,364.00 | JHU_17.5415683 | 2.8649 | 22_bit_twopoint_parametric_dominant |
| 17 | 7,744,908.00 | 17:7648226-G-A | 2.0285 | 21_bit_twopoint_parametric_dominant |
| 17 | 9,238,006.00 | rs3744656 | 1.9385 | 21_bit_twopoint_parametric_recessive |
| 17 | 9,813,594.00 | JHU_17.9716910 | 2.0562 | 21_bit_twopoint_parametric_recessive |
| 17 | 10,328,994.00 | rs8067532 | 1.9404 | 22_bit_twopoint_parametric_recessive |
| 17 | 10,337,647.00 | GSA-rs12952386 | 1.9404 | 22_bit_twopoint_parametric_recessive |
| 17 | 10,805,428.00 | rs11655506 | 2.0002 | 21_bit_twopoint_parametric_dominant |
| 17 | 11,277,406.00 | rs62062007 | 2.2959 | 21_bit_twopoint_parametric_dominant |
| 17 | 12,288,034.00 | rs12453677 | 1.966 | 21_bit_twopoint_parametric_dominant |
| 17 | 12,411,179.00 | GSA-rs62061493 | 2.8914 | 22_bit_twopoint_parametric_dominant |
| 17 | 13,837,532.00 | rs7226344 | 2.6586 | 22_bit_twopoint_parametric_recessive |
| 17 | 14,312,129.00 | JHU_17.14215445 | 2.117 | 21_bit_twopoint_parametric_dominant |
| 17 | 14,566,051.00 | rs9903098 | 2.1293 | 22_bit_twopoint_parametric_recessive |
| 17 | 14,777,067.00 | rs8064366 | 2.526 | 21_bit_twopoint_parametric_dominant |
| 17 | 15,022,059.00 | JHU_17.14925375 | 1.8777 | 22_bit_twopoint_parametric_dominant |
| 17 | 17,303,035.00 | JHU_17.17206348 | 2.1654 | 22_bit_twopoint_parametric_recessive |
| 17 | 17,758,488.00 | rs4925109 | 2.2415 | 22_bit_twopoint_parametric_recessive |
| 17 | 27,404,394.00 | rs10853142 | 2.2005 | 21_bit_twopoint_parametric_recessive |
| 17 | 27,794,895.00 | rs3794766 | 1.9335 | 22_bit_twopoint_parametric_dominant |
| 17 | 31,417,481.00 | exm2268040 | 2.155 | 22_bit_twopoint_parametric_recessive |
| 17 | 31,417,481.00 | exm2268040 | 3.2052 | 22_bit_twopoint_parametric_dominant |
| 17 | 32,614,910.00 | rs321148 | 2.3847 | 22_bit_twopoint_parametric_recessive |
| 17 | 32,621,104.00 | GSA-rs321152 | 2.3109 | 22_bit_twopoint_parametric_dominant |
| 17 | 32,621,104.00 | GSA-rs321152 | 2.4281 | 22_bit_twopoint_parametric_recessive |
| 17 | 32,633,519.00 | exm2267999 | 2.2291 | 22_bit_twopoint_parametric_dominant |
| 17 | 32,633,519.00 | exm2267999 | 2.548 | 22_bit_twopoint_parametric_recessive |
| 17 | 33,123,810.00 | rs72819115 | 2.0675 | 22_bit_twopoint_parametric_dominant |
| 17 | 34,624,718.00 | rs12952673 | 1.9788 | 22_bit_twopoint_parametric_recessive |
| 17 | 35,295,755.00 | rs812800 | 3.0745 | 21_bit_twopoint_parametric_recessive |
| 17 | 35,769,046.00 | rs77838845 | 1.9771 | 21_bit_twopoint_parametric_recessive |
| 17 | 38,996,116.00 | rs8070153 | 2.0074 | 21_bit_twopoint_parametric_recessive |
| 17 | 39,029,511.00 | rs11868358 | 1.9619 | 21_bit_twopoint_parametric_recessive |
| 17 | 39,087,980.00 | rs9896717 | 2.1419 | 21_bit_twopoint_parametric_recessive |
| 17 | 39,132,034.00 | rs3025170 | 1.9318 | 21_bit_twopoint_parametric_recessive |
| 17 | 40,854,610.00 | 17:39010862-A-C | 2.4848 | 21_bit_twopoint_parametric_recessive |
| 17 | 40,854,610.00 | 17:39010862-A-C | 2.4848 | 21_bit_twopoint_parametric_recessive |
| 17 | 41,529,805.00 | rs11650388 | 1.9641 | 22_bit_twopoint_parametric_recessive |
| 17 | 42,214,635.00 | rs185478092 | 2.4166 | 21_bit_twopoint_parametric_dominant |
| 17 | 42,214,635.00 | rs185478092 | 2.4166 | 21_bit_twopoint_parametric_dominant |
| 17 | 42,223,863.00 | rs9900213 | 1.8702 | 21_bit_twopoint_parametric_dominant |
| 17 | 42,223,863.00 | rs9900213 | 1.8702 | 21_bit_twopoint_parametric_dominant |
| 17 | 42,246,955.00 | rs8082391 | 1.8702 | 21_bit_twopoint_parametric_dominant |
| 17 | 42,246,955.00 | rs8082391 | 1.8702 | 21_bit_twopoint_parametric_dominant |
| 17 | 42,308,985.00 | rs2293154 | 2.4166 | 21_bit_twopoint_parametric_dominant |
| 17 | 42,308,985.00 | rs2293154 | 2.4166 | 21_bit_twopoint_parametric_dominant |
| 17 | 42,400,776.00 | rs2883456 | 1.8798 | 21_bit_twopoint_parametric_recessive |
| 17 | 42,985,484.00 | rs12944462 | 2.8158 | 21_bit_twopoint_parametric_dominant |
| 17 | 42,988,250.00 | GSA-rs10852953 | 2.5124 | 21_bit_twopoint_parametric_dominant |
| 17 | 42,988,250.00 | GSA-rs10852953 | 2.5284 | 21_bit_twopoint_parametric_recessive |
| 17 | 43,092,412.00 | GSA-rs4986852 | 1.9857 | 21_bit_twopoint_parametric_dominant |
| 17 | 45,342,556.00 | JHU_17.43419921 | 1.9658 | 21_bit_twopoint_parametric_dominant |
| 17 | 48,755,657.00 | GSA-rs77097215 | 2.2029 | 21_bit_twopoint_parametric_dominant |
| 17 | 49,120,881.00 | rs2898834 | 1.9819 | 21_bit_twopoint_parametric_recessive |
| 17 | 49,698,723.00 | rs4794061 | 2.0493 | 22_bit_twopoint_parametric_recessive |
| 17 | 52,984,063.00 | GSA-rs1512868 | 1.9483 | 21_bit_multipoint_parametric_dominant |
| 17 | 52,984,063.00 | GSA-rs1512868 | 2.0979 | 22_bit_multipoint_parametric_dominant |
| 17 | 52,984,063.00 | GSA-rs1512868 | 2.101 | 22_bit_twopoint_parametric_dominant |
| 17 | 55,615,598.00 | rs1426748 | 1.8841 | 21_bit_twopoint_parametric_recessive |
| 17 | 55,849,135.00 | rs1454113 | 2.0782 | 21_bit_twopoint_parametric_dominant |
| 17 | 55,856,592.00 | rs12603590 | 2.2576 | 21_bit_twopoint_parametric_dominant |
| 17 | 56,729,478.00 | rs13341098 | 1.9836 | 22_bit_twopoint_parametric_recessive |
| 17 | 56,729,478.00 | rs13341098 | 2.3166 | 22_bit_twopoint_parametric_dominant |
| 17 | 57,051,752.00 | GSA-rs12944393 | 1.9415 | 21_bit_twopoint_parametric_dominant |
| 17 | 57,053,472.00 | rs7208729 | 1.891 | 21_bit_twopoint_parametric_dominant |
| 17 | 57,095,801.00 | rs2270277 | 2.5861 | 21_bit_twopoint_parametric_dominant |
| 17 | 57,413,888.00 | rs1542125 | 1.9677 | 22_bit_twopoint_parametric_dominant |
| 17 | 61,468,389.00 | rs7214481 | 2.3598 | 21_bit_twopoint_parametric_recessive |
| 17 | 65,376,202.00 | GSA-rs11871336 | 1.928 | 21_bit_twopoint_parametric_dominant |
| 17 | 67,404,658.00 | rs34696122 | 1.8841 | 21_bit_twopoint_parametric_recessive |
| 17 | 67,516,557.00 | GSA-rs4791287 | 2.0782 | 21_bit_twopoint_parametric_dominant |
| 17 | 67,520,533.00 | rs4791284 | 2.2576 | 21_bit_twopoint_parametric_dominant |
| 17 | 68,713,085.00 | rs7214054 | 1.9415 | 21_bit_twopoint_parametric_dominant |
| 17 | 68,714,557.00 | JHU_17.66710697 | 1.891 | 21_bit_twopoint_parametric_dominant |
| 17 | 68,747,749.00 | GSA-rs78191388 | 2.5861 | 21_bit_twopoint_parametric_dominant |
| 17 | 68,771,439.00 | rs11654057 | 1.9965 | 22_bit_twopoint_parametric_dominant |
| 17 | 68,771,439.00 | rs11654057 | 1.9988 | 22_bit_multipoint_parametric_dominant |
| 17 | 68,776,160.00 | rs1348372 | 1.9267 | 22_bit_twopoint_parametric_dominant |
| 17 | 70,181,624.00 | rs4328485 | 1.944 | 21_bit_multipoint_NPL |
| 17 | 71,602,076.00 | rs9900249 | 2.3598 | 21_bit_twopoint_parametric_recessive |
| 17 | 73,139,631.00 | rs11077665 | 1.928 | 21_bit_twopoint_parametric_dominant |
| 17 | 74,050,040.00 | rs73997403 | 1.8675 | 22_bit_twopoint_parametric_dominant |
| 17 | 74,274,989.00 | rs9912615 | 1.8841 | 21_bit_twopoint_parametric_recessive |
| 17 | 74,336,016.00 | rs2100896 | 3.1616 | 22_bit_twopoint_parametric_dominant |
| 17 | 74,361,476.00 | rs28430711 | 2.0782 | 21_bit_twopoint_parametric_dominant |
| 17 | 74,369,940.00 | JHU_17.72366078 | 2.2576 | 21_bit_twopoint_parametric_dominant |
| 17 | 75,723,177.00 | rs62088208 | 1.9415 | 21_bit_twopoint_parametric_dominant |
| 17 | 75,727,159.00 | rs2584100 | 1.891 | 21_bit_twopoint_parametric_dominant |
| 17 | 75,786,110.00 | exm-rs2125345 | 2.5861 | 21_bit_twopoint_parametric_dominant |
| 17 | 77,635,512.00 | GSA-rs2411134 | 1.9061 | 22_bit_twopoint_parametric_recessive |
| 17 | 77,853,419.00 | rs4789007 | 2.3598 | 21_bit_twopoint_parametric_recessive |
| 17 | 78,643,632.00 | JHU_17.76639713 | 1.9819 | 22_bit_twopoint_parametric_dominant |
| 17 | 78,647,944.00 | rs9896451 | 2.0088 | 22_bit_twopoint_parametric_dominant |
| 17 | 78,739,730.00 | rs74802261 | 2.263 | 22_bit_twopoint_parametric_recessive |
| 17 | 79,221,018.00 | rs11870893 | 1.928 | 21_bit_twopoint_parametric_dominant |
| 17 | 80,495,557.00 | rs9898443 | 1.8841 | 21_bit_twopoint_parametric_recessive |
| 17 | 80,703,105.00 | GSA-rs4062178 | 2.0782 | 21_bit_twopoint_parametric_dominant |
| 17 | 80,710,571.00 | JHU_17.78684370 | 2.2576 | 21_bit_twopoint_parametric_dominant |
| 17 | 81,628,614.00 | JHU_17.79595639 | 1.8903 | 22_bit_twopoint_parametric_dominant |
| 17 | 81,862,798.00 | JHU_17.79820673 | 1.9415 | 21_bit_twopoint_parametric_dominant |
| 17 | 81,866,231.00 | JHU_17.79824106 | 1.891 | 21_bit_twopoint_parametric_dominant |
| 17 | 82,036,150.00 | rs6502047 | 2.5861 | 21_bit_twopoint_parametric_dominant |
| 18 | 965,525.00 | rs1818867 | 1.9284 | 22_bit_twopoint_parametric_recessive |
| 18 | 1,635,747.00 | rs1988035 | 1.9146 | 22_bit_twopoint_parametric_dominant |
| 18 | 2,200,876.00 | rs513990 | 2.4789 | 22_bit_twopoint_parametric_dominant |
| 18 | 2,409,325.00 | rs17524891 | 1.8731 | 21_bit_twopoint_parametric_dominant |
| 18 | 2,518,698.00 | rs3810071 | 2.6515 | 21_bit_twopoint_parametric_dominant |
| 18 | 2,634,429.00 | rs12455652 | 2.0339 | 21_bit_twopoint_parametric_dominant |
| 18 | 2,825,420.00 | GSA-rs673783 | 1.9095 | 22_bit_twopoint_parametric_dominant |
| 18 | 2,825,420.00 | GSA-rs673783 | 2.6129 | 21_bit_twopoint_parametric_dominant |
| 18 | 3,424,360.00 | JHU_18.3424357 | 2.0163 | 21_bit_twopoint_parametric_dominant |
| 18 | 3,448,757.00 | rs238136 | 2.9938 | 21_bit_twopoint_parametric_dominant |
| 18 | 3,471,642.00 | rs9963977 | 2.0216 | 21_bit_twopoint_parametric_dominant |
| 18 | 5,748,544.00 | GSA-rs1917915 | 2.067 | 22_bit_twopoint_parametric_dominant |
| 18 | 5,748,544.00 | GSA-rs1917915 | 2.4545 | 22_bit_twopoint_parametric_recessive |
| 18 | 6,979,622.00 | JHU_18.6979620 | 1.9521 | 22_bit_twopoint_parametric_recessive |
| 18 | 7,041,987.00 | JHU_18.7041985 | 1.8841 | 22_bit_twopoint_parametric_dominant |
| 18 | 7,388,213.00 | rs9955412 | 2.8265 | 22_bit_twopoint_parametric_recessive |
| 18 | 7,486,108.00 | rs554825 | 2.5337 | 21_bit_twopoint_parametric_dominant |
| 18 | 7,706,349.00 | rs9953589 | 2.6336 | 22_bit_twopoint_parametric_dominant |
| 18 | 8,585,098.00 | rs17498343 | 2.0137 | 22_bit_twopoint_parametric_recessive |
| 18 | 8,828,574.00 | rs1564484 | 1.8875 | 22_bit_twopoint_parametric_recessive |
| 18 | 8,882,681.00 | rs8094017 | 2.0423 | 22_bit_multipoint_parametric_dominant |
| 18 | 8,882,681.00 | rs8094017 | 2.0886 | 22_bit_twopoint_parametric_dominant |
| 18 | 9,752,942.00 | rs4798850 | 2.0646 | 22_bit_twopoint_parametric_recessive |
| 18 | 9,973,665.00 | GSA-rs29172 | 2.0814 | 22_bit_twopoint_parametric_recessive |
| 18 | 10,341,254.00 | rs12970879 | 1.8776 | 22_bit_twopoint_parametric_recessive |
| 18 | 10,663,883.00 | rs73945128 | 2.2929 | 22_bit_twopoint_parametric_recessive |
| 18 | 10,663,883.00 | rs73945128 | 2.4269 | 22_bit_twopoint_parametric_dominant |
| 18 | 11,688,980.00 | rs8087266 | 2.0398 | 21_bit_twopoint_parametric_recessive |
| 18 | 11,688,980.00 | rs8087266 | 2.1699 | 22_bit_twopoint_parametric_recessive |
| 18 | 12,236,901.00 | rs59321156 | 1.8719 | 21_bit_twopoint_parametric_dominant |
| 18 | 12,236,901.00 | rs59321156 | 1.8719 | 21_bit_twopoint_parametric_dominant |
| 18 | 13,558,816.00 | rs9947149 | 1.879 | 21_bit_twopoint_NPL |
| 18 | 23,311,063.00 | GSA-rs4800466 | 2.196 | 21_bit_twopoint_parametric_recessive |
| 18 | 24,803,353.00 | rs4800595 | 1.9762 | 21_bit_twopoint_parametric_recessive |
| 18 | 25,833,980.00 | exm2268083 | 2.0129 | 22_bit_twopoint_parametric_dominant |
| 18 | 26,637,593.00 | rs11083175 | 2.5802 | 21_bit_twopoint_parametric_dominant |
| 18 | 29,466,446.00 | rs9954312 | 2.5053 | 21_bit_twopoint_parametric_dominant |
| 18 | 29,466,446.00 | rs9954312 | 2.5053 | 21_bit_twopoint_parametric_dominant |
| 18 | 31,004,808.00 | rs8096598 | 2.0726 | 22_bit_twopoint_parametric_recessive |
| 18 | 31,030,266.00 | rs2640847 | 2.0323 | 22_bit_twopoint_parametric_recessive |
| 18 | 31,597,008.00 | rs3794884 | 2.6125 | 21_bit_twopoint_parametric_dominant |
| 18 | 31,607,316.00 | rs1667255 | 2.0352 | 21_bit_twopoint_parametric_dominant |
| 18 | 31,646,481.00 | rs4799311 | 2.9807 | 21_bit_twopoint_parametric_dominant |
| 18 | 32,760,397.00 | rs12606529 | 2.7076 | 22_bit_twopoint_parametric_recessive |
| 18 | 34,391,122.00 | rs12958537 | 1.9614 | 21_bit_twopoint_parametric_recessive |
| 18 | 35,705,645.00 | GSA-rs613252 | 1.8623 | 21_bit_twopoint_parametric_recessive |
| 18 | 38,116,132.00 | GSA-rs9783925 | 1.9459 | 22_bit_twopoint_parametric_recessive |
| 18 | 38,116,132.00 | GSA-rs9783925 | 2.425 | 21_bit_twopoint_parametric_recessive |
| 18 | 39,379,046.00 | rs12458935 | 1.8666 | 21_bit_twopoint_parametric_dominant |
| 18 | 39,379,046.00 | rs12458935 | 2.7712 | 21_bit_twopoint_parametric_recessive |
| 18 | 41,887,867.00 | JHU_18.39467831 | 2.1408 | 22_bit_twopoint_parametric_recessive |
| 18 | 41,887,867.00 | JHU_18.39467831 | 2.1408 | 22_bit_twopoint_parametric_recessive |
| 18 | 41,984,912.00 | rs9956832 | 2.2535 | 22_bit_twopoint_parametric_recessive |
| 18 | 41,984,912.00 | rs9956832 | 2.2535 | 22_bit_twopoint_parametric_recessive |
| 18 | 42,023,739.00 | JHU_18.39603702 | 2.2535 | 22_bit_twopoint_parametric_recessive |
| 18 | 42,023,739.00 | JHU_18.39603702 | 2.2535 | 22_bit_twopoint_parametric_recessive |
| 18 | 42,217,011.00 | rs7239017 | 1.9883 | 21_bit_twopoint_parametric_dominant |
| 18 | 46,369,350.00 | rs9962113 | 1.8875 | 21_bit_twopoint_parametric_dominant |
| 18 | 47,498,977.00 | rs6507753 | 1.9076 | 22_bit_twopoint_parametric_dominant |
| 18 | 49,940,339.00 | rs1787296 | 2.0192 | 21_bit_twopoint_parametric_dominant |
| 18 | 53,021,171.00 | JHU_18.50547540 | 1.9447 | 21_bit_twopoint_parametric_recessive |
| 18 | 53,021,171.00 | JHU_18.50547540 | 2.234 | 21_bit_twopoint_parametric_dominant |
| 18 | 53,021,171.00 | JHU_18.50547540 | 2.4254 | 22_bit_twopoint_parametric_dominant |
| 18 | 54,814,352.00 | rs8085272 | 2.6484 | 21_bit_twopoint_parametric_recessive |
| 18 | 54,814,352.00 | rs8085272 | 2.9331 | 22_bit_twopoint_parametric_recessive |
| 18 | 57,558,545.00 | rs1736442 | 2.7518 | 22_bit_twopoint_parametric_dominant |
| 18 | 57,599,781.00 | rs4940928 | 1.9884 | 22_bit_twopoint_parametric_dominant |
| 18 | 57,741,643.00 | rs7234711 | 1.905 | 22_bit_twopoint_parametric_recessive |
| 18 | 57,777,506.00 | rs2019535 | 2.1493 | 21_bit_twopoint_parametric_recessive |
| 18 | 57,777,506.00 | rs2019535 | 2.153 | 22_bit_twopoint_parametric_recessive |
| 18 | 63,708,449.00 | rs6567383 | 2.2654 | 22_bit_twopoint_parametric_recessive |
| 18 | 64,242,805.00 | rs4940617 | 2.0752 | 21_bit_twopoint_parametric_dominant |
| 18 | 66,320,442.00 | rs12960184 | 3.2292 | 22_bit_twopoint_parametric_dominant |
| 18 | 66,428,248.00 | rs4544336 | 2.0684 | 22_bit_twopoint_parametric_dominant |
| 18 | 66,536,337.00 | JHU_18.64203573 | 1.9732 | 22_bit_twopoint_parametric_dominant |
| 18 | 69,561,643.00 | rs9949854 | 2.0627 | 21_bit_twopoint_parametric_dominant |
| 18 | 69,561,643.00 | rs9949854 | 2.0627 | 21_bit_twopoint_parametric_dominant |
| 18 | 69,701,120.00 | GSA-rs9963608 | 2.1996 | 21_bit_twopoint_parametric_dominant |
| 18 | 69,701,120.00 | GSA-rs9963608 | 2.1996 | 21_bit_twopoint_parametric_dominant |
| 18 | 72,994,684.00 | rs7236409 | 2.0727 | 21_bit_twopoint_parametric_dominant |
| 18 | 73,417,368.00 | JHU_18.71084602 | 2.7031 | 21_bit_twopoint_parametric_recessive |
| 18 | 73,803,856.00 | rs2115978 | 1.9689 | 21_bit_twopoint_parametric_dominant |
| 18 | 77,298,576.00 | rs11659917 | 2.1789 | 21_bit_twopoint_parametric_dominant |
| 18 | 77,306,598.00 | rs7226437 | 1.8656 | 22_bit_twopoint_parametric_dominant |
| 18 | 77,306,598.00 | rs7226437 | 2.6756 | 21_bit_twopoint_parametric_recessive |
| 18 | 77,306,598.00 | rs7226437 | 2.9311 | 21_bit_twopoint_parametric_dominant |
| 18 | 77,308,214.00 | rs7232802 | 2.0387 | 22_bit_twopoint_parametric_dominant |
| 18 | 77,308,214.00 | rs7232802 | 2.5759 | 21_bit_twopoint_parametric_recessive |
| 18 | 77,308,214.00 | rs7232802 | 2.9843 | 21_bit_twopoint_parametric_dominant |
| 18 | 77,371,899.00 | GSA-rs7235392 | 2.1837 | 21_bit_twopoint_parametric_recessive |
| 18 | 77,765,395.00 | rs2727046 | 2.3207 | 22_bit_twopoint_parametric_dominant |
| 18 | 77,765,395.00 | rs2727046 | 2.37 | 22_bit_twopoint_parametric_recessive |
| 18 | 77,765,395.00 | rs2727046 | 2.4466 | 21_bit_twopoint_parametric_recessive |
| 18 | 77,765,395.00 | rs2727046 | 2.6255 | 21_bit_twopoint_parametric_dominant |
| 18 | 78,322,352.00 | rs596176 | 2.3148 | 21_bit_twopoint_parametric_dominant |
| 18 | 78,473,783.00 | GSA-rs10853399 | 1.9783 | 21_bit_twopoint_parametric_dominant |
| 18 | 78,955,856.00 | GSA-rs1789250 | 2.2253 | 22_bit_twopoint_parametric_dominant |
| 19 | 452,595.00 | JHU_19.452594 | 1.9549 | 21_bit_twopoint_parametric_dominant |
| 19 | 1,958,309.00 | GSA-rs12977121 | 2.027 | 21_bit_twopoint_parametric_recessive |
| 19 | 2,550,109.00 | rs758473 | 2.448 | 22_bit_twopoint_parametric_dominant |
| 19 | 6,422,877.00 | rs2075755 | 2.1172 | 22_bit_twopoint_parametric_recessive |
| 19 | 8,220,723.00 | rs11666866 | 2.04 | 21_bit_twopoint_parametric_dominant |
| 19 | 8,677,305.00 | rs4804078 | 2.5406 | 22_bit_twopoint_parametric_dominant |
| 19 | 8,997,717.00 | rs8111512 | 2.597 | 22_bit_twopoint_parametric_dominant |
| 19 | 11,131,368.00 | rs14158 | 2.2807 | 21_bit_twopoint_parametric_dominant |
| 19 | 11,132,089.00 | 19:11242765-A-G | 2.2135 | 21_bit_twopoint_parametric_dominant |
| 19 | 12,581,187.00 | exm2268218 | 1.8668 | 22_bit_twopoint_parametric_dominant |
| 19 | 12,581,187.00 | exm2268218 | 2.1661 | 21_bit_twopoint_parametric_dominant |
| 19 | 15,478,890.00 | rs1961562 | 2.1702 | 21_bit_twopoint_parametric_recessive |
| 19 | 16,265,335.00 | rs2124905 | 2.8334 | 22_bit_twopoint_parametric_dominant |
| 19 | 16,265,335.00 | rs2124905 | 2.8334 | 22_bit_twopoint_parametric_dominant |
| 19 | 16,265,335.00 | rs2124905 | 3.0633 | 22_bit_twopoint_parametric_recessive |
| 19 | 16,265,335.00 | rs2124905 | 3.0633 | 22_bit_twopoint_parametric_recessive |
| 19 | 16,314,202.00 | rs12973410 | 2.6557 | 22_bit_twopoint_parametric_recessive |
| 19 | 16,315,846.00 | rs386869 | 2.2481 | 22_bit_twopoint_parametric_recessive |
| 19 | 16,776,634.00 | rs12974841 | 2.4107 | 21_bit_twopoint_parametric_recessive |
| 19 | 17,970,182.00 | rs7252014 | 1.8632 | 21_bit_twopoint_parametric_dominant |
| 19 | 18,786,630.00 | GSA-rs61739916 | 2.0789 | 21_bit_twopoint_parametric_recessive |
| 19 | 18,786,630.00 | GSA-rs61739916 | 2.7462 | 21_bit_twopoint_parametric_dominant |
| 19 | 29,452,313.00 | rs4805409 | 2.2896 | 21_bit_twopoint_parametric_dominant |
| 19 | 30,035,852.00 | JHU_19.30526758 | 2.3799 | 22_bit_twopoint_parametric_dominant |
| 19 | 33,383,852.00 | JHU_19.33874757 | 1.8684 | 22_bit_twopoint_parametric_recessive |
| 19 | 33,383,852.00 | JHU_19.33874757 | 1.8723 | 22_bit_multipoint_parametric_recessive |
| 19 | 33,779,347.00 | JHU_19.34270251 | 1.881 | 22_bit_twopoint_parametric_dominant |
| 19 | 33,818,627.00 | exm-rs29941 | 1.9142 | 21_bit_twopoint_parametric_recessive |
| 19 | 33,818,627.00 | exm-rs29941 | 2.4542 | 22_bit_twopoint_parametric_recessive |
| 19 | 34,646,097.00 | rs8105937 | 1.9547 | 22_bit_multipoint_parametric_dominant |
| 19 | 34,646,097.00 | rs8105937 | 1.9912 | 22_bit_twopoint_parametric_dominant |
| 19 | 35,345,627.00 | exm1455869 | 2.7257 | 22_bit_twopoint_parametric_dominant |
| 19 | 36,337,315.00 | rs1129376 | 2.0599 | 22_bit_twopoint_parametric_recessive |
| 19 | 36,337,315.00 | rs1129376 | 2.8906 | 22_bit_twopoint_parametric_dominant |
| 19 | 38,652,649.00 | rs13344413 | 2.071 | 21_bit_multipoint_NPL |
| 19 | 38,951,581.00 | rs35575208 | 1.9469 | 22_bit_twopoint_parametric_dominant |
| 19 | 39,518,826.00 | rs28596775 | 2.1265 | 22_bit_twopoint_parametric_recessive |
| 19 | 39,686,196.00 | rs4803286 | 1.9148 | 21_bit_twopoint_parametric_recessive |
| 19 | 39,691,145.00 | JHU_19.40181784 | 1.8742 | 21_bit_twopoint_parametric_recessive |
| 19 | 41,137,161.00 | JHU_19.41643065 | 2.4351 | 22_bit_twopoint_parametric_dominant |
| 19 | 41,137,161.00 | JHU_19.41643065 | 2.4351 | 22_bit_twopoint_parametric_dominant |
| 19 | 43,963,329.00 | rs112590975 | 1.9921 | 21_bit_twopoint_parametric_recessive |
| 19 | 47,722,470.00 | rs2974237 | 2.1419 | 22_bit_twopoint_parametric_dominant |
| 19 | 48,954,681.00 | 19:49457938-A-G | 1.9745 | 22_bit_twopoint_parametric_dominant |
| 19 | 48,954,681.00 | 19:49457938-A-G | 2.2377 | 21_bit_twopoint_parametric_dominant |
| 19 | 48,957,658.00 | rs1010104 | 1.97 | 22_bit_twopoint_parametric_dominant |
| 19 | 51,457,686.00 | exm1497186 | 1.9986 | 21_bit_twopoint_parametric_recessive |
| 19 | 52,114,045.00 | rs10418739 | 2.4121 | 21_bit_twopoint_parametric_dominant |
| 19 | 54,245,208.00 | rs11668526 | 1.908 | 21_bit_twopoint_parametric_recessive |
| 19 | 55,372,491.00 | rs117240443 | 1.9817 | 21_bit_twopoint_parametric_dominant |
| 19 | 55,372,491.00 | rs117240443 | 2.0737 | 22_bit_twopoint_parametric_recessive |
| 19 | 55,389,731.00 | rs10425596 | 1.9003 | 21_bit_twopoint_parametric_dominant |
| 19 | 55,805,430.00 | rs299169 | 2.1668 | 22_bit_twopoint_parametric_dominant |
| 19 | 55,805,430.00 | rs299169 | 2.1668 | 22_bit_twopoint_parametric_dominant |
| 20 | 276,391.00 | GSA-rs3827153 | 2.1445 | 22_bit_twopoint_parametric_dominant |
| 20 | 789,716.00 | rs73071720 | 2.1582 | 22_bit_twopoint_parametric_recessive |
| 20 | 3,058,469.00 | rs8184236 | 1.9838 | 21_bit_twopoint_parametric_dominant |
| 20 | 4,704,905.00 | rs6037937 | 2.1031 | 22_bit_twopoint_parametric_recessive |
| 20 | 6,065,649.00 | rs745297 | 1.9749 | 22_bit_twopoint_parametric_dominant |
| 20 | 6,065,649.00 | rs745297 | 2.4572 | 21_bit_twopoint_parametric_dominant |
| 20 | 7,634,255.00 | rs2206394 | 1.9361 | 22_bit_twopoint_parametric_recessive |
| 20 | 7,634,255.00 | rs2206394 | 2.2357 | 22_bit_twopoint_parametric_dominant |
| 20 | 10,129,589.00 | GSA-rs3761161 | 2.236 | 22_bit_twopoint_parametric_dominant |
| 20 | 10,129,589.00 | GSA-rs3761161 | 2.2661 | 22_bit_multipoint_parametric_dominant |
| 20 | 11,603,340.00 | rs7260891 | 2.927 | 22_bit_twopoint_parametric_dominant |
| 20 | 12,732,422.00 | rs6033504 | 2.0891 | 22_bit_twopoint_parametric_dominant |
| 20 | 16,918,932.00 | rs2057245 | 2.06 | 22_bit_twopoint_parametric_recessive |
| 20 | 17,523,456.00 | rs2269051 | 1.9041 | 22_bit_twopoint_parametric_recessive |
| 20 | 18,918,138.00 | JHU_20.18898781 | 2.0165 | 22_bit_twopoint_parametric_recessive |
| 20 | 18,918,138.00 | JHU_20.18898781 | 2.1468 | 21_bit_twopoint_parametric_recessive |
| 20 | 18,981,664.00 | rs2104196 | 2.4314 | 22_bit_twopoint_parametric_recessive |
| 20 | 19,089,508.00 | rs6132156 | 1.9903 | 22_bit_twopoint_parametric_recessive |
| 20 | 19,094,560.00 | GSA-rs2208795 | 2.0655 | 22_bit_twopoint_parametric_dominant |
| 20 | 19,217,646.00 | rs6112259 | 2.2273 | 22_bit_twopoint_parametric_recessive |
| 20 | 19,217,646.00 | rs6112259 | 2.3162 | 21_bit_twopoint_parametric_recessive |
| 20 | 19,217,646.00 | rs6112259 | 2.4286 | 21_bit_twopoint_parametric_dominant |
| 20 | 19,217,646.00 | rs6112259 | 2.4505 | 22_bit_twopoint_parametric_dominant |
| 20 | 20,402,268.00 | rs6106244 | 1.9934 | 22_bit_twopoint_parametric_dominant |
| 20 | 24,509,809.00 | rs34320942 | 2.342 | 22_bit_multipoint_parametric_recessive |
| 20 | 24,509,809.00 | rs34320942 | 2.35 | 22_bit_twopoint_parametric_recessive |
| 20 | 32,004,611.00 | rs6061088 | 1.9194 | 22_bit_twopoint_parametric_dominant |
| 20 | 35,355,478.00 | 20:33943281-C-T | 2.047 | 22_bit_twopoint_parametric_recessive |
| 20 | 35,393,057.00 | 20:33980860-T-C | 2.0896 | 22_bit_twopoint_parametric_recessive |
| 20 | 38,286,018.00 | rs6064324 | 1.9395 | 22_bit_twopoint_parametric_dominant |
| 20 | 39,245,170.00 | GSA-rs932426 | 2.3884 | 21_bit_twopoint_parametric_dominant |
| 20 | 40,649,002.00 | rs735031 | 1.9541 | 21_bit_twopoint_parametric_dominant |
| 20 | 40,649,002.00 | rs735031 | 2.1627 | 21_bit_twopoint_parametric_recessive |
| 20 | 40,660,250.00 | rs10485673 | 2.5921 | 22_bit_twopoint_parametric_dominant |
| 20 | 40,670,812.00 | GSA-rs6016412 | 2.1997 | 22_bit_twopoint_parametric_dominant |
| 20 | 40,692,111.00 | rs6102095 | 2.7325 | 22_bit_twopoint_parametric_dominant |
| 20 | 41,880,396.00 | rs6102580 | 2.1188 | 21_bit_twopoint_parametric_recessive |
| 20 | 41,880,396.00 | rs6102580 | 2.1188 | 21_bit_twopoint_parametric_recessive |
| 20 | 41,880,396.00 | rs6102580 | 2.3108 | 21_bit_twopoint_parametric_dominant |
| 20 | 41,880,396.00 | rs6102580 | 2.3108 | 21_bit_twopoint_parametric_dominant |
| 20 | 42,029,278.00 | GSA-rs2223913 | 2.6664 | 21_bit_twopoint_parametric_dominant |
| 20 | 42,029,278.00 | GSA-rs2223913 | 2.6664 | 21_bit_twopoint_parametric_dominant |
| 20 | 42,931,754.00 | rs761027 | 2.1606 | 22_bit_twopoint_parametric_recessive |
| 20 | 44,122,832.00 | rs4810411 | 2.4474 | 22_bit_twopoint_parametric_dominant |
| 20 | 48,216,323.00 | rs6095016 | 1.954 | 21_bit_multipoint_NPL |
| 20 | 50,254,742.00 | rs6020400 | 2.0467 | 21_bit_twopoint_parametric_recessive |
| 20 | 50,254,742.00 | rs6020400 | 2.0467 | 21_bit_twopoint_parametric_recessive |
| 20 | 50,288,310.00 | rs6122897 | 2.7513 | 21_bit_twopoint_parametric_dominant |
| 20 | 50,288,310.00 | rs6122897 | 2.7513 | 21_bit_twopoint_parametric_dominant |
| 20 | 50,431,903.00 | GSA-rs13038695 | 1.9606 | 22_bit_twopoint_parametric_recessive |
| 20 | 50,431,903.00 | GSA-rs13038695 | 1.9922 | 21_bit_twopoint_parametric_recessive |
| 20 | 50,611,452.00 | JHU_20.49227988 | 1.9459 | 21_bit_twopoint_parametric_recessive |
| 20 | 50,621,890.00 | rs768175 | 2.0757 | 21_bit_twopoint_parametric_recessive |
| 20 | 53,451,602.00 | rs6063947 | 2.4701 | 22_bit_twopoint_parametric_dominant |
| 20 | 53,495,477.00 | GSA-rs1999602 | 2.1089 | 22_bit_twopoint_parametric_dominant |
| 20 | 53,766,245.00 | rs6126943 | 2.2628 | 22_bit_twopoint_parametric_dominant |
| 20 | 53,949,222.00 | JHU_20.52565760 | 2.154 | 21_bit_twopoint_parametric_recessive |
| 20 | 53,949,222.00 | JHU_20.52565760 | 2.457 | 21_bit_twopoint_parametric_dominant |
| 20 | 53,958,200.00 | rs6091793 | 2.181 | 21_bit_twopoint_NPL |
| 20 | 53,958,200.00 | rs6091793 | 2.8844 | 21_bit_twopoint_parametric_dominant |
| 20 | 53,958,200.00 | rs6091793 | 3.2287 | 21_bit_twopoint_parametric_recessive |
| 20 | 56,695,437.00 | rs4583526 | 1.9596 | 22_bit_twopoint_parametric_recessive |
| 20 | 57,443,029.00 | rs328502 | 2.136 | 22_bit_twopoint_parametric_recessive |
| 20 | 57,443,029.00 | rs328502 | 2.136 | 22_bit_twopoint_parametric_recessive |
| 20 | 58,496,992.00 | rs1077350 | 1.9679 | 21_bit_twopoint_parametric_dominant |
| 20 | 58,549,716.00 | rs6064671 | 2.1218 | 21_bit_twopoint_parametric_recessive |
| 20 | 58,549,716.00 | rs6064671 | 2.1498 | 22_bit_twopoint_parametric_recessive |
| 20 | 58,549,716.00 | rs6064671 | 2.2625 | 21_bit_twopoint_parametric_dominant |
| 20 | 58,606,374.00 | rs76032679 | 1.8916 | 22_bit_twopoint_parametric_dominant |
| 20 | 59,249,254.00 | rs259964 | 1.9277 | 21_bit_twopoint_parametric_recessive |
| 20 | 59,249,254.00 | rs259964 | 2.0815 | 21_bit_twopoint_parametric_dominant |
| 20 | 60,598,272.00 | rs846201 | 2.4955 | 22_bit_twopoint_parametric_dominant |
| 20 | 60,788,783.00 | rs1321715 | 2.1945 | 22_bit_twopoint_parametric_recessive |
| 20 | 60,788,783.00 | rs1321715 | 2.2688 | 22_bit_twopoint_parametric_dominant |
| 20 | 61,261,906.00 | rs1106437 | 2.0098 | 22_bit_twopoint_parametric_recessive |
| 20 | 61,261,906.00 | rs1106437 | 2.0309 | 21_bit_twopoint_parametric_recessive |
| 20 | 61,555,702.00 | GSA-rs1936203 | 1.9817 | 21_bit_twopoint_parametric_dominant |
| 20 | 61,963,596.00 | rs6089573 | 2.0729 | 22_bit_twopoint_parametric_dominant |
| 20 | 62,160,042.00 | rs6121926 | 2.9329 | 22_bit_twopoint_parametric_dominant |
| 20 | 62,896,026.00 | rs6090155 | 2.222 | 21_bit_twopoint_parametric_recessive |
| 20 | 62,905,586.00 | 20:61536938-GA | 2.222 | 21_bit_twopoint_parametric_recessive |
| 20 | 63,355,597.00 | rs2273500 | 1.9723 | 21_bit_twopoint_parametric_recessive |
| 21 | 16,190,949.00 | rs2051347 | 2.4383 | 21_bit_twopoint_parametric_dominant |
| 21 | 16,370,733.00 | rs427761 | 2.0529 | 22_bit_twopoint_parametric_recessive |
| 21 | 16,675,887.00 | rs2823966 | 2.8424 | 22_bit_twopoint_parametric_dominant |
| 21 | 16,799,511.00 | rs2824052 | 2.1438 | 21_bit_twopoint_parametric_recessive |
| 21 | 17,011,414.00 | rs765143 | 1.8953 | 22_bit_twopoint_parametric_recessive |
| 21 | 17,517,899.00 | rs211953 | 2.477 | 21_bit_twopoint_parametric_dominant |
| 21 | 19,034,091.00 | rs2825298 | 2.1291 | 22_bit_twopoint_parametric_recessive |
| 21 | 19,034,091.00 | rs2825298 | 2.1636 | 21_bit_twopoint_parametric_recessive |
| 21 | 19,548,780.00 | rs2825631 | 1.9381 | 22_bit_twopoint_parametric_dominant |
| 21 | 20,311,891.00 | rs2826165 | 2.305 | 21_bit_twopoint_parametric_recessive |
| 21 | 20,333,984.00 | rs2826182 | 1.963 | 21_bit_twopoint_parametric_recessive |
| 21 | 20,333,984.00 | rs2826182 | 2.274 | 22_bit_twopoint_parametric_recessive |
| 21 | 21,109,929.00 | rs2826655 | 2.4344 | 21_bit_twopoint_parametric_dominant |
| 21 | 21,109,929.00 | rs2826655 | 3.2544 | 21_bit_twopoint_parametric_recessive |
| 21 | 21,115,166.00 | rs3920995 | 2.8039 | 21_bit_twopoint_parametric_recessive |
| 21 | 21,115,536.00 | rs2826659 | 2.0967 | 21_bit_twopoint_parametric_recessive |
| 21 | 23,696,631.00 | GSA-rs115142075 | 2.5614 | 21_bit_twopoint_parametric_recessive |
| 21 | 23,730,737.00 | rs2828473 | 2.0646 | 21_bit_twopoint_parametric_recessive |
| 21 | 24,069,583.00 | rs1783354 | 2.3742 | 21_bit_twopoint_parametric_recessive |
| 21 | 26,077,619.00 | GSA-rs117349040 | 1.9987 | 22_bit_twopoint_parametric_recessive |
| 21 | 26,077,619.00 | GSA-rs117349040 | 2.2332 | 21_bit_twopoint_parametric_recessive |
| 21 | 26,348,344.00 | GSA-rs2830182 | 2.2469 | 21_bit_twopoint_parametric_recessive |
| 21 | 27,111,042.00 | rs2830701 | 1.9117 | 21_bit_twopoint_parametric_recessive |
| 21 | 27,111,042.00 | rs2830701 | 2.281 | 22_bit_twopoint_parametric_recessive |
| 21 | 27,111,042.00 | rs2830701 | 2.4819 | 22_bit_twopoint_parametric_dominant |
| 21 | 27,117,386.00 | rs2212824 | 1.8748 | 21_bit_twopoint_parametric_recessive |
| 21 | 27,117,386.00 | rs2212824 | 2.1144 | 22_bit_twopoint_parametric_recessive |
| 21 | 27,117,386.00 | rs2212824 | 2.3249 | 22_bit_twopoint_parametric_dominant |
| 21 | 27,618,127.00 | rs8184916 | 2.5697 | 21_bit_twopoint_parametric_dominant |
| 21 | 28,109,155.00 | rs2831503 | 2.2452 | 22_bit_twopoint_parametric_recessive |
| 21 | 28,109,155.00 | rs2831503 | 2.4799 | 21_bit_twopoint_parametric_recessive |
| 21 | 28,115,469.00 | GSA-rs34030490 | 2.3331 | 21_bit_twopoint_parametric_dominant |
| 21 | 28,142,290.00 | rs2831534 | 1.9469 | 21_bit_twopoint_parametric_dominant |
| 21 | 28,187,913.00 | rs10482989 | 1.9527 | 22_bit_twopoint_parametric_recessive |
| 21 | 28,336,083.00 | JHU_21.29708401 | 1.9599 | 22_bit_twopoint_parametric_recessive |
| 21 | 28,345,820.00 | rs2831715 | 2.0178 | 22_bit_twopoint_parametric_recessive |
| 21 | 29,151,972.00 | rs3787662 | 2.0368 | 22_bit_twopoint_parametric_dominant |
| 21 | 31,965,137.00 | GSA-rs9976085 | 1.9999 | 21_bit_twopoint_parametric_dominant |
| 21 | 31,965,137.00 | GSA-rs9976085 | 1.9999 | 21_bit_twopoint_parametric_dominant |
| 21 | 31,984,156.00 | rs8130810 | 1.9083 | 22_bit_twopoint_parametric_recessive |
| 21 | 31,984,156.00 | rs8130810 | 2.4749 | 21_bit_twopoint_parametric_recessive |
| 21 | 31,992,308.00 | rs12165267 | 1.9402 | 21_bit_twopoint_parametric_dominant |
| 21 | 31,992,308.00 | rs12165267 | 2.4094 | 21_bit_twopoint_parametric_recessive |
| 21 | 31,994,840.00 | rs9981748 | 2.0305 | 21_bit_twopoint_parametric_recessive |
| 21 | 31,994,840.00 | rs9981748 | 2.1726 | 21_bit_twopoint_parametric_dominant |
| 21 | 32,142,576.00 | rs2833678 | 2.2271 | 22_bit_twopoint_parametric_dominant |
| 21 | 32,142,576.00 | rs2833678 | 2.344 | 22_bit_twopoint_parametric_recessive |
| 21 | 33,785,207.00 | rs2256797 | 2.1391 | 21_bit_twopoint_parametric_recessive |
| 21 | 34,598,414.00 | 21:35970712-G-T | 2.1843 | 22_bit_twopoint_parametric_recessive |
| 21 | 34,613,596.00 | rs915540 | 2.2013 | 22_bit_twopoint_parametric_recessive |
| 21 | 34,754,342.00 | rs2834630 | 1.9881 | 21_bit_twopoint_parametric_dominant |
| 21 | 35,155,502.00 | exm2268389 | 1.872 | 21_bit_twopoint_parametric_recessive |
| 21 | 35,155,502.00 | exm2268389 | 1.8999 | 22_bit_twopoint_parametric_recessive |
| 21 | 37,866,307.00 | rs762147 | 1.8735 | 22_bit_twopoint_parametric_recessive |
| 21 | 37,866,307.00 | rs762147 | 1.9117 | 21_bit_twopoint_parametric_recessive |
| 21 | 39,914,089.00 | rs2837291 | 2.2745 | 22_bit_twopoint_parametric_dominant |
| 21 | 43,493,593.00 | GSA-rs73365815 | 2.0748 | 21_bit_twopoint_parametric_recessive |
| 21 | 43,528,804.00 | rs75836229 | 2.0771 | 21_bit_twopoint_parametric_recessive |
| 21 | 44,239,562.00 | JHU_21.45659444 | 2.771 | 21_bit_twopoint_parametric_recessive |
| 21 | 44,239,562.00 | JHU_21.45659444 | 3.1657 | 22_bit_twopoint_parametric_recessive |
| 21 | 44,239,562.00 | JHU_21.45659444 | 3.2541 | 21_bit_twopoint_parametric_dominant |
| 21 | 44,528,188.00 | rs2838579 | 2.7055 | 21_bit_twopoint_parametric_dominant |
| 21 | 44,917,311.00 | JHU_21.46337225 | 1.881 | 21_bit_multipoint_NPL |
| 22 | 19,312,180.00 | rs5993595 | 2.1255 | 22_bit_twopoint_parametric_recessive |
| 22 | 19,350,110.00 | rs885981 | 2.0191 | 22_bit_twopoint_parametric_recessive |
| 22 | 19,393,191.00 | GSA-rs5993624 | 2.1213 | 22_bit_twopoint_parametric_recessive |
| 22 | 20,168,892.00 | rs28384 | 1.8905 | 22_bit_twopoint_parametric_dominant |
| 22 | 20,205,591.00 | rs640836 | 2.0758 | 21_bit_twopoint_parametric_recessive |
| 22 | 20,205,591.00 | rs640836 | 2.6717 | 21_bit_twopoint_parametric_dominant |
| 22 | 20,596,277.00 | JHU_22.20950563 | 1.9685 | 22_bit_twopoint_parametric_recessive |
| 22 | 20,596,277.00 | JHU_22.20950563 | 2.5327 | 21_bit_twopoint_parametric_recessive |
| 22 | 23,557,777.00 | rs9620289 | 2.4677 | 22_bit_twopoint_parametric_dominant |
| 22 | 26,983,867.00 | rs7289738 | 2.5543 | 22_bit_twopoint_parametric_recessive |
| 22 | 26,983,867.00 | rs7289738 | 3.059 | 22_bit_twopoint_parametric_dominant |
| 22 | 27,045,417.00 | rs3091387 | 2.3969 | 21_bit_twopoint_parametric_dominant |
| 22 | 27,892,148.00 | rs1016495 | 2.0903 | 21_bit_twopoint_parametric_recessive |
| 22 | 27,892,148.00 | rs1016495 | 2.5547 | 22_bit_twopoint_parametric_recessive |
| 22 | 30,650,600.00 | rs5994328 | 1.9171 | 22_bit_twopoint_parametric_recessive |
| 22 | 30,650,600.00 | rs5994328 | 2.1632 | 22_bit_twopoint_parametric_dominant |
| 22 | 32,554,853.00 | JHU_22.32950838 | 2.4743 | 22_bit_twopoint_parametric_recessive |
| 22 | 33,131,419.00 | rs137343 | 2.0236 | 22_bit_twopoint_parametric_dominant |
| 22 | 33,739,021.00 | rs239330 | 1.9748 | 21_bit_twopoint_parametric_dominant |
| 22 | 34,164,387.00 | rs12485116 | 2.2366 | 21_bit_twopoint_parametric_dominant |
| 22 | 36,592,607.00 | rs2283985 | 1.9026 | 22_bit_twopoint_parametric_recessive |
| 22 | 36,845,745.00 | rs4821540 | 2.6872 | 21_bit_twopoint_parametric_dominant |
| 22 | 37,066,896.00 | exm1605376 | 1.9247 | 22_bit_twopoint_parametric_dominant |
| 22 | 37,066,896.00 | exm1605376 | 2.4737 | 21_bit_twopoint_parametric_recessive |
| 22 | 37,066,896.00 | exm1605376 | 2.8194 | 21_bit_twopoint_parametric_dominant |
| 22 | 37,411,264.00 | rs6000725 | 1.9443 | 22_bit_twopoint_parametric_recessive |
| 22 | 37,555,444.00 | rs5995460 | 1.9848 | 22_bit_twopoint_parametric_dominant |
| 22 | 37,562,261.00 | rs2235336 | 2.0879 | 22_bit_twopoint_parametric_dominant |
| 22 | 39,696,859.00 | rs5757777 | 2.01 | 21_bit_twopoint_parametric_dominant |
| 22 | 42,821,074.00 | rs17003362 | 2.1122 | 21_bit_twopoint_parametric_dominant |
| 22 | 44,003,881.00 | rs6006610 | 2.9041 | 22_bit_twopoint_parametric_dominant |
| 22 | 44,438,412.00 | rs8135880 | 2.0926 | 21_bit_twopoint_parametric_dominant |
| 22 | 44,438,412.00 | rs8135880 | 2.0926 | 21_bit_twopoint_parametric_dominant |
| 22 | 44,782,943.00 | JHU_22.45178822 | 1.9471 | 21_bit_twopoint_parametric_recessive |
| 22 | 44,783,767.00 | JHU_22.45179646 | 1.9539 | 21_bit_twopoint_parametric_recessive |
| 22 | 44,784,674.00 | GSA-rs5766005 | 1.9571 | 21_bit_twopoint_parametric_recessive |
| 22 | 45,915,087.00 | GSA-rs76895390 | 2.057 | 22_bit_twopoint_parametric_recessive |
| 22 | 45,915,087.00 | GSA-rs76895390 | 2.8717 | 22_bit_twopoint_parametric_dominant |
| 22 | 47,873,762.00 | rs2064803 | 1.8601 | 21_bit_twopoint_parametric_dominant |
| 22 | 47,873,762.00 | rs2064803 | 2.3738 | 22_bit_twopoint_parametric_dominant |
| 22 | 47,968,221.00 | rs4823509 | 2.1311 | 22_bit_twopoint_parametric_dominant |
| 22 | 48,426,578.00 | GSA-rs4823768 | 1.8784 | 21_bit_twopoint_parametric_dominant |
| 22 | 48,616,850.00 | rs28621643 | 2.1492 | 21_bit_twopoint_parametric_recessive |
| 22 | 48,616,850.00 | rs28621643 | 2.5924 | 22_bit_twopoint_parametric_recessive |
| 22 | 48,616,850.00 | rs28621643 | 2.6595 | 22_bit_twopoint_parametric_dominant |
| 22 | 49,443,116.00 | rs5770437 | 1.8815 | 21_bit_twopoint_parametric_recessive |
| 22 | 49,443,116.00 | rs5770437 | 1.8815 | 21_bit_twopoint_parametric_recessive |
| 23 | 8,516,982.00 | rs16985016 | 1.9493 | 22_bit_twopoint_parametric_recessive |
| 23 | 9,535,270.00 | rs2066887 | 2.1408 | 22_bit_twopoint_parametric_dominant |
| 23 | 11,671,173.00 | rs4830744 | 2.4561 | 22_bit_twopoint_parametric_dominant |
| 23 | 12,902,616.00 | rs5978593 | 2.4982 | 22_bit_twopoint_parametric_dominant |
| 23 | 13,337,818.00 | rs5978624 | 2.1093 | 21_bit_twopoint_parametric_dominant |
| 23 | 15,761,058.00 | rs145432050 | 2.1664 | 22_bit_twopoint_parametric_dominant |
| 23 | 23,588,173.00 | rs4562492 | 2.0443 | 22_bit_twopoint_parametric_recessive |
| 23 | 23,588,173.00 | rs4562492 | 2.2585 | 21_bit_twopoint_parametric_recessive |
| 23 | 23,588,173.00 | rs4562492 | 2.7401 | 21_bit_twopoint_parametric_dominant |
| 23 | 25,036,812.00 | rs7064660 | 2.3361 | 21_bit_twopoint_parametric_dominant |
| 23 | 25,702,515.00 | JHU_X.25720631 | 2.2784 | 21_bit_twopoint_parametric_recessive |
| 23 | 26,818,613.00 | rs4263917 | 2.1154 | 21_bit_twopoint_parametric_recessive |
| 23 | 26,818,613.00 | rs4263917 | 2.4832 | 22_bit_twopoint_parametric_recessive |
| 23 | 27,842,305.00 | rs12557438 | 2.322 | 21_bit_twopoint_parametric_recessive |
| 23 | 27,860,545.00 | rs139745373 | 2.3259 | 21_bit_twopoint_parametric_recessive |
| 23 | 27,958,260.00 | rs149871106 | 1.961 | 22_bit_twopoint_parametric_recessive |
| 23 | 27,958,260.00 | rs149871106 | 2.5325 | 21_bit_twopoint_parametric_recessive |
| 23 | 39,960,297.00 | rs5917336 | 2.1322 | 22_bit_twopoint_parametric_recessive |
| 23 | 47,108,225.00 | rs6609458 | 1.9777 | 22_bit_twopoint_parametric_dominant |
| 23 | 47,772,836.00 | rs5952451 | 2.1612 | 22_bit_twopoint_parametric_dominant |
| 23 | 47,792,766.00 | rs5952455 | 1.8792 | 22_bit_twopoint_parametric_dominant |
| 23 | 71,184,495.00 | rs12007241 | 1.8777 | 22_bit_twopoint_parametric_dominant |
| 23 | 94,096,802.00 | rs5983343 | 1.8903 | 22_bit_twopoint_parametric_dominant |
| 23 | 94,174,579.00 | rs808745 | 2.0066 | 22_bit_twopoint_parametric_dominant |
| 23 | 95,986,447.00 | rs1323752 | 2.2457 | 22_bit_twopoint_parametric_dominant |
| 23 | 95,986,447.00 | rs1323752 | 2.4537 | 22_bit_twopoint_parametric_recessive |
| 23 | 122,519,985.00 | rs2840655 | 1.957 | 21_bit_twopoint_parametric_recessive |
| 23 | 122,586,359.00 | rs1454658 | 1.9451 | 21_bit_twopoint_parametric_recessive |
| 23 | 124,219,312.00 | rs17330749 | 2.1998 | 21_bit_twopoint_parametric_recessive |
| 23 | 124,226,042.00 | rs9887524 | 2.1992 | 21_bit_twopoint_parametric_recessive |
| 23 | 124,228,004.00 | JHU_X.123361853 | 2.1256 | 21_bit_twopoint_parametric_recessive |
| 23 | 124,448,459.00 | rs2283769 | 2.5131 | 21_bit_twopoint_parametric_recessive |
| 23 | 132,634,492.00 | rs243451 | 2.2168 | 22_bit_twopoint_parametric_recessive |
| 23 | 134,908,302.00 | rs5978060 | 1.8698 | 22_bit_twopoint_parametric_recessive |
| 23 | 137,574,263.00 | rs113378176 | 1.9077 | 22_bit_twopoint_parametric_recessive |
| 23 | 137,633,975.00 | rs112297323 | 2.1748 | 22_bit_twopoint_parametric_recessive |
| 23 | 140,046,443.00 | rs11796241 | 1.9775 | 22_bit_twopoint_parametric_recessive |
| 23 | 140,785,027.00 | rs5954093 | 1.9683 | 22_bit_twopoint_parametric_recessive |
| 23 | 140,799,636.00 | rs72617949 | 2.3633 | 22_bit_twopoint_parametric_recessive |
| 23 | 141,340,425.00 | rs6528829 | 2.0908 | 22_bit_twopoint_parametric_recessive |
| 23 | 141,957,398.00 | rs7880927 | 2.0755 | 22_bit_twopoint_parametric_recessive |
| 23 | 141,957,398.00 | rs7880927 | 2.3002 | 21_bit_twopoint_parametric_recessive |
| 23 | 141,967,822.00 | rs1980465 | 1.9228 | 21_bit_twopoint_parametric_recessive |
| 23 | 141,967,822.00 | rs1980465 | 2.3633 | 22_bit_twopoint_parametric_recessive |
| 23 | 141,973,074.00 | rs3135488 | 2.3117 | 22_bit_twopoint_parametric_recessive |
| 23 | 141,973,074.00 | rs3135488 | 2.4488 | 21_bit_twopoint_parametric_recessive |
| 23 | 141,982,218.00 | rs5954476 | 1.8924 | 21_bit_twopoint_parametric_recessive |
| 23 | 141,982,218.00 | rs5954476 | 2.0998 | 22_bit_twopoint_parametric_recessive |
| 23 | 143,427,970.00 | rs2903103 | 2.0612 | 22_bit_twopoint_parametric_recessive |
| 23 | 144,601,046.00 | JHU_X.143682566 | 2.1415 | 22_bit_twopoint_parametric_dominant |
| 23 | 144,660,549.00 | rs2742604 | 1.8883 | 22_bit_twopoint_parametric_dominant |
| 23 | 144,688,088.00 | rs17271843 | 1.9412 | 22_bit_twopoint_parametric_recessive |
| 23 | 144,748,012.00 | JHU_X.143829532 | 2.2811 | 22_bit_twopoint_parametric_recessive |
| 23 | 144,809,050.00 | rs35113120 | 2.6131 | 21_bit_twopoint_parametric_dominant |
| 23 | 144,993,486.00 | rs1120527 | 2.2564 | 21_bit_twopoint_parametric_recessive |
| 23 | 145,858,875.00 | rs142777228 | 2.2112 | 21_bit_twopoint_parametric_dominant |
| 23 | 147,500,348.00 | rs5951989 | 1.887 | 21_bit_twopoint_parametric_recessive |
| 23 | 151,925,424.00 | rs2855263 | 1.9166 | 21_bit_twopoint_parametric_dominant |
| 23 | 151,930,634.00 | rs5970164 | 1.9422 | 21_bit_twopoint_parametric_dominant |
| 23 | 153,801,095.00 | rs5987158 | 1.8646 | 21_bit_twopoint_parametric_dominant |
| 23 | 153,801,095.00 | rs5987158 | 2.088 | 21_bit_twopoint_parametric_recessive |

SUPPLEMENTARY TABLE 5 (Leighanne Main)

**Supplementary Table 5 – Suggestive Linkage Analysis results.** Loci that surpassed the suggestive linkage threshold (≥ 1.86, < 3.3) for all analyses. For each SNP, chromosome number is followed by genomic coordinates (hg38), SNP identifier, LOD score, and to which analysis this LOD score corresponds.
