## Supplemental Table 6 for "Genetic analysis of cognitive preservation in the midwestern Amish reveals a novel locus on chromosome 2"

| Population | Genotype Frequency | | | | | | Allele Frequency | |
| --- | --- | --- | --- | --- | --- | --- | --- | --- |
|  | TT | | TC | | CC | |  |  |
|  | CU | CI | CU | CI | CU | CI | C | T |
| Amish chromosome 2-linked large pedigree (MORGAN analysis) | 7 (32%) | 0 | 12 (55%) | 0 | 3 (14%) | 2 (100%) | 46% | 54% |
| Amish Population^a^ | 19 (3%) | 6 (2%) | 118 (19%) | 64 (20%) | 489 (78%) | 250 (78%) | 88% | 12% |
| dbSNP European Reference Population |  |  |  |  |  |  | 20% | 80% |

SUPPLEMENTARY TABLE 6 (Leighanne Main)

Supplementary Table 6 – **rs1402906 Genotype and Allele Frequencies**. Genotype count and frequency is listed separately for CU and CI individuals; frequency is calculated as a percentage for each group (e.g., CU or CI). ^a^8,222 individuals from the large Amish pedigree were used to calculate allele frequencies. ALFA allele frequency of European ancestry on dbSNP was used as a reference population because it includes all non-Hispanic Whites of European ancestry.
